# Use of routine health data to monitor malaria intervention effectiveness: a scoping review

**DOI:** 10.1101/2024.12.01.24318260

**Authors:** Richard Reithinger, Donal Bisanzio, Anya Cushnie, Jessica Craig

## Abstract

The expansive scale-up of malaria interventions has contributed to substantial reductions in malaria morbidity and mortality in the past 15–20 years. The effectiveness of these interventions has traditionally been estimated through research studies and trials, nationally representative surveys, and mathematical modelling. Because of their sheer volume across space and time, programmatic data collected and reported routinely through health management information systems (HMIS) can complement and even offer an alternative to nationally representative and other ad hoc surveys to assess health intervention effectiveness, and ultimately impact on health outcomes. The objective of this scoping review was to describe the different analytical approaches for estimating the impact and effectiveness of malaria interventions using routine HMIS and surveillance data.

We examined PubMed using combination searches of the following terms: “malaria” AND “intervention” AND “effect*” OR “impact” AND “system” OR “surveillance”. We limited inclusion to studies and analyses that were conducted in the past decade. We purposefully chose this time cut-off, as that is when countries’ routine HMIS began to substantially mature, with data reported by these systems progressively becoming more robust. Out of 957 records generated from the PubMed search, following title and abstract screening, 93 were included for full-text review, with 49 records ultimately meeting the inclusion criteria and being included in the scoping review.

We summarize included studies by publication year, geography, outcome variables, target populations, interventions assessed, HMIS data platform used—we show that analytical approaches used a range of modelling and non-modelling approaches to assess intervention effectiveness.

This scoping review shows that routine HMIS data can also be used to regularly assess the effectiveness of various malaria interventions—an important exercise to ensure that implemented malaria interventions continue to be effective, have the desired effect, and ultimately help countries progress towards their national strategic goals and targets.

**Strengths and Limitations of this Study:**

► This scoping review describes the different analytical approaches for estimating the impact and effectiveness of malaria interventions using routine health management information system (HMIS) and surveillance data.
► A range of analytical approaches to assess malaria intervention effectiveness using routine HMIS and surveillance data were identified in the records (studies) reviewed, which broadly can be categorized into modelling and non-modelling approaches.
► Limitations lie in the inclusion criteria and main literature database used for the review: some papers and grey literature may not have been included, as well as papers in languages other than English may have been missed.

**Article Summary Line:** Monitoring malaria intervention effectiveness

## BACKGROUND

Malaria is an acute febrile illness caused by a parasitic infection transmitted by *Anopheles* mosquitoes. Human malaria is caused by five different *Plasmodium* parasites, with *P. falciparum* being the predominant species in sub-Saharan Africa (SSA).[1] In the past 15–20 years, the combined efforts of Ministries of Health (MOHs) and National Malaria Programs (NMPs), and their partners, have made tremendous progress against malaria. Thus, in 2022, the estimated global malaria incidence was 58.43 per 1,000 people at risk, a 25% reduction since 2002; similarly, in the same time period, malaria mortality rates have decreased by nearly 50% to 14.82 per 100,000 people in 2021.[2] This progress resulted from the massive scale-up of various malaria prevention and control interventions, including facility and community-based confirmatory testing and treatment of malaria cases, intermittent preventive treatment during pregnancy (IPTp), and seasonal malaria chemoprevention (SMC), along with indoor residual spraying of households with insecticide (IRS) and insecticide-treated nets (ITNs).[1–3]

MOH / NMPs use programmatic intervention coverage and effectiveness data to regularly monitor impact of interventions, modify intervention implementation approaches (e.g., if coverage estimates are sub-par), or switch interventions altogether (e.g., when the effectiveness of an intervention is observed to be lower than expected or waning). Intervention coverage and effectiveness has traditionally been assessed by post-intervention campaign surveys (e.g., post-ITN distribution campaign surveys),[4–6] periodic nationally representative surveys (e.g., Demographic and Health Surveys [DHS], Malaria Indicator Surveys [MIS], Multiple Indicator Cluster Surveys [MICS]),[7–10] or estimated by complex mathematical modelling.[11, 12] While these approaches are generally robust, they have limitations. For example, nationally-representative surveys only occur every 2–5 years; take time; require significant human, logistical and financial resources, and capabilities; and may not be powered sufficiently to provide sub-national intervention estimates. Similarly, mathematical modelling may be limited by the available data and the significant technical expertise needed to develop and run the models, let alone run them continuously. Additionally, neither surveys nor modelling may avail necessary estimates at key strategic moments in the malaria programming planning, implementation, and monitoring cycle, such as the development of key national strategies or design of necessary donor documents (e.g., national malaria strategic plans, Global Fund to Fight AIDS, Tuberculosis and Malaria Concept Notes or U.S. President’s Malaria Initiative Malaria Operational Plans).

Countries’ national health management information systems (HMIS) have been dramatically strengthened in the past decade,[13] with countries being able to consistently and fully report on outpatient, inpatient and other programmatic data—much of this progress has been made following the wide adoption, piloting and roll out of the district health management information system 2 (DHIS2), an open-source data-system software specifically developed to capture health data in lower-and-middle income countries.[14] Because of their sheer volume across space and time, data collected and reported through HMIS like DHIS2 can complement and even offer an alternative to nationally representative and other ad hoc surveys to assess health intervention coverage and effectiveness, and ultimately impact on health outcomes.[15–17]

The objective of this scoping review was to describe the different analytical approaches for estimating the impact and effectiveness of malaria interventions using routine HMIS and surveillance data. To our knowledge, such a review has not been conducted. A preliminary search for existing scoping and systematic reviews on the topic was conducted on December 15, 2023, using PubMed, and no similar reviews were found.

## MATERIALS & METHODS

To conduct and report this scoping review, we followed the PRISMA-ScR (Preferred Reporting Items for Systematic Reviews and Meta-Analysis extension for Scoping Reviews) (**S1 File: PRISMA-ScR Checklist**).[18]. The detailed published protocol is available on Protocol.io [19].

A systematic search of PubMed was conducted on January 2, 2024, to identify studies and analyses that had used routine surveillance and HMIS data to assess the effectiveness of malaria interventions. A detailed search strategy was designed and piloted to identify the optimal combination of keywords used. We examined PubMed using combination searches of the following terms: “malaria” AND “intervention” AND “effect*” OR “impact” AND “system” OR “surveillance”. Other key terms such as "routine” or “information systems” were not included in the search strategy to have a more comprehensive initial search and were used during abstract and full text screening. All identified studies were imported into Rayyan, a systematic review management software, to screen (title, abstract, and full text) and manage the results of the search.[20]

The first stage of the review involved two reviewers (JC, RR) independently identifying potentially relevant articles based on information provided in the title and abstract. Inclusion criteria were: (1) they addressed malaria, and (2) described an approach using routine HMIS and surveillance data to evaluate the effectiveness of a malaria intervention. Articles were also included if the information provided in the title and abstract was not sufficient to determine if it met the inclusion criteria (i.e., it was tagged as “Maybe” in the Rayyan database). In the event of discordance between the two reviewers (e.g., “Included” / “Excluded” and “Excluded” / “Maybe” tag dyad), a third reviewer (DB) reviewed the titles and abstracts and came to a final decision (i.e., “Included”, “Excluded”, “Maybe”).

The second stage of the review involved at least two of the three reviewers independently reviewing articles’ full text and determining which publications were relevant to the current review. We limited inclusion to studies and analyses that were conducted in the past decade (i.e., the study publication date had to be 2014 onwards, with the study period having to include 2013 at the least). We purposefully chose this time cut-off, as that is the time point when countries’ routine HMIS began to substantially mature, with data reported by these systems progressively becoming more robust.[13]

From the included articles, one reviewer (RR) extracted the data from the articles into a pre-specified MS Excel spreadsheet, specifying the following data elements: (1) author; (2) year of publication; (3) geography; (4) study design; (5) study or time period covered; (6) intervention(s) for which effectiveness and impact was measured; (7) approach to measure effectiveness and impact; (8) health information system platform used for the analyses; (9) indicator variables included in the analyses; (10) target population; (11) key findings; and (12) items from the Template for Intervention Description and Replication (TIDieR) checklist. TIDieR is a 12-item checklist that includes the brief name, why, what (materials), what (procedure), who provided, how, where, when and how much, tailoring, modifications, how well (planned), how well (actual) of a program.[21]

## RESULTS

The PubMed search generated 912 records. Following title and abstract screening, 93 were included for full-text review, with 49 records ultimately meeting the inclusion criteria and being included in the scoping review (**Figure 1**).[22–70] A complete list of all screened records can be found as a supplementary file (**S2 File: Complete Data File**). Main characteristics of the included records (studies) are summarized in **Table 1**. All articles were published in English from 2014–2024.

**Figure 1.**
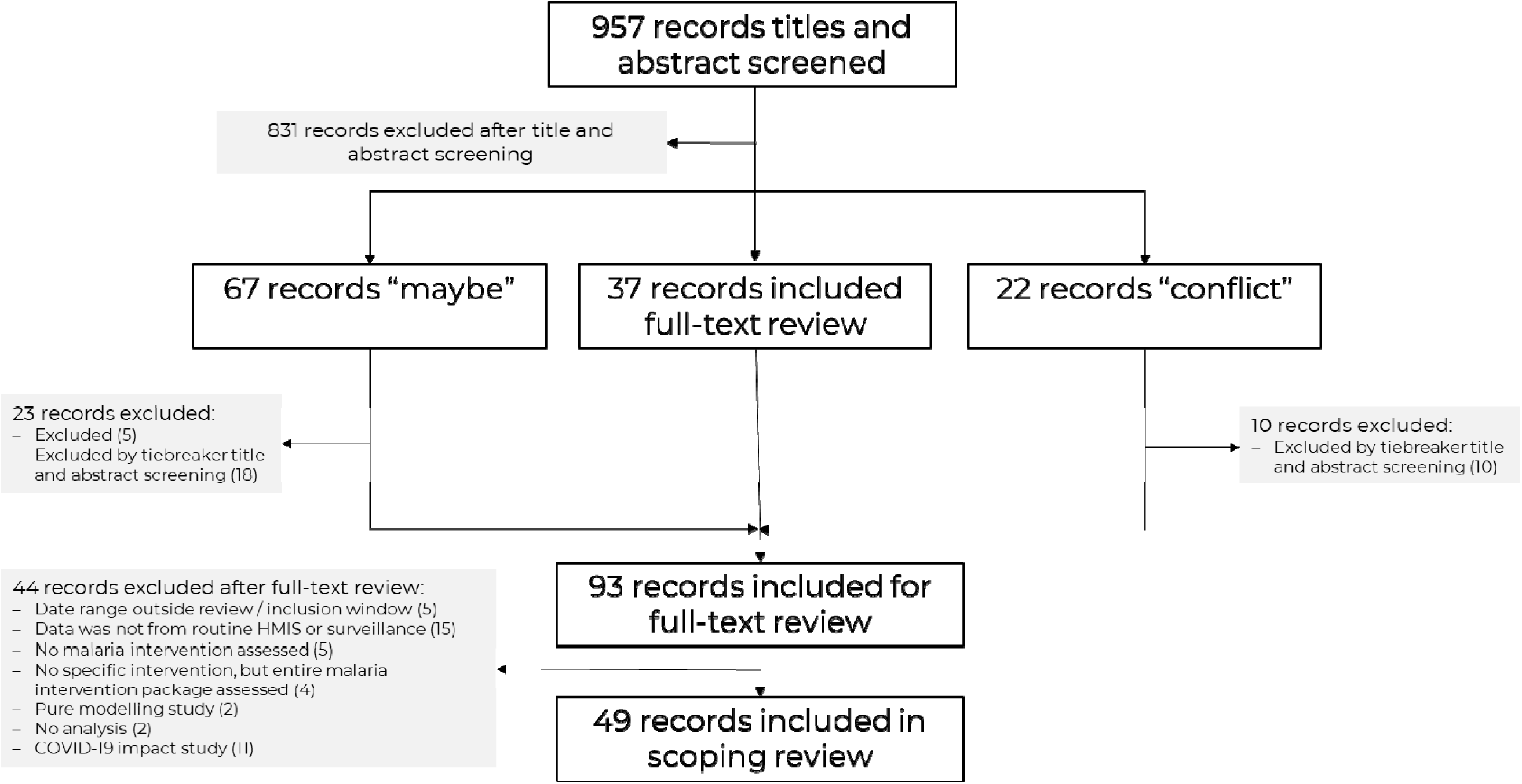
PRISMA Flowchart of Records (Studies) included in Scoping Review.

**Table 1.**
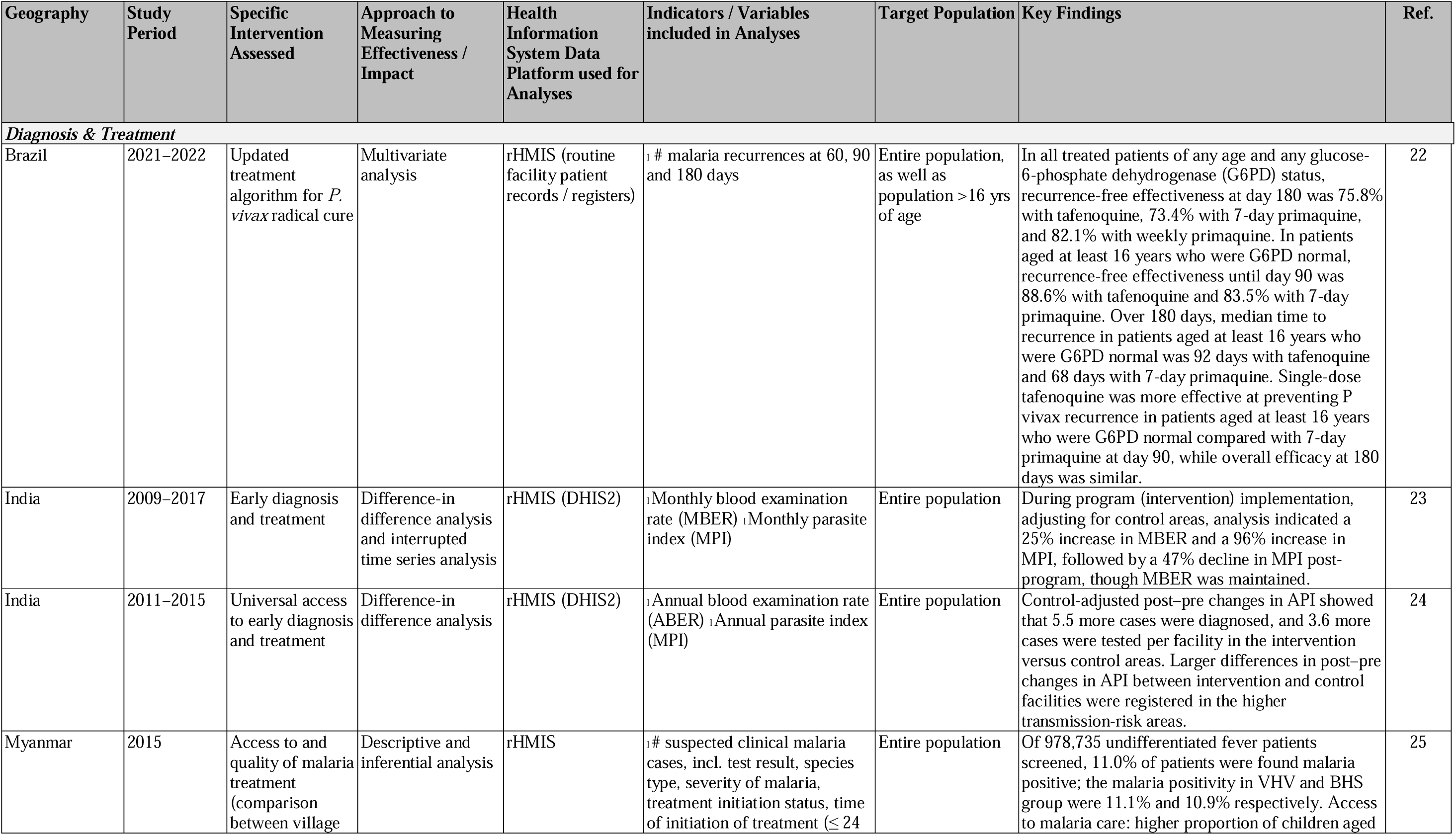
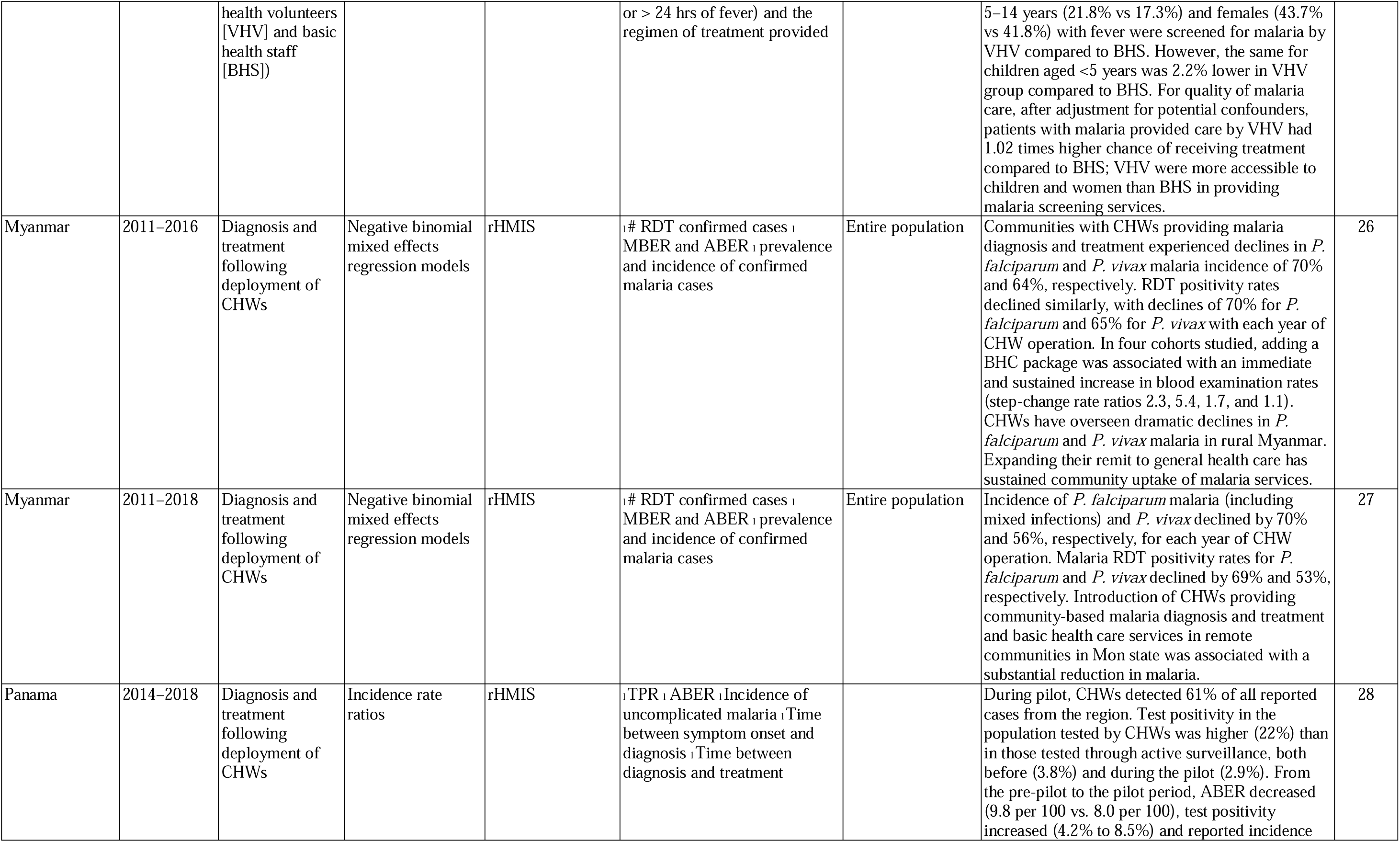
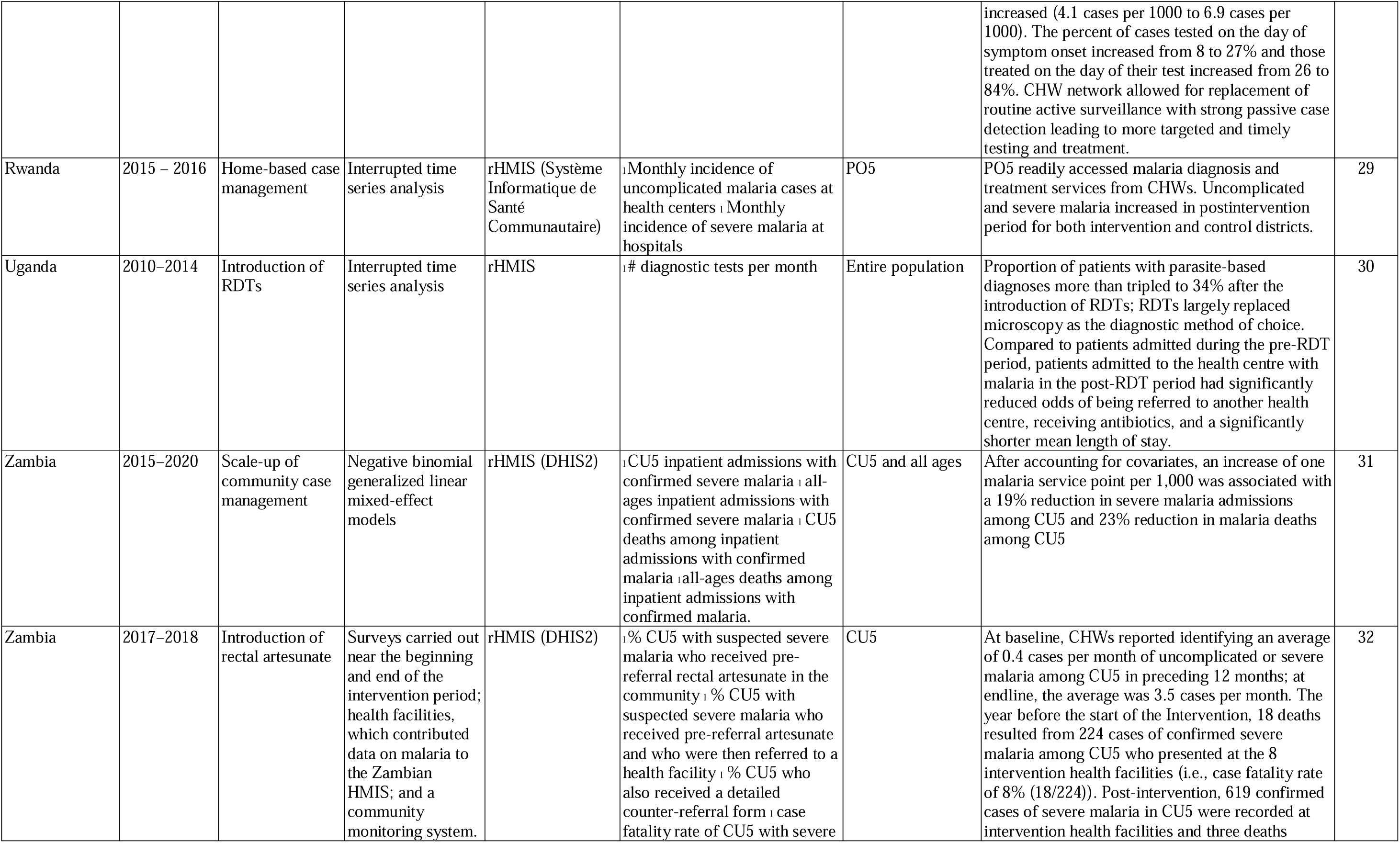
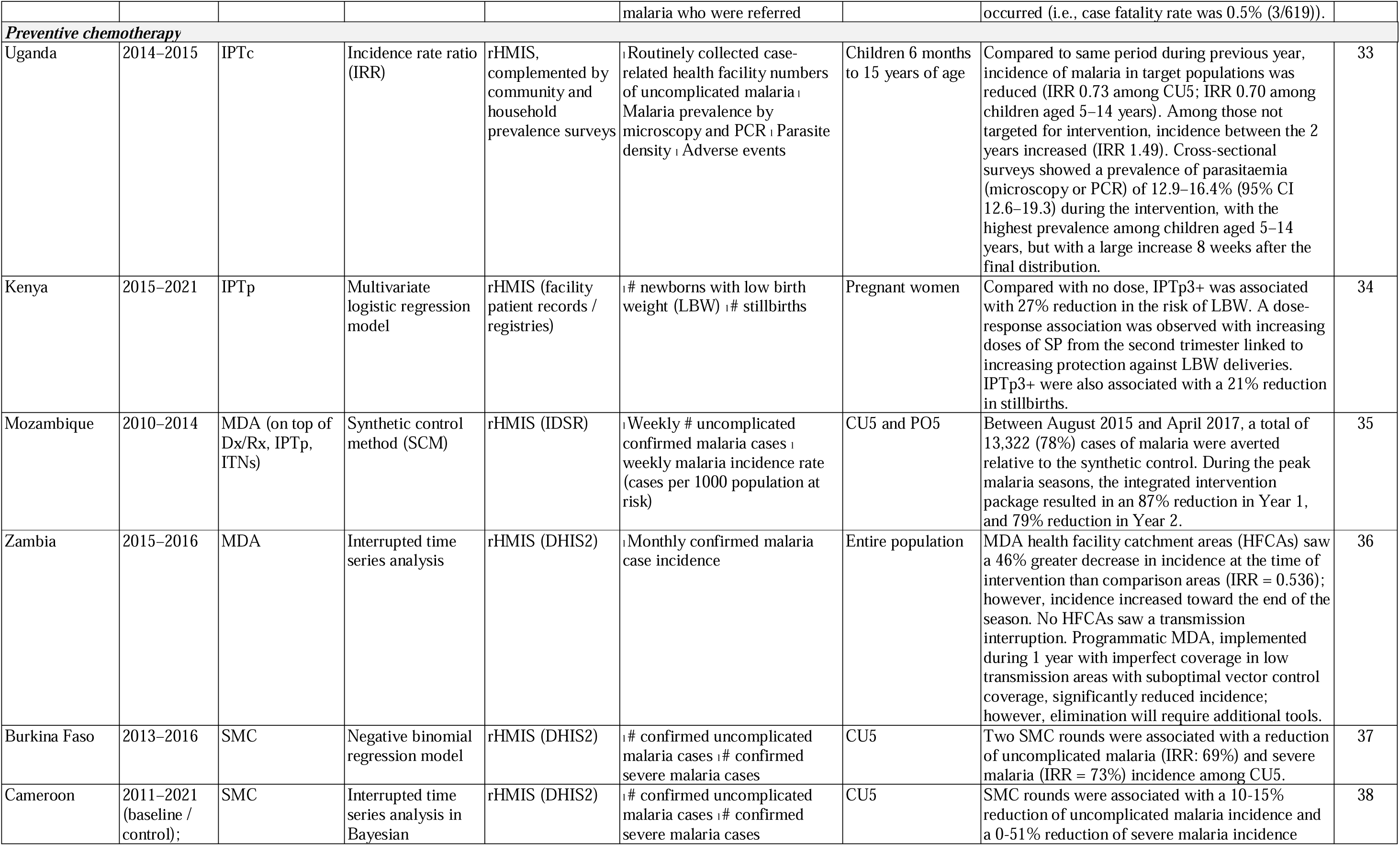
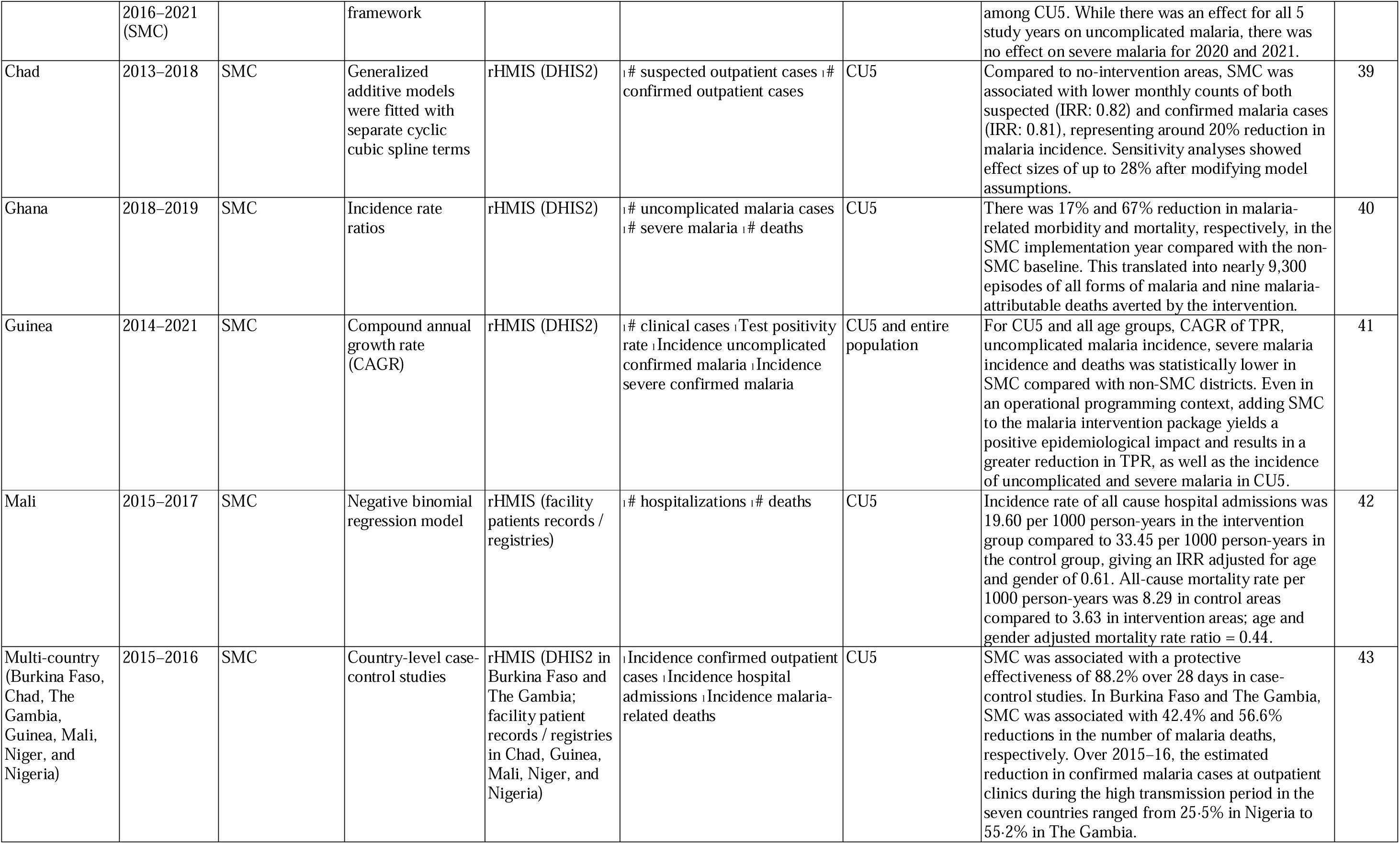
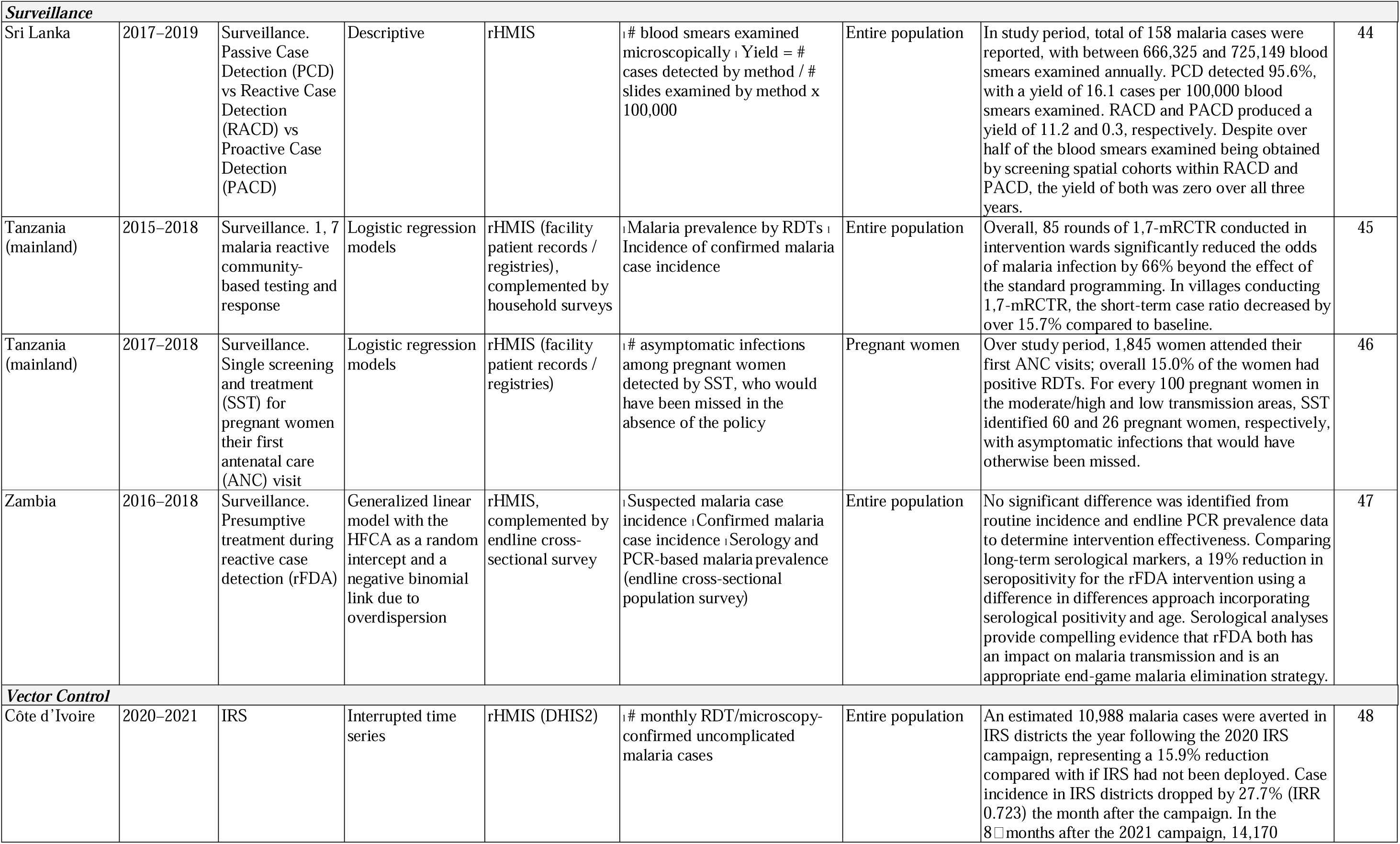
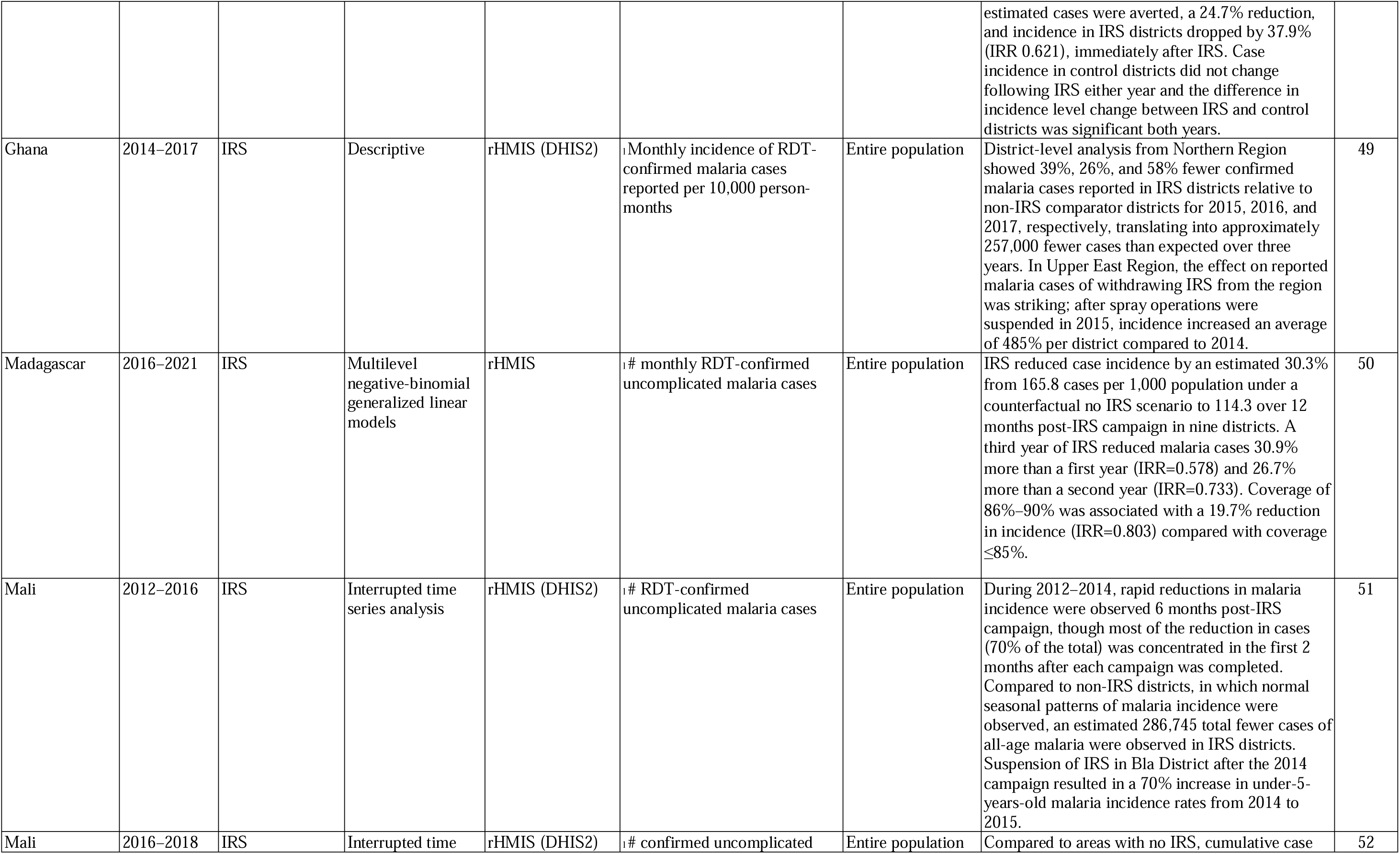
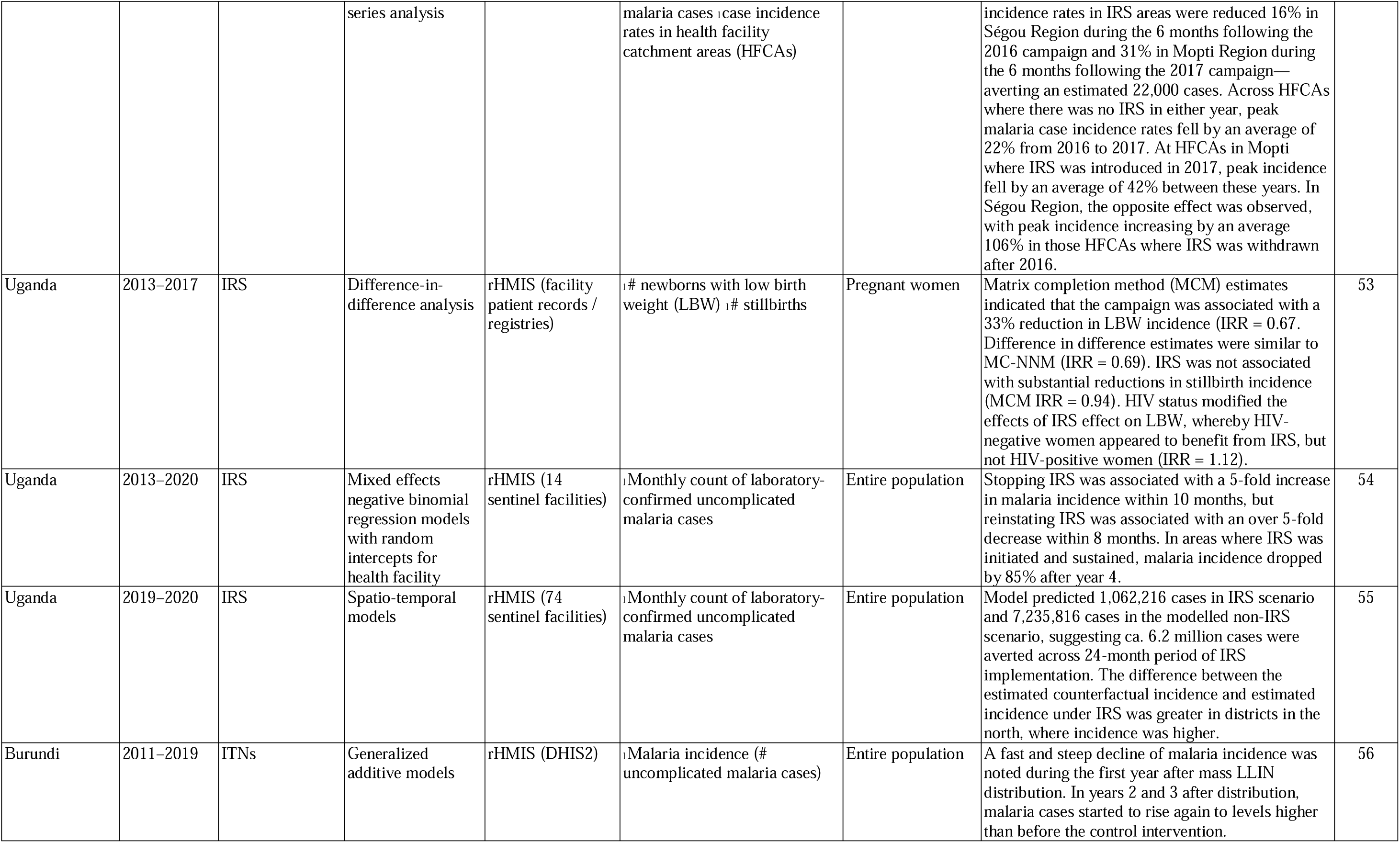
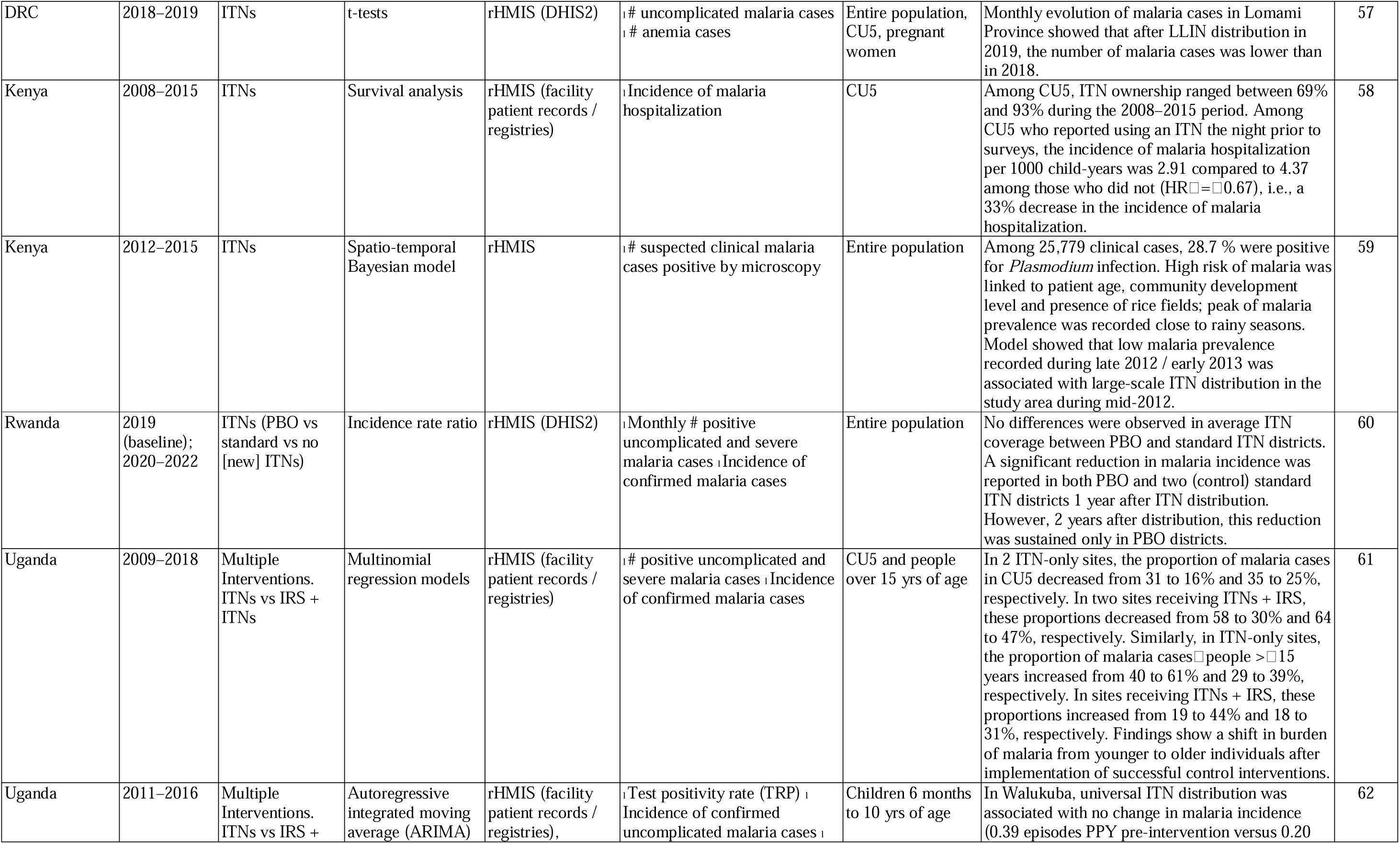
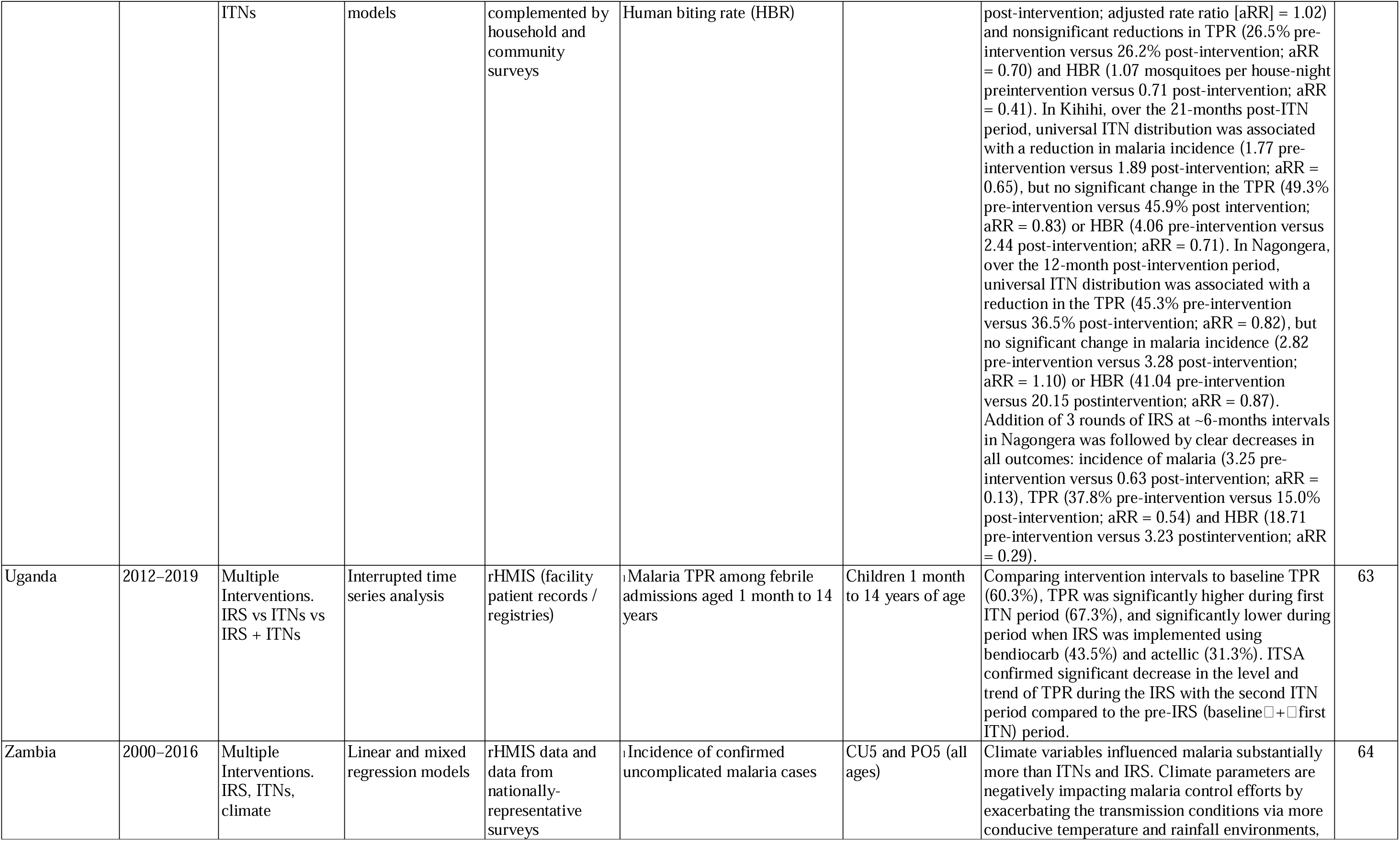
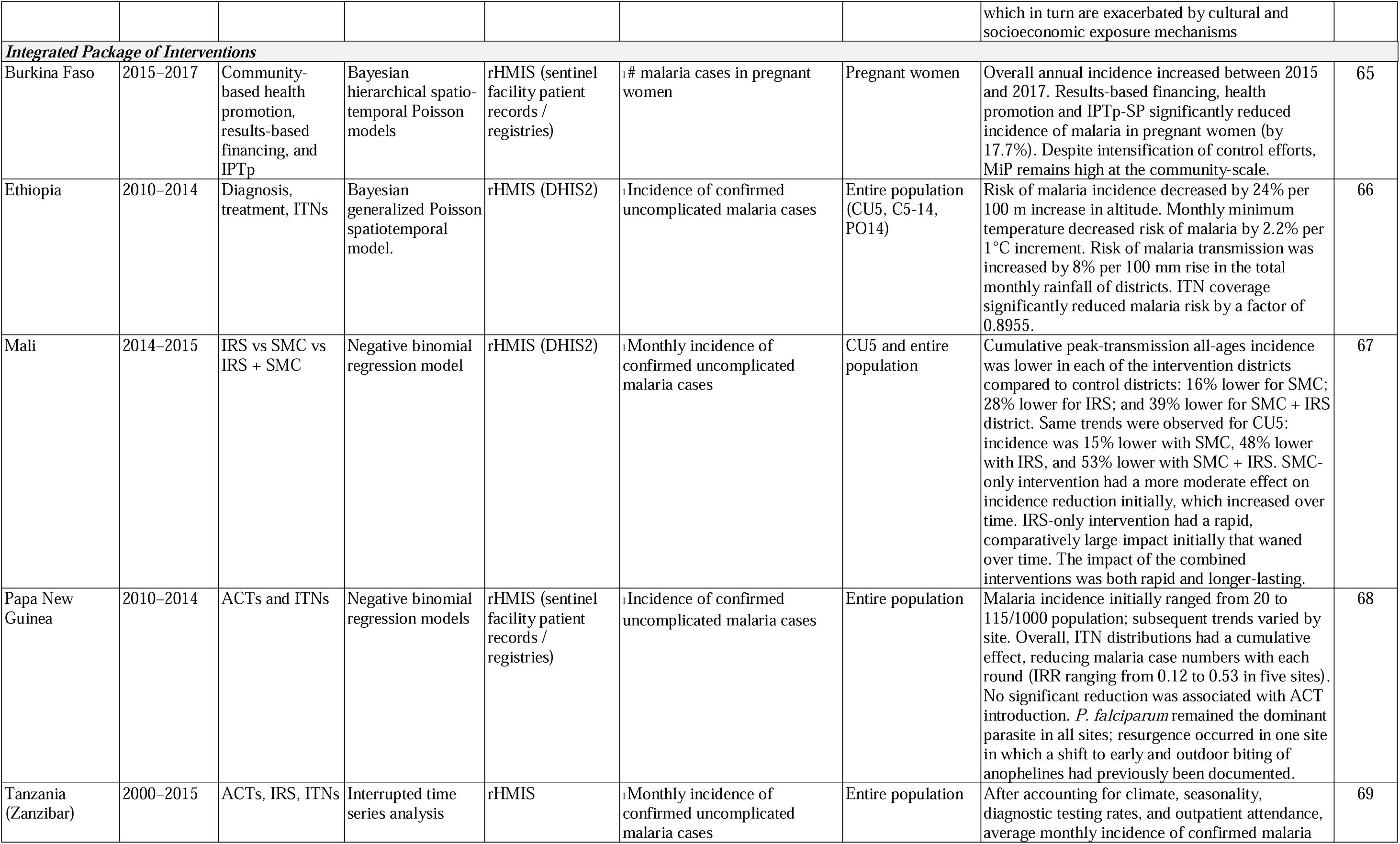
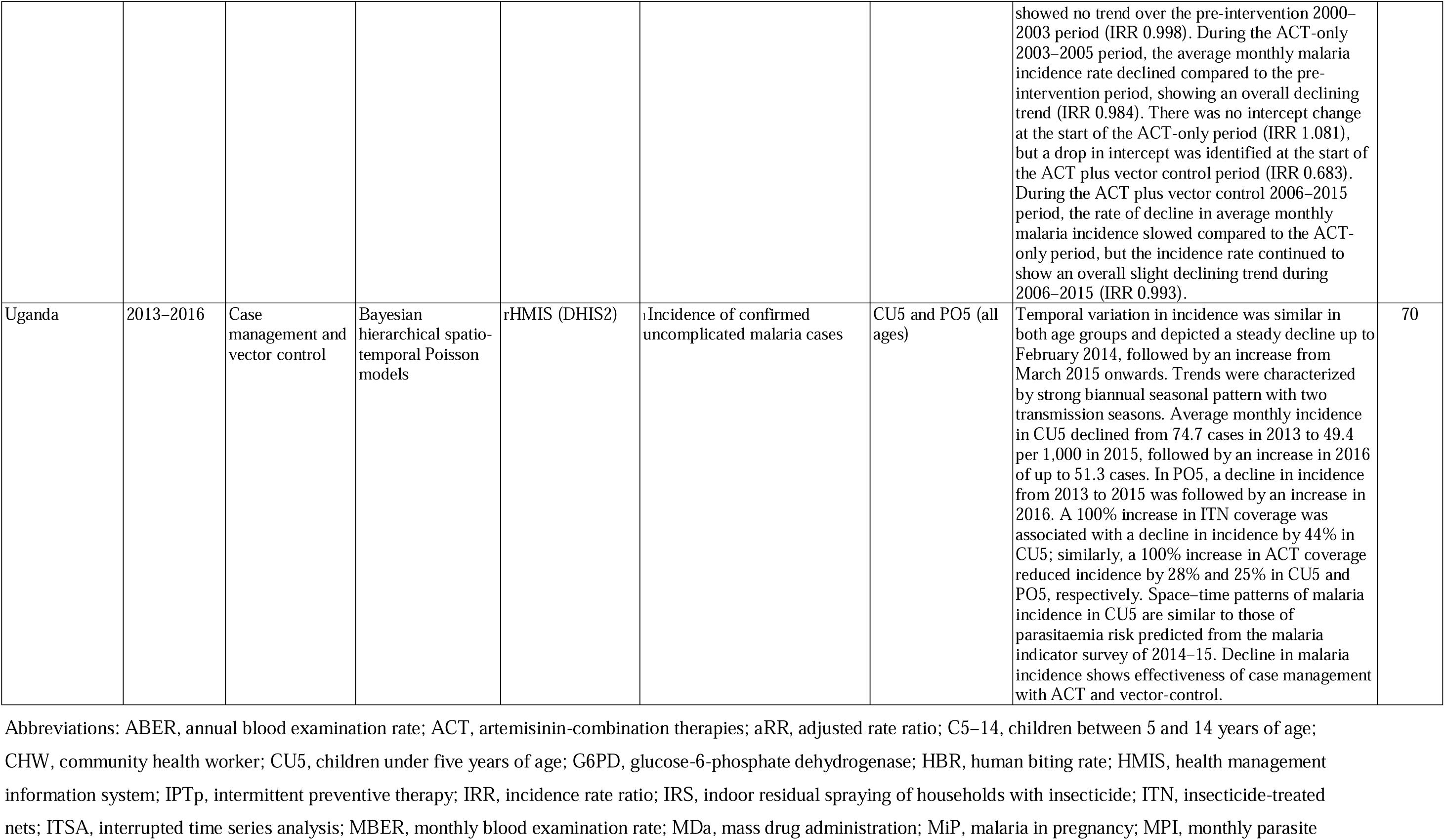
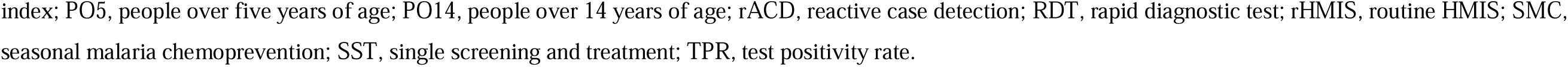
Characteristics of 49 records included in scoping review.

### Publication year

Of the 49 included studies that were conducted between 2013 and 2023, 35 (71.4%) were published between 2020 and 2024; most studies were published in 2022 (n = 11) and 2020 (n = 10).

### Geography

The 49 studies included in the scoping review were conducted in 23 countries; one multi-country study was conducted in five countries.[41] Of included studies, 40 (81.6%) were conducted in Africa, compared to seven (14.3%) and two (4.1%) in Asia and Latin America and the Caribbean, respectively. Countries in which more than three studies were conducted included Uganda (n = 9), Zambia (n = 5), and Mali (n = 4).

### Outcome Variables

Studies included in the scoping review used a range of malariometric outcome indicators, with the majority (n = 36, 73.5%) using either the number of confirmed uncomplicated malaria cases or uncomplicated malaria case incidence (usually defined as 1,000 per population). Many studies included more than just a single outcome indicator in their analyses. Other indicators used in the studies included clinical or suspected malaria, malaria test positivity rate, hospitalizations, deaths, and low birth weight.

### Target populations

Generally, interventions’ effectiveness was assessed for the entire population (n = 23), children under five years of age (n = 8), pregnant women (n = 4); or other specific age groups (e.g., children under 10 years of age, people over five years of age) (n = 4); ten studies included analyses on two or more age groups.

### Interventions evaluated

Interventions, whose effectiveness was assessed included ITNs and/or IRS (n = 18 [36.7%]), diagnosis and/or treatment (n = 12 [24.5%]), SMC (n = 7 [14.3%]), IPTp or intermittent preventive treatment in children (n = 3 [6.1%]), mass drug administration (n = 3 [6.1%]), and some form of active and responsive surveillance (n = 2 [4.1%]). Of the 49 studies included, four assessed the effectiveness of integrated, complementary interventions (e.g., artemisinin-based combination therapies [ACT] and ITNs, or SMC and IRS).

### HMIS data platform used

All studies included in the scoping review used routine HMIS data, some in combination with other data sources (e.g., cross-sectional community and household surveys) (n = 5). Of included studies, 20 (40.8%) specifically stated using the DHIS2 platform, and nine (18.4%) used facility registries (e.g., for antenatal care, outpatients, or inpatients).

### Analytical Methods

Modelling approaches were used in 35 (71.4%) studies to estimate interventions’ effectiveness, including regression models (n = 18), interrupted time series models (n = 12), or Bayesian spatio-temporal models (n = 5). In the remaining 14 studies, non-modelling approaches were used, including comparisons of incidence trends (n = 8), rate ratios (n = 3), difference-in-difference analysis (n = 1), and the compound annual growth rate (n = 1). No study was found comparing modelling and non-modelling approaches.

### Collaborative in-country partnership

Of the 49 studies included in the scoping review, 46 (94%) had co-authors affiliated with in-country institutions, notably the MOH or NMP (n = 31), and/or academic research institutions (n = 24). Three studies did not have a co-author affiliated with MOH, NMP, or local academic research institutions.[36]

## DISCUSSION

As support for malaria control and elimination continues, and additional malaria interventions are introduced (e.g., vaccines, spatial repellents),[1–3] alternative methods for measuring intervention effectiveness and impact must be explored. Controlled and observational research studies or nationally representative surveys to investigate intervention coverage and effectiveness at scale are resource-intensive and costly. Similarly, modelling efforts are complex—they usually require specific types of data and a substantial level of technical expertise, which may not always be available at country level. Following substantial investments by country governments and external assistance partners such as the Global Fund to Fight AIDS, Tuberculosis and Malaria, the U.S. President’s Malaria Initiative, and GAVI, countries’ HMIS have strengthened considerably over the past decade.[13] There is an increasing acknowledgement that routinely (passively) collected data can not only be used to monitor malariometric trends, but can also be used to assess the impact of malaria interventions, and that such an approach—in complement to surveys and modelling—represents a great opportunity to substantially inform programmatic decision-making.[70–74] Moreover, such approach can also be seen as more representative of the true effect and impact of an intervention within a programmatic context rather than the testing and monitoring of intervention effectiveness in a gold-standard, resource-intensive and rich environment (e.g., randomized controlled trials). This change in thinking is reflected in the increasing number of studies conducted and published in the last 3 years that used routine HMIS data to assess the effectiveness of malaria interventions (i.e., of the 49 studies included in our review, 35 [71.4%] were published between 2020–2024). Using routine HMIS data—whether from health facility registries or monthly aggregate, facility-or district-level DHIS2 data—we show that countries can estimate the programmatic impact of most commonly-implemented malaria interventions across facilities, districts, provinces or regions, and at national levels.

For routine HMIS data to be successfully used to assess malaria intervention effectiveness, several pre-requisites need to be in place. First, for countries to be able to use routine HMIS data for such purpose, available data needs to originate from the majority of health service delivery points (i.e., from community-level to tertiary health facilities), as well as be complete, up-to-date and timely, and of high quality.[75] Second, ideally, interventions for which effectiveness is assessed should have high programmatic coverage and be deployed or implemented at a high-quality standard (e.g., SMC-eligible children should receive all preventive treatment doses to protect them from malaria throughout the entire malaria transmission season). Third, the data for all outcome indicators should be robust (e.g., if reported confirmed malaria case incidence is used as outcome indicator, all suspected clinical malaria cases should be tested and confirmed, as otherwise the data will not be a robust representation of malaria trends in a given geographic area). Fourth, efforts to estimate intervention effectiveness using routine HMIS data should be cognizant of possible confounding variables. Thus, if there are geographic variances in the proportion of the population accessing health services (e.g., due to physical distance, commodity stock-outs or disasters), or accessing health services in the public versus private sector in areas where a specific intervention is being implemented, outcome indicator data may under-or over-estimate the effectiveness of the intervention. Similarly, if there are geographic and temporal variances of rainfall and temperature between intervention and non-intervention areas, transmission as well as intervention coverage and use may be impacted—again, under-or over-estimating the implemented interventions’ effectiveness. Consequently, analytical approaches should adjust for these confounders or at least fully address these in the interpretation of the analyses’ findings.

As shown in our scoping review, studies using routine HMIS data to assess intervention effectiveness have been conducted for all main malaria interventions, i.e., for case management, preventive chemotherapy, vector control, or surveillance (**Table 1**). For example, several studies showed the impact of vector control, specifically IRS, in reducing malaria case incidence by 16% in moderate transmission districts in Mali [50] to 85% in high-transmission districts in Uganda [52]. Several studies assessed the impact of SMC in children, showing reductions of 10–55%, 27–51% and 56–67% in uncomplicated malaria case incidence, hospital admissions, and malaria-related deaths, respectively.[36–41] Similarly, several studies showed how the introduction of community health workers and community case management lead to a reduction in malaria incidence,[26, 27] severe malaria admissions,[31] and malaria-related deaths.[31] Importantly, analyses using routine HMIS data also showed the changes in epidemiological trends when intervention are waning or are being discontinued. Thus, in a study in the Burundian highlands, a decline of malaria incidence was noted 20 to 40 weeks after a national mass ITN distribution campaign; yet, from week 80 post-ITN distribution onwards, malaria incidence had rebounded to levels higher than before the intervention.[55] In Uganda, discontinuation of IRS was associated with a 5-fold increase in malaria case incidence within 10 months; subsequent reinstating of IRS was associated with an over 5-fold decrease within 8 months.[53] In Mali, discontinuation of IRS resulted in a 70% increase in malaria case incidence, compared to areas where IRS was continued and malaria case incidence remained the same.[50].

As reported in our scoping review, different analytical approaches have been applied to estimate interventions’ effectiveness from routine HMIS and surveillance data, which broadly can be categorized into modelling and non-modelling approaches. While the former is inherently more robust, as analyses can be adjusted for covariates and confounding variables, they can be complex as requiring a certain level of expertise and specific types of data—ultimately, this also could hamper the approaches’ routine and long-term adoption and use by in-country malaria policymakers and stakeholders. While not having the same level of robustness, non-modelling analytical approaches are simpler to conduct and more readily communicated to and ultimately applied and used by policymakers and stakeholders (e.g., because the approaches are often algebra based, they do not require advanced data analytics or modelling expertise, and thus can be routinely applied by programme personnel with a basic understanding of algebra)—a crucial element in changing a countries’ data analysis and use culture.[75] Interestingly, we found no study that specifically compared different sets of analytical approaches (e.g., modelling vs non-modelling approaches; regression vs Bayesian spatio-temporal models). Ultimately, deciding on which analytical approach to use to assess interventions’ effectiveness will depend on the body of data available for the analyses, the level of robustness and reproducibility required, the skills and expertise available to conduct the analyses, software and computing requirements, and needs for capacity transferability and sustainability (**Table 2**).

**Table 2.**
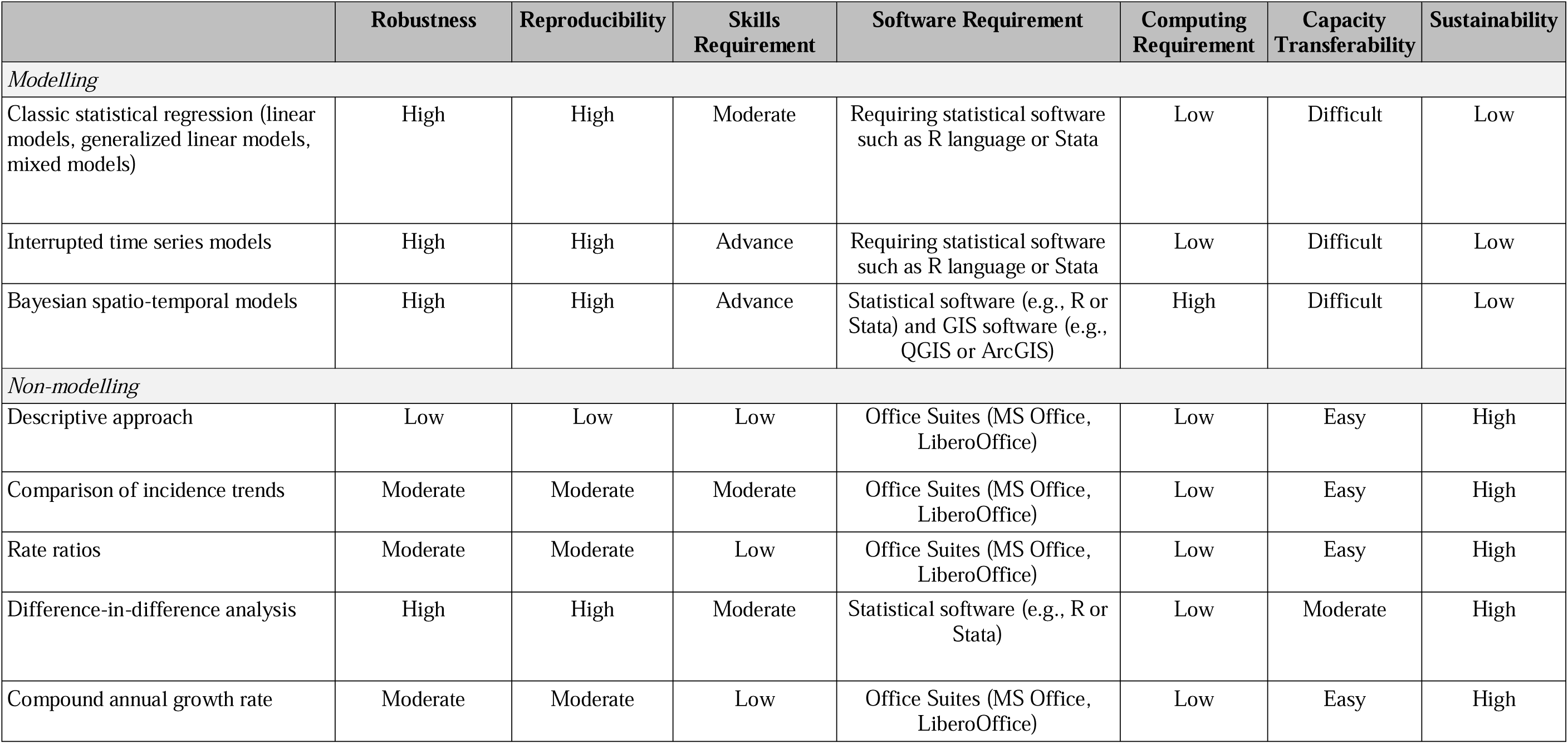
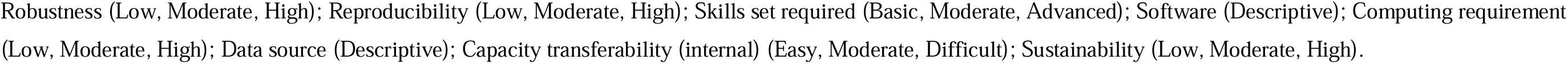
Qualitative comparison of different analytical approaches conducted to assess interventions’ effectiveness using routine HMIS and surveillance data, in terms of technical and other resource requirements, as well as sustainability.

This scoping review has several potential limitations. First, we only searched one database (i.e., PubMed), and some relevant studies may have been excluded. To mitigate against this, we used broad literature database search terms, which resulted in the identification of over 900 records for title and abstract screening. Second, scoping reviews are exploratory in nature and are meant to address broad questions—such as whether routine HMIS data have been used to estimate malaria interventions’ effectiveness. Although the scope was wide, it is evident and clear from this scoping review that this topic is an area of increased interest for researchers and policy makers as they are contemplating the most effective combination of malaria interventions to use when, where and at what scale, in a context of increasingly constrained resources.[76] Third, we restricted interventions as tools and approaches that public health practitioners deploy and implement to reduce the malaria burden, rather than using an even broader definition of intervention (e.g., intervention being any “outside event” affecting intervention effectiveness). For example, several studies assessed the impact of climate change [77–79] or even the recent COVID-19 pandemic [80–82] on malaria burden, which we excluded from our scoping review. Lastly, we did not assess the quality of the studies reported on in the records included in our review, with studies varying in their approach to measure intervention effectiveness, first-and-foremost using either modelling or non-modelling approaches. Parameters to assess the studies’ quality would have to be defined. Of interest could be a study comparing different analytical approaches, not only in terms of the outcome of the analyses (i.e., quantitative metrics of intervention effectiveness), but also resource requirements and programmatic decision-making outcome (i.e., would differences in the analyses’ outcomes have led to a different programmatic decision, such as continuing or discontinuing a specific intervention).

## CONCLUSION

In its 2016–2030 Global Technical Strategy, WHO defines surveillance as a core intervention for malaria prevention and control, making it one of three pillars the strategy is built upon. A strong routine HMIS is critical for efficient and effective malaria surveillance in order to document and report on disease trends at national and subnational levels, both spatially and temporally. This in turn supports strategic and programmatic planning, including for targeting interventions geographically and epidemiologically, thereby optimizing and maximizing the public health impact of available operational, human, and financial resources. The effectiveness of malaria interventions has traditionally been estimated through research studies and trials, nationally representative surveys, and mathematical modelling. This scoping review shows that routine HMIS and surveillance data can also be used to regularly assess the effectiveness of various malaria interventions—an important exercise to ensure that implemented malaria interventions continue to be effective, have the desired effect, and ultimately help countries progress towards their national strategic goals and targets.

## Data Availability

All data produced in the present study are available upon reasonable request to the authors.

## Acknowledgements

The findings and conclusions expressed herein are those of the authors and do not necessarily represent the official position of their employing organizations, the U.S. Agency for International Development, or the United States Government.

## Financial Support

The work was performed with funding provided by the US President’s Malaria Initiative via the U.S. Agency for International Development Country Health Information System and Data Use (CHISU) program (Cooperative Agreement #7200AA20CA00009).

## Conflict of Interest

None to declare.

## Patient Statement

Patients and/or the public were not involved in the design, or conduct, or reporting, or dissemination plans of this research.

## Notes

### Competing Interest Statement

The authors have declared no competing interest.

