## Supplementary material for "Use of routine health data to monitor malaria intervention effectiveness: a scoping review": Supp File 2

<table>
<tr><th>key</th><th>authors</th><th>title</th><th>year</th><th>journal</th><th>volume</th><th>pages</th><th>notes</th><th>doi</th><th>keywords</th><th>pmc_id</th><th>url</th></tr>
<tr><td>rayyan-651229517</td><td>Bhatt S||Weiss DJ||Cameron E||Bisanzio D||Mappin B||Dalrymple U||Battle K||Moyes CL||Henry A||Eck...</td><td>The effect of malaria control on Plasmodium falciparum in Africa between 2000 and 2015.</td><td></td><td></td><td>526</td><td>207-211</td><td>RAYYAN-INCLUSION: {"Donal"=>"Excluded", "Richard"=>"Maybe", "Jessica "=>"Maybe"} | RAYYAN-LABELS: Excluded (tiebreaker) | RAYYAN-EXCLUSION-REASONS: wrong study design | USER-NOTES: {"Richard"=>["Unsure of routine data used, rather than survey data only", "Survey data only"]}</td><td></td><td>Plasmodium falciparum;Malaria;Plasmodium;Africa</td><td>PMC4820050</td><td></td></tr>
<tr><td>rayyan-651229518</td><td>Gonzalez-Sanz M||Berzosa P||Norman FF</td><td>Updates on Malaria Epidemiology and Prevention Strategies.</td><td></td><td></td><td></td><td></td><td>RAYYAN-INCLUSION: {"Richard"=>"Excluded", "Jessica "=>"Excluded"} | RAYYAN-EXCLUSION-REASONS: background article</td><td></td><td>Malaria</td><td>PMC10248987</td><td></td></tr>
<tr><td>rayyan-651229519</td><td>Legendre E||Lehot L||Dieng S||Rebaudet S||Thu AM||Rae JD||Delmas G||Girond F||Herbreteau V||Noste...</td><td>Malaria temporal dynamic clustering for surveillance and intervention planning.</td><td></td><td></td><td>43</td><td>100682</td><td>RAYYAN-INCLUSION: {"Donal"=>"Included", "Richard"=>"Included", "Jessica Craig"=>"Included", "Jessica "=>"Excluded"} | RAYYAN-LABELS: Include (tiebreaker)</td><td></td><td>Malaria</td><td>PMC10280692</td><td></td></tr>
<tr><td>rayyan-651229520</td><td>Gutman JR||Thwing J||Mwesigwa J||McElroy PD||Robertson M</td><td>Routine Healthcare Facility- and Antenatal Care-Based Malaria Surveillance: Challenges and Opportunities.</td><td></td><td></td><td>108</td><td></td><td>RAYYAN-INCLUSION: {"Richard"=>"Excluded", "Jessica "=>"Excluded"} | RAYYAN-EXCLUSION-REASONS: background article,wrong publication type | USER-NOTES: {"Richard"=>["Review"]}</td><td></td><td>Malaria</td><td>PMC9904165</td><td></td></tr>
<tr><td>rayyan-651229521</td><td>Newby G||Cotter C||Roh ME||Harvard K||Bennett A||Hwang J||Chitnis N||Fine S||Stresman G||Chen I||...</td><td>Testing and treatment for malaria elimination: a systematic review.</td><td></td><td></td><td>22</td><td>254</td><td>RAYYAN-INCLUSION: {"Richard"=>"Excluded", "Jessica "=>"Excluded"} | RAYYAN-EXCLUSION-REASONS: background article,wrong publication type | USER-NOTES: {"Richard"=>["Review"]}</td><td></td><td>Malaria</td><td>PMC10476355</td><td></td></tr>
<tr><td>rayyan-651229522</td><td>Sun D||Chen Y||Wang L||Hu X||Wu Q||Liu Y||Liu P||Zeng X||Li S||Wang G||Zhang Y</td><td>Surveillance and Control of Malaria Vectors in Hainan Province, China from 1950 to 2021: A Retrospective Review.</td><td></td><td></td><td>8</td><td></td><td>RAYYAN-INCLUSION: {"Donal"=>"Excluded", "Richard"=>"Excluded", "Jessica Craig"=>"Excluded", "Jessica "=>"Maybe"} | RAYYAN-LABELS: Excluded | RAYYAN-EXCLUSION-REASONS: background article,wrong publication type | USER-NOTES: {"Richard"=>["Review"]}</td><td></td><td>China;Malaria</td><td>PMC10051372</td><td></td></tr>
<tr><td>rayyan-651229523</td><td>Riggle BA||Miller LH||Pierce SK</td><td>Desperately Seeking Therapies for Cerebral Malaria.</td><td></td><td></td><td>204</td><td>327-334</td><td>RAYYAN-INCLUSION: {"Richard"=>"Excluded", "Jessica "=>"Excluded"} | RAYYAN-EXCLUSION-REASONS: background article,wrong publication type | USER-NOTES: {"Richard"=>["Review"]}</td><td></td><td>Malaria, Cerebral;Malaria</td><td>PMC6951433</td><td></td></tr>
<tr><td>rayyan-651229524</td><td>Paaijmans KP||Lobo NF</td><td>Gaps in protection: the actual challenge in malaria elimination.</td><td></td><td></td><td>22</td><td>46</td><td>RAYYAN-INCLUSION: {"Richard"=>"Excluded", "Jessica "=>"Excluded"} | RAYYAN-EXCLUSION-REASONS: background article,wrong publication type | USER-NOTES: {"Richard"=>["Perspective article"]}</td><td></td><td>Malaria</td><td>PMC9902240</td><td></td></tr>
<tr><td>rayyan-651229525</td><td>Auburn S||Barry AE</td><td>Dissecting malaria biology and epidemiology using population genetics and genomics.</td><td></td><td></td><td>47</td><td>77-85</td><td>RAYYAN-INCLUSION: {"Richard"=>"Excluded", "Jessica "=>"Excluded"} | RAYYAN-EXCLUSION-REASONS: wrong outcome,background article,wrong publication type | USER-NOTES: {"Richard"=>["Review"]}</td><td></td><td>Genome;Malaria;Genomics</td><td></td><td></td></tr>
<tr><td>rayyan-651229526</td><td>Karim A||de Savigny D</td><td>Effective Coverage in Health Systems: Evolution of a Concept.</td><td></td><td></td><td>11</td><td></td><td>RAYYAN-INCLUSION: {"Richard"=>"Excluded", "Jessica "=>"Excluded"} | RAYYAN-EXCLUSION-REASONS: wrong outcome,background article,wrong publication type | USER-NOTES: {"Richard"=>["Review"]}</td><td></td><td></td><td>PMC10047489</td><td></td></tr>
<tr><td>rayyan-651229527</td><td>Diawara SI||Konate D||Kayentao K||Mihigo J||Shaffer JG||Sangare M||Ndabamenye P||Swedberg E||Garg...</td><td>Effect of seasonal malaria chemoprevention in children between 5 and 9 years old in Kita and Bafoulabe districts, Mali.</td><td></td><td></td><td>18</td><td>e00258</td><td>RAYYAN-INCLUSION: {"Richard"=>"Excluded", "Jessica "=>"Excluded"} | RAYYAN-EXCLUSION-REASONS: wrong study design | USER-NOTES: {"Richard"=>["Unclear if routine data used", "RCT with cross-sectional surveys"]}</td><td></td><td>Only Child;Child;Chemoprevention;Seasons;Malaria</td><td>PMC9249800</td><td></td></tr>
<tr><td>rayyan-651229528</td><td>Chang W||Cohen J||Wang DQ||Abdulla S||Mahende MK||Gavana T||Scott V||Msuya HM||Mwanyika-Sando M||...</td><td>Impact of 1,7-malaria reactive community-based testing and response (1,7-mRCTR) approach on malaria prevalence in Tanzania.</td><td></td><td></td><td>12</td><td>116</td><td>RAYYAN-INCLUSION: {"Richard"=>"Excluded", "Jessica "=>"Excluded"} | RAYYAN-EXCLUSION-REASONS: wrong study design | USER-NOTES: {"Richard"=>["Unclear if routine data used", "Cross-sectional surveys only to measure impact"]}</td><td></td><td>Prevalence;Malaria</td><td>PMC10726614</td><td></td></tr>
<tr><td>rayyan-651229529</td><td>Shahandeh K||Basseri HR</td><td>Challenges and the Path Forward on Malaria Elimination Intervention: A Systematic Review.</td><td></td><td></td><td>48</td><td>1004-1013</td><td>RAYYAN-INCLUSION: {"Richard"=>"Excluded", "Jessica "=>"Excluded"} | RAYYAN-EXCLUSION-REASONS: wrong study design,wrong publication type</td><td></td><td>Malaria</td><td>PMC6635336</td><td></td></tr>
<tr><td>rayyan-651229530</td><td>Rants'o TA||Koekemoer LL||Panayides JL||van Zyl RL</td><td>Potential of Essential Oil-Based Anticholinesterase Insecticides against Anopheles Vectors: A Review.</td><td></td><td></td><td>27</td><td></td><td>RAYYAN-INCLUSION: {"Richard"=>"Excluded", "Jessica "=>"Excluded"} | RAYYAN-EXCLUSION-REASONS: wrong outcome,wrong publication type | USER-NOTES: {"Richard"=>["Review"]}</td><td></td><td>Oils, Volatile;Insecticides;Cholinesterase Inhibitors</td><td>PMC9611659</td><td></td></tr>
<tr><td>rayyan-651229531</td><td>Fountain A||Ye Y||Roca-Feltrer A||Rowe AK||Camara A||Fofana A||Candrinho B||Hamainza B||Ndiop M||...</td><td>Surveillance as a Core Intervention to Strengthen Malaria Control Programs in Moderate to High Transmission Settings.</td><td></td><td></td><td>108</td><td>13-Aug</td><td>RAYYAN-INCLUSION: {"Richard"=>"Excluded", "Jessica "=>"Excluded"} | RAYYAN-EXCLUSION-REASONS: background article,wrong publication type | USER-NOTES: {"Richard"=>["Review"]}</td><td></td><td>Malaria</td><td>PMC9904156</td><td></td></tr>
<tr><td>rayyan-651229532</td><td>Su XZ||Zhang C||Joy DA</td><td>Host-Malaria Parasite Interactions and Impacts on Mutual Evolution.</td><td></td><td></td><td>10</td><td>587933</td><td>RAYYAN-INCLUSION: {"Richard"=>"Excluded", "Jessica "=>"Excluded"} | RAYYAN-EXCLUSION-REASONS: wrong outcome,wrong publication type</td><td></td><td>Malaria</td><td>PMC7652737</td><td></td></tr>
<tr><td>rayyan-651229533</td><td>Naing C||Whittaker MA||Tanner M</td><td>Inter-sectoral approaches for the prevention and control of malaria among the mobile and migrant populations: a scoping review.</td><td></td><td></td><td>17</td><td>430</td><td>RAYYAN-INCLUSION: {"Donal"=>"Excluded", "Richard"=>"Excluded", "Jessica Craig"=>"Excluded", "Jessica "=>"Maybe"} | RAYYAN-LABELS: Excluded | RAYYAN-EXCLUSION-REASONS: background article,wrong publication type | USER-NOTES: {"Richard"=>["Review"]}</td><td></td><td>Malaria</td><td>PMC6240188</td><td></td></tr>
<tr><td>rayyan-651229534</td><td>Tay DA||Ocansey RTA</td><td>Impact of Urbanization on Health and Well-Being in Ghana. Status of Research, Intervention Strategies and Future Directions: A Rapid Review.</td><td></td><td></td><td>10</td><td>877920</td><td>RAYYAN-INCLUSION: {"Richard"=>"Excluded", "Jessica "=>"Excluded"} | RAYYAN-EXCLUSION-REASONS: background article,wrong publication type | USER-NOTES: {"Richard"=>["Review"]}</td><td></td><td>Urban Health;Ghana</td><td>PMC9273841</td><td></td></tr>
<tr><td>rayyan-651229535</td><td></td><td>malERA: An updated research agenda for combination interventions and modelling in malaria elimination and eradication.</td><td></td><td></td><td>14</td><td>e1002453</td><td>RAYYAN-INCLUSION: {"Richard"=>"Excluded", "Jessica "=>"Excluded"} | RAYYAN-EXCLUSION-REASONS: background article,wrong publication type | USER-NOTES: {"Richard"=>["Review"]}</td><td></td><td>Malaria</td><td>PMC5708628</td><td></td></tr>
<tr><td>rayyan-651229536</td><td>Xu M||Hu YX||Lu SN||Idris MA||Zhou SD||Yang J||Feng XN||Huang YM||Xu X||Chen Y||Wang DQ</td><td>Seasonal malaria chemoprevention in Africa and China's upgraded role as a contributor: a scoping review.</td><td></td><td></td><td>12</td><td>63</td><td>RAYYAN-INCLUSION: {"Richard"=>"Excluded", "Jessica "=>"Excluded"} | RAYYAN-EXCLUSION-REASONS: background article,wrong publication type | USER-NOTES: {"Richard"=>["Review"]}</td><td></td><td>China;Chemoprevention;Seasons;Malaria</td><td>PMC10320994</td><td></td></tr>
<tr><td>rayyan-651229537</td><td>Ashton RA||Bennett A||Yukich J||Bhattarai A||Keating J||Eisele TP</td><td>Methodological Considerations for Use of Routine Health Information System Data to Evaluate Malaria Program Impact in an Era of Declining Malaria Transmission.</td><td></td><td></td><td>97</td><td>46-57</td><td>RAYYAN-INCLUSION: {"Richard"=>"Excluded", "Jessica "=>"Excluded"} | RAYYAN-EXCLUSION-REASONS: background article,wrong publication type | USER-NOTES: {"Richard"=>["Likely a review; unsure if routine HIS example provided", "Probably nice review for context setting"]}</td><td></td><td>Malaria;Information Systems</td><td>PMC5619932</td><td></td></tr>
<tr><td>rayyan-651229538</td><td>Lourenco C||Tatem AJ||Atkinson PM||Cohen JM||Pindolia D||Bhavnani D||Le Menach A</td><td>Strengthening surveillance systems for malaria elimination: a global landscaping of system performance, 2015-2017.</td><td></td><td></td><td>18</td><td>315</td><td>RAYYAN-INCLUSION: {"Richard"=>"Excluded", "Jessica "=>"Excluded"} | RAYYAN-EXCLUSION-REASONS: background article,wrong publication type | USER-NOTES: {"Richard"=>["Review"]}</td><td></td><td>Malaria</td><td>PMC6751607</td><td></td></tr>
<tr><td>rayyan-651229539</td><td>Bisanzio D||Lalji S||Abbas FB||Ali MH||Hassan W||Mkali HR||Al-Mafazy AW||Joseph JJ||Nyinondi S||K...</td><td>Spatiotemporal dynamics of malaria in Zanzibar, 2015-2020.</td><td></td><td></td><td>8</td><td></td><td>RAYYAN-INCLUSION: {"Richard"=>"Excluded", "Jessica "=>"Excluded"} | RAYYAN-EXCLUSION-REASONS: wrong outcome,wrong study design | USER-NOTES: {"Richard"=>["Use of routine data, but no intervention"]}</td><td></td><td>Malaria</td><td>PMC9843203</td><td></td></tr>
<tr><td>rayyan-651229540</td><td>Oduola AO||Obembe A||Adelaja OJ||Adeneye AK||Akilah J||Awolola TS</td><td>Outcome of capacity building intervention for malaria vector surveillance, control and research in Nigerian higher institutions.</td><td></td><td></td><td>17</td><td>193</td><td>RAYYAN-INCLUSION: {"Richard"=>"Excluded", "Jessica "=>"Excluded"} | RAYYAN-EXCLUSION-REASONS: wrong outcome,wrong study design</td><td></td><td>Malaria</td><td>PMC5952629</td><td></td></tr>
<tr><td>rayyan-651229541</td><td>Seijas-Chavez JA||Nolan MS||Lynn MK||da Rocha MJF||da Costa Araujo M||Fonseca FLA||Laporta GZ</td><td>Causal effects on low Apgar at 5-min and stillbirth in a malaria maternal-fetal health outcome investigation: a large perinatal surveillance study in the Brazilian Amazon.</td><td></td><td></td><td>20</td><td>444</td><td>RAYYAN-INCLUSION: {"Richard"=>"Excluded", "Jessica "=>"Excluded"} | RAYYAN-EXCLUSION-REASONS: wrong outcome,wrong study design | USER-NOTES: {"Richard"=>["Epi study"]}</td><td></td><td>Malaria</td><td>PMC8614005</td><td></td></tr>
<tr><td>rayyan-651229542</td><td>Namuganga JF||Epstein A||Nankabirwa JI||Mpimbaza A||Kiggundu M||Sserwanga A||Kapisi J||Arinaitwe ...</td><td>The impact of stopping and starting indoor residual spraying on malaria burden in Uganda.</td><td></td><td></td><td>12</td><td>2635</td><td>RAYYAN-INCLUSION: {"Richard"=>"Included", "Jessica "=>"Included"}</td><td></td><td>Uganda;Malaria</td><td>PMC8113470</td><td></td></tr>
<tr><td>rayyan-651229543</td><td>Asiedu A||Haws RA||Gyasi A||Boateng P||Malm K||Ntumy R||Oseni L||Tetteh G</td><td>Improving Malaria Case Management and Referral Relationships at the Primary Care Level in Ghana: Evaluation of a Quality Assurance Internship.</td><td></td><td></td><td>11</td><td></td><td>RAYYAN-INCLUSION: {"Richard"=>"Excluded", "Jessica "=>"Excluded"} | RAYYAN-EXCLUSION-REASONS: wrong outcome,wrong study design</td><td></td><td>Internship and Residency;Case Management;Referral and Consultation;Ghana</td><td>PMC10749655</td><td></td></tr>
<tr><td>rayyan-651229544</td><td>Ampofo GD||Osarfo J||Aberese-Ako M||Asem L||Komey MN||Mohammed W||Ofosu AA||Tagbor H</td><td>Malaria in pregnancy control and pregnancy outcomes: a decade's overview using Ghana's DHIMS II data.</td><td></td><td></td><td>21</td><td>303</td><td>RAYYAN-INCLUSION: {"Richard"=>"Included", "Jessica "=>"Included"}</td><td></td><td>Pregnancy;Pregnancy Outcome;Ghana;Malaria</td><td>PMC9615308</td><td></td></tr>
<tr><td>rayyan-651229545</td><td>Smith JL||Ghimire P||Rijal KR||Maglior A||Hollis S||Andrade-Pacheco R||Das Thakur G||Adhikari N||...</td><td>Designing malaria surveillance strategies for mobile and migrant populations in Nepal: a mixed-methods study.</td><td></td><td></td><td>18</td><td>158</td><td>RAYYAN-INCLUSION: {"Donal"=>"Excluded", "Richard"=>"Excluded", "Jessica Craig"=>"Excluded", "Jessica "=>"Maybe"} | RAYYAN-LABELS: Excluded | RAYYAN-EXCLUSION-REASONS: wrong outcome,wrong study design | USER-NOTES: {"Richard"=>["UNclear if intervention effect was</td><td></td><td>Nepal;Malaria</td><td>PMC6500027</td><td></td></tr>
<tr><td>rayyan-651229546</td><td>Wangdi K||Wetzler E||Marchesini P||Villegas L||Canavati S</td><td>Cross-border malaria drivers and risk factors on the Brazil-Venezuela border between 2016 and 2018.</td><td></td><td></td><td>12</td><td>6058</td><td>RAYYAN-INCLUSION: {"Richard"=>"Excluded", "Jessica "=>"Excluded"} | RAYYAN-EXCLUSION-REASONS: wrong outcome,wrong study design | USER-NOTES: {"Richard"=>["Epi study"]}</td><td></td><td>Emigration and Immigration;Risk Factors;Malaria</td><td>PMC9001644</td><td></td></tr>
<tr><td>rayyan-651229547</td><td>Beloconi A||Nyawanda BO||Bigogo G||Khagayi S||Obor D||Danquah I||Kariuki S||Munga S||Vounatsou P</td><td>Malaria, climate variability, and interventions: modelling transmission dynamics.</td><td></td><td></td><td>13</td><td>7367</td><td>RAYYAN-INCLUSION: {"Donal"=>"Excluded", "Richard"=>"Maybe", "Jessica "=>"Excluded"} | RAYYAN-LABELS: Excluded (tiebreaker) | RAYYAN-EXCLUSION-REASONS: wrong outcome,wrong study design | USER-NOTES: {"Richard"=>["UNclear if intervention effect measured / adjusted for"]}</td><td></td><td>Malaria</td><td>PMC10161998</td><td></td></tr>
</table>

<table>
<tr><th></th><th></th><th></th><th></th><th></th><th></th><th></th><th></th></tr>
<tr><td>rayyan-651229548</td><td>Killeen GF||Reed TE</td><td>The portfolio effect cushions mosquito populations and malaria transmission against vector control interventions.</td><td>17</td><td>291</td><td>RAYYAN-INCLUSION: {"Richard"=>"Excluded", "Jessica "=>"Excluded"} | RAYYAN-EXCLUSION-REASONS: wrong outcome,wrong study design</td><td>Malaria;Culicidae</td><td>PMC6086012</td></tr>
<tr><td>rayyan-651229549</td><td>Epstein A||Namuganga JF||Nabende I||Kamya EV||Kamya MR||Dorsey G||Sturrock H||Bhatt S||Rodriguez-...</td><td>Mapping malaria incidence using routine health facility surveillance data in Uganda.</td><td>8</td><td></td><td>RAYYAN-INCLUSION: {"Richard"=>"Included", "Jessica "=>"Included"}</td><td>Uganda;Health Facilities;Malaria</td><td>PMC10201255</td></tr>
<tr><td>rayyan-651229550</td><td>Alencar Filho AC||Lacerda MV||Okoshi K||Okoshi MP</td><td>Malaria and vascular endothelium.</td><td>103</td><td>165-9</td><td>RAYYAN-INCLUSION: {"Richard"=>"Excluded", "Jessica "=>"Excluded"} | RAYYAN-EXCLUSION-REASONS: wrong outcome,wrong study design</td><td>Endothelium, Vascular;Malaria</td><td>PMC4150669</td></tr>
<tr><td>rayyan-651229551</td><td>Kone S||Baikoro N||N'Guessan Y||Jaeger FN||Silue KD||Furst T||Hurlimann E||Ouattara M||Seka MC||N...</td><td>Health & Demographic Surveillance System Profile: The Taabo Health and Demographic Surveillance System, Cote d'Ivoire.</td><td>44</td><td>87-97</td><td>RAYYAN-INCLUSION: {"Donal"=>"Excluded", "Richard"=>"Excluded", "Jessica Craig"=>"Excluded", "Jessica "=>"Maybe"} | RAYYAN-LABELS: Excluded | RAYYAN-EXCLUSION-REASONS: wrong study design,wrong publication type,wrong infection / disease | USER-NOTES: {"Richard"=>["Unclear</td><td></td><td></td></tr>
<tr><td>rayyan-651229552</td><td>Davis L||Prosnitz D||Ye Y</td><td>The Value Added of Incorporating Qualitative Approaches into Malaria Surveillance, Monitoring, and Evaluation.</td><td>108</td><td>32-37</td><td>RAYYAN-INCLUSION: {"Richard"=>"Excluded", "Jessica "=>"Excluded"} | RAYYAN-EXCLUSION-REASONS: wrong outcome,wrong study design</td><td>Malaria</td><td>PMC9904161</td></tr>
<tr><td>rayyan-651229553</td><td>Jacobson JO||Cueto C||Smith JL||Hwang J||Gosling R||Bennett A</td><td>Surveillance and response for high-risk populations: what can malaria elimination programmes learn from the experience of HIV?</td><td>16</td><td>33</td><td>RAYYAN-INCLUSION: {"Richard"=>"Excluded", "Jessica "=>"Excluded"} | RAYYAN-EXCLUSION-REASONS: wrong outcome,background article,wrong study design</td><td>Malaria</td><td>PMC5241929</td></tr>
<tr><td>rayyan-651229554</td><td>Vigodny A||Ben Aharon M||Wharton-Smith A||Fialkoff Y||Houri-Yafin A||Braganca F||Soares Da Graca ...</td><td>Digitally managed larviciding as a cost-effective intervention for urban malaria: operational lessons from a pilot in Sao Tome and Principe guided by the Zzapp system.</td><td>22</td><td>114</td><td>RAYYAN-INCLUSION: {"Donal"=>"Excluded", "Richard"=>"Excluded", "Jessica Craig"=>"Excluded", "Jessica "=>"Maybe"} | RAYYAN-LABELS: Excluded | RAYYAN-EXCLUSION-REASONS: wrong outcome,wrong study design | USER-NOTES: {"Richard"=>["Don't think routine data were used"]}</td><td>Cost-Benefit Analysis;Malaria</td><td>PMC10080920</td></tr>
<tr><td>rayyan-651229555</td><td>Ssempiira J||Kissa J||Nambuusi B||Mukooyo E||Opigo J||Makumbi F||Kasasa S||Vounatsou P</td><td>Interactions between climatic changes and intervention effects on malaria spatio-temporal dynamics in Uganda.</td><td>3</td><td>e00070</td><td>RAYYAN-INCLUSION: {"Donal"=>"Included", "Richard"=>"Maybe", "Jessica "=>"Maybe"} | RAYYAN-LABELS: Include (tiebreaker) | USER-NOTES: {"Richard"=>["While focus is on climate change, abstract mention how climate change affects can affect interventions"]}</td><td>Uganda;Malaria</td><td>PMC6020080</td></tr>
<tr><td>rayyan-651229556</td><td>Visser T||Bruxvoort K||Maloney K||Leslie T||Barat LM||Allan R||Ansah EK||Anyanti J||Boulton I||Cl...</td><td>Introducing malaria rapid diagnostic tests in private medicine retail outlets: A systematic literature review.</td><td>12</td><td>e0173093</td><td>RAYYAN-INCLUSION: {"Richard"=>"Excluded", "Jessica "=>"Excluded"} | RAYYAN-EXCLUSION-REASONS: wrong outcome,background article,wrong study design</td><td>Malaria</td><td>PMC5333947</td></tr>
<tr><td>rayyan-651229557</td><td>Mbuagbaw L||Medley N||Darzi AJ||Richardson M||Habiba Garga K||Ongolo-Zogo P</td><td>Health system and community level interventions for improving antenatal care coverage and health outcomes.</td><td>2015</td><td>CD010994</td><td>RAYYAN-INCLUSION: {"Richard"=>"Excluded", "Jessica "=>"Excluded"} | RAYYAN-EXCLUSION-REASONS: background article,wrong publication type</td><td></td><td>PMC4676908</td></tr>
<tr><td>rayyan-651229558</td><td>Kirakoya-Samadoulougou F||De Brouwere V||Fokam AF||Ouedraogo M||Ye Y</td><td>Assessing the effect of seasonal malaria chemoprevention on malaria burden among children under 5 years in Burkina Faso.</td><td>21</td><td>143</td><td>RAYYAN-INCLUSION: {"Richard"=>"Included", "Jessica "=>"Included"}</td><td>Chemoprevention;Seasons</td><td>PMC9074217</td></tr>
<tr><td>rayyan-651229559</td><td>Perera R||Caldera A||Wickremasinghe AR</td><td>Reactive Case Detection (RACD) and foci investigation strategies in malaria control and elimination: a review.</td><td>19</td><td>401</td><td>RAYYAN-INCLUSION: {"Richard"=>"Excluded", "Jessica "=>"Excluded"} | RAYYAN-EXCLUSION-REASONS: background article,wrong study design</td><td>Malaria</td><td>PMC7653886</td></tr>
<tr><td>rayyan-651229560</td><td>Kouassi BL||Edi C||Ouattara AF||Ekra AK||Bellai LG||Gouamene J||Kacou YAK||Kouame JKI||Beke AO||Y...</td><td>Entomological monitoring data driving decision-making for appropriate and sustainable malaria vector control in Cote d'Ivoire.</td><td>22</td><td>14</td><td>RAYYAN-INCLUSION: {"Richard"=>"Excluded", "Jessica "=>"Excluded"} | RAYYAN-EXCLUSION-REASONS: wrong outcome,wrong study design</td><td>Decision Making;Malaria;Poly dA-dT</td><td>PMC9835745</td></tr>
<tr><td>rayyan-651229561</td><td>Winskill P||Walker PG||Cibulskis RE||Ghani AC</td><td>Prioritizing the scale-up of interventions for malaria control and elimination.</td><td>18</td><td>122</td><td>RAYYAN-INCLUSION: {"Richard"=>"Excluded", "Jessica "=>"Excluded"} | RAYYAN-EXCLUSION-REASONS: background article,wrong study design</td><td>Malaria</td><td>PMC6454681</td></tr>
<tr><td>rayyan-651229562</td><td>DePina A||Barros H||Tiffany A||Stresman G</td><td>Sustaining surveillance as an intervention during the COVID-19 pandemic in Cabo Verde and implications for malaria elimination.</td><td>13</td><td>956864</td><td>RAYYAN-INCLUSION: {"Donal"=>"Excluded", "Richard"=>"Maybe", "Jessica "=>"Maybe"} | RAYYAN-LABELS: Excluded (C19),Excluded (tiebreaker) | USER-NOTES: {"Richard"=>["Unclear if intervention effect measured"]}</td><td>Malaria</td><td>PMC9582766</td></tr>
<tr><td>rayyan-651229563</td><td>Pradhan MM||Pradhan S||Dutta A||Shah NK||Valecha N||Joshi PL||Pradhan K||Grewal Daumerie P||Baner...</td><td>Impact of the malaria comprehensive case management programme in Odisha, India.</td><td>17</td><td>e0265352</td><td>RAYYAN-INCLUSION: {"Donal"=>"Included", "Richard"=>"Included", "Jessica "=>"Excluded"} | RAYYAN-LABELS: Include (tiebreaker)</td><td>Case Management;India;Malaria</td><td>PMC8947122</td></tr>
<tr><td>rayyan-651229564</td><td>Coleman S||Yihdego Y||Gyamfi F||Kolyada L||Tongren JE||Zigirumugabe S||Dery DB||Badu K||Obiri-Dan...</td><td>Estimating malaria transmission risk through surveillance of human-vector interactions in northern Ghana.</td><td>16</td><td>205</td><td>RAYYAN-INCLUSION: {"Richard"=>"Excluded", "Jessica "=>"Excluded"} | RAYYAN-EXCLUSION-REASONS: wrong outcome,wrong study design</td><td>Humanities;Humanism;Humans;Ghana;Malaria</td><td>PMC10280856</td></tr>
<tr><td>rayyan-651229565</td><td>Mwangungulu SP||Dorothea D||Ngereja ZR||Kaindoa EW</td><td>Geospatial based model for malaria risk prediction in Kilombero valley, South-eastern, Tanzania.</td><td>18</td><td>e0293201</td><td>RAYYAN-INCLUSION: {"Richard"=>"Excluded", "Jessica "=>"Excluded"} | RAYYAN-EXCLUSION-REASONS: wrong outcome,wrong study design | USER-NOTES: {"Richard"=>["Risk map"]}</td><td>Malaria</td><td>PMC10597495</td></tr>
<tr><td>rayyan-651229566</td><td>Green C||Quigley P||Kureya T||Barber C||Chanda E||Moyo B||Mpande B||Mubuyaeta K||Mudenda M||Mundi...</td><td>Rectal artesunate for severe malaria, implementation research, Zambia.</td><td>101</td><td>371-380A</td><td>RAYYAN-INCLUSION: {"Donal"=>"Excluded", "Richard"=>"Included", "Jessica "=>"Excluded"} | RAYYAN-LABELS: Excluded (tiebreaker)</td><td>Malaria</td><td>PMC10225942</td></tr>
<tr><td>rayyan-651229567</td><td>Hardy A||Haji K||Abbas F||Hassan J||Ali A||Yussuf Y||Cook J||Rosu L||Houri-Yafin A||Vigodny A||Oa...</td><td>Cost and quality of operational larviciding using drones and smartphone technology.</td><td>22</td><td>286</td><td>RAYYAN-INCLUSION: {"Donal"=>"Excluded", "Richard"=>"Excluded", "Jessica "=>"Maybe"} | RAYYAN-LABELS: Excluded (tiebreaker) | RAYYAN-EXCLUSION-REASONS: wrong outcome,wrong study design</td><td></td><td>PMC10523724</td></tr>
<tr><td>rayyan-651229568</td><td>Desai MR||Samuels AM||Odongo W||Williamson J||Odero NA||Otieno K||Shi YP||Kachur SP||Hamel MJ||Ka...</td><td>Impact of Intermittent Mass Testing and Treatment on Incidence of Malaria Infection in a High Transmission Area of Western Kenya.</td><td>103</td><td>369-377</td><td>RAYYAN-INCLUSION: {"Donal"=>"Excluded", "Richard"=>"Included", "Jessica "=>"Excluded"} | RAYYAN-LABELS: Excluded (tiebreaker)</td><td>Kenya;Malaria</td><td>PMC7356446</td></tr>
<tr><td>rayyan-651229569</td><td>Baliga BS||Baliga S||Jain A||Kulal N||Kumar M||Koduvattat N||Kumar BGP||Kumar A||Ghosh SK</td><td>Digitized smart surveillance and micromanagement using information technology for malaria elimination in Mangaluru, India: an analysis of five-year post-digitization data.</td><td>20</td><td>139</td><td>RAYYAN-INCLUSION: {"Donal"=>"Excluded", "Richard"=>"Excluded", "Jessica "=>"Included"} | RAYYAN-LABELS: Excluded (tiebreaker) | RAYYAN-EXCLUSION-REASONS: wrong outcome,wrong study design | USER-NOTES: {"Richard"=>["Unclear what interventions were included, if any"]}</td><td>India;Malaria</td><td>PMC7938374</td></tr>
<tr><td>rayyan-651229570</td><td>Fondjo E||Toto JC||Tchouakui M||Eyisap WE||Patchoke S||Menze B||Njeambosay B||Zeukeug F||Ngomdjum...</td><td>High vector diversity and malaria transmission dynamics in five sentinel sites in Cameroon.</td><td>22</td><td>123</td><td>RAYYAN-INCLUSION: {"Richard"=>"Excluded", "Jessica "=>"Excluded"} | RAYYAN-EXCLUSION-REASONS: wrong outcome,wrong study design</td><td>Cameroon;Malaria</td><td>PMC10100606</td></tr>
<tr><td>rayyan-651229571</td><td>Beshir KB||Muwanguzi J||Nader J||Mansukhani R||Traore A||Gamougam K||Ceesay S||Bazie T||Kolie F||...</td><td>Prevalence of Plasmodium falciparum haplotypes associated with resistance to sulfadoxine-pyrimethamine and amodiaquine before and after upscaling of seasonal malaria chemoprevention in seven African countries: a genomic surveillance study.</td><td>23</td><td>361-370</td><td>RAYYAN-INCLUSION: {"Richard"=>"Excluded", "Jessica "=>"Excluded"} | RAYYAN-EXCLUSION-REASONS: wrong outcome,wrong study design</td><td>Sulfadoxine;Plasmodium falciparum;Prevalence;Haplotypes;Genomics;Pyrimethamine;</td><td></td></tr>
<tr><td>rayyan-651229572</td><td>Ali R||Wan Mohamad Ali WN||Wilson Putit P</td><td>Updating the Data on Malaria Vectors in Malaysia: Protocol for a Scoping Review.</td><td>12</td><td>e39798</td><td>RAYYAN-INCLUSION: {"Richard"=>"Excluded", "Jessica "=>"Excluded"} | RAYYAN-EXCLUSION-REASONS: wrong outcome,background article,wrong study design</td><td>Malaysia;Malaria</td><td>PMC10028518</td></tr>
<tr><td>rayyan-651229573</td><td>Issiaka D||Barry A||Traore T||Diarra B||Cook D||Keita M||Sagara I||Duffy P||Fried M||Dicko A</td><td>Impact of seasonal malaria chemoprevention on hospital admissions and mortality in children under 5 years of age in Ouelessebougou, Mali.</td><td>19</td><td>103</td><td>RAYYAN-INCLUSION: {"Donal"=>"Included", "Richard"=>"Excluded", "Jessica "=>"Excluded"} | RAYYAN-LABELS: Excluded (tiebreaker) | RAYYAN-EXCLUSION-REASONS: wrong study design</td><td>Only Child;Child;Seasons;Chemoprevention;Malaria</td><td>PMC7055064</td></tr>
<tr><td>rayyan-651229574</td><td>Runge M||Snow RW||Molteni F||Thawer S||Mohamed A||Mandike R||Giorgi E||Macharia PM||Smith TA||Len...</td><td>Simulating the council-specific impact of anti-malaria interventions: A tool to support malaria strategic planning in Tanzania.</td><td>15</td><td>e0228469</td><td>RAYYAN-INCLUSION: {"Donal"=>"Excluded", "Richard"=>"Maybe", "Jessica "=>"Excluded"} | RAYYAN-LABELS: Excluded (tiebreaker) | USER-NOTES: {"Richard"=>["Unclear if routine data was used"]}</td><td>Malaria</td><td>PMC7029840</td></tr>
<tr><td>rayyan-651229575</td><td>Kavuma M||Mars M</td><td>The effect of an integrated electronic medical record system on malaria out-patient case management in a Ugandan health facility.</td><td>28</td><td>14604582<br>22113744<br>6</td><td>RAYYAN-INCLUSION: {"Donal"=>"Excluded", "Richard"=>"Excluded", "Jessica "=>"Maybe"} | RAYYAN-LABELS: Excluded (tiebreaker) | RAYYAN-EXCLUSION-REASONS: wrong outcome,wrong study design</td><td>Medical Records;Health Facilities;Case Management</td><td></td></tr>
<tr><td>rayyan-651229576</td><td>Zhou G||Zhong D||Lee MC||Wang X||Atieli HE||Githure JI||Githeko AK||Kazura J||Yan G</td><td>Multi-Indicator and Multistep Assessment of Malaria Transmission Risks in Western Kenya.</td><td>104</td><td>1359-1370</td><td>RAYYAN-INCLUSION: {"Donal"=>"Included", "Richard"=>"Excluded", "Jessica "=>"Excluded"} | RAYYAN-LABELS: Excluded (tiebreaker) | RAYYAN-EXCLUSION-REASONS: wrong study design</td><td>Kenya;Malaria</td><td>PMC8045647</td></tr>
<tr><td>rayyan-651229577</td><td>Shi B||Lin S||Tan Q||Cao J||Zhou X||Xia S||Zhou XN||Liu J</td><td>Inference and prediction of malaria transmission dynamics using time series data.</td><td>9</td><td>95</td><td>RAYYAN-INCLUSION: {"Richard"=>"Excluded", "Jessica "=>"Excluded"} | RAYYAN-EXCLUSION-REASONS: wrong study design,wrong study duration | USER-NOTES: {"Richard"=>["Unclear if routine data was used and which interventions were included"]}</td><td>Malaria</td><td>PMC7367373</td></tr>
<tr><td>rayyan-651229578</td><td>Anwar MN||Hickson RI||Mehra S||Price DJ||McCaw JM||Flegg MB||Flegg JA</td><td>Optimal Interruption of P. vivax Malaria Transmission Using Mass Drug Administration.</td><td>85</td><td>43</td><td>RAYYAN-INCLUSION: {"Richard"=>"Excluded", "Jessica "=>"Excluded"} | RAYYAN-EXCLUSION-REASONS: wrong study design | USER-NOTES: {"Richard"=>["Model, not using routine data"]}</td><td>Malaria, Vivax;Malaria</td><td>PMC10115738</td></tr>
<tr><td>rayyan-651229579</td><td>Karema C||Wen S||Sidibe A||Smith JL||Gosling R||Hakizimana E||Tanner M||Noor AM||Tatarsky A</td><td>History of malaria control in Rwanda: implications for future elimination in Rwanda and other malaria-endemic countries.</td><td>19</td><td>356</td><td>RAYYAN-INCLUSION: {"Richard"=>"Excluded", "Jessica "=>"Excluded"} | RAYYAN-EXCLUSION-REASONS: background article,wrong study design</td><td>Malaria</td><td>PMC7539391</td></tr>
</table>

| | | | | | | | |
|---|---|---|---|---|---|---|---|
| rayyan-651229580 | Boggild AK\|\|Geduld J\|\|Libman M\|\|Ward BJ\|\|McCarthy AE\|\|Doyle PW\|\|Ghesquiere W\|\|Vincelette J\|\|Kuhn ... | Travel-acquired infections and illnesses in Canadians: surveillance report from CanTravNet surveillance data, 2009-2011. | 8 | e20-32 | RAYYAN-INCLUSION: {"Richard"=>"Excluded", "Jessica "=>"Excluded"} \| RAYYAN-EXCLUSION-REASONS: wrong outcome,wrong study design,wrong study duration | Travel | PMC4085092 |
| rayyan-651229581 | Graham HR\|\|Maher J\|\|Bakare AA\|\|Nguyen CD\|\|Ayede AI\|\|Oyewole OB\|\|Gray A\|\|Izadnegahdar R\|\|Duke T\|\|F... | Oxygen systems and quality of care for children with pneumonia, malaria and diarrhoea: Analysis of a stepped-wedge trial in Nigeria. | 16 | e0254229 | RAYYAN-INCLUSION: {"Richard"=>"Excluded", "Jessica "=>"Excluded"} \| RAYYAN-EXCLUSION-REASONS: wrong outcome,wrong study design | Oxygenators;Nigeria;Malaria;Only Child;Child | PMC8266122 |
| rayyan-651229582 | Adjei MR\|\|Kubio C\|\|Buamah M\|\|Sarfo A\|\|Suuri T\|\|Ibrahim S\|\|Sadiq A\|\|Abubakari II\|\|Baafi JV | Effectiveness of seasonal malaria chemoprevention in reducing under-five malaria morbidity and mortality in the Savannah Region, Ghana. | 56 | 64-70 | RAYYAN-INCLUSION: {"Richard"=>"Included", "Jessica "=>"Included"} | Ghana;Malaria;Seasons;Chemoprevention | PMC10336463 |
| rayyan-651229583 | Shretta R\|\|Silal SP\|\|Celhay OJ\|\|Gran Mercado CE\|\|Kyaw SS\|\|Avancena A\|\|Fox K\|\|Zelman B\|\|Baral R\|\|W... | Malaria elimination transmission and costing in the Asia-Pacific: Developing an investment case. | 4 | 60 | RAYYAN-INCLUSION: {"Richard"=>"Excluded", "Jessica "=>"Excluded"} \| RAYYAN-EXCLUSION-REASONS: wrong outcome,wrong study design | Malaria;Asia | PMC6974926 |
| rayyan-651229584 | Gutman JR\|\|Mwesigwa JN\|\|Arnett K\|\|Kangale C\|\|Aaron S\|\|Babarinde D\|\|Buekens J\|\|Candrinho B\|\|Debe S... | Using antenatal care as a platform for malaria surveillance data collection: study protocol. | 22 | 99 | RAYYAN-INCLUSION: {"Richard"=>"Excluded", "Jessica "=>"Excluded"} \| RAYYAN-EXCLUSION-REASONS: wrong publication type \| USER-NOTES: {"Richard"=>["Protocol publication"]} | Malaria | PMC10022568 |
| rayyan-651229585 | Messenger LA\|\|Furnival-Adams J\|\|Chan K\|\|Pelloquin B\|\|Paris L\|\|Rowland M | Vector control for malaria prevention during humanitarian emergencies: a systematic review and meta-analysis. | 11 | e534-e545 | RAYYAN-INCLUSION: {"Richard"=>"Excluded", "Jessica "=>"Excluded"} \| RAYYAN-EXCLUSION-REASONS: wrong outcome,background article,wrong study design | Malaria;Emergencies | |
| rayyan-651229586 | Chen-Hussey V\|\|Behrens R\|\|Logan JG | Assessment of methods used to determine the safety of the topical insect repellent N,N-diethyl-m-toluamide (DEET). | 7 | 173 | RAYYAN-INCLUSION: {"Richard"=>"Excluded", "Jessica "=>"Excluded"} \| RAYYAN-EXCLUSION-REASONS: wrong outcome,wrong study design | | PMC4041896 |
| rayyan-651229587 | Torre S\|\|Langlais D\|\|Gros P | Genetic analysis of cerebral malaria in the mouse model infected with Plasmodium berghei. | 29 | 488-506 | RAYYAN-INCLUSION: {"Richard"=>"Excluded", "Jessica "=>"Excluded"} \| RAYYAN-EXCLUSION-REASONS: wrong outcome,wrong study design | Mice;Malaria;Plasmodium | |
| rayyan-651229588 | Geubbels E\|\|Amri S\|\|Levira F\|\|Schellenberg J\|\|Masanja H\|\|Nathan R | Health & Demographic Surveillance System Profile: The Ifakara Rural and Urban Health and Demographic Surveillance System (Ifakara HDSS). | 44 | 848-61 | RAYYAN-INCLUSION: {"Richard"=>"Excluded", "Jessica "=>"Excluded"} \| RAYYAN-EXCLUSION-REASONS: background article,wrong study design | Urban Health | |
| rayyan-651229589 | Homan T\|\|Hiscox A\|\|Mweresa CK\|\|Masiga D\|\|Mukabana WR\|\|Oria P\|\|Maire N\|\|Pasquale AD\|\|Silkey M\|\|Ala... | The effect of mass mosquito trapping on malaria transmission and disease burden (SolarMal): a stepped-wedge cluster-randomised trial. | 388 | 1193-201 | RAYYAN-INCLUSION: {"Donal"=>"Excluded", "Richard"=>"Excluded", "Jessica "=>"Excluded"} | Culicidae;Malaria | |
| rayyan-651229590 | Burkot TR\|\|Farlow R\|\|Min M\|\|Espino E\|\|Mnzava A\|\|Russell TL | A global analysis of National Malaria Control Programme vector surveillance by elimination and control status in 2018. | 18 | 399 | RAYYAN-INCLUSION: {"Richard"=>"Excluded", "Jessica "=>"Excluded"} \| RAYYAN-EXCLUSION-REASONS: background article,wrong study design | Malaria | PMC6894334 |
| rayyan-651229591 | Sarker AR\|\|Sultana M | Cost-effective analysis of childhood malaria vaccination in endemic hotspots of Bangladesh. | 15 | e0233902 | RAYYAN-INCLUSION: {"Richard"=>"Excluded", "Jessica "=>"Excluded"} \| RAYYAN-EXCLUSION-REASONS: wrong study design | Bangladesh;Cost-Benefit Analysis;Vaccination;Malaria | PMC7259743 |
| rayyan-651229592 | Kittichai V\|\|Kaewthamasorn M\|\|Samung Y\|\|Jomtarak R\|\|Naing KM\|\|Tongloy T\|\|Chuwongin S\|\|Boonsang S | Automatic identification of medically important mosquitoes using embedded learning approach-based image-retrieval system. | 13 | 10609 | RAYYAN-INCLUSION: {"Richard"=>"Excluded", "Jessica "=>"Excluded"} \| RAYYAN-EXCLUSION-REASONS: wrong outcome,wrong study design | Culicidae | PMC10313673 |
| rayyan-651229593 | Minkah NK\|\|Schafer C\|\|Kappe SHI | Humanized Mouse Models for the Study of Human Malaria Parasite Biology, Pathogenesis, and Immunity. | 9 | 807 | RAYYAN-INCLUSION: {"Richard"=>"Excluded", "Jessica "=>"Excluded"} \| RAYYAN-EXCLUSION-REASONS: wrong outcome,wrong study design | Humans;Humanities;Humanism;Mice;Malaria | PMC5917005 |
| rayyan-651229594 | Martin S\|\|Rassi C\|\|Antonio V\|\|Graham K\|\|Leitao J\|\|King R\|\|Jive E | Evaluating the feasibility and acceptability of a community dialogue intervention in the prevention and control of schistosomiasis in Nampula province, Mozambique. | 16 | e0255647 | RAYYAN-INCLUSION: {"Richard"=>"Excluded", "Jessica "=>"Excluded"} \| RAYYAN-EXCLUSION-REASONS: wrong outcome,wrong study design,wrong infection / disease | Schistosomiasis | PMC8341517 |
| rayyan-651229595 | Selemani M\|\|Msengwa AS\|\|Mrema S\|\|Shamte A\|\|Mahande MJ\|\|Yeates K\|\|Mbago MC\|\|Lutambi AM | Assessing the effects of mosquito nets on malaria mortality using a space time model: a case study of Rufiji and Ifakara Health and Demographic Surveillance System sites in rural Tanzania. | 15 | 257 | RAYYAN-INCLUSION: {"Donal"=>"Excluded", "Richard"=>"Excluded", "Jessica "=>"Included"} \| RAYYAN-LABELS: Excluded (tiebreaker) \| RAYYAN-EXCLUSION-REASONS: wrong study duration \| USER-NOTES: {"Richard"=>["analysis period ends in 2012"]} | Culicidae | PMC4857246 |
| rayyan-651229596 | Das AM\|\|Hetzel MW\|\|Yukich JO\|\|Stuck L\|\|Fakih BS\|\|Al-Mafazy AH\|\|Ali A\|\|Chitnis N | The impact of reactive case detection on malaria transmission in Zanzibar in the presence of human mobility. | 41 | 100639 | RAYYAN-INCLUSION: {"Richard"=>"Excluded", "Jessica "=>"Excluded"} \| RAYYAN-EXCLUSION-REASONS: wrong outcome,wrong study design | Humanities;Humanism;Humans;Malaria | PMC9758615 |
| rayyan-651229597 | Santos-Vega M\|\|Lowe R\|\|Anselin L\|\|Desai V\|\|Vaishnav KG\|\|Naik A\|\|Pascual M | Quantifying climatic and socioeconomic drivers of urban malaria in Surat, India: a statistical spatiotemporal modelling study. | 7 | e985-e998 | RAYYAN-INCLUSION: {"Richard"=>"Excluded", "Jessica "=>"Excluded"} \| RAYYAN-EXCLUSION-REASONS: wrong outcome,wrong study design | India;Malaria | |
| rayyan-651229598 | Chirombo J\|\|Ceccato P\|\|Lowe R\|\|Terlouw DJ\|\|Thomson MC\|\|Gumbo A\|\|Diggle PJ\|\|Read JM | Childhood malaria case incidence in Malawi between 2004 and 2017: spatio-temporal modelling of climate and non-climate factors. | 19 | 5 | RAYYAN-INCLUSION: {"Donal"=>"Included", "Richard"=>"Included", "Jessica "=>"Excluded"} \| RAYYAN-LABELS: Include (tiebreaker) \| USER-NOTES: {"Richard"=>["Unclear if intervention effect measured"]} | | PMC6945411 |
| rayyan-651229599 | Rassi C\|\|Martin S\|\|Graham K\|\|de Cola MA\|\|Christiansen-Jucht C\|\|Smith LE\|\|Jive E\|\|Phillips AE\|\|New... | Knowledge, attitudes and practices with regard to schistosomiasis prevention and control: Two cross-sectional household surveys before and after a Community Dialogue intervention in Nampula province, Mozambique. | 13 | e0007138 | RAYYAN-INCLUSION: {"Richard"=>"Excluded", "Jessica "=>"Excluded"} \| RAYYAN-EXCLUSION-REASONS: wrong study design | Schistosomiasis;Cesarean Section | PMC6382216 |
| rayyan-651229600 | Aheto JMK | Mapping under-five child malaria risk that accounts for environmental and climatic factors to aid malaria preventive and control efforts in Ghana: Bayesian geospatial and interactive web-based mapping methods. | 21 | 384 | RAYYAN-INCLUSION: {"Richard"=>"Excluded", "Jessica "=>"Excluded"} \| RAYYAN-EXCLUSION-REASONS: wrong study design \| USER-NOTES: {"Richard"=>["Model exercise"]} | Malaria | PMC9756577 |
| rayyan-651229601 | Ralaidovy AH\|\|Lauer JA\|\|Pretorius C\|\|Briet OJ\|\|Patouillard E | Priority Setting in HIV, Tuberculosis, and Malaria - New Cost-Effectiveness Results From WHO-CHOICE. | 10 | 678-696 | RAYYAN-INCLUSION: {"Richard"=>"Excluded", "Jessica "=>"Excluded"} \| RAYYAN-EXCLUSION-REASONS: wrong outcome,wrong study design | Cost-Benefit Analysis;Malaria | PMC9278379 |
| rayyan-651229602 | Iessa N\|\|Macolic Sarinic V\|\|Ghazaryan L\|\|Romanova N\|\|Alemu A\|\|Rungapiromnan W\|\|Jiamsuchon P\|\|Pokh... | Smart Safety Surveillance (3S): Multi-Country Experience of Implementing the 3S Concepts and Principles. | 44 | 1085-1098 | RAYYAN-INCLUSION: {"Richard"=>"Excluded", "Jessica "=>"Excluded"} \| RAYYAN-EXCLUSION-REASONS: wrong outcome,wrong study design | | PMC8325038 |
| rayyan-651229603 | Hopkins H\|\|Bruxvoort KJ\|\|Cairns ME\|\|Chandler CI\|\|Leurent B\|\|Ansah EK\|\|Baiden F\|\|Baltzell KA\|\|Bjor... | Impact of introduction of rapid diagnostic tests for malaria on antibiotic prescribing: analysis of observational and randomised studies in public and private healthcare settings. | 356 | j1054 | RAYYAN-INCLUSION: {"Richard"=>"Excluded", "Jessica "=>"Excluded"} \| RAYYAN-EXCLUSION-REASONS: background article,wrong study design,wrong publication type | Anti-Bacterial Agents;Malaria | PMC5370398 |
| rayyan-651229604 | Thomas R\|\|Cirera L\|\|Brew J\|\|Saute F\|\|Sicuri E | The short-term impact of a malaria elimination initiative in Southern Mozambique: Application of the synthetic control method to routine surveillance data. | 30 | 2168-2184 | RAYYAN-INCLUSION: {"Donal"=>"Included", "Richard"=>"Included", "Jessica "=>"Maybe"} \| RAYYAN-LABELS: Include (tiebreaker) | Malaria | |
| rayyan-651229605 | Carrel M\|\|Kim S\|\|Mwandagalirwa MK\|\|Mvuama N\|\|Bala JA\|\|Nkalani M\|\|Kihuma G\|\|Atibu J\|\|Diallo AO\|\|Go... | Individual, household and neighborhood risk factors for malaria in the Democratic Republic of the Congo support new approaches to programmatic intervention. | 70 | 102581 | RAYYAN-INCLUSION: {"Richard"=>"Excluded", "Jessica "=>"Excluded"} \| RAYYAN-EXCLUSION-REASONS: wrong study design | Risk Factors | PMC8328915 |
| rayyan-651229606 | Anderson CM\|\|Eloff I\|\|Kruger T | Could a culturally and age-appropriate song contribute towards malaria prevention in primary school learners? | 21 | 113 | RAYYAN-INCLUSION: {"Richard"=>"Excluded", "Jessica "=>"Excluded"} \| RAYYAN-EXCLUSION-REASONS: wrong study design | Malaria;Music;Primary Prevention | PMC8976955 |
| rayyan-651229607 | Kamau A\|\|Musau M\|\|Mwakio S\|\|Amadi D\|\|Nyaguara A\|\|Bejon P\|\|Seale AC\|\|Berkley JA\|\|Snow RW | Impact of Intermittent Presumptive Treatment for Malaria in Pregnancy on Hospital Birth Outcomes on the Kenyan Coast. | 76 | e875-e883 | RAYYAN-INCLUSION: {"Richard"=>"Included", "Jessica "=>"Included"} | Pregnancy;Malaria | PMC9907553 |
| rayyan-651229608 | Folegatti PM\|\|Siqueira AM\|\|Monteiro WM\|\|Lacerda MV\|\|Drakeley CJ\|\|Braga EM | A systematic review on malaria sero-epidemiology studies in the Brazilian Amazon: insights into immunological markers for exposure and protection. | 16 | 107 | RAYYAN-INCLUSION: {"Richard"=>"Excluded", "Jessica "=>"Excluded"} \| RAYYAN-EXCLUSION-REASONS: background article,wrong study design,wrong publication type | Biological Markers;Malaria | PMC5341168 |
| rayyan-651229609 | Wagman J\|\|Cisse I\|\|Kone D\|\|Fomba S\|\|Eckert E\|\|Mihigo J\|\|Bankineza E\|\|Bah M\|\|Diallo D\|\|Gogue C\|\|Ty... | Combining next-generation indoor residual spraying and drug-based malaria control strategies: observational evidence of a combined effect in Mali. | 19 | 293 | RAYYAN-INCLUSION: {"Richard"=>"Included", "Jessica "=>"Included"} | Malaria | PMC7429948 |
| rayyan-651229610 | Hutchinson E\|\|Nayiga S\|\|Nabirye C\|\|Taaka L\|\|Westercamp N\|\|Rowe AK\|\|Staedke SG | Opening the 'black box' of collaborative improvement: a qualitative evaluation of a pilot intervention to improve quality of malaria surveillance data in public health centres in Uganda. | 20 | 289 | RAYYAN-INCLUSION: {"Richard"=>"Excluded", "Jessica "=>"Excluded"} \| RAYYAN-EXCLUSION-REASONS: wrong study design | Uganda;African Continental Ancestry Group;Malaria | PMC8243860 |
| rayyan-651229611 | Ashton RA\|\|Bennett A\|\|Al-Mafazy AW\|\|Abass AK\|\|Msellem MI\|\|McElroy P\|\|Kachur SP\|\|Ali AS\|\|Yukich J\|... | Use of Routine Health Information System Data to Evaluate Impact of Malaria Control Interventions in Zanzibar, Tanzania from 2000 to 2015. | 12 | 19-Nov | RAYYAN-INCLUSION: {"Richard"=>"Included", "Jessica "=>"Included"} | Information Systems;Malaria | PMC6677660 |

| | | | | | | | |
|---|---|---|---|---|---|---|---|
| rayyan-651229612 | Green C\|\|Quigley P\|\|Kureya T\|\|Barber C\|\|Chizema E\|\|Moonga H\|\|Chanda E\|\|Simfukwe V\|\|Mpande B\|\|Simu... | Use of rectal artesunate for severe malaria at the community level, Zambia. | 97 | 810-817 | RAYYAN-INCLUSION: {"Richard"=>"Included", "Jessica "=>"Included"} | Malaria | PMC6883271 |
| rayyan-651229613 | Stanton MC\|\|Mkwanda S\|\|Mzilahowa T\|\|Bockarie MJ\|\|Kelly-Hope LA | Quantifying filariasis and malaria control activities in relation to lymphatic filariasis elimination: a multiple intervention score map (MISM) for Malawi. | 19 | 224-35 | RAYYAN-INCLUSION: {"Donal"=>"Excluded", "Richard"=>"Excluded", "Jessica "=>"Maybe"} \| RAYYAN-LABELS: Excluded (tiebreaker) \| RAYYAN-EXCLUSION-REASONS: wrong infection / disease | Malaria;Filariasis | |
| rayyan-651229614 | Pradhan S\|\|Pradhan MM\|\|Dutta A\|\|Shah NK\|\|Joshi PL\|\|Pradhan K\|\|Sharma SK\|\|Grewal Daumerie P\|\|Baner... | Improved access to early diagnosis and complete treatment of malaria in Odisha, India. | 14 | e0208943 | RAYYAN-INCLUSION: {"Donal"=>"Included", "Richard"=>"Included", "Jessica "=>"Excluded"} \| RAYYAN-LABELS: Include (tiebreaker) | India;Malaria | PMC6314604 |
| rayyan-651229615 | Holmes KK\|\|Bertozzi S\|\|Bloom BR\|\|Jha P\|\|Gelband H\|\|DeMaria LM\|\|Horton S | Major Infectious Diseases: Key Messages from Disease Control Priorities, Third Edition. | | | RAYYAN-INCLUSION: {"Richard"=>"Excluded", "Jessica "=>"Excluded"} \| RAYYAN-EXCLUSION-REASONS: background article,wrong study design,wrong publication type | Communicable Diseases | |
| rayyan-651229616 | Homan T\|\|di Pasquale A\|\|Onoka K\|\|Kiche I\|\|Hiscox A\|\|Mweresa C\|\|Mukabana WR\|\|Masiga D\|\|Takken W\|\|M... | Profile: The Rusinga Health and Demographic Surveillance System, Western Kenya. | 45 | 718-27 | RAYYAN-INCLUSION: {"Richard"=>"Excluded", "Jessica "=>"Excluded"} \| RAYYAN-EXCLUSION-REASONS: wrong outcome,wrong study design,wrong publication type | Kenya | |
| rayyan-651229617 | Ashton RA\|\|Prosnitz D\|\|Andrada A\|\|Herrera S\|\|Ye Y | Evaluating malaria programmes in moderate- and low-transmission settings: practical ways to generate robust evidence. | 19 | 75 | RAYYAN-INCLUSION: {"Richard"=>"Excluded", "Jessica "=>"Excluded"} \| RAYYAN-EXCLUSION-REASONS: background article,wrong study design,wrong publication type | Malaria | PMC7027277 |
| rayyan-651229618 | Chang MA\|\|Impoinvil D\|\|Hamre KES\|\|Dalexis PE\|\|Merilien JB\|\|Dismer AM\|\|Fouche B\|\|Desir L\|\|Holmes K... | Acceptability, Feasibility, Drug Safety, and Effectiveness of a Pilot Mass Drug Administration with a Single Round of Sulfadoxine-Pyrimethamine Plus Primaquine and Indoor Residual Spraying in Communities with Malaria Transmission in Haiti, 2018. | 108 | 1127-1139 | RAYYAN-INCLUSION: {"Richard"=>"Excluded", "Jessica "=>"Excluded"} \| RAYYAN-EXCLUSION-REASONS: wrong study design \| USER-NOTES: {"Richard"=>["Unclear if routine data used", "Pre/post cross-sectional surveys"]} | Sulfadoxine;Pyrimethamine;Malaria | PMC10540127 |
| rayyan-651229619 | Soumare HM\|\|Dabira ED\|\|Camara MM\|\|Jadama L\|\|Gaye PM\|\|Kanteh S\|\|Jawara EA\|\|Njie AK\|\|Sanneh F\|\|Ndia... | Entomological impact of mass administration of ivermectin and dihydroartemisinin-piperaquine in The Gambia: a cluster-randomized controlled trial. | 15 | 435 | RAYYAN-INCLUSION: {"Richard"=>"Excluded", "Jessica "=>"Excluded"} \| RAYYAN-EXCLUSION-REASONS: wrong outcome,wrong study design | | PMC9673448 |
| rayyan-651229620 | Daniels RF\|\|Rice BL\|\|Daniels NM\|\|Volkman SK\|\|Hartl DL | The utility of genomic data for Plasmodium vivax population surveillance. | 109 | 153-61 | RAYYAN-INCLUSION: {"Richard"=>"Excluded", "Jessica "=>"Excluded"} \| RAYYAN-EXCLUSION-REASONS: wrong outcome,wrong study design | Population Surveillance;Genome;Plasmodium;Genomics | PMC4455356 |
| rayyan-651229621 | Tewara MA\|\|Mbah-Fongkimeh PN\|\|Dayimu A\|\|Kang F\|\|Xue F | Small-area spatial statistical analysis of malaria clusters and hotspots in Cameroon;2000-2015. | 18 | 636 | RAYYAN-INCLUSION: {"Richard"=>"Excluded", "Jessica "=>"Excluded"} \| RAYYAN-EXCLUSION-REASONS: wrong outcome,wrong study design \| USER-NOTES: {"Richard"=>["Epi study"]} | Cameroon;Malaria | PMC6286522 |
| rayyan-651229622 | Tegegne E\|\|Alemu Gelaye K\|\|Dessie A\|\|Shimelash A\|\|Asmare B\|\|Deml YA\|\|Lamore Y\|\|Temesgen T\|\|Demiss... | Spatio-Temporal Variation of Malaria Incidence and Risk Factors in West Gojjam Zone, Northwest Ethiopia. | 16 | 11786302 22109570 2 | RAYYAN-INCLUSION: {"Richard"=>"Excluded", "Jessica "=>"Excluded"} \| RAYYAN-EXCLUSION-REASONS: wrong outcome,wrong study design \| USER-NOTES: {"Richard"=>["Epi study"]} | Northwestern United States;Risk Factors;Malaria | PMC9087229 |
| rayyan-651229623 | Bal M\|\|Das A\|\|Ghosal J\|\|Pradhan MM\|\|Khuntia HK\|\|Pati S\|\|Dutta A\|\|Ranjit M | Assessment of effectiveness of DAMaN: A malaria intervention program initiated by Government of Odisha, India. | 15 | e0238323 | RAYYAN-INCLUSION: {"Donal"=>"Excluded", "Richard"=>"Excluded", "Jessica "=>"Included"} \| RAYYAN-LABELS: Excluded (tiebreaker) \| RAYYAN-EXCLUSION-REASONS: wrong publication type \| USER-NOTES: {"Richard"=>["Protocol"]} | India;Malaria | PMC7478908 |
| rayyan-651229624 | Longbottom J\|\|Krause A\|\|Torr SJ\|\|Stanton MC | Quantifying geographic accessibility to improve efficiency of entomological monitoring. | 14 | e0008096 | RAYYAN-INCLUSION: {"Richard"=>"Excluded", "Jessica "=>"Excluded"} \| RAYYAN-EXCLUSION-REASONS: background article,wrong study design,wrong infection / disease | | PMC7117774 |
| rayyan-651229625 | Karim A\|\|de Savigny D\|\|Awor P\|\|Cobos Munoz D\|\|Mausezahl D\|\|Kitoto Tshefu A\|\|Ngaima JS\|\|Enebeli U\|... | The building blocks of community health systems: a systems framework for the design, implementation and evaluation of iCCM programs and community-based interventions. | 7 | | RAYYAN-INCLUSION: {"Richard"=>"Excluded", "Jessica "=>"Excluded"} \| RAYYAN-EXCLUSION-REASONS: wrong outcome,wrong study design | | PMC9247653 |
| rayyan-651229626 | Mphwatiwa T\|\|Witek-McManus S\|\|Mtali A\|\|Okello G\|\|Nguluwe P\|\|Chatsika H\|\|Roschnik N\|\|Halliday KE\|\|... | School-based diagnosis and treatment of malaria by teachers using rapid diagnostic tests and artemisinin-based combination therapy: experiences and perceptions of users and implementers of the Learner Treatment Kit, southern Malawi. | 16 | 318 | RAYYAN-INCLUSION: {"Richard"=>"Excluded", "Jessica "=>"Excluded"} \| RAYYAN-EXCLUSION-REASONS: wrong outcome,wrong study design | | PMC5547577 |
| rayyan-651229627 | Lana R\|\|Nekkab N\|\|Siqueira AM\|\|Peterka C\|\|Marchesini P\|\|Lacerda M\|\|Mueller I\|\|White M\|\|Villela D | The top 1%: quantifying the unequal distribution of malaria in Brazil. | 20 | 87 | RAYYAN-INCLUSION: {"Richard"=>"Excluded", "Jessica "=>"Excluded"} \| RAYYAN-EXCLUSION-REASONS: wrong outcome,wrong study design | Brazil;Malaria | PMC7880522 |
| rayyan-651229628 | Levitz L\|\|Janko M\|\|Mwandagalirwa K\|\|Thwai KL\|\|Likwela JL\|\|Tshefu AK\|\|Emch M\|\|Meshnick SR | Effect of individual and community-level bed net usage on malaria prevalence among under-fives in the Democratic Republic of Congo. | 17 | 39 | RAYYAN-INCLUSION: {"Donal"=>"Excluded", "Richard"=>"Excluded", "Jessica "=>"Included"} \| RAYYAN-LABELS: Excluded (tiebreaker) \| RAYYAN-EXCLUSION-REASONS: wrong study design,wrong study duration \| USER-NOTES: {"Richard"=>["Nationally representative survey (DHS)"]} | Prevalence;Malaria | PMC5774036 |
| rayyan-651229629 | Liu H\|\|Zhou Y\|\|Deng Y\|\|Lin Z\|\|Zhang C\|\|Chen Q\|\|Wei C\|\|Duan K\|\|Tian P\|\|Zhou H\|\|Xu J | Malaria from hyperendemicity to elimination along international borders in Yunnan, China during 2003â€'2020: a case study. | 11 | 51 | RAYYAN-INCLUSION: {"Donal"=>"Excluded", "Richard"=>"Excluded", "Jessica "=>"Maybe"} \| RAYYAN-LABELS: Excluded (tiebreaker) \| RAYYAN-EXCLUSION-REASONS: wrong outcome,wrong study design | China;Malaria | PMC9088148 |
| rayyan-651229630 | Mukhtar AYA\|\|Munyakazi JB\|\|Ouifki R | Assessing the role of human mobility on malaria transmission. | 320 | 108304 | RAYYAN-INCLUSION: {"Richard"=>"Excluded", "Jessica "=>"Excluded"} \| RAYYAN-EXCLUSION-REASONS: wrong outcome,wrong study design | Humanities;Humanism;Humans;Malaria | |
| rayyan-651229631 | Mlacha YP\|\|Wang D\|\|Chaki PP\|\|Gavana T\|\|Zhou Z\|\|Michael MG\|\|Khatib R\|\|Chila G\|\|Msuya HM\|\|Chaki E\|\|... | Effectiveness of the innovative 1,7-malaria reactive community-based testing and response (1, 7-mRCTR) approach on malaria burden reduction in Southeastern Tanzania. | 19 | 292 | RAYYAN-INCLUSION: {"Donal"=>"Included", "Richard"=>"Included", "Jessica "=>"Excluded"} \| RAYYAN-LABELS: Include (tiebreaker) | Malaria | PMC7429894 |
| rayyan-651229632 | Medeiros DN\|\|Ferranti JF\|\|Delgado AF\|\|de Carvalho WB | Colloids for the Initial Management of Severe Sepsis and Septic Shock in Pediatric Patients: A Systematic Review. | 31 | e11-6 | RAYYAN-INCLUSION: {"Richard"=>"Excluded", "Jessica "=>"Excluded"} \| RAYYAN-EXCLUSION-REASONS: wrong outcome,wrong study design,wrong publication type | Sepsis | |
| rayyan-651229633 | Vavilala H\|\|Yaladanda N\|\|Krishna Kondeti P\|\|Rafiq Unissa\|\|Mopuri R\|\|Gouda KC\|\|Rao Bhimala K\|\|Rao ... | Weather integrated malaria prediction system using Bayesian structural time series model for northeast states of India. | 29 | 68232-68246 | RAYYAN-INCLUSION: {"Richard"=>"Excluded", "Jessica "=>"Excluded"} \| RAYYAN-EXCLUSION-REASONS: wrong outcome,wrong study design | India;Malaria | |
| rayyan-651229634 | Wang X\|\|Zhao XQ | A climate-based malaria model with the use of bed nets. | 77 | 25-Jan | RAYYAN-INCLUSION: {"Richard"=>"Excluded", "Jessica "=>"Excluded"} \| RAYYAN-EXCLUSION-REASONS: wrong outcome,wrong study design | Malaria | |
| rayyan-651229635 | Jongdeepaisal M\|\|Khonputsa P\|\|Prasert O\|\|Maneenet S\|\|Pongsoipetch K\|\|Jatapai A\|\|Rotejanaprasert C... | Forest malaria and prospects for anti-malarial chemoprophylaxis among forest goers: findings from a qualitative study in Thailand. | 21 | 47 | RAYYAN-INCLUSION: {"Richard"=>"Excluded", "Jessica "=>"Excluded"} \| RAYYAN-EXCLUSION-REASONS: wrong outcome,wrong study design | Antimalarials;Thailand;Chemoprevention | PMC8845363 |
| rayyan-651229636 | Camponovo F\|\|Ockenhouse CF\|\|Lee C\|\|Penny MA | Mass campaigns combining antimalarial drugs and anti-infective vaccines as seasonal interventions for malaria control, elimination and prevention of resurgence: a modelling study. | 19 | 920 | RAYYAN-INCLUSION: {"Richard"=>"Excluded", "Jessica "=>"Excluded"} \| RAYYAN-EXCLUSION-REASONS: wrong study design | Antimalarials;Vaccination;Malaria;Seasons | PMC6820916 |
| rayyan-651229637 | Awine T\|\|Silal SP | Accounting for regional transmission variability and the impact of malaria control interventions in Ghana: a population level mathematical modelling approach. | 19 | 423 | RAYYAN-INCLUSION: {"Donal"=>"Included", "Richard"=>"Included", "Jessica "=>"Excluded"} \| RAYYAN-LABELS: Include (tiebreaker) | Mathematics;Ghana;Malaria | PMC7684904 |
| rayyan-651229638 | Staedke SG\|\|Kamya MR\|\|Dorsey G\|\|Maiteki-Sebuguzi C\|\|Gonahasa S\|\|Yeka A\|\|Lynd A\|\|Opigo J\|\|Hemingwa... | LLIN Evaluation in Uganda Project (LLINEUP) - Impact of long-lasting insecticidal nets with, and without, piperonyl butoxide on malaria indicators in Uganda: study protocol for a cluster-randomised trial. | 20 | 321 | RAYYAN-INCLUSION: {"Donal"=>"Excluded", "Richard"=>"Excluded", "Jessica "=>"Excluded"} \| USER-NOTES: {"Richard"=>["Protocol. No routine data, but innovative trial design"]} | Uganda;Malaria | PMC6547536 |
| rayyan-651229639 | Hill J\|\|Ouma P\|\|Oluoch S\|\|Bruce J\|\|Kariuki S\|\|Desai M\|\|Webster J | Intermittent screening and treatment with dihydroartemisinin-piperaquine for the prevention of malaria in pregnancy: implementation feasibility in a routine healthcare system setting in western Kenya. | 19 | 433 | RAYYAN-INCLUSION: {"Richard"=>"Excluded", "Jessica "=>"Excluded"} \| RAYYAN-EXCLUSION-REASONS: wrong outcome,wrong study design \| USER-NOTES: {"Richard"=>["Intervention and routine data unclear"]} | Pregnancy;Kenya;Mass Screening;Malaria | PMC7690090 |
| rayyan-651229640 | Young N\|\|Achieng F\|\|Desai M\|\|Phillips-Howard P\|\|Hill J\|\|Aol G\|\|Bigogo G\|\|Laserson K\|\|Ter Kuile F\|... | Integrated point-of-care testing (POCT) for HIV, syphilis, malaria and anaemia at antenatal facilities in western Kenya: a qualitative study exploring end-users' perspectives of appropriateness, acceptability and feasibility. | 19 | 74 | RAYYAN-INCLUSION: {"Richard"=>"Excluded", "Jessica "=>"Excluded"} \| RAYYAN-EXCLUSION-REASONS: wrong outcome,wrong study design,wrong infection / disease | Kenya;Malaria | PMC6348645 |
| rayyan-651229641 | Oliff MS\|\|Muniina P\|\|Babigumira K\|\|Phuka J\|\|Rietveld H\|\|Sande J\|\|Nsona H\|\|Lugand MM | The five continuum of care criteria that should accompany rectal artesunate interventions: lessons learned from an implementation study in Malawi. | 22 | 108 | RAYYAN-INCLUSION: {"Richard"=>"Excluded", "Jessica "=>"Excluded"} \| RAYYAN-EXCLUSION-REASONS: wrong outcome,wrong study design | Continuity of Patient Care | PMC10039536 |
| rayyan-651229642 | Waiswa P\|\|Mpanga F\|\|Bagenda D\|\|Kananura RM\|\|O'Connell T\|\|Henriksson DK\|\|Diaz T\|\|Ayebare F\|\|Kataho... | Child health and the implementation of Community and District-management Empowerment for Scale-up (CODES) in Uganda: a randomised controlled trial. | 6 | | RAYYAN-INCLUSION: {"Richard"=>"Excluded", "Jessica "=>"Excluded"} \| RAYYAN-EXCLUSION-REASONS: wrong outcome,wrong study design | Child Welfare;Uganda;Only Child;Child | PMC8189926 |
| rayyan-651229643 | Moukam Kakmeni FM\|\|Guimapi RYA\|\|Ndjomatchoua FT\|\|Pedro SA\|\|Mutunga J\|\|Tonnang HEZ | Spatial panorama of malaria prevalence in Africa under climate change and interventions scenarios. | 17 | 2 | RAYYAN-INCLUSION: {"Richard"=>"Excluded", "Jessica "=>"Excluded"} \| RAYYAN-EXCLUSION-REASONS: wrong outcome,wrong study design | Prevalence;Malaria | PMC5771136 |

| | | | | | | | |
|---|---|---|---|---|---|---|---|
| rayyan-651229644 | Lesego A\|\|Tsegaye T\|\|Were LPO\|\|Sakvarelidze G\|\|Garg S\|\|Morrison L\|\|Nigussie S\|\|Githendu P\|\|Achoki T | Assessment of the Global Fund-supported procurement and supply chain reforms at the Ethiopian Pharmaceuticals Supply Agency: a mixed-methods study. | 13 | e073390 | RAYYAN-INCLUSION: {"Richard"=>"Excluded", "Jessica "=>"Excluded"} \| RAYYAN-EXCLUSION-REASONS: wrong outcome,wrong study design | | PMC10729206 |
| rayyan-651229645 | Chung AM\|\|Case P\|\|Gosling J\|\|Gosling R\|\|Madinga M\|\|Chikodzore R\|\|Hove M\|\|Viljoen G\|\|Chitapi P\|\|Gu... | Scaling up malaria elimination management and leadership: a pilot in three provinces in Zimbabwe, 2016-2018. | 19 | 185 | RAYYAN-INCLUSION: {"Richard"=>"Excluded", "Jessica "=>"Excluded"} \| RAYYAN-EXCLUSION-REASONS: wrong outcome,wrong study design | Malaria | PMC7238623 |
| rayyan-651229646 | Ghansah A\|\|Tiedje KE\|\|Argyropoulos DC\|\|Onwona CO\|\|Deed SL\|\|Labbe F\|\|Oduro AR\|\|Koram KA\|\|Pascual M... | Comparison of molecular surveillance methods to assess changes in the population genetics of Plasmodium falciparum in high transmission. | 2 | | RAYYAN-INCLUSION: {"Richard"=>"Excluded", "Jessica "=>"Excluded"} \| RAYYAN-EXCLUSION-REASONS: wrong outcome,wrong study design | Plasmodium falciparum | PMC10686283 |
| rayyan-651229647 | Sharma PK\|\|Tilak R | Outbreak prone communicable diseases of public health importance in the northern districts of West Bengal - Current status & the way forward. | 153 | 358-366 | RAYYAN-INCLUSION: {"Richard"=>"Excluded", "Jessica "=>"Excluded"} \| RAYYAN-EXCLUSION-REASONS: wrong outcome,wrong study design,wrong infection / disease | Disease Outbreaks;Communicable Diseases | PMC8204839 |
| rayyan-651229648 | Dambach P\|\|Traore I\|\|Becker N\|\|Kaiser A\|\|Sie A\|\|Sauerborn R | EMIRA: Ecologic Malaria Reduction for Africa--innovative tools for integrated malaria control. | 7 | 25908 | RAYYAN-INCLUSION: {"Richard"=>"Excluded", "Jessica "=>"Excluded"} \| RAYYAN-EXCLUSION-REASONS: wrong publication type | Malaria;Africa | PMC4223283 |
| rayyan-651229649 | Liu J\|\|Wu X\|\|Li C\|\|Zhou S | Decline in malaria incidence in a typical county of China: Role of climate variance and anti-malaria intervention measures. | 167 | 276-282 | RAYYAN-INCLUSION: {"Richard"=>"Excluded", "Jessica "=>"Excluded"} \| RAYYAN-EXCLUSION-REASONS: wrong outcome,wrong study design | Malaria;China | |
| rayyan-651229650 | Wang T\|\|Chen Z\|\|Chen H\|\|Yu X\|\|Wang L\|\|Liu X | Brusatol inhibits the growth of renal cell carcinoma by regulating the PTEN/PI3K/AKT pathway. | 288 | 115020 | RAYYAN-INCLUSION: {"Richard"=>"Excluded", "Jessica "=>"Excluded"} \| RAYYAN-EXCLUSION-REASONS: wrong outcome,wrong study design,wrong infection / disease | Carcinoma, Renal Cell | |
| rayyan-651229651 | Singh MP\|\|Rajvanshi H\|\|Bharti PK\|\|Jayswar H\|\|Singh S\|\|Mehra RK\|\|Pandey M\|\|Sahu RS\|\|Patel B\|\|Bhala... | Evaluation of the model malaria elimination strategy in Mandla district along with its neighbouring districts: a time series analysis from 2008 to 2020. | 22 | 45 | RAYYAN-INCLUSION: {"Donal"=>"Excluded", "Richard"=>"Excluded", "Jessica "=>"Maybe"} \| RAYYAN-LABELS: Excluded (tiebreaker) \| RAYYAN-EXCLUSION-REASONS: wrong outcome,wrong study design | Malaria | PMC9901400 |
| rayyan-651229652 | Some JW\|\|Abbeddou S\|\|Yakes Jimenez E\|\|Hess SY\|\|Ouedraogo ZP\|\|Guissou RM\|\|Vosti SA\|\|Ouedraogo JB\|\|... | Effect of zinc added to a daily small-quantity lipid-based nutrient supplement on diarrhoea, malaria, fever and respiratory infections in young children in rural Burkina Faso: a cluster-randomised trial. | 5 | e007828 | RAYYAN-INCLUSION: {"Richard"=>"Excluded", "Jessica "=>"Excluded"} \| RAYYAN-EXCLUSION-REASONS: wrong outcome,wrong study design,wrong infection / disease | Respiratory Tract Infections;Q Fever;Zinc;Fever;Malaria | PMC4567679 |
| rayyan-651229653 | Cepeda JA\|\|Eritsyan K\|\|Vickerman P\|\|Lyubimova A\|\|Shegay M\|\|Odinokova V\|\|Beletsky L\|\|Borquez A\|\|Hi... | Potential impact of implementing and scaling up harm reduction and antiretroviral therapy on HIV prevalence and mortality and overdose deaths among people who inject drugs in two Russian cities: a modelling study. | 5 | e578-e587 | RAYYAN-INCLUSION: {"Richard"=>"Excluded", "Jessica "=>"Excluded"} \| RAYYAN-EXCLUSION-REASONS: wrong outcome,wrong study design,wrong infection / disease | Prevalence | PMC6188805 |
| rayyan-651229654 | Zhou G\|\|Lo E\|\|Zhong D\|\|Wang X\|\|Wang Y\|\|Malla S\|\|Lee MC\|\|Yang Z\|\|Cui L\|\|Yan G | Impact of interventions on malaria in internally displaced persons along the China-Myanmar border: 2011 2014. | 15 | 471 | RAYYAN-INCLUSION: {"Donal"=>"Excluded", "Richard"=>"Maybe", "Jessica "=>"Maybe"} \| RAYYAN-LABELS: Excluded (tiebreaker) \| USER-NOTES: {"Richard"=>["Unclear re interventions and routine data"]} | China;Malaria | PMC5024476 |
| rayyan-651229655 | Bahia AC\|\|Dong Y\|\|Blumberg BJ\|\|Mlambo G\|\|Tripathi A\|\|BenMarzouk-Hidalgo OJ\|\|Chandra R\|\|Dimopoulos G | Exploring Anopheles gut bacteria for Plasmodium blocking activity. | 16 | 2980-94 | RAYYAN-INCLUSION: {"Richard"=>"Excluded", "Jessica "=>"Excluded"} \| RAYYAN-EXCLUSION-REASONS: wrong outcome,wrong study design | Bacteria;Plasmodium | PMC4099322 |
| rayyan-651229656 | Canavati SE\|\|Kelly GC\|\|Quintero CE\|\|Vo TH\|\|Tran LK\|\|Ngo TD\|\|Tran DT\|\|Edgel KA\|\|Martin NJ | Targeting high risk forest goers for malaria elimination: a novel approach for investigating forest malaria to inform program intervention in Vietnam. | 20 | 757 | RAYYAN-INCLUSION: {"Richard"=>"Excluded", "Jessica "=>"Excluded"} \| RAYYAN-EXCLUSION-REASONS: wrong outcome,wrong study design | Malaria;Vietnam;Trees | PMC7559775 |
| rayyan-651229657 | Munsey A\|\|Kinyina A\|\|Assenga M\|\|Almeida A\|\|Kitojo C\|\|Reaves E\|\|Simeo J\|\|Aron S\|\|Chacky F\|\|Nhiga S... | Assessing the utility of pregnant women as a sentinel surveillance population for malaria in Geita, Tanzania, 2019 - 2021. | 136 | 57-63 | RAYYAN-INCLUSION: {"Donal"=>"Excluded", "Richard"=>"Maybe", "Jessica "=>"Excluded"} \| RAYYAN-LABELS: Excluded (tiebreaker) | Population Surveillance;Malaria | |
| rayyan-651229658 | Akogbeto MC\|\|Aikpon RY\|\|Azondekon R\|\|Padonou GG\|\|Osse RA\|\|Agossa FR\|\|Beach R\|\|Sezonlin M | Six years of experience in entomological surveillance of indoor residual spraying against malaria transmission in Benin: lessons learned, challenges and outlooks. | 14 | 242 | RAYYAN-INCLUSION: {"Richard"=>"Excluded", "Jessica "=>"Excluded"} \| RAYYAN-EXCLUSION-REASONS: wrong outcome,wrong study design | Malaria;Learning | PMC4465467 |
| rayyan-651229659 | Manning J\|\|Lon C\|\|Spring M\|\|Wojnarski M\|\|Somethy S\|\|Chann S\|\|Gosi P\|\|Soveasna K\|\|Sriwichai S\|\|Kun... | Cluster-randomized trial of monthly malaria prophylaxis versus focused screening and treatment: a study protocol to define malaria elimination strategies in Cambodia. | 19 | 558 | RAYYAN-INCLUSION: {"Richard"=>"Excluded", "Jessica "=>"Excluded"} \| RAYYAN-EXCLUSION-REASONS: wrong publication type | Malaria | PMC6192281 |
| rayyan-651229660 | Odero NA\|\|Samuels AM\|\|Odongo W\|\|Abong'o B\|\|Gimnig J\|\|Otieno K\|\|Odero C\|\|Obor D\|\|Ombok M\|\|Were V\|\|... | Community-based intermittent mass testing and treatment for malaria in an area of high transmission intensity, western Kenya: development of study site infrastructure and lessons learned. | 18 | 255 | RAYYAN-INCLUSION: {"Richard"=>"Excluded", "Jessica "=>"Excluded"} \| RAYYAN-EXCLUSION-REASONS: wrong outcome,wrong study design | Kenya;Malaria | PMC6664589 |
| rayyan-651229661 | Nuwa A\|\|Baker K\|\|Bonnington C\|\|Odongo M\|\|Kyagulanyi T\|\|Bwanika JB\|\|Richardson S\|\|Nabakooza J\|\|Ach... | A non-randomized controlled trial to assess the protective effect of SMC in the context of high parasite resistance in Uganda. | 22 | 63 | RAYYAN-INCLUSION: {"Donal"=>"Excluded", "Richard"=>"Maybe", "Jessica "=>"Excluded"} \| RAYYAN-LABELS: Excluded (tiebreaker) \| USER-NOTES: {"Richard"=>["Passive surveillance of kids in cohort"]} | Uganda | PMC9945593 |
| rayyan-651229663 | Mpimbaza A\|\|Sserwanga A\|\|Rutazaana D\|\|Kapisi J\|\|Walemwa R\|\|Suiyanka L\|\|Kyalo D\|\|Kamya M\|\|Opigo J\|... | Changing malaria fever test positivity among paediatric admissions to Tororo district hospital, Uganda 2012-2019. | 19 | 416 | RAYYAN-INCLUSION: {"Donal"=>"Included", "Richard"=>"Included", "Jessica "=>"Excluded"} \| RAYYAN-LABELS: Include (tiebreaker) | Uganda;Q Fever;Fever;Malaria | PMC7678291 |
| rayyan-651229664 | Massoda Tonye SG\|\|Kouambeng C\|\|Wounang R\|\|Vounatsou P | Challenges of DHS and MIS to capture the entire pattern of malaria parasite risk and intervention effects in countries with different ecological zones: the case of Cameroon. | 17 | 156 | RAYYAN-INCLUSION: {"Richard"=>"Excluded", "Jessica "=>"Excluded"} \| RAYYAN-EXCLUSION-REASONS: wrong study design,wrong study duration | Cameroon;Malaria | PMC5889563 |
| rayyan-651229665 | Xu JW\|\|Li JJ\|\|Guo HP\|\|Pu SW\|\|Li SM\|\|Wang RH\|\|Liu H\|\|Wang WJ | Malaria from hyperendemicity to elimination in Hekou County on China-Vietnam border: an ecological study. | 16 | 66 | RAYYAN-INCLUSION: {"Richard"=>"Excluded", "Jessica "=>"Excluded"} \| RAYYAN-EXCLUSION-REASONS: wrong outcome,wrong study design | China;Vietnam;Malaria | PMC5297092 |
| rayyan-651229666 | Ferriss E\|\|Chaponda M\|\|Muleba M\|\|Kabuya JB\|\|Lupiya JS\|\|Riley C\|\|Winters A\|\|Moulton LH\|\|Mulenga M\|... | The Impact of Household and Community Indoor Residual Spray Coverage with Fludora Fusion in a High Malaria Transmission Setting in Northern Zambia. | 109 | 248-257 | RAYYAN-INCLUSION: {"Richard"=>"Included", "Jessica "=>"Included"} | Malaria | PMC10397455 |
| rayyan-651229667 | Kabeya TK\|\|Kasongo JCM\|\|Matumba NB\|\|Tshibangu DI\|\|Garcia-Morzon LA\|\|Burgueno E | Impact of mass distribution of long-lasting insecticide nets on the incidence of malaria in Lomami, Democratic Republic of Congo: a study based on electronic health record data (2018 - 2019). | 45 | 89 | RAYYAN-INCLUSION: {"Richard"=>"Included", "Jessica "=>"Included"} | Malaria;Insecticides | PMC10474805 |
| rayyan-651229668 | Awine T\|\|Silal SP | Assessing the effectiveness of malaria interventions at the regional level in Ghana using a mathematical modelling application. | 2 | e0000474 | RAYYAN-INCLUSION: {"Richard"=>"Excluded", "Jessica "=>"Excluded"} \| RAYYAN-EXCLUSION-REASONS: wrong study design | Mathematics;Malaria;Ghana | PMC10021332 |
| rayyan-651229669 | Mkali HR\|\|Reaves EJ\|\|Lalji SM\|\|Al-Mafazy AW\|\|Joseph JJ\|\|Ali AS\|\|Abbas FB\|\|Ali MH\|\|Hassan WS\|\|Kito... | Risk factors associated with malaria infection identified through reactive case detection in Zanzibar, 2012-2019. | 20 | 485 | RAYYAN-INCLUSION: {"Donal"=>"Included", "Richard"=>"Included", "Jessica "=>"Excluded"} \| RAYYAN-LABELS: Include (tiebreaker) | Risk Factors;Malaria | PMC8710018 |
| rayyan-651229670 | Fan Y\|\|Shen S\|\|Wei G\|\|Tang J\|\|Zhao Y\|\|Wang F\|\|He X\|\|Guo G\|\|Shang X\|\|Yu X\|\|Ma Z\|\|He X\|\|Liu M\|\|Zhu ... | Rrp6 Regulates Heterochromatic Gene Silencing via ncRNA RUF6 Decay in Malaria Parasites. | 11 | | RAYYAN-INCLUSION: {"Richard"=>"Excluded", "Jessica "=>"Excluded"} \| RAYYAN-EXCLUSION-REASONS: wrong outcome,wrong study design | | PMC7267889 |
| rayyan-651229671 | Smith Gueye C\|\|Newby G\|\|Gosling RD\|\|Whittaker MA\|\|Chandramohan D\|\|Slutsker L\|\|Tanner M | Strategies and approaches to vector control in nine malaria-eliminating countries: a cross-case study analysis. | 15 | 2 | RAYYAN-INCLUSION: {"Richard"=>"Excluded", "Jessica "=>"Excluded"} \| RAYYAN-EXCLUSION-REASONS: wrong outcome,background article,wrong study design,wrong publication type | Malaria | PMC4700736 |
| rayyan-651229672 | Nyanza EC\|\|Kapesa A\|\|Maduka T\|\|Madullu MT | Disability inclusion in malaria services in western Tanzania: A rapid barrier analysis. | 12 | 1270 | RAYYAN-INCLUSION: {"Richard"=>"Excluded", "Jessica "=>"Excluded"} \| RAYYAN-EXCLUSION-REASONS: wrong outcome,wrong study design | Malaria | PMC10696584 |
| rayyan-651229673 | Kassegne K\|\|Fei SW\|\|Ananou K\|\|Noussougnon KS\|\|Komi Koukoura K\|\|Abe EM\|\|Guo XK\|\|Chen JH\|\|Zhou XN | A Molecular Investigation of Malaria Infections From High-Transmission Areas of Southern Togo Reveals Different Species of Plasmodium Parasites. | 12 | 732923 | RAYYAN-INCLUSION: {"Richard"=>"Excluded", "Jessica "=>"Excluded"} \| RAYYAN-EXCLUSION-REASONS: wrong outcome,wrong study design | Plasmodium;Malaria | PMC8674532 |
| rayyan-651229674 | Lu G\|\|Liu Y\|\|Wang J\|\|Li X\|\|Liu X\|\|Beiersmann C\|\|Feng Y\|\|Cao J\|\|Muller O | Malaria training for community health workers in the setting of elimination: a qualitative study from China. | 17 | 95 | RAYYAN-INCLUSION: {"Richard"=>"Excluded", "Jessica "=>"Excluded"} \| RAYYAN-EXCLUSION-REASONS: wrong outcome,wrong study design | China;Malaria | PMC5824442 |
| rayyan-651229675 | Yasuoka J\|\|Jimba M\|\|Levins R | Application of loop analysis for evaluation of malaria control interventions. | 13 | 140 | RAYYAN-INCLUSION: {"Richard"=>"Excluded", "Jessica "=>"Excluded"} \| RAYYAN-EXCLUSION-REASONS: wrong outcome,wrong study design | Malaria | PMC4017832 |
| rayyan-651229676 | Ajayi IO\|\|Jegede AS\|\|Ogunwale AO\|\|Ogundairo J\|\|Olaleye OS\|\|Oshiname FO\|\|Falade CO | Community acceptance, satisfaction, and support for case management of malaria of various degrees in selected rural communities in Ibadan, Oyo-State. | 55 | 198-205 | RAYYAN-INCLUSION: {"Richard"=>"Excluded", "Jessica "=>"Excluded"} \| RAYYAN-EXCLUSION-REASONS: wrong outcome,wrong study design | Rural Population;Case Management;Malaria | PMC9334944 |

<table>
<tr><td>rayyan-651229677</td><td>Ding XY||Mao WH||Lu W||Yu H||Liu Q||Lu P||Jiang H||Zhang X||Lu F||Xu J||Zhong CQ||Hu JL||Jiang WX...</td><td>Impact of multiple policy interventions on the screening and diagnosis of drug-resistant tuberculosis patients: a cascade analysis on six prefectures in China.</td><td>10</td><td>8</td><td>RAYYAN-INCLUSION: {"Donal"=>"Excluded", "Richard"=>"Maybe", "Jessica "=>"Excluded"} | RAYYAN-LABELS: Excluded (tiebreaker) | RAYYAN-EXCLUSION-REASONS: wrong infection / disease</td><td>China;Tuberculosis;Drug Resistance</td><td>PMC7814633</td></tr>
<tr><td>rayyan-651229678</td><td>Zhou G||Lee MC||Atieli HE||Githure JI||Githeko AK||Kazura JW||Yan G</td><td>Adaptive interventions for optimizing malaria control: an implementation study protocol for a block-cluster randomized, sequential multiple assignment trial.</td><td>21</td><td>665</td><td>RAYYAN-INCLUSION: {"Richard"=>"Excluded", "Jessica "=>"Excluded"} | RAYYAN-EXCLUSION-REASONS: wrong publication type</td><td>Malaria</td><td>PMC7372887</td></tr>
<tr><td>rayyan-651229679</td><td>Okitawutshu J||Signorell A||Kalenga JC||Mukomena E||Delvento G||Burri C||Mwaluke F||Buj V||Sangar...</td><td>Key factors predicting suspected severe malaria case management and health outcomes: an operational study in the Democratic Republic of the Congo.</td><td>21</td><td>274</td><td>RAYYAN-INCLUSION: {"Richard"=>"Excluded", "Jessica "=>"Excluded"} | RAYYAN-EXCLUSION-REASONS: wrong outcome,wrong study design</td><td>Case Management;Malaria</td><td>PMC9513903</td></tr>
<tr><td>rayyan-651229680</td><td>Tizifa TA||Gowelo S||Kabaghe AN||McCann RS||Malenga T||Nkhata RM||Kadama A||Chapeta Y||Takken W||...</td><td>Community-based house improvement for malaria control in southern Malawi: Stakeholder perceptions, experiences, and acceptability.</td><td>2</td><td>e0000627</td><td>RAYYAN-INCLUSION: {"Richard"=>"Excluded", "Jessica "=>"Excluded"} | RAYYAN-EXCLUSION-REASONS: wrong outcome,wrong study design</td><td>Malaria</td><td>PMC10021647</td></tr>
<tr><td>rayyan-651229681</td><td>Vassall A||Mangham-Jefferies L||Gomez GB||Pitt C||Foster N</td><td>Incorporating Demand and Supply Constraints into Economic Evaluations in Low-Income and Middle-Income Countries.</td><td>25</td><td>95-115</td><td>RAYYAN-INCLUSION: {"Richard"=>"Excluded", "Jessica "=>"Excluded"} | RAYYAN-EXCLUSION-REASONS: wrong outcome,wrong study design</td><td></td><td>PMC5042074</td></tr>
<tr><td>rayyan-651229682</td><td>Kabaghe AN||Chipeta MG||McCann RS||Terlouw DJ||Tizifa T||Truwah Z||Phiri KS||van Vugt M</td><td>Access and adequate utilization of malaria control interventions in rural Malawi: a descriptive quantitative study.</td><td>17</td><td>104</td><td>RAYYAN-INCLUSION: {"Richard"=>"Excluded", "Jessica "=>"Excluded"} | RAYYAN-EXCLUSION-REASONS: wrong outcome,wrong study design</td><td>Malaria</td><td>PMC5838945</td></tr>
<tr><td>rayyan-651229683</td><td>Nigussie TZ||Zewotir T||Muluneh EK</td><td>Effects of climate variability and environmental factors on the spatiotemporal distribution of malaria incidence in the Amhara national regional state, Ethiopia.</td><td>40</td><td>100475</td><td>RAYYAN-INCLUSION: {"Donal"=>"Included", "Richard"=>"Maybe", "Jessica "=>"Excluded"} | RAYYAN-LABELS: Include (tiebreaker) | USER-NOTES: {"Richard"=>["Unclear of intervention outcome"]}</td><td>Malaria</td><td></td></tr>
<tr><td>rayyan-651229685</td><td>Golumbeanu M||Yang GJ||Camponovo F||Stuckey EM||Hamon N||Mondy M||Rees S||Chitnis N||Cameron E||P...</td><td>Leveraging mathematical models of disease dynamics and machine learning to improve development of novel malaria interventions.</td><td>11</td><td>61</td><td>RAYYAN-INCLUSION: {"Richard"=>"Excluded", "Jessica "=>"Excluded"} | RAYYAN-EXCLUSION-REASONS: wrong outcome,wrong study design,wrong publication type</td><td>Mathematics;Malaria</td><td>PMC9167503</td></tr>
<tr><td>rayyan-651229686</td><td>Singh H||Gupta SK||Vikram K||Saxena R||Sharma A</td><td>The impact of mosquito proof lids of underground tanks "tanka" on the breeding of Anopheles stephensi in a village in western Rajasthan, India.</td><td>20</td><td>412</td><td>RAYYAN-INCLUSION: {"Richard"=>"Excluded", "Jessica "=>"Excluded"} | RAYYAN-EXCLUSION-REASONS: wrong outcome,wrong study design</td><td>Breeding;India;Culicidae</td><td>PMC8527675</td></tr>
<tr><td>rayyan-651229687</td><td>Muhammad MI||Abdulsalam AH||Petit-Clair F||Acquaye A||Nobles A||Fleischer B||Konkwo C||Ransome Y|...</td><td>Almajiri health; a scoping review on disease, health literacy and space for participatory research.</td><td>3</td><td>e0001641</td><td>RAYYAN-INCLUSION: {"Richard"=>"Excluded", "Jessica "=>"Excluded"} | RAYYAN-EXCLUSION-REASONS: wrong publication type,wrong infection / disease</td><td></td><td>PMC10566680</td></tr>
<tr><td>rayyan-651229688</td><td>Gerth-Guyette E||Nguyen HT||Nowak S||Hoang NT||Mai DTT||Thi Sang V||Long ND||Mvundura M||Nguyen N...</td><td>Assessing the Operational Feasibility of Integrating Point-of-Care G6PD Testing into Plasmodium vivax Malaria Management in Vietnam.</td><td>12</td><td></td><td>RAYYAN-INCLUSION: {"Richard"=>"Excluded", "Jessica "=>"Excluded"} | RAYYAN-EXCLUSION-REASONS: wrong outcome,wrong study design</td><td>Malaria, Vivax</td><td>PMC10222310</td></tr>
<tr><td>rayyan-651229689</td><td>Chersich MF||Wright CY</td><td>Climate change adaptation in South Africa: a case study on the role of the health sector.</td><td>15</td><td>22</td><td>RAYYAN-INCLUSION: {"Richard"=>"Excluded", "Jessica "=>"Excluded"} | RAYYAN-EXCLUSION-REASONS: wrong outcome,background article,wrong study design,wrong publication type</td><td>South Africa</td><td>PMC6423888</td></tr>
<tr><td>rayyan-651229690</td><td>Kabaghe AN||Phiri MD||Phiri KS||van Vugt M</td><td>Challenges in implementing uncomplicated malaria treatment in children: a health facility survey in rural Malawi.</td><td>16</td><td>419</td><td>RAYYAN-INCLUSION: {"Richard"=>"Excluded", "Jessica "=>"Excluded"} | RAYYAN-EXCLUSION-REASONS: wrong outcome,wrong study design</td><td>Only Child;Health Facilities;Child;Malaria</td><td>PMC5648442</td></tr>
<tr><td>rayyan-651229691</td><td>Rezaei-Hemami M||Akbari-Sari A||Raiesi A||Vatandoost H||Majdzadeh R</td><td>Cost Effectiveness of Malaria Interventions from Preelimination through Elimination: a Study in Iran.</td><td>8</td><td>43-52</td><td>RAYYAN-INCLUSION: {"Donal"=>"Excluded", "Richard"=>"Excluded", "Jessica "=>"Included"} | RAYYAN-LABELS: Excluded (tiebreaker) | RAYYAN-EXCLUSION-REASONS: wrong outcome,wrong study design | USER-NOTES: {"Richard"=>["Cost-effectiveness study"]}</td><td>Iran;Cost-Benefit Analysis;Malaria</td><td>PMC4289510</td></tr>
<tr><td>rayyan-651229692</td><td>Odongkara B||Nankabirwa V||Ndeezi G||Achora V||Arach AA||Napyo A||Musaba M||Mukunya D||Tumwine JK...</td><td>Incidence and Risk Factors for Low Birthweight and Preterm Birth in Post-Conflict Northern Uganda: A Community-Based Cohort Study.</td><td>19</td><td></td><td>RAYYAN-INCLUSION: {"Richard"=>"Excluded", "Jessica "=>"Excluded"} | RAYYAN-EXCLUSION-REASONS: wrong outcome,wrong study design,wrong infection / disease</td><td>Uganda;Cohort Studies</td><td>PMC9564590</td></tr>
<tr><td>rayyan-651229693</td><td>Gerardin J||Bever CA||Bridenbecker D||Hamainza B||Silumbe K||Miller JM||Eisele TP||Eckhoff PA||We...</td><td>Effectiveness of reactive case detection for malaria elimination in three archetypical transmission settings: a modelling study.</td><td>16</td><td>248</td><td>RAYYAN-INCLUSION: {"Richard"=>"Excluded", "Jessica "=>"Excluded"} | RAYYAN-EXCLUSION-REASONS: wrong outcome,wrong study design</td><td>Malaria</td><td>PMC5469005</td></tr>
<tr><td>rayyan-651229694</td><td>Nekorchuk DM||Gebrehiwot T||Lake M||Awoke W||Mihretie A||Wimberly MC</td><td>Comparing malaria early detection methods in a declining transmission setting in northwestern Ethiopia.</td><td>21</td><td>788</td><td>RAYYAN-INCLUSION: {"Richard"=>"Excluded", "Jessica "=>"Excluded"} | RAYYAN-EXCLUSION-REASONS: wrong outcome,wrong study design</td><td>Malaria</td><td>PMC8067323</td></tr>
<tr><td>rayyan-651229695</td><td>Thomas S||Ravishankaran S||Justin NA||Asokan A||Mathai MT||Valecha N||Montgomery J||Thomas MB||Ea...</td><td>Resting and feeding preferences of Anopheles stephensi in an urban setting, perennial for malaria.</td><td>16</td><td>111</td><td>RAYYAN-INCLUSION: {"Richard"=>"Excluded", "Jessica "=>"Excluded"} | RAYYAN-EXCLUSION-REASONS: wrong outcome,wrong study design</td><td>Malaria;Rest</td><td>PMC5346253</td></tr>
<tr><td>rayyan-651229696</td><td>Lanou HB||Osendarp SJM||Argaw A||De Polnay K||Ouedraogo C||Kouanda S||Kolsteren P</td><td>Micronutrient powder supplements combined with nutrition education marginally improve growth amongst children aged 6-23 months in rural Burkina Faso: A cluster randomized controlled trial.</td><td>15</td><td>e12820</td><td>RAYYAN-INCLUSION: {"Richard"=>"Excluded", "Jessica "=>"Excluded"} | RAYYAN-EXCLUSION-REASONS: wrong outcome,wrong study design,wrong infection / disease</td><td>Only Child;Powders;Child</td><td>PMC6859995</td></tr>
<tr><td>rayyan-651229697</td><td>Carrasco-Escobar G||Qquellon J||Villa D||Cava R||Llanos-Cuentas A||Benmarhnia T</td><td>Time-Varying Effects of Meteorological Variables on Malaria Epidemiology in the Context of Interrupted Control Efforts in the Amazon Rainforest, 2000-2017.</td><td>8</td><td>721515</td><td>RAYYAN-INCLUSION: {"Richard"=>"Excluded", "Jessica "=>"Excluded"} | RAYYAN-EXCLUSION-REASONS: wrong outcome,wrong study design,wrong study duration</td><td>Malaria</td><td>PMC8511324</td></tr>
<tr><td>rayyan-651229698</td><td>Mayor A||da Silva C||Rovira-Vallbona E||Roca-Feltrer A||Bonnington C||Wharton-Smith A||Greenhouse...</td><td>Prospective surveillance study to detect antimalarial drug resistance, gene deletions of diagnostic relevance and genetic diversity of Plasmodium falciparum in Mozambique: protocol.</td><td>12</td><td>e063456</td><td>RAYYAN-INCLUSION: {"Richard"=>"Excluded", "Jessica "=>"Excluded"} | RAYYAN-EXCLUSION-REASONS: wrong outcome,wrong study design</td><td>Genetic Variation;Antimalarials;Gene Deletion;Plasmodium</td><td>PMC9274532</td></tr>
<tr><td>rayyan-651229699</td><td>Stuckey EM||Stevenson J||Galactionova K||Baidjoe AY||Bousema T||Odongo W||Kariuki S||Drakeley C||...</td><td>Modeling the cost effectiveness of malaria control interventions in the highlands of western Kenya.</td><td>9</td><td>e107700</td><td>RAYYAN-INCLUSION: {"Richard"=>"Excluded", "Jessica "=>"Excluded"} | RAYYAN-EXCLUSION-REASONS: wrong outcome,wrong study design</td><td>Cost-Benefit Analysis;Kenya;Malaria</td><td>PMC4188563</td></tr>
<tr><td>rayyan-651229700</td><td>Malenga T||Griffiths FE||van den Berg M||van den Berg H||van Vugt M||Phiri KS||Manda-Taylor L||Um...</td><td>A qualitative exploration of the experiences of community health animation on malaria control in rural Malawi.</td><td>16</td><td>25</td><td>RAYYAN-INCLUSION: {"Richard"=>"Excluded", "Jessica "=>"Excluded"} | RAYYAN-EXCLUSION-REASONS: wrong outcome,wrong study design</td><td>Animals;Animal Shells;Malaria</td><td>PMC7085180</td></tr>
<tr><td>rayyan-651229701</td><td>Faujdar DS||Sahay S||Singh T||Kaur M||Kumar R</td><td>Field testing of a digital health information system for primary health care: A quasi-experimental study from India.</td><td>141</td><td>104235</td><td>RAYYAN-INCLUSION: {"Richard"=>"Excluded", "Jessica "=>"Excluded"} | RAYYAN-EXCLUSION-REASONS: wrong outcome,wrong study design</td><td>Information Systems;India</td><td></td></tr>
<tr><td>rayyan-651229702</td><td>Druetz T||Stresman G||Ashton RA||van den Hoogen LL||Joseph V||Fayette C||Monestime F||Hamre KE||C...</td><td>Programmatic options for monitoring malaria in elimination settings: easy access group surveys to investigate Plasmodium falciparum epidemiology in two regions with differing endemicity in Haiti.</td><td>18</td><td>141</td><td>RAYYAN-INCLUSION: {"Richard"=>"Excluded", "Jessica "=>"Excluded"} | RAYYAN-EXCLUSION-REASONS: wrong outcome</td><td>Plasmodium falciparum;Malaria;Plasmodium</td><td>PMC7310408</td></tr>
<tr><td>rayyan-651229703</td><td>Ciapponi A||Lewin S||Herrera CA||Opiyo N||Pantoja T||Paulsen E||Rada G||Wiysonge CS||Bastias G||D...</td><td>Delivery arrangements for health systems in low-income countries: an overview of systematic reviews.</td><td>9</td><td>CD011083</td><td>RAYYAN-INCLUSION: {"Richard"=>"Excluded", "Jessica "=>"Excluded"} | RAYYAN-EXCLUSION-REASONS: wrong outcome,background article,wrong study design,wrong publication type</td><td></td><td>PMC5621087</td></tr>
<tr><td>rayyan-651229704</td><td>Verma M||Panwar S||Sahoo SS||Grover GS||Aggarwal S||Tripathy JP||Shah J||Kakkar R</td><td>Mapping the stability of febrile illness hotspots in Punjab from 2012 to 2019- a spatial clustering and regression analysis.</td><td>23</td><td>2014</td><td>RAYYAN-INCLUSION: {"Richard"=>"Excluded", "Jessica "=>"Excluded"} | RAYYAN-EXCLUSION-REASONS: wrong outcome,wrong study design,wrong infection / disease</td><td>Regression Analysis</td><td>PMC10580620</td></tr>
<tr><td>rayyan-651229705</td><td>Lwetoijera D||Kiware S||Okumu F||Devine GJ||Majambere S</td><td>Autodissemination of pyriproxyfen suppresses stable populations of Anopheles arabiensis under semi-controlled settings.</td><td>18</td><td>166</td><td>RAYYAN-INCLUSION: {"Richard"=>"Excluded", "Jessica "=>"Excluded"} | RAYYAN-EXCLUSION-REASONS: wrong outcome,wrong study design</td><td>Population Dynamics</td><td>PMC6507228</td></tr>
<tr><td>rayyan-651229706</td><td>Wisniewski J||Worges M||Lusamba-Dikassa PS</td><td>Impact of a free care policy on routine health service volumes during a protracted Ebola virus disease outbreak in the Democratic Republic of Congo.</td><td>322</td><td>115815</td><td>RAYYAN-INCLUSION: {"Donal"=>"Excluded", "Richard"=>"Excluded", "Jessica "=>"Included"} | RAYYAN-LABELS: Excluded (tiebreaker) | RAYYAN-EXCLUSION-REASONS: wrong infection / disease</td><td>Virus Diseases;Disease Outbreaks</td><td></td></tr>
<tr><td>rayyan-651229707</td><td>Essendi WM||Vardo-Zalik AM||Lo E||Machani MG||Zhou G||Githeko AK||Yan G||Afrane YA</td><td>Epidemiological risk factors for clinical malaria infection in the highlands of Western Kenya.</td><td>18</td><td>211</td><td>RAYYAN-INCLUSION: {"Richard"=>"Excluded", "Jessica "=>"Excluded"} | RAYYAN-EXCLUSION-REASONS: wrong outcome,wrong study design</td><td>Risk Factors;Kenya;Malaria</td><td>PMC6591804</td></tr>
<tr><td>rayyan-651229708</td><td>Tugwell P||Petkovic J||Welch V||Vincent J||Bhutta ZA||Churchill R||deSavigny D||Mbuagbaw L||Panto...</td><td>Setting priorities for knowledge translation of Cochrane reviews for health equity: Evidence for Equity.</td><td>16</td><td>208</td><td>RAYYAN-INCLUSION: {"Richard"=>"Excluded", "Jessica "=>"Excluded"} | RAYYAN-EXCLUSION-REASONS: wrong outcome,wrong study design,wrong publication type</td><td></td><td>PMC5712153</td></tr>
<tr><td>rayyan-651229709</td><td>Sisya TJ||Kamn'gona RM||Vareta JA||Fulakeza JM||Mukaka MF||Seydel KB||Laufer MK||Taylor TE||Nkhom...</td><td>Subtle changes in Plasmodium falciparum infection complexity following enhanced intervention in Malawi.</td><td>142</td><td>108-14</td><td>RAYYAN-INCLUSION: {"Richard"=>"Excluded", "Jessica "=>"Excluded"} | RAYYAN-EXCLUSION-REASONS: wrong outcome,wrong study design,wrong study duration</td><td>Plasmodium falciparum;Plasmodium</td><td>PMC4296692</td></tr>
</table>

| | | | | | | | |
|---|---|---|---|---|---|---|---|
| rayyan-651229710 | Moreno-Gutierrez D\|\|Llanos-Cuentas A\|\|Luis Barboza J\|\|Contreras-Mancilla J\|\|Gamboa D\|\|Rodriguez H... | Effectiveness of a Malaria Surveillance Strategy Based on Active Case Detection during High Transmission Season in the Peruvian Amazon. | 15 | | RAYYAN-INCLUSION: {"Richard"=>"Excluded", "Jessica "=>"Excluded"} \| RAYYAN-EXCLUSION-REASONS: wrong outcome,wrong study design | Malaria;Seasons | PMC6314008 |
| rayyan-651229711 | Cook J\|\|Hergott D\|\|Phiri W\|\|Rivas MR\|\|Bradley J\|\|Segura L\|\|Garcia G\|\|Schwabe C\|\|Kleinschmidt I | Trends in parasite prevalence following 13 years of malaria interventions on Bioko island, Equatorial Guinea: 2004-2016. | 17 | 62 | RAYYAN-INCLUSION: {"Donal"=>"Included", "Richard"=>"Included", "Jessica "=>"Excluded"} \| RAYYAN-LABELS: Include (tiebreaker) | Prevalence;Malaria | PMC5799938 |
| rayyan-651229712 | Chow WK\|\|Beebe NW\|\|Ambrose L\|\|Pickering P\|\|Cooper RD | Seasonal assessment on the effects of time of night, temperature and humidity on the biting profile of Anopheles farauti in north Queensland, Australia using a population naive to malaria vector control pressures. | 22 | 85 | RAYYAN-INCLUSION: {"Richard"=>"Excluded", "Jessica "=>"Excluded"} \| RAYYAN-EXCLUSION-REASONS: wrong outcome,wrong study design | Seasons | PMC9996873 |
| rayyan-651229713 | Wollum A\|\|Burstein R\|\|Fullman N\|\|Dwyer-Lindgren L\|\|Gakidou E | Benchmarking health system performance across states in Nigeria: a systematic analysis of levels and trends in key maternal and child health interventions and outcomes, 2000-2013. | 13 | 208 | RAYYAN-INCLUSION: {"Donal"=>"Excluded", "Richard"=>"Excluded", "Jessica "=>"Maybe"} \| RAYYAN-LABELS: Excluded (tiebreaker) \| RAYYAN-EXCLUSION-REASONS: wrong outcome,background article,wrong study design,wrong study duration,wrong publication type | Only Child;Child;Child Welfare;Nigeria;Benchmarking | PMC4557921 |
| rayyan-651229714 | Coldiron ME\|\|Lasry E\|\|Bouhenia M\|\|Das D\|\|Okui P\|\|Nyehangane D\|\|Mwanga J\|\|Langendorf C\|\|Elder G\|\|S... | Intermittent preventive treatment for malaria among children in a refugee camp in Northern Uganda: lessons learned. | 16 | 218 | RAYYAN-INCLUSION: {"Donal"=>"Included", "Richard"=>"Included", "Jessica "=>"Excluded"} \| RAYYAN-LABELS: Include (tiebreaker) | Uganda;Malaria;Only Child;Child | PMC5442684 |
| rayyan-651229715 | Mwakalinga VM\|\|Sartorius BK\|\|Mlacha YP\|\|Msellemu DF\|\|Limwagu AJ\|\|Mageni ZD\|\|Paliga JM\|\|Govella NJ... | Spatially aggregated clusters and scattered smaller loci of elevated malaria vector density and human infection prevalence in urban Dar es Salaam, Tanzania. | 15 | 135 | RAYYAN-INCLUSION: {"Richard"=>"Excluded", "Jessica "=>"Excluded"} \| RAYYAN-EXCLUSION-REASONS: wrong study design,wrong study duration | Humanities;Humanism;Humans;Prevalence;Malaria | PMC4774196 |
| rayyan-651229716 | Foy BD\|\|Some A\|\|Magalhaes T\|\|Gray L\|\|Rao S\|\|Sougue E\|\|Jackson CL\|\|Kittelson J\|\|Slater HC\|\|Bousema... | Repeat Ivermectin Mass Drug Administrations for Malaria Control II: Protocol for a Double-blind, Cluster-Randomized, Placebo-Controlled Trial for the Integrated Control of Malaria. | 12 | e41197 | RAYYAN-INCLUSION: {"Richard"=>"Excluded", "Jessica "=>"Excluded"} \| RAYYAN-EXCLUSION-REASONS: wrong publication type | Malaria | PMC10132043 |
| rayyan-651229717 | Sanou A\|\|Nelli L\|\|Guelbeogo WM\|\|Cisse F\|\|Tapsoba M\|\|Ouedraogo P\|\|Sagnon N\|\|Ranson H\|\|Matthiopoulo... | Insecticide resistance and behavioural adaptation as a response to long-lasting insecticidal net deployment in malaria vectors in the Cascades region of Burkina Faso. | 11 | 17569 | RAYYAN-INCLUSION: {"Richard"=>"Excluded", "Jessica "=>"Excluded"} \| RAYYAN-EXCLUSION-REASONS: wrong outcome,wrong study design | Insecticides;Malaria | PMC8413378 |
| rayyan-651229718 | Soniran OT\|\|Abuaku B\|\|Ahorlu CS | Evaluating interventions to improve test, treat, and track (T3) malaria strategy among over-the-counter medicine sellers (OTCMS) in some rural communities of Fanteakwa North district, Ghana: study protocol for a cluster randomized controlled trial. | 21 | 623 | RAYYAN-INCLUSION: {"Richard"=>"Excluded", "Jessica "=>"Excluded"} \| RAYYAN-EXCLUSION-REASONS: wrong publication type | Rural Population;Ghana;Malaria | PMC7346649 |
| rayyan-651229719 | Ngo TD\|\|Canavati SE\|\|Dinh HS\|\|Ngo TD\|\|Tran DT\|\|Martin NJ\|\|Kelly GC | Addressing operational challenges of combatting malaria in a remote forest area of Vietnam using spatial decision support system approaches. | 14 | | RAYYAN-INCLUSION: {"Richard"=>"Excluded", "Jessica "=>"Excluded"} \| RAYYAN-EXCLUSION-REASONS: wrong outcome,wrong study design | Vietnam;Malaria;Trees | |
| rayyan-651229720 | Kabaria CW\|\|Molteni F\|\|Mandike R\|\|Chacky F\|\|Noor AM\|\|Snow RW\|\|Linard C | Mapping intra-urban malaria risk using high resolution satellite imagery: a case study of Dar es Salaam. | 15 | 26 | RAYYAN-INCLUSION: {"Richard"=>"Excluded", "Jessica "=>"Excluded"} \| RAYYAN-EXCLUSION-REASONS: wrong outcome,wrong study design | Imagery (Psychotherapy);Malaria | PMC4967308 |
| rayyan-651229721 | Hast M\|\|Searle KM\|\|Chaponda M\|\|Lupiya J\|\|Lubinda J\|\|Sikalima J\|\|Kobayashi T\|\|Shields T\|\|Mulenga M... | The use of GPS data loggers to describe the impact of spatio-temporal movement patterns on malaria control in a high-transmission area of northern Zambia. | 18 | 19 | RAYYAN-INCLUSION: {"Richard"=>"Excluded", "Jessica "=>"Excluded"} \| RAYYAN-EXCLUSION-REASONS: wrong outcome,wrong study design | Malaria | PMC6701131 |
| rayyan-651229722 | Okebe J\|\|Ribera JM\|\|Balen J\|\|Jaiteh F\|\|Masunaga Y\|\|Nwakanma D\|\|Bradley J\|\|Yeung S\|\|Peeters Griete... | Reactive community-based self-administered treatment against residual malaria transmission: study protocol for a randomized controlled trial. | 19 | 126 | RAYYAN-INCLUSION: {"Richard"=>"Excluded", "Jessica "=>"Excluded"} \| RAYYAN-EXCLUSION-REASONS: wrong outcome,wrong study design | Malaria | PMC5819183 |
| rayyan-651229723 | Alafo C\|\|Marti-Soler H\|\|Maquina M\|\|Malheia A\|\|Aswat AS\|\|Koekemoer LL\|\|Colborn J\|\|Lobo NF\|\|Tatarsk... | To spray or target mosquitoes another way: focused entomological intelligence guides the implementation of indoor residual spraying in southern Mozambique. | 21 | 215 | RAYYAN-INCLUSION: {"Richard"=>"Excluded", "Jessica "=>"Excluded"} \| RAYYAN-EXCLUSION-REASONS: wrong outcome,wrong study design | Intelligence;Culicidae | PMC9275269 |
| rayyan-651229724 | Githinji S\|\|Jones C\|\|Malinga J\|\|Snow RW\|\|Talisuna A\|\|Zurovac D | Development of a text-messaging intervention to improve treatment adherence and post-treatment review of children with uncomplicated malaria in western Kenya. | 14 | 320 | RAYYAN-INCLUSION: {"Richard"=>"Excluded", "Jessica "=>"Excluded"} \| RAYYAN-EXCLUSION-REASONS: wrong outcome,wrong study design | Kenya;Malaria | PMC4539928 |
| rayyan-651229725 | Martineau P\|\|Behera SK\|\|Nonaka M\|\|Jayanthi R\|\|Ikeda T\|\|Minakawa N\|\|Kruger P\|\|Mabunda QE | Predicting malaria outbreaks from sea surface temperature variability up to 9 months ahead in Limpopo, South Africa, using machine learning. | 10 | 962377 | RAYYAN-INCLUSION: {"Richard"=>"Excluded", "Jessica "=>"Excluded"} \| RAYYAN-EXCLUSION-REASONS: wrong outcome,wrong study design | South Africa;Disease Outbreaks;Temperature | PMC9453600 |
| rayyan-651229726 | Florez D\|\|Young AJ\|\|Bernabe KJ\|\|Hyman JM\|\|Qu Z | Modeling Sustained Transmission of Wolbachia among Anopheles Mosquitoes: Implications for Malaria Control in Haiti. | 8 | | RAYYAN-INCLUSION: {"Richard"=>"Excluded", "Jessica "=>"Excluded"} \| RAYYAN-EXCLUSION-REASONS: wrong outcome,wrong study design | Culicidae;Malaria | PMC10056903 |
| rayyan-651229727 | Haenssgen MJ\|\|Charoenboon N\|\|Althaus T\|\|Greer RC\|\|Intralawan D\|\|Lubell Y | The social role of C-reactive protein point-of-care testing to guide antibiotic prescription in Northern Thailand. | 202 | 12-Jan | RAYYAN-INCLUSION: {"Richard"=>"Excluded", "Jessica "=>"Excluded"} \| RAYYAN-EXCLUSION-REASONS: wrong outcome,wrong study design | C-Reactive Protein;Prescriptions | PMC5910303 |
| rayyan-651229728 | Kumari J\|\|Kumar V\|\|Behl A\|\|Kumar Sah R\|\|Kumari G\|\|Garg S\|\|Gupta A\|\|Nazar Mohomed Mohaideen S\|\|Sha... | 'Erythritol', a safe natural sweetener exhibits multi-stage anti-malarial activity by permeating into Plasmodium falciparum through aquaglyceroporin channel. | 205 | 115287 | RAYYAN-INCLUSION: {"Richard"=>"Excluded", "Jessica "=>"Excluded"} \| RAYYAN-EXCLUSION-REASONS: wrong outcome,wrong study design | Plasmodium falciparum;Antimalarials;Plasmodium | |
| rayyan-651229729 | Westercamp N\|\|Staedke SG\|\|Maiteki-Sebuguzi C\|\|Ndyabakira A\|\|Okiring JM\|\|Kigozi SP\|\|Dorsey G\|\|Brou... | Effectiveness of in-service training plus the collaborative improvement strategy on the quality of routine malaria surveillance data: results of a pilot study in Kayunga District, Uganda. | 20 | 290 | RAYYAN-INCLUSION: {"Richard"=>"Excluded", "Jessica "=>"Excluded"} \| RAYYAN-EXCLUSION-REASONS: wrong outcome,wrong study design | Uganda;Pilot Projects;Malaria | PMC8243434 |
| rayyan-651229730 | Kaunda-Khangamwa BN\|\|van den Berg H\|\|McCann RS\|\|Kabaghe A\|\|Takken W\|\|Phiri K\|\|van Vugt M\|\|Manda-T... | The role of health animators in malaria control: a qualitative study of the health animator (HA) approach within the Majete malaria project (MMP) in Chikwawa District, Malawi. | 19 | 478 | RAYYAN-INCLUSION: {"Richard"=>"Excluded", "Jessica "=>"Excluded"} \| RAYYAN-EXCLUSION-REASONS: wrong outcome,wrong study design | Animal Shells;Animals;Malaria;Matrix Metalloproteinases | PMC6624973 |
| rayyan-651229731 | Nabyonga-Orem J\|\|Ssengooba F\|\|Macq J\|\|Criel B | Malaria treatment policy change in Uganda: what role did evidence play? | 13 | 345 | RAYYAN-INCLUSION: {"Richard"=>"Excluded", "Jessica "=>"Excluded"} \| RAYYAN-EXCLUSION-REASONS: wrong outcome,wrong study design,wrong publication type | Uganda;Malaria | PMC4164770 |
| rayyan-651229732 | Van Bortel W\|\|Marien J\|\|Jacobs BKM\|\|Sinzinkayo D\|\|Sinarinzi P\|\|Lampaert E\|\|D'hondt R\|\|Mafuko JM\|\|... | Long-lasting insecticidal nets provide protection against malaria for only a single year in Burundi, an African highland setting with marked malaria seasonality. | 7 | | RAYYAN-INCLUSION: {"Richard"=>"Included", "Jessica "=>"Included"} | Seasons;Malaria;Insecticides | PMC9772646 |
| rayyan-651229733 | Coulibaly A\|\|Diop MF\|\|Kone A\|\|Dara A\|\|Ouattara A\|\|Mulder N\|\|Miotto O\|\|Diakite M\|\|Djimde A\|\|Amambu... | Genome-wide SNP analysis of Plasmodium falciparum shows differentiation at drug-resistance-associated loci among malaria transmission settings in southern Mali. | 13 | 943445 | RAYYAN-INCLUSION: {"Richard"=>"Excluded", "Jessica "=>"Excluded"} \| RAYYAN-EXCLUSION-REASONS: wrong outcome,wrong study design | Genome;Genomics;Plasmodium falciparum;Malaria;Polymorphi | PMC9576839 |
| rayyan-651229734 | Zelman BW\|\|Baral R\|\|Zarlinda I\|\|Coutrier FN\|\|Sanders KC\|\|Cotter C\|\|Herdiana H\|\|Greenhouse B\|\|Shre... | Costs and cost-effectiveness of malaria reactive case detection using loop-mediated isothermal amplification compared to microscopy in the low transmission setting of Aceh Province, Indonesia. | 17 | 220 | RAYYAN-INCLUSION: {"Richard"=>"Excluded", "Jessica "=>"Excluded"} \| RAYYAN-EXCLUSION-REASONS: wrong outcome,wrong study design | Malaria;Cost-Benefit Analysis | PMC5984760 |
| rayyan-651229735 | Lengeler C\|\|Burri C\|\|Awor P\|\|Athieno P\|\|Kimera J\|\|Tumukunde G\|\|Angiro I\|\|Tshefu A\|\|Okitawutshu J\|... | Community access to rectal artesunate for malaria (CARAMAL): A large-scale observational implementation study in the Democratic Republic of the Congo, Nigeria and Uganda. | 2 | e0000464 | RAYYAN-INCLUSION: {"Richard"=>"Excluded", "Jessica "=>"Excluded"} \| RAYYAN-EXCLUSION-REASONS: wrong publication type | Uganda;Nigeria;Malaria | PMC10022208 |
| rayyan-651229736 | Strachan CE\|\|Kana M\|\|Martin S\|\|Dada J\|\|Wandera N\|\|Marasciulo M\|\|Counihan H\|\|Kolawole M\|\|Babale T\|... | The use of formative research to inform the design of a seasonal malaria chemoprevention intervention in northern Nigeria. | 15 | 474 | RAYYAN-INCLUSION: {"Richard"=>"Excluded", "Jessica "=>"Excluded"} \| RAYYAN-EXCLUSION-REASONS: wrong outcome,wrong study design | Nigeria;Chemoprevention;Malaria;Seasons | PMC5025607 |
| rayyan-651229737 | Mukhtar AYA\|\|Munyakazi JB\|\|Ouifki R\|\|Clark AE | Modelling the effect of bednet coverage on malaria transmission in South Sudan. | 13 | e0198280 | RAYYAN-INCLUSION: {"Richard"=>"Excluded", "Jessica "=>"Excluded"} \| RAYYAN-EXCLUSION-REASONS: wrong study design | Sudan;Malaria | PMC5991726 |
| rayyan-651229738 | Hiscox A\|\|Homan T\|\|Mweresa CK\|\|Maire N\|\|Di Pasquale A\|\|Masiga D\|\|Oria PA\|\|Alaii J\|\|Leeuwis C\|\|Muk... | Mass mosquito trapping for malaria control in western Kenya: study protocol for a stepped wedge cluster-randomised trial. | 17 | 356 | RAYYAN-INCLUSION: {"Richard"=>"Excluded", "Jessica "=>"Excluded"} \| RAYYAN-EXCLUSION-REASONS: wrong outcome,wrong study design,wrong publication type | Culicidae;Kenya;Malaria | PMC4962350 |
| rayyan-651229739 | Ide N\|\|Hardy V\|\|Chirambo G\|\|Heavin C\|\|O'Connor Y\|\|O'Donoghue J\|\|Mastellos N\|\|Dharmayat K\|\|Anderss... | People Welcomed This Innovation with Two Hands: A Qualitative Report of an mHealth Intervention for Community Case Management in Malawi. | 85 | | RAYYAN-INCLUSION: {"Richard"=>"Excluded", "Jessica "=>"Excluded"} \| RAYYAN-EXCLUSION-REASONS: wrong outcome,wrong study design | Case Management | PMC6634480 |
| rayyan-651229740 | Ma Y\|\|Kim H\|\|Cho Y\|\|Lee J\|\|Degley JK\|\|Adam AG\|\|Lee G\|\|Lee H\|\|Cha S | Effects of community health volunteers on infectious diseases of children under five in Volta Region, Ghana: study protocol for a cluster randomized controlled trial. | 17 | 95 | RAYYAN-INCLUSION: {"Richard"=>"Excluded", "Jessica "=>"Excluded"} \| RAYYAN-EXCLUSION-REASONS: wrong outcome,wrong study design,wrong publication type | Ghana;Communicable Diseases | PMC5244532 |
| rayyan-651229741 | Duke T\|\|Hwaihwanje I\|\|Kaupa M\|\|Karubi J\|\|Panauwe D\|\|Sa'avu M\|\|Pulsan F\|\|Prasad P\|\|Maru F\|\|Tenambo... | Solar powered oxygen systems in remote health centers in Papua New Guinea: a large scale implementation effectiveness trial. | 7 | 010411 | RAYYAN-INCLUSION: {"Richard"=>"Excluded", "Jessica "=>"Excluded"} \| RAYYAN-EXCLUSION-REASONS: wrong outcome,wrong study design,wrong infection / disease | Oxygenators | PMC5441450 |

| | | | | | | | |
|---|---|---|---|---|---|---|---|
| rayyan-651229742 | Kitutu FE\|\|Mayora C\|\|Johansson EW\|\|Peterson S\|\|Wamani H\|\|Bigdeli M\|\|Shroff ZC | Health system effects of implementing integrated community case management (iCCM) intervention in private retail drug shops in South Western Uganda: a qualitative study. | 2 | e000334 | RAYYAN-INCLUSION: {"Richard"=>"Excluded", "Jessica "=>"Excluded"} \| RAYYAN-EXCLUSION-REASONS: wrong outcome,wrong study design | Uganda;Case Management | PMC5717929 |
| rayyan-651229743 | Talisuna AO\|\|Zurovac D\|\|Githinji S\|\|Oburu A\|\|Malinga J\|\|Nyandigisi A\|\|Jones CO\|\|Snow RW | Efficacy of Mobile Phone Short Message Service (SMS) Reminders on Malaria Treatment Adherence and Day 3 Post-Treatment Reviews (SMS-RES-MAL) in Kenya: A Study Protocol. | 5 | 217 | RAYYAN-INCLUSION: {"Richard"=>"Excluded", "Jessica "=>"Excluded"} \| RAYYAN-EXCLUSION-REASONS: wrong publication type | Kenya;Malaria | PMC6614027 |
| rayyan-651229744 | Deng C\|\|Wang Q\|\|Zheng S\|\|Zhou C\|\|Gao Y\|\|Guo J\|\|Mliva AM\|\|Oithik F\|\|Bacar A\|\|Attoumane R\|\|Song J | Mass Drug Administration of Artemisinin-piperaquine on High Malaria Epidemic Area. | 42 | 33-41 | RAYYAN-INCLUSION: {"Richard"=>"Excluded", "Jessica "=>"Excluded"} \| RAYYAN-EXCLUSION-REASONS: wrong outcome,background article,wrong study design,wrong publication type | Malaria | PMC4204058 |
| rayyan-651229745 | Dun-Dery F\|\|Meissner P\|\|Beiersmann C\|\|Kuunibe N\|\|Winkler V\|\|Albrecht J\|\|Muller O | Uptake challenges of intermittent preventive malaria therapy among pregnant women and their health care providers in the Upper West Region of Ghana: A mixed-methods study. | 15 | e00222 | RAYYAN-INCLUSION: {"Richard"=>"Excluded", "Jessica "=>"Excluded"} \| RAYYAN-EXCLUSION-REASONS: wrong outcome,wrong study design | Women's Health;Ghana;Malaria | PMC8488310 |
| rayyan-651229746 | Offor C\|\|Ade-Banjo O\|\|Nwankwo C\|\|Nwaononiwu G\|\|Adukwu F\|\|Egharevba B\|\|Owoyemi J\|\|Odo C\|\|Olatunji M | Evidence for "Whole Family Approach" in accelerating uptake of COVID-19 and routine immunizations among integrated primary health services in Nigeria. | 3 | 1157377 | RAYYAN-INCLUSION: {"Richard"=>"Excluded", "Jessica "=>"Excluded"} \| RAYYAN-EXCLUSION-REASONS: wrong outcome,wrong study design,wrong infection / disease | Nigeria | PMC10232860 |
| rayyan-651229747 | Wu L\|\|Hsiang MS\|\|Prach LM\|\|Schrubbe L\|\|Ntuku H\|\|Dufour MK\|\|Whittemore B\|\|Scott V\|\|Yala J\|\|Roberts... | Serological evaluation of the effectiveness of reactive focal mass drug administration and reactive vector control to reduce malaria transmission in Zambezi Region, Namibia: Results from a secondary analysis of a cluster randomised trial. | 44 | 101272 | RAYYAN-INCLUSION: {"Richard"=>"Excluded", "Jessica "=>"Excluded"} \| RAYYAN-EXCLUSION-REASONS: wrong study design | Malaria | PMC8851292 |
| rayyan-651229748 | Murhandarwati EE\|\|Fuad A\|\|Sulistyawati\|\|Wijayanti MA\|\|Bia MB\|\|Widartono BS\|\|Kuswantoro\|\|Lobo NF\|\|... | Change of strategy is required for malaria elimination: a case study in Purworejo District, Central Java Province, Indonesia. | 14 | 318 | RAYYAN-INCLUSION: {"Richard"=>"Excluded", "Jessica "=>"Excluded"} \| RAYYAN-EXCLUSION-REASONS: wrong outcome,wrong study design,wrong study duration | Malaria | PMC4537575 |
| rayyan-651229749 | Sanchez-Cano A\|\|Ruiz-Vega G\|\|Vicente-Gomez S\|\|de la Serna E\|\|Sulleiro E\|\|Molina I\|\|Sanchez-Montal... | Development of a Fast Chemiluminescent Magneto-Immunoassay for Sensitive Plasmodium falciparum Detection in Whole Blood. | 93 | 12793-12800 | RAYYAN-INCLUSION: {"Richard"=>"Excluded", "Jessica "=>"Excluded"} \| RAYYAN-EXCLUSION-REASONS: wrong outcome,wrong study design | Plasmodium falciparum;Immunoassay;Plasmodium | |
| rayyan-651229750 | Badger-Emeka LI | The malaria burden: A look at 3 years outpatient malaria clinic visits in a university community town in Southeast of Nigeria. | 23 | 711-719 | RAYYAN-INCLUSION: {"Donal"=>"Excluded", "Richard"=>"Excluded", "Jessica "=>"Maybe"} \| RAYYAN-LABELS: Excluded (tiebreaker) \| RAYYAN-EXCLUSION-REASONS: wrong outcome,wrong study design | Ambulatory Care;Southeastern United States;Nigeria;Malaria | |
| rayyan-651229751 | Chirambo GB\|\|Thompson M\|\|Hardy V\|\|Ide N\|\|Hwang PH\|\|Dharmayat K\|\|Mastellos N\|\|Heavin C\|\|O'Connor Y... | Effectiveness of Smartphone-Based Community Case Management on the Urgent Referral, Reconsultation, and Hospitalization of Children Aged Under 5 Years in Malawi: Cluster-Randomized, Stepped-Wedge Trial. | 23 | e25777 | RAYYAN-INCLUSION: {"Donal"=>"Excluded", "Richard"=>"Maybe", "Jessica "=>"Excluded"} \| RAYYAN-LABELS: Excluded (tiebreaker) \| USER-NOTES: {"Richard"=>["Unclear if routine data were used"]} | Case Management;Referral and Consultation | PMC8567152 |
| rayyan-651229752 | Manrique-Saide P\|\|Dean NE\|\|Halloran ME\|\|Longini IM\|\|Collins MH\|\|Waller LA\|\|Gomez-Dantes H\|\|Lenhar... | The TIRS trial: protocol for a cluster randomized controlled trial assessing the efficacy of preventive targeted indoor residual spraying to reduce Aedes-borne viral illnesses in Merida, Mexico. | 21 | 839 | RAYYAN-INCLUSION: {"Richard"=>"Excluded", "Jessica "=>"Excluded"} \| RAYYAN-EXCLUSION-REASONS: background article,wrong publication type,wrong infection / disease | Mexico;Aedes | PMC7542575 |
| rayyan-651229753 | Rumisha SF\|\|Smith T\|\|Abdulla S\|\|Masanja H\|\|Vounatsou P | Modelling heterogeneity in malaria transmission using large sparse spatio-temporal entomological data. | 7 | 22682 | RAYYAN-INCLUSION: {"Richard"=>"Excluded", "Jessica "=>"Excluded"} \| RAYYAN-EXCLUSION-REASONS: wrong outcome,wrong study design | Malaria | PMC4071307 |
| rayyan-651229754 | Li-Dan L\|\|Dong A\|\|Dan-Cheng Y\|\|Jian-Jun X\|\|Guo-Yan W\|\|Xiao-Yan P\|\|Qiong L\|\|Le-Mei Z\|\|Zhong-Hao Y | [Pilot study on comprehensive malaria control in Judong Village, Congjiang County, Guizhou Province]. | 31 | 175-177 | RAYYAN-INCLUSION: {"Richard"=>"Excluded", "Jessica "=>"Excluded"} \| RAYYAN-EXCLUSION-REASONS: wrong outcome,wrong study design | Pilot Projects;Malaria | |
| rayyan-651229755 | Silal SP\|\|Little F\|\|Barnes KI\|\|White LJ | Towards malaria elimination in Mpumalanga, South Africa: a population-level mathematical modelling approach. | 13 | 297 | RAYYAN-INCLUSION: {"Richard"=>"Excluded", "Jessica "=>"Excluded"} \| RAYYAN-EXCLUSION-REASONS: wrong study duration | South Africa;Mathematics;Malaria | PMC4127654 |
| rayyan-651229756 | Chaccour CJ\|\|Alonso S\|\|Zulliger R\|\|Wagman J\|\|Saifodine A\|\|Candrinho B\|\|Macete E\|\|Brew J\|\|Fornadel... | Combination of indoor residual spraying with long-lasting insecticide-treated nets for malaria control in Zambezia, Mozambique: a cluster randomised trial and cost-effectiveness study protocol. | 3 | e000610 | RAYYAN-INCLUSION: {"Richard"=>"Excluded", "Jessica "=>"Excluded"} \| RAYYAN-EXCLUSION-REASONS: background article,wrong publication type | Cost-Benefit Analysis;Malaria;Insecticides | PMC5859815 |
| rayyan-651229757 | Hetzel MW\|\|Pulford J\|\|Maraga S\|\|Barnadas C\|\|Reimer LJ\|\|Tavul L\|\|Jamea-Maiasa S\|\|Tandrapah T\|\|Maal... | Evaluation of the Global Fund-supported National Malaria Control Program in Papua New Guinea, 2009-2014. | 57 | 29-Jul | RAYYAN-INCLUSION: {"Richard"=>"Excluded", "Jessica "=>"Excluded"} \| RAYYAN-EXCLUSION-REASONS: wrong outcome,wrong study design | Malaria | |
| rayyan-651229758 | Diawara H\|\|Walker P\|\|Cairns M\|\|Steinhardt LC\|\|Diawara F\|\|Kamate B\|\|Duval L\|\|Sicuri E\|\|Sagara I\|\|S... | Cost-effectiveness of district-wide seasonal malaria chemoprevention when implemented through routine malaria control programme in Kita, Mali using fixed point distribution. | 20 | 128 | RAYYAN-INCLUSION: {"Richard"=>"Excluded", "Jessica "=>"Excluded"} \| RAYYAN-EXCLUSION-REASONS: wrong outcome,wrong study design | Cost-Benefit Analysis;Chemoprevention;Seasons | PMC7934250 |
| rayyan-651229759 | Pal P\|\|Daniels BP\|\|Oskman A\|\|Diamond MS\|\|Klein RS\|\|Goldberg DE | Plasmodium falciparum Histidine-Rich Protein II Compromises Brain Endothelial Barriers and May Promote Cerebral Malaria Pathogenesis. | 7 | | RAYYAN-INCLUSION: {"Richard"=>"Excluded", "Jessica "=>"Excluded"} \| RAYYAN-EXCLUSION-REASONS: wrong outcome,wrong study design | Plasmodium falciparum;Brain | PMC4959673 |
| rayyan-651229760 | Adhikari SR | Towards universal health coverage: an example of malaria intervention in Nepal. | 3 | 103-112 | RAYYAN-INCLUSION: {"Richard"=>"Excluded", "Jessica "=>"Excluded"} \| RAYYAN-EXCLUSION-REASONS: wrong outcome,wrong study design | Nepal;Malaria | |
| rayyan-651229761 | Zinszer K\|\|Kigozi R\|\|Charland K\|\|Dorsey G\|\|Brewer TF\|\|Brownstein JS\|\|Kamya MR\|\|Buckeridge DL | Forecasting malaria in a highly endemic country using environmental and clinical predictors. | 14 | 245 | RAYYAN-INCLUSION: {"Richard"=>"Excluded", "Jessica "=>"Excluded"} \| RAYYAN-EXCLUSION-REASONS: wrong outcome,wrong study design | Malaria | PMC4470343 |
| rayyan-651229763 | Suchdev PS\|\|Addo OY\|\|Martorell R\|\|Grant FK\|\|Ruth LJ\|\|Patel MK\|\|Juliao PC\|\|Quick R\|\|Flores-Ayala R | Effects of community-based sales of micronutrient powders on morbidity episodes in preschool children in Western Kenya. | 103 | 934-41 | RAYYAN-INCLUSION: {"Richard"=>"Excluded", "Jessica "=>"Excluded"} \| RAYYAN-EXCLUSION-REASONS: wrong outcome,wrong study design,wrong infection / disease | Child, Preschool;Powders;Kenya;Child;Only Child | PMC4845747 |
| rayyan-651229764 | Hess SY\|\|Abbeddou S\|\|Jimenez EY\|\|Some JW\|\|Vosti SA\|\|Ouedraogo ZP\|\|Guissou RM\|\|Ouedraogo JB\|\|Brown KH | Small-quantity lipid-based nutrient supplements, regardless of their zinc content, increase growth and reduce the prevalence of stunting and wasting in young burkinabe children: a cluster-randomized trial. | 10 | e0122242 | RAYYAN-INCLUSION: {"Richard"=>"Excluded", "Jessica "=>"Excluded"} \| RAYYAN-EXCLUSION-REASONS: wrong outcome,wrong study design,wrong infection / disease | Prevalence;Only Child;Child;Zinc | PMC4376671 |
| rayyan-651229765 | Fernandez-Ruiz D\|\|Ng WY\|\|Holz LE\|\|Ma JZ\|\|Zaid A\|\|Wong YC\|\|Lau LS\|\|Mollard V\|\|Cozijnsen A\|\|Collins... | Liver-Resident Memory CD8(+) T Cells Form a Front-Line Defense against Malaria Liver-Stage Infection. | 45 | 889-902 | RAYYAN-INCLUSION: {"Richard"=>"Excluded", "Jessica "=>"Excluded"} \| RAYYAN-EXCLUSION-REASONS: wrong outcome,wrong study design | Liver | |
| rayyan-651229766 | Makadzange K\|\|Dlamini N\|\|Zulu Z\|\|Dlamini S\|\|Kunene S\|\|Sikhondze W\|\|Owiti P\|\|Geoffroy E\|\|Zachariah... | Low uptake of preventive interventions among malaria cases in Swaziland: towards malaria elimination. | 8 | S29-S33 | RAYYAN-INCLUSION: {"Donal"=>"Included", "Richard"=>"Excluded", "Jessica "=>"Included"} \| RAYYAN-LABELS: Include (tiebreaker) \| RAYYAN-EXCLUSION-REASONS: wrong outcome,wrong study design | Malaria | PMC5912418 |
| rayyan-651229767 | Spjeldnaes AO\|\|Kitua AY\|\|Blomberg B | Education and knowledge helps combating malaria, but not degedege: a cross-sectional study in Rufiji, Tanzania. | 13 | 200 | RAYYAN-INCLUSION: {"Richard"=>"Excluded", "Jessica "=>"Excluded"} \| RAYYAN-EXCLUSION-REASONS: wrong outcome,wrong study design | Cross-Sectional Studies;Cesarean Section;Malaria | PMC4058719 |
| rayyan-651229768 | Keen S\|\|Begum H\|\|Friedman HS\|\|James CD | Scaling up family planning in Sierra Leone: A prospective cost-benefit analysis. | 13 | 43-57 | RAYYAN-INCLUSION: {"Richard"=>"Excluded", "Jessica "=>"Excluded"} \| RAYYAN-EXCLUSION-REASONS: wrong outcome,wrong study design,wrong infection / disease | Cost-Benefit Analysis | PMC7789032 |
| rayyan-651229769 | Singh H\|\|Gupta SK\|\|Vikram K\|\|Saxena R\|\|Srivastava A\|\|Nagpal BN | Sustainable control of malaria employing Gambusia fishes as biological control in Jalore and Barmer districts of Western Rajasthan. | 59 | 91-97 | RAYYAN-INCLUSION: {"Richard"=>"Excluded", "Jessica "=>"Excluded"} \| RAYYAN-EXCLUSION-REASONS: wrong outcome,wrong study design | Malaria | |
| rayyan-651229770 | File T\|\|Dinka H\|\|Golassa L | A retrospective analysis on the transmission of Plasmodium falciparum and Plasmodium vivax: the case of Adama City, East Shoa Zone, Oromia, Ethiopia. | 18 | 193 | RAYYAN-INCLUSION: {"Richard"=>"Excluded", "Jessica "=>"Excluded"} \| RAYYAN-EXCLUSION-REASONS: wrong outcome,wrong study design | Plasmodium falciparum;Plasmodium | PMC6558791 |
| rayyan-651229771 | Sternberg ED\|\|Ng'habi KR\|\|Lyimo IN\|\|Kessy ST\|\|Farenhorst M\|\|Thomas MB\|\|Knols BG\|\|Mnyone LL | Eave tubes for malaria control in Africa: initial development and semi-field evaluations in Tanzania. | 15 | 447 | RAYYAN-INCLUSION: {"Richard"=>"Excluded", "Jessica "=>"Excluded"} \| RAYYAN-EXCLUSION-REASONS: wrong outcome,wrong study design | Malaria | PMC5009540 |
| rayyan-651229772 | Aninanya GA\|\|Williams JE\|\|Williams A\|\|Otupiri E\|\|Howard N | Effects of computerized decision support on maternal and neonatal health-worker performance in the context of combined implementation with performance-based incentivisation in Upper East Region, Ghana: a qualitative study of professional perspectives. | 22 | 1581 | RAYYAN-INCLUSION: {"Richard"=>"Excluded", "Jessica "=>"Excluded"} \| RAYYAN-EXCLUSION-REASONS: wrong outcome,wrong study design | Infant, Newborn;Ghana;Middle East;Far East | PMC9791727 |
| rayyan-651229773 | Hagenlocher M\|\|Castro MC | Mapping malaria risk and vulnerability in the United Republic of Tanzania: a spatial explicit model. | 13 | 2 | RAYYAN-INCLUSION: {"Richard"=>"Excluded", "Jessica "=>"Excluded"} \| RAYYAN-EXCLUSION-REASONS: wrong study design,wrong study duration | Malaria | PMC4324856 |
| rayyan-651229774 | Wagman JM\|\|Grieco JP\|\|Bautista K\|\|Polanco J\|\|Briceno I\|\|King R\|\|Achee NL | The field evaluation of a push-pull system to control malaria vectors in northern Belize, Central America. | 14 | 184 | RAYYAN-INCLUSION: {"Richard"=>"Excluded", "Jessica "=>"Excluded"} \| RAYYAN-EXCLUSION-REASONS: wrong outcome,wrong study design | Malaria | PMC4425932 |

<table>
<tr><td>rayyan-651229775</td><td>Hast MA||Stevenson JC||Muleba M||Chaponda M||Kabuya JB||Mulenga M||Lessler J||Shields T||Moss WJ||...</td><td>Risk Factors for Household Vector Abundance Using Indoor CDC Light Traps in a High Malaria Transmission Area of Northern Zambia.</td><td>101</td><td>126-136</td><td>RAYYAN-INCLUSION: {"Richard"=>"Excluded", "Jessica "=>"Excluded"} | RAYYAN-EXCLUSION-REASONS: wrong outcome,wrong study design</td><td>Risk Factors;Malaria</td><td>PMC6609184</td></tr>
<tr><td>rayyan-651229776</td><td>Usman-Yamman H||Omalu C J I||Abubakar A||Abolarinwa S O||Eke S S||Otuu CA</td><td>Genetic diversity of plasmodium falciparum isolates in Minna, North Central Nigeria inferred by PCR genotyping of Merozoite surface protein 1 and 2.</td><td>96</td><td>105143</td><td>RAYYAN-INCLUSION: {"Richard"=>"Excluded", "Jessica "=>"Excluded"} | RAYYAN-EXCLUSION-REASONS: wrong outcome,wrong study design</td><td>Genetic Variation;Membrane Proteins;Plasmodium falciparum;Nigeria;Genotype</td><td></td></tr>
<tr><td>rayyan-651229777</td><td>Nkosi-Gondwe T||Robberstad B||Blomberg B||Phiri KS||Lange S</td><td>Introducing post-discharge malaria chemoprevention (PMC) for management of severe anemia in Malawian children: a qualitative study of community health workers' perceptions and motivation.</td><td>18</td><td>984</td><td>RAYYAN-INCLUSION: {"Richard"=>"Excluded", "Jessica "=>"Excluded"} | RAYYAN-EXCLUSION-REASONS: wrong outcome,wrong study design</td><td>Anemia;Chemoprevention</td><td>PMC6299958</td></tr>
<tr><td>rayyan-651229778</td><td>George A||Rodriguez DC||Rasanathan K||Brandes N||Bennett S</td><td>iCCM policy analysis: strategic contributions to understanding its character, design and scale up in sub-Saharan Africa.</td><td>30</td><td>ii3-ii11</td><td>RAYYAN-INCLUSION: {"Richard"=>"Excluded", "Jessica "=>"Excluded"} | RAYYAN-EXCLUSION-REASONS: wrong outcome,background article,wrong study design,wrong publication type</td><td>Africa South of the Sahara</td><td></td></tr>
<tr><td>rayyan-651229779</td><td>Dambach P||Traore I||Kaiser A||Sie A||Sauerborn R||Becker N</td><td>Challenges of implementing a large scale larviciding campaign against malaria in rural Burkina Faso - lessons learned and recommendations derived from the EMIRA project.</td><td>16</td><td>1023</td><td>RAYYAN-INCLUSION: {"Richard"=>"Excluded", "Jessica "=>"Excluded"} | RAYYAN-EXCLUSION-REASONS: wrong outcome,wrong study design</td><td>Malaria;Learning</td><td>PMC5041282</td></tr>
<tr><td>rayyan-651229780</td><td>Pretorius E||Kristan M||Bradley J||da Silva ET||Hutchins H||Barri F||Cassama A||Ceesay S||Ndiath ...</td><td>Validation of a method for the dry preservation and rehydration of Anopheles gambiae sensu lato for parity analysis to assess the impact of vector control measures in the field.</td><td>16</td><td>236</td><td>RAYYAN-INCLUSION: {"Richard"=>"Excluded", "Jessica "=>"Excluded"} | RAYYAN-EXCLUSION-REASONS: wrong outcome,wrong study design</td><td>Fluid Therapy</td><td>PMC10349412</td></tr>
<tr><td>rayyan-651229781</td><td>Mukonka VM||Chanda E||Haque U||Kamuliwo M||Mushinge G||Chileshe J||Chibwe KA||Norris DE||Mulenga ...</td><td>High burden of malaria following scale-up of control interventions in Nchelenge District, Luapula Province, Zambia.</td><td>13</td><td>153</td><td>RAYYAN-INCLUSION: {"Richard"=>"Included", "Jessica "=>"Included"} | RAYYAN-LABELS: Excluded (outside study period)</td><td>Malaria</td><td>PMC4016669</td></tr>
<tr><td>rayyan-651229782</td><td>Abaza H||Marschollek M</td><td>mHealth Application Areas and Technology Combinations*. A Comparison of Literature from High and Low/Middle Income Countries.</td><td>56</td><td>e105-e122</td><td>RAYYAN-INCLUSION: {"Richard"=>"Excluded", "Jessica "=>"Excluded"} | RAYYAN-EXCLUSION-REASONS: wrong outcome,background article,wrong study design</td><td></td><td>PMC6291822</td></tr>
<tr><td>rayyan-651229783</td><td>Dhiman RC||Sarkar S</td><td>El Nino Southern Oscillation as an early warning tool for malaria outbreaks in India.</td><td>16</td><td>122</td><td>RAYYAN-INCLUSION: {"Richard"=>"Excluded", "Jessica "=>"Excluded"} | RAYYAN-EXCLUSION-REASONS: wrong outcome,wrong study design</td><td>Disease Outbreaks;India;Malaria</td><td>PMC5359847</td></tr>
<tr><td>rayyan-651229784</td><td>Helinski ME||Nuwa A||Protopopoff N||Feldman M||Ojuka P||Oguttu DW||Abeku TA||Meek S</td><td>Entomological surveillance following a long-lasting insecticidal net universal coverage campaign in Midwestern Uganda.</td><td>8</td><td>458</td><td>RAYYAN-INCLUSION: {"Richard"=>"Excluded", "Jessica "=>"Excluded"} | RAYYAN-EXCLUSION-REASONS: wrong outcome,wrong study design</td><td>Uganda;Insecticides</td><td>PMC4574096</td></tr>
<tr><td>rayyan-651229785</td><td>Masquelier B||Waltisperger D||Ralijaona O||Pison G||Ravelo A</td><td>The epidemiological transition in Antananarivo, Madagascar: an assessment based on death registers (1900-2012).</td><td>7</td><td>23237</td><td>RAYYAN-INCLUSION: {"Richard"=>"Excluded", "Jessica "=>"Excluded"} | RAYYAN-EXCLUSION-REASONS: wrong outcome,wrong study design,wrong study duration</td><td></td><td>PMC4028933</td></tr>
<tr><td>rayyan-651229786</td><td>Bempah S||Curtis A||Awandare G||Ajayakumar J</td><td>Appreciating the complexity of localized malaria risk in Ghana: Spatial data challenges and solutions.</td><td>64</td><td>102382</td><td>RAYYAN-INCLUSION: {"Richard"=>"Excluded", "Jessica "=>"Excluded"} | RAYYAN-EXCLUSION-REASONS: wrong outcome,wrong study design</td><td>Ghana;Malaria</td><td></td></tr>
<tr><td>rayyan-651229787</td><td>Amouzou A||Morris S||Moulton LH||Mukanga D</td><td>Assessing the impact of integrated community case management (iCCM) programs on child mortality: Review of early results and lessons learned in sub-Saharan Africa.</td><td>4</td><td>020411</td><td>RAYYAN-INCLUSION: {"Richard"=>"Excluded", "Jessica "=>"Excluded"} | RAYYAN-EXCLUSION-REASONS: wrong outcome,background article,wrong study design</td><td>Case Management;Africa South of the Sahara</td><td>PMC4267100</td></tr>
<tr><td>rayyan-651229788</td><td>Ssempiira J||Nambuusi B||Kissa J||Agaba B||Makumbi F||Kasasa S||Vounatsou P</td><td>Geostatistical modelling of malaria indicator survey data to assess the effects of interventions on the geographical distribution of malaria prevalence in children less than 5 years in Uganda.</td><td>12</td><td>e0174948</td><td>RAYYAN-INCLUSION: {"Richard"=>"Excluded", "Jessica "=>"Excluded"} | RAYYAN-EXCLUSION-REASONS: wrong study design | USER-NOTES: {"Richard"=>["Unclear if also routine data was used", "Seems that MIS data only was used", "Seems that MIS data only was used"]}</td><td>Uganda;Prevalence;Malaria</td><td>PMC5380319</td></tr>
<tr><td>rayyan-651229789</td><td>Rusk A||Highfield L||Wilkerson JM||Harrell M||Obala A||Amick B</td><td>Geographically-weighted regression of knowledge and behaviour determinants to anti-malarial recommending and dispensing practice among medicine retailers in western Kenya: capacitating targeted interventions.</td><td>15</td><td>562</td><td>RAYYAN-INCLUSION: {"Richard"=>"Excluded", "Jessica "=>"Excluded"} | RAYYAN-EXCLUSION-REASONS: wrong outcome,wrong study design</td><td>Antimalarials;Kenya</td><td>PMC5117492</td></tr>
<tr><td>rayyan-651229790</td><td>Baliga BS||Jain A||Koduvattat N||Kumar BGP||Kumar M||Kumar A||Ghosh SK</td><td>Indigenously developed digital handheld Android-based Geographic Information System (GIS)-tagged tablets (TABs) in malaria elimination programme in Mangaluru city, Karnataka, India.</td><td>18</td><td>444</td><td>RAYYAN-INCLUSION: {"Richard"=>"Excluded", "Jessica "=>"Excluded"} | RAYYAN-EXCLUSION-REASONS: wrong outcome,wrong study design</td><td>Poverty;Methyltestosterone;Information Systems;India;Malaria</td><td>PMC6933888</td></tr>
<tr><td>rayyan-651229791</td><td>Horn S||Snoep JL||van Niekerk DD</td><td>Uncovering the effects of heterogeneity and parameter sensitivity on within-host dynamics of disease: malaria as a case study.</td><td>22</td><td>384</td><td>RAYYAN-INCLUSION: {"Richard"=>"Excluded", "Jessica "=>"Excluded"} | RAYYAN-EXCLUSION-REASONS: wrong outcome,wrong study design</td><td>Malaria</td><td>PMC8305899</td></tr>
<tr><td>rayyan-651229792</td><td>Siddiqui AJ||Bhardwaj J||Goyal M||Prakash K||Soni A||Tiwari V||Puri SK</td><td>Assessment of real-time method to detect liver parasite burden under different experimental conditions in mice infected with Plasmodium yoelii sporozoites.</td><td>89</td><td>35-42</td><td>RAYYAN-INCLUSION: {"Richard"=>"Excluded", "Jessica "=>"Excluded"} | RAYYAN-EXCLUSION-REASONS: wrong outcome,wrong study design</td><td>Liver</td><td></td></tr>
<tr><td>rayyan-651229793</td><td>Verguet S||Olson ZD||Babigumira JB||Desalegn D||Johansson KA||Kruk ME||Levin CE||Nugent RA||Pecen...</td><td>Health gains and financial risk protection afforded by public financing of selected interventions in Ethiopia: an extended cost-effectiveness analysis.</td><td>3</td><td>e288-96</td><td>RAYYAN-INCLUSION: {"Donal"=>"Excluded", "Richard"=>"Excluded", "Jessica "=>"Maybe"} | RAYYAN-LABELS: Excluded (tiebreaker) | RAYYAN-EXCLUSION-REASONS: wrong outcome,wrong study design</td><td>Cost-Benefit Analysis;Financing, Government</td><td></td></tr>
<tr><td>rayyan-651229794</td><td>Pemberton-Ross P||Smith TA||Hodel EM||Kay K||Penny MA</td><td>Age-shifting in malaria incidence as a result of induced immunological deficit: a simulation study.</td><td>14</td><td>287</td><td>RAYYAN-INCLUSION: {"Richard"=>"Excluded", "Jessica "=>"Excluded"} | RAYYAN-EXCLUSION-REASONS: wrong outcome,wrong study design</td><td>Malaria</td><td>PMC4513612</td></tr>
<tr><td>rayyan-651229795</td><td>Mwingira F||Genton B||Kabanywanyi AN||Felger I</td><td>Comparison of detection methods to estimate asexual Plasmodium falciparum parasite prevalence and gametocyte carriage in a community survey in Tanzania.</td><td>13</td><td>433</td><td>RAYYAN-INCLUSION: {"Richard"=>"Excluded", "Jessica "=>"Excluded"} | RAYYAN-EXCLUSION-REASONS: wrong outcome,wrong study design</td><td>Prevalence;Plasmodium falciparum</td><td>PMC4246543</td></tr>
<tr><td>rayyan-651229796</td><td>Kweka EJ||Munga S||Himeidan Y||Githeko AK||Yan G</td><td>Assessment of mosquito larval productivity among different land use types for targeted malaria vector control in the western Kenya highlands.</td><td>8</td><td>356</td><td>RAYYAN-INCLUSION: {"Richard"=>"Excluded", "Jessica "=>"Excluded"} | RAYYAN-EXCLUSION-REASONS: wrong outcome,wrong study design</td><td>Kenya;Culicidae;Malaria</td><td>PMC4491214</td></tr>
<tr><td>rayyan-651229797</td><td>Yang X||Liu J||Zhou XN||Cheung WK</td><td>Inferring disease transmission networks at a metapopulation level.</td><td>2</td><td>8</td><td>RAYYAN-INCLUSION: {"Richard"=>"Excluded", "Jessica "=>"Excluded"} | RAYYAN-EXCLUSION-REASONS: wrong outcome,wrong study design</td><td></td><td>PMC4375841</td></tr>
<tr><td>rayyan-651229798</td><td>Brindle E||Lillis L||Barney R||Hess SY||Wessells KR||Ouedraogo CT||Stinca S||Kalnoky M||Peck R||T...</td><td>Simultaneous assessment of iodine, iron, vitamin A, malarial antigenemia, and inflammation status biomarkers via a multiplex immunoassay method on a population of pregnant women from Niger.</td><td>12</td><td>e0185868</td><td>RAYYAN-INCLUSION: {"Richard"=>"Excluded", "Jessica "=>"Excluded"} | RAYYAN-EXCLUSION-REASONS: wrong outcome,wrong study design</td><td>Vitamins</td><td>PMC5628875</td></tr>
<tr><td>rayyan-651229799</td><td>Shamu S||Rusakaniko S||Hongoro C</td><td>Prioritizing health system and disease burden factors: an evaluation of the net benefit of transferring health technology interventions to different districts in Zimbabwe.</td><td>8</td><td>695-705</td><td>RAYYAN-INCLUSION: {"Richard"=>"Excluded", "Jessica "=>"Excluded"} | RAYYAN-EXCLUSION-REASONS: wrong outcome,wrong study design,wrong infection / disease</td><td></td><td>PMC5125992</td></tr>
<tr><td>rayyan-651229800</td><td>Katureebe A||Zinszer K||Arinaitwe E||Rek J||Kakande E||Charland K||Kigozi R||Kilama M||Nankabirwa...</td><td>Measures of Malaria Burden after Long-Lasting Insecticidal Net Distribution and Indoor Residual Spraying at Three Sites in Uganda: A Prospective Observational Study.</td><td>13</td><td>e1002167</td><td>RAYYAN-INCLUSION: {"Donal"=>"Included", "Richard"=>"Included", "Jessica "=>"Maybe"} | RAYYAN-LABELS: Include (tiebreaker)</td><td>Uganda;Malaria;Insecticides</td><td>PMC5100985</td></tr>
<tr><td>rayyan-651229801</td><td>Aleman Resto Y||Fernandez Robledo JA</td><td>Identification of MMV Malaria Box inhibitors of Perkinsus marinus using an ATP-based bioluminescence assay.</td><td>9</td><td>e111051</td><td>RAYYAN-INCLUSION: {"Richard"=>"Excluded", "Jessica "=>"Excluded"} | RAYYAN-EXCLUSION-REASONS: wrong outcome,wrong study design</td><td>Malaria</td><td>PMC4206467</td></tr>
<tr><td>rayyan-651229802</td><td>Mweresa CK||Mukabana WR||Omusula P||Otieno B||Gheysens T||Takken W||van Loon JJ</td><td>Evaluation of textile substrates for dispensing synthetic attractants for malaria mosquitoes.</td><td>7</td><td>376</td><td>RAYYAN-INCLUSION: {"Richard"=>"Excluded", "Jessica "=>"Excluded"} | RAYYAN-EXCLUSION-REASONS: wrong outcome,wrong study design</td><td>Culicidae;Malaria</td><td>PMC4152566</td></tr>
<tr><td>rayyan-651229803</td><td>Fernando SD||Ihalamulla RL||Wickremasinghe R||de Silva NL||Thilakarathne JH||Wijeyaratne P||Prema...</td><td>Effects of modifying the World Health Organization standard operating procedures for malaria microscopy to improve surveillance in resource poor settings.</td><td>13</td><td>98</td><td>RAYYAN-INCLUSION: {"Richard"=>"Excluded", "Jessica "=>"Excluded"} | RAYYAN-EXCLUSION-REASONS: wrong outcome,wrong study design</td><td>Surgical Procedures, Operative;Malaria</td><td>PMC3995539</td></tr>
<tr><td>rayyan-651229804</td><td>Sluydts V||Durnez L||Heng S||Gryseels C||Canier L||Kim S||Van Roey K||Kerkhof K||Khim N||Mao S||U...</td><td>Efficacy of topical mosquito repellent (picaridin) plus long-lasting insecticidal nets versus long-lasting insecticidal nets alone for control of malaria: a cluster randomised controlled trial.</td><td>16</td><td>1169-1177</td><td>RAYYAN-INCLUSION: {"Richard"=>"Excluded", "Jessica "=>"Excluded"} | RAYYAN-EXCLUSION-REASONS: wrong study design</td><td>Culicidae;Insecticides;Malaria</td><td></td></tr>
<tr><td>rayyan-651229805</td><td>Roy M||Bouma M||Dhiman RC||Pascual M</td><td>Predictability of epidemic malaria under non-stationary conditions with process-based models combining epidemiological updates and climate variability.</td><td>14</td><td>419</td><td>RAYYAN-INCLUSION: {"Donal"=>"Excluded", "Richard"=>"Maybe", "Jessica "=>"Excluded"} | RAYYAN-LABELS: Excluded (tiebreaker) | USER-NOTES: {"Richard"=>["Unclear if/how HMIS data was used"]}</td><td>Malaria</td><td>PMC4623260</td></tr>
<tr><td>rayyan-651229806</td><td>Okwaro FM||Chandler CI||Hutchinson E||Nabirye C||Taaka L||Kayendeke M||Nayiga S||Staedke SG</td><td>Challenging logics of complex intervention trials: community perspectives of a health care improvement intervention in rural Uganda.</td><td>131</td><td>7-Oct</td><td>RAYYAN-INCLUSION: {"Richard"=>"Excluded", "Jessica "=>"Excluded"} | RAYYAN-EXCLUSION-REASONS: wrong outcome,wrong study design,wrong study duration</td><td>Uganda</td><td></td></tr>
</table>

| | | | | | | | | | | | |
|---|---|---|---|---|---|---|---|---|---|---|---|
| rayyan-651229807 | Muhwezi WW\|\|Palchik EA\|\|Kiwanuka DH\|\|Mpanga F\|\|Mukundane M\|\|Nanungi A\|\|Bataringaya D\|\|Ssesanga P\|... | Community participation to improve health services for children: a methodology for a community dialogue intervention in Uganda. | | | 19 | 1574-1581 | RAYYAN-INCLUSION: {"Richard"=>"Excluded", "Jessica "=>"Excluded"} \| RAYYAN-EXCLUSION-REASONS: wrong outcome,wrong study design | | Uganda | PMC6531984 | |
| rayyan-651229808 | Mbonye AK\|\|Clarke SE\|\|Lal S\|\|Chandler CI\|\|Hutchinson E\|\|Hansen KS\|\|Magnussen P | Introducing rapid diagnostic tests for malaria into registered drug shops in Uganda: lessons learned and policy implications. | | | 14 | 448 | RAYYAN-INCLUSION: {"Richard"=>"Excluded", "Jessica "=>"Excluded"} \| RAYYAN-EXCLUSION-REASONS: wrong outcome,wrong study design | | Uganda;Malaria | PMC4647310 | |
| rayyan-651229809 | Lynd A\|\|Gonahasa S\|\|Staedke SG\|\|Oruni A\|\|Maiteki-Sebuguzi C\|\|Dorsey G\|\|Opigo J\|\|Yeka A\|\|Katureebe... | LLIN Evaluation in Uganda Project (LLINEUP): a cross-sectional survey of species diversity and insecticide resistance in 48 districts of Uganda. | | | 12 | 94 | RAYYAN-INCLUSION: {"Richard"=>"Excluded", "Jessica "=>"Excluded"} \| RAYYAN-EXCLUSION-REASONS: wrong outcome,wrong study design | | Uganda;Cross-Sectional Studies;Cesarean Section;Insecticides | PMC6417037 | |
| rayyan-651229810 | Sibanda M\|\|Focke W\|\|Braack L\|\|Leuteritz A\|\|Brunig H\|\|Tran NHA\|\|Wieczorek F\|\|Trumper W | Bicomponent fibres for controlled release of volatile mosquito repellents. | | | 91 | 754-761 | RAYYAN-INCLUSION: {"Richard"=>"Excluded", "Jessica "=>"Excluded"} \| RAYYAN-EXCLUSION-REASONS: wrong outcome,wrong study design | | Culicidae;Volatilization | | |
| rayyan-651229811 | Yohannes YB\|\|Ikenaka Y\|\|Ito G\|\|Nakayama SMM\|\|Mizukawa H\|\|Wepener V\|\|Smit NJ\|\|Van Vuren JHJ\|\|Ishiz... | Assessment of DDT contamination in house rat as a possible bioindicator in DDT-sprayed areas from Ethiopia and South Africa. | | | 24 | 23763-23770 | RAYYAN-INCLUSION: {"Richard"=>"Excluded", "Jessica "=>"Excluded"} \| RAYYAN-EXCLUSION-REASONS: wrong outcome,wrong study design | | South Africa;Rats | | |
| rayyan-651229812 | Zhang T\|\|Zhang X\|\|Lin C\|\|Wu S\|\|Wang F\|\|Wang H\|\|Wang Y\|\|Peng Y\|\|Hutchinson MR\|\|Li H\|\|Wang X | Artemisinin inhibits TLR4 signaling by targeting co-receptor MD2 in microglial BV-2 cells and prevents lipopolysaccharide-induced blood-brain barrier leakage in mice. | | | 157 | 611-623 | RAYYAN-INCLUSION: {"Richard"=>"Excluded", "Jessica "=>"Excluded"} \| RAYYAN-EXCLUSION-REASONS: wrong outcome,wrong study design | | Blood-Brain Barrier;Artemisinins;Brain | | |
| rayyan-651229813 | Njoroge H\|\|Van't Hof A\|\|Oruni A\|\|Pipini D\|\|Nagi SC\|\|Lynd A\|\|Lucas ER\|\|Tomlinson S\|\|Grau-Bove X\|\|M... | Identification of a rapidly-spreading triple mutant for high-level metabolic insecticide resistance in Anopheles gambiae provides a real-time molecular diagnostic for antimalarial intervention deployment. | | | 31 | 4307-4318 | RAYYAN-INCLUSION: {"Richard"=>"Excluded", "Jessica "=>"Excluded"} \| RAYYAN-EXCLUSION-REASONS: wrong outcome,wrong study design | | Antimalarials | PMC9424592 | |
| rayyan-651229814 | Ruktanonchai NW\|\|Bhavnani D\|\|Sorichetta A\|\|Bengtsson L\|\|Carter KH\|\|Cordoba RC\|\|Le Menach A\|\|Lu X\|... | Census-derived migration data as a tool for informing malaria elimination policy. | | | 15 | 273 | RAYYAN-INCLUSION: {"Richard"=>"Excluded", "Jessica "=>"Excluded"} \| RAYYAN-EXCLUSION-REASONS: wrong outcome,wrong study design,wrong study duration | | Malaria | PMC4864939 | |
| rayyan-651229815 | Rorick MM\|\|Baskerville EB\|\|Rask TS\|\|Day KP\|\|Pascual M | Identifying functional groups among the diverse, recombining antigenic var genes of the malaria parasite Plasmodium falciparum from a local community in Ghana. | | | 14 | e1006174 | RAYYAN-INCLUSION: {"Richard"=>"Excluded", "Jessica "=>"Excluded"} \| RAYYAN-EXCLUSION-REASONS: wrong outcome,wrong study design | | | PMC6016947 | |
| rayyan-651229816 | Mengistu B\|\|Karim AM\|\|Eniyew A\|\|Yitabrek A\|\|Eniyew A\|\|Tsegaye S\|\|Muluye F\|\|Tesfaye H\|\|Demeke B\|\|M... | Effect of performance review and clinical mentoring meetings (PRCMM) on recording of community case management by health extension workers in Ethiopia. | | | 52 | 73-81 | RAYYAN-INCLUSION: {"Richard"=>"Excluded", "Jessica "=>"Excluded"} \| RAYYAN-EXCLUSION-REASONS: wrong outcome,wrong study design,wrong study duration | | Case Management | | |
| rayyan-651229817 | Kraft TE\|\|Heitmeier MR\|\|Putanko M\|\|Edwards RL\|\|Ilagan MX\|\|Payne MA\|\|Autry JM\|\|Thomas DD\|\|Odom AR\|... | A Novel Fluorescence Resonance Energy Transfer-Based Screen in High-Throughput Format To Identify Inhibitors of Malarial and Human Glucose Transporters. | | | 60 | 7407-7414 | RAYYAN-INCLUSION: {"Richard"=>"Excluded", "Jessica "=>"Excluded"} \| RAYYAN-EXCLUSION-REASONS: wrong outcome,wrong study design | | Humanities;Humanism;Humans;Energy Transfer | PMC5119023 | |
| rayyan-651229818 | Li H\|\|van der Linden WA\|\|Verdoes M\|\|Florea BI\|\|McAllister FE\|\|Govindaswamy K\|\|Elias JE\|\|Bhanot P\|... | Assessing subunit dependency of the Plasmodium proteasome using small molecule inhibitors and active site probes. | | | 9 | 1869-76 | RAYYAN-INCLUSION: {"Richard"=>"Excluded", "Jessica "=>"Excluded"} \| RAYYAN-EXCLUSION-REASONS: wrong outcome,wrong study design | | | PMC4136710 | |
| rayyan-651229819 | Jaeger MS\|\|Briet OJ\|\|Keating J\|\|Ahorlu CK\|\|Yukich JO\|\|Oppong S\|\|Nardini P\|\|Pfeiffer C | Perceptions on the effect of small electric fans on comfort inside bed nets in southern Ghana: a qualitative study. | | | 15 | 580 | RAYYAN-INCLUSION: {"Richard"=>"Excluded", "Jessica "=>"Excluded"} \| RAYYAN-EXCLUSION-REASONS: wrong outcome,wrong study design | | Ghana | PMC5134074 | |
| rayyan-751570029 | Raele DA and Severini F and Toma L and Menegon M and Boccolini D and Tortorella G and Di Luca M and Cafiero MA | Anopheles sacharovi in Italy: first record of the historical malaria vector after over 50Â years. | 2024 | Parasites & vectors | 17 | 182 | RAYYAN-INCLUSION: {"Richard"=>"Excluded"} \| RAYYAN-EXCLUSION-REASONS: wrong outcome,wrong study design \| USER-NOTES: {"Richard"=>["Ento study"]} | 10.1186/s13071-024-06252-2 | Animals;*Malaria/epidemiology;*Anopheles/genetics;Mosquito | PMC11005165 | https://pubmed.ncbi.nlm.nih.gov/38600589/ |
| rayyan-751570030 | Dhawan S and Dittrich S and Arafah S and Ongarello S and Mace A and Panapruksachat S and Boutthasavong L and Adsamouth A and Thongpaseuth S and Davong V and | Diagnostic accuracy of DPP Fever Panel II Asia tests for tropical fever diagnosis. | 2024 | PLoS neglected tropical | 18 | e0012077 | RAYYAN-INCLUSION: {"Richard"=>"Excluded"} \| RAYYAN-EXCLUSION-REASONS: wrong outcome,wrong study design,wrong infection / disease \| USER-NOTES: {"Richard"=>["Non malaria study"]} | 10.1371/journal.pntd.0012077 | Humans;*Sensitivity and Specificity;*Immunoglobulin M/blood;Female;Male;Laos;Ad | PMC11034646 | https://pubmed.ncbi.nlm.nih.gov/38598549/ |
| rayyan-751570031 | Aheto JMK and Menezes LJ and Takramah W and Cui L | Modelling spatiotemporal variation in under-five malaria risk in Ghana in 2016-2021. | 2024 | Malaria journal | 23 | 102 | RAYYAN-INCLUSION: {"Richard"=>"Excluded"} \| RAYYAN-EXCLUSION-REASONS: wrong outcome,wrong study design \| USER-NOTES: {"Richard"=>["No intervention data"]} | 10.1186/s12936-024-04918-x | Male;Child;Humans;Ghana/epidemiology;Bayes Theorem;*Malaria/epidemiolo | PMC11005246 | https://pubmed.ncbi.nlm.nih.gov/38594716/ |
| rayyan-751570032 | Rerolle F and Jacobson JO and Smith Gueye C and Bennett A and Carrillo S and Ntuku H and Smith JL | Estimating the size of populations at risk for malaria: a case study in cattle herders and agricultural workers in Northern Namibia. | 2024 | Scientific reports | 14 | 7160 | RAYYAN-INCLUSION: {"Richard"=>"Excluded"} \| RAYYAN-EXCLUSION-REASONS: wrong outcome,wrong study design \| USER-NOTES: {"Richard"=>["Risk factor / population study"]} | 10.1038/s41598-024-56810-y | Cattle;Animals;Humans;Namibia/epidemiology;*Farmers;*Malaria/epidemiology;Risk | PMC10965975 | https://pubmed.ncbi.nlm.nih.gov/38531921/ |
| rayyan-751570033 | Doucoure M and Zeguime A and Niangaly A and Guindo MA and Doritchamou JYA and Assadou MH and Katile A and Kanoute MB and Perou S and Ouattara A and Sanogo S and | Normal Clinical Laboratory Ranges by Age and Sex, and Impact on Study Screening Outcomes in Rural Mali. | 2024 | The American journal of | | | RAYYAN-INCLUSION: {"Richard"=>"Excluded"} \| RAYYAN-EXCLUSION-REASONS: wrong outcome,wrong study design \| USER-NOTES: {"Richard"=>["Immunological study"]} | 10.4269/ajtmh.23-0719 | | | https://pubmed.ncbi.nlm.nih.gov/38531104/ |
| rayyan-751570034 | Skjefte M and Cooper S and Poyer S and LourenÃ§o C and Smedinghoff S and Keller B and Wambua T and Oduor C and Frade S and Waweru W | Use of a health worker-targeted smartphone app to support quality malaria RDT implementation in Busia County, Kenya: A feasibility and acceptability study. | 2024 | PloS one | 19 | e0295049 | RAYYAN-INCLUSION: {"Richard"=>"Excluded"} \| RAYYAN-EXCLUSION-REASONS: wrong outcome,wrong study design \| USER-NOTES: {"Richard"=>["No intervention included"]} | 10.1371/journal.pone.0295049 | Humans;*Mobile Applications;Kenya;Feasibility Studies;Pilot | PMC10965099 | https://pubmed.ncbi.nlm.nih.gov/38530827/ |
| rayyan-751570035 | Sampa M and Fisa R and Mukuma C and Mwanza M and Hamainza B and Musonda P | The effect of COVID-19 on malaria cases in Zambia: a mixed effect multilevel analysis. | 2024 | Malaria journal | 23 | 85 | RAYYAN-INCLUSION: {"Richard"=>"Maybe"} \| RAYYAN-LABELS: Excluded (C19) \| USER-NOTES: {"Richard"=>["Not an intervention study. COVID-19 is the \"intervention\""]} | 10.1186/s12936-024-04882-6 | Humans;Adolescent;Zambia/epidemiology;*COVID-19/epidemiology;Cross- | PMC10958942 | https://pubmed.ncbi.nlm.nih.gov/38519929/ |
| rayyan-751570036 | Hilton ER and Gning-Cisse N and Assi A and Eyakou M and Koffi J and Gnakou B and Kouassi B and Flatley C and Chabi J and Gbalegba C and Alex Aimain S and Yah Kokrasset C and | Reduction of malaria case incidence following the introduction of clothianidin-based indoor residual spraying in previously unsprayed districts: an observational analysis using health facility register data from CÃ´te d'Ivoire, 2018-2022. | 2024 | BMJ global health | 9 | | RAYYAN-INCLUSION: {"Richard"=>"Included"} | 10.1136/bmjgh-2023-013324 | Humans;Incidence;Mosquito Control;Cote d'Ivoire/epidemiology;Neonico | PMC10961507 | https://pubmed.ncbi.nlm.nih.gov/38519096/ |
| rayyan-751570037 | Nengnong CB and Passah M and Wilson ML and Bellotti E and Kessler A and Marak BR and Carlton JM and Sarkar R and Albert S | Community and health worker perspectives on malaria in Meghalaya, India: covering the last mile of elimination by 2030. | 2024 | Malaria journal | 23 | 83 | RAYYAN-INCLUSION: {"Richard"=>"Excluded"} \| RAYYAN-EXCLUSION-REASONS: wrong outcome,wrong study design \| USER-NOTES: {"Richard"=>["Health systems study qualitative"]} | 10.1186/s12936-024-04905-2 | Animals;Humans;Health Personnel;*Malaria, Cerebral;India/epidemiology;H | PMC10949573 | https://pubmed.ncbi.nlm.nih.gov/38500097/ |
| rayyan-751570038 | Elias S and Massebo F | Plasmodium falciparum remains the dominant parasite affecting children despite decades of implementing vector control in two villages of Wolaita Zone, Southwest Ethiopia. | 2023 | Frontiers in epidemiol | 3 | 1305074 | RAYYAN-INCLUSION: {"Richard"=>"Excluded"} \| RAYYAN-EXCLUSION-REASONS: wrong study design \| USER-NOTES: {"Richard"=>["No routine data used"]} | 10.3389/fepid.2023.1305074 | Plasmodium falciparum;Only Child;Child | PMC10911012 | https://pubmed.ncbi.nlm.nih.gov/38455893/ |
| rayyan-751570039 | Brito M and Rufatto R and Brito-Sousa JD and Murta F and Sampaio V and Balieiro P and BaÃ-a-Silva D and Castro V and Alves B and Alencar A and Duparc S and Grewal Daumerie P | Operational effectiveness of tafenoquine and primaquine for the prevention of Plasmodium vivax recurrence in Brazil: a retrospective observational study. | 2024 | The Lancet. Infectious | | | RAYYAN-INCLUSION: {"Richard"=>"Included"} | 10.1016/S1473-3099(24)00074-4 | Recurrence;Brazil;Plasmodium | | https://pubmed.ncbi.nlm.nih.gov/38452779/ |
| rayyan-751570040 | Chawla K and Khanna V and Ayer VS and Khanna R | Comparative evaluation of traditional and molecular diagnostic methods for malaria: An analysis of performance. | 2024 | Tropical parasitology | 14 | 30-35 | RAYYAN-INCLUSION: {"Richard"=>"Excluded"} \| RAYYAN-EXCLUSION-REASONS: wrong outcome,background article,wrong study design \| USER-NOTES: {"Richard"=>["Diagnostic study"]} | 10.4103/tp.tp_38_23 | Malaria | PMC10911188 | https://pubmed.ncbi.nlm.nih.gov/38444788/ |
| rayyan-751570041 | Gari T and LindtjÃ¸rn B | Insecticide-treated bed nets and residual indoor spraying reduce malaria in areas with low transmission: a reanalysis of the Maltrials study. | 2024 | Malaria journal | 23 | 67 | RAYYAN-INCLUSION: {"Richard"=>"Maybe"} \| RAYYAN-LABELS: Excluded (after full text) \| USER-NOTES: {"Richard"=>["Not clear if routine data used"]} | 10.1186/s12936-024-04894-2 | Animals;*Insecticide-Treated Bednets;*Culicidae;Environment;Ethiopia;*Insecticides;*Mal | PMC10913548 | https://pubmed.ncbi.nlm.nih.gov/38439099/ |
| rayyan-751570042 | Bisanzio D and Keita MS and Camara A and Guilavogui T and Diallo T and Barry H and Preston A and Bangoura L and Mbounga E and Florey LS and Taton JL and Fofana A and | Malaria trends in districts that were targeted and not-targeted for seasonal malaria chemoprevention in children under 5 years of age in Guinea, 2014-2021. | 2024 | BMJ global health | 9 | | RAYYAN-INCLUSION: {"Richard"=>"Included"} | 10.1136/bmjgh-2023-013898 | Child;Humans;Child, Preschool;*Antimalarials/therapeutic | PMC10900330 | https://pubmed.ncbi.nlm.nih.gov/38413098/ |
| rayyan-751570046 | Kamau L and Bennett KL and Ochomo E and Herren J and Agumba S and Otieno S and Omoke D and Matoke-Muhia D and Mburu D and Mwangangi J and Ramaita E and Juma EO | The Anopheles coluzzii range extends into Kenya: Detection, insecticide resistance profiles and population genetic structure in relation to conspecific populations in West and Central Africa. | 2024 | Research square | | | RAYYAN-INCLUSION: {"Richard"=>"Excluded"} \| RAYYAN-EXCLUSION-REASONS: wrong outcome,wrong study design \| USER-NOTES: {"Richard"=>["Ento study"]} | 10.21203/rs.3.rs-3953608/v1 | Kenya;Insecticides | PMC10896386 | https://pubmed.ncbi.nlm.nih.gov/38410447/ |
| rayyan-751570049 | Degefa T and Yewhalaw D and Yan G | Methods of sampling malaria vectors and their reliability in estimating entomological indices in Africa. | 2024 | Journal of medical entomolo | | | RAYYAN-INCLUSION: {"Richard"=>"Excluded"} \| RAYYAN-EXCLUSION-REASONS: wrong outcome,wrong study design \| USER-NOTES: {"Richard"=>["Ento study"]} | 10.1093/jme/tjae015 | Malaria;Africa | | https://pubmed.ncbi.nlm.nih.gov/38394375/ |
| rayyan-751570056 | Chiphwanya J and Mkwanda S and Kabuluzi S and Mzilahowa T and Ngwira B and Matipula DE and Chaponda L and Ndhlova P and Katchika P and Mahebere Chirambo C and Moses P and | Elimination of lymphatic filariasis as a public health problem in Malawi. | 2024 | PLoS neglected tropical | 18 | e0011957 | RAYYAN-INCLUSION: {"Richard"=>"Excluded"} \| RAYYAN-EXCLUSION-REASONS: wrong infection / disease \| USER-NOTES: {"Richard"=>["LF not malaria"]} | 10.1371/journal.pntd.0011957 | Animals;Humans;*Elephantiasis, Filarial/drug therapy/epidemiology/prevent | PMC10903958 | https://pubmed.ncbi.nlm.nih.gov/38363794/ |
| rayyan-751570061 | Srivastava B and Sharma S and Swarnkar D and Ahmed N and Valecha N and Anvikar AR | Benefits of Lot Testing to Improve the Quality of Malaria Rapid Diagnostic Tests in India. | 2024 | The American journal of | 110 | 431-435 | RAYYAN-INCLUSION: {"Richard"=>"Excluded"} \| RAYYAN-EXCLUSION-REASONS: wrong outcome,wrong study design \| USER-NOTES: {"Richard"=>["Diagnostic study"]} | 10.4269/ajtmh.23-0339 | Humans;*Malaria, Falciparum/diagnosis;Plasmodium falciparum;Rapid | PMC10919188 | https://pubmed.ncbi.nlm.nih.gov/38350136/ |
| rayyan-751570067 | Malm KL and Peprah NY and Mohammed W and Adomako BY and Oppong S and Boateng P and Adu GA and Dadzie D and Adza G and Forson I and Frimpong J | A retrospective analysis of malaria deaths in the pre- and intra- COVID 19 pandemic era, Ghana, 2016-2021. | 2024 | PloS one | 19 | e0286212 | RAYYAN-INCLUSION: {"Richard"=>"Maybe"} \| RAYYAN-LABELS: Excluded (C19) \| USER-NOTES: {"Richard"=>["COVID-19 is the \"intervention\""]} | 10.1371/journal.pone.0286212 | Child;Humans;Child, Preschool;*COVID-19/epidemiology;Ghana/epide | PMC10846731 | https://pubmed.ncbi.nlm.nih.gov/38319929/ |

| | | | | | | | | | | | |
|---|---|---|---|---|---|---|---|---|---|---|---|
| rayyan-751570071 | Thomas A and Bakai TA and Atcha-Oubou T and Tchadjobo T and Rabilloud M and Voirin N | Exploring malaria prediction models in Togo: a time series forecasting by health district and target group. | 2024 | BMJ open | 14 | e066547 | RAYYAN-INCLUSION: {"Richard"=>"Excluded"} \| RAYYAN-EXCLUSION-REASONS: wrong outcome,wrong study design \| USER-NOTES: {"Richard"=>["Climate is the intervention"]} | 10.1136/bmjopen-2022-066547 | Child;Humans;Female;Pregnancy;Child, Preschool;Time Factors;Togo/epidemiology;Inc | PMC10828885 | https://pubmed.ncbi.nlm.nih.gov/38296296/ |
| rayyan-751570078 | Toure M and Shaffer JG and Sanogo D and Keita S and Keita M and Kane F and Traore B and Dabitao D and Kone A and Doumbia CO and Keating J and Yukich J and Hansson HH and | Seasonal Malaria Chemoprevention Therapy in Children Up To 9 Years of Age: Protocol for a Cluster-Randomized Trial Study. | 2024 | JMIR research protocols | 13 | e51660 | RAYYAN-INCLUSION: {"Richard"=>"Excluded"} \| RAYYAN-EXCLUSION-REASONS: background article,wrong publication type \| USER-NOTES: {"Richard"=>["Protocol article"]} | 10.2196/51660 | Only Child;Child;Seasons;Chemoprevention | PMC10845024 | https://pubmed.ncbi.nlm.nih.gov/38252481/ |
| rayyan-751570082 | Ciesielski TH and Sirũgo G and Iyengar SK and Williams SM | Characterizing the pathogenicity of genetic variants: the consequences of context. | 2024 | NPJ genomic medicine | 9 | 3 | RAYYAN-INCLUSION: {"Richard"=>"Excluded"} \| RAYYAN-EXCLUSION-REASONS: wrong outcome,wrong study design \| USER-NOTES: {"Richard"=>["Genomic surveillance study"]} | 10.1038/s41525-023-00386-5 | | PMC10776585 | https://pubmed.ncbi.nlm.nih.gov/38195641/ |
| rayyan-751570086 | Aidoo EK and Aboagye FT and Agginie GE and Botchway FA and Osei-Adjei G and Appiah M and Takyi RD and Sakyi SA and Amoah L and Arthur G and Lawson BW and Asmah RH and | Malaria elimination in Ghana: recommendations for reactive case detection strategy implementation in a low endemic area of Asutsuare, Ghana. | 2024 | Malaria journal | 23 | 5 | RAYYAN-INCLUSION: {"Richard"=>"Excluded"} \| RAYYAN-EXCLUSION-REASONS: wrong outcome,wrong study design \| USER-NOTES: {"Richard"=>["Risk factor study, no routine data used"]} | 10.1186/s12936-023-04792-z | Humans;Asymptomatic Infections/epidemiology;Cross-Sectional Studies;Diagnostic | PMC10759473 | https://pubmed.ncbi.nlm.nih.gov/38167067/ |
| rayyan-751570090 | Agaba BB and Smith D and Travis J and Pasay C and Nabatanzi M and Arinaitwe E and Ssewanyana I and Nabadda S and Cunningham J and Kamya MR and Cheng Q | Limited threat of Plasmodium falciparum pfhrp2 and pfhrp3 gene deletion to the utility of HRP2-based malaria RDTs in Northern Uganda. | 2024 | Malaria journal | 23 | 3 | RAYYAN-INCLUSION: {"Richard"=>"Excluded"} \| RAYYAN-EXCLUSION-REASONS: wrong outcome,wrong study design \| USER-NOTES: {"Richard"=>["Diagnostic study"]} | 10.1186/s12936-023-04830-w | Humans;Antigens, Protozoan/genetics;Cross-Sectional Studies;Diagnostic | PMC10759665 | https://pubmed.ncbi.nlm.nih.gov/38167003/ |
| rayyan-751570094 | Golumbeanu M and Edi CAV and Hetzel MW and Koepfli C and Nsanzabana C | Bridging the Gap from Molecular Surveillance to Programmatic Decisions for Malaria Control and Elimination. | 2023 | The American journal of | | | RAYYAN-INCLUSION: {"Richard"=>"Excluded"} \| RAYYAN-EXCLUSION-REASONS: wrong outcome,background article,wrong study design \| USER-NOTES: {"Richard"=>["Genomic surveillance"]} | 10.4269/ajtmh.22-0749 | Malaria | | https://pubmed.ncbi.nlm.nih.gov/38150733/ |
| rayyan-751570099 | Singh A and Singh MP and Ali NA and Poriya R and Rajvanshi H and Nisar S and Bhandari S and Sahu RS and Jayswar H and Mishra AK and Das A and Kaur H and Anvikar AR and | Assessment of Plasmodium falciparum drug resistance associated molecular markers in Mandla, Madhya Pradesh, India. | 2023 | Malaria journal | 22 | 375 | RAYYAN-INCLUSION: {"Richard"=>"Excluded"} \| RAYYAN-EXCLUSION-REASONS: wrong outcome,wrong study design \| USER-NOTES: {"Richard"=>["Genomic surveillance"]} | 10.1186/s12936-023-04817-7 | Humans;Plasmodium falciparum;*Antimalarials/pharmacology/therapeutic | PMC10712044 | https://pubmed.ncbi.nlm.nih.gov/38072967/ |
| rayyan-751570105 | Ashton RA and Worges M and Zeh Meka A and Yikpotey P and Domkam Kammogne I and Chanda-Kapata P and Vanderick C and Streat E and Yukich J | Can Outreach Training and Supportive Supervision Improve Competency in Malaria Service Delivery? An Evaluation in Cameroon, Ghana, Niger, and Zambia. | 2024 | The American journal of | 110 | 19-Oct | RAYYAN-INCLUSION: {"Richard"=>"Excluded"} \| RAYYAN-EXCLUSION-REASONS: wrong outcome,wrong study design \| USER-NOTES: {"Richard"=>["Diagnostic study"]} | 10.4269/ajtmh.23-0150 | Female;Humans;Pregnancy;Zambia/epidemiology;Cameroon/epidemiology;Ghana;Niger;* | PMC10919235 | https://pubmed.ncbi.nlm.nih.gov/38052082/ |
| rayyan-751570106 | Zaw AS and Win ESS and Yan SW and Thein KS and Verma V and McLean ARD and Kyaw TT and White NJ and Smithuis FM | Successful elimination of falciparum malaria following the introduction of community-based health workers in Eastern Myanmar: A retrospective analysis. | 2023 | PLoS medicine | 20 | e1004318 | RAYYAN-INCLUSION: {"Richard"=>"Included"} | 10.1371/journal.pmed.1004318 | Humans;Retrospective Studies;Community Health Workers;Myanmar/epidemiolo | PMC10721164 | https://pubmed.ncbi.nlm.nih.gov/38033155/ |
| rayyan-751570107 | Hoyt J and Okello G and Bange T and Kariuki S and Jalloh MF and Webster J and Hill J | RTS,S/AS01 malaria vaccine pilot implementation in western Kenya: a qualitative longitudinal study to understand immunisation barriers and optimise uptake. | 2023 | BMC public health | 23 | 2283 | RAYYAN-INCLUSION: {"Richard"=>"Excluded"} \| RAYYAN-EXCLUSION-REASONS: wrong outcome,wrong study design \| USER-NOTES: {"Richard"=>["QUal study"]} | 10.1186/s12889-023-17194-2 | Child;Humans;Infant;*Malaria Vaccines;*Malaria, Falciparum/prevention & | PMC10657022 | https://pubmed.ncbi.nlm.nih.gov/37980467/ |
| rayyan-751570108 | Thriemer K and Degaga TS and Christian M and Alam MS and Rajasekhar M and Ley B and Hossain MS and Kibria MG and Tego TT and Abate DT and Weston S and Mnjala H and | Primaquine radical cure in patients with Plasmodium falciparum malaria in areas co-endemic for P falciparum and Plasmodium vivax (PRIMA): a multicentre, open-label, superiority randomised controlled trial. | 2023 | Lancet (London, England) | 402 | 2101-2110 | RAYYAN-INCLUSION: {"Richard"=>"Excluded"} \| RAYYAN-EXCLUSION-REASONS: wrong outcome,wrong study design \| USER-NOTES: {"Richard"=>["No routine data was used"]} | 10.1016/S0140-6736(23)01553-2 | Humans;Primaquine/adverse effects;*Antimalarials/adverse effects;Plasmodium | PMC10714037 | https://pubmed.ncbi.nlm.nih.gov/37979594/ |
| rayyan-751570109 | Vesely BA and Cheng Q | Force Protection Risks in AFRICOM, INDOPACOM and SOUTHCOM Due to Rapid Diagnostic Test Failures for Falciparum Malaria, 2016-2022. | 2023 | MSMR | 30 | 11-Jul | RAYYAN-INCLUSION: {"Richard"=>"Excluded"} \| RAYYAN-EXCLUSION-REASONS: wrong outcome,wrong study design \| USER-NOTES: {"Richard"=>["Diagnostic study"]} | | Humans;Protozoan Proteins/genetics;Antigens, Protozoan/genetics;Rapid | | https://pubmed.ncbi.nlm.nih.gov/37963222/ |
| rayyan-751570110 | Fottsoh Fokam A and Rouamba T and Samadoulougou S and Ye Y and Kirakoya-Samadoulougou F | A Bayesian spatio-temporal framework to assess the effect of seasonal malaria chemoprevention on children under 5 years in Cameroon from 2016 to 2021 using routine data. | 2023 | Malaria journal | 22 | 347 | RAYYAN-INCLUSION: {"Richard"=>"Included"} | 10.1186/s12936-023-04677-1 | Humans;Child;Infant;Child, Preschool;*Antimalarials/therapeutic | PMC10640753 | https://pubmed.ncbi.nlm.nih.gov/37951942/ |
| rayyan-751570111 | Gondard M and Lane M and Barratt J and Talundzic E and Qvarnstrom Y | Simultaneous targeted amplicon deep sequencing and library preparation for a time and cost-effective universal parasite diagnostic sequencing approach. | 2023 | Parasitology research | 122 | 3243-3256 | RAYYAN-INCLUSION: {"Richard"=>"Excluded"} \| RAYYAN-EXCLUSION-REASONS: wrong outcome,wrong study design \| USER-NOTES: {"Richard"=>["Diagnostic study"]} | 10.1007/s00436-023-07991-4 | Animals;Humans;*Parasites/genetics;Cost-Benefit Analysis;*Plasmodium/genetic | PMC10772880 | https://pubmed.ncbi.nlm.nih.gov/37940706/ |
| rayyan-751570112 | Tarimo FS and Dillip A and Kosia EM and Lwetoijera DW | Community perception of the autodissemination of pyriproxyfen for controlling malaria vectors in south-eastern Tanzania. | 2023 | Malaria journal | 22 | 333 | RAYYAN-INCLUSION: {"Richard"=>"Excluded"} \| RAYYAN-EXCLUSION-REASONS: wrong outcome,wrong study design \| USER-NOTES: {"Richard"=>["QUal study"]} | 10.1186/s12936-023-04773-2 | Animals;Child;Humans;*Anopheles;Mosquito Vectors;Tanzania;*Malaria/pre | PMC10625276 | https://pubmed.ncbi.nlm.nih.gov/37924148/ |
| rayyan-751570115 | Segala FV and Ictho J and L'Episcopia M and Onapa E and De Vita E and Novara R and Olung N and Totaro V and Olal L and Patti G and Bingom C and Farina U and Papagni R and Agaro C | Impact of the COVID-19 pandemic on malaria in pregnancy indicators in Northern Uganda: a joinpoint regression analysis. | 2023 | Pathogens and global | | 9-Jan | RAYYAN-INCLUSION: {"Richard"=>"Included"} \| RAYYAN-LABELS: Excluded (C19) | 10.1080/20477724.2023.2273023 | Pregnancy;Uganda;Regression Analysis;Malaria | | https://pubmed.ncbi.nlm.nih.gov/37872763/ |
| rayyan-751570116 | Kabera M and Mangala JN and Soebiyanto R and Mukarugwiro B and Munguti K and Mbituyumuremyi A and Lucchi NW and Hakizimana E | Impact of Pyrethroid Plus Piperonyl Butoxide Synergist-Treated Nets on Malaria Incidence 24 Months after a National Distribution Campaign in Rwanda. | 2023 | The American journal of | 109 | 1356-1362 | RAYYAN-INCLUSION: {"Richard"=>"Included"} | 10.4269/ajtmh.23-0317 | Humans;*Pyrethrins/pharmacology;Piperonyl Butoxide/pharmacology;Incide | PMC10793038 | https://pubmed.ncbi.nlm.nih.gov/37871590/ |
| rayyan-751570117 | Marcombe S and Maithaviphet S and Reyburn R and Kunlaya K and Silavong K and Hongvanthong B and Vanisaveth V and Sengsavath V and Banouvong V and Chindavongsa K and | Bionomics of malaria vectors in Lao PDR, 2018-2020: entomological surveillance as a key tool for malaria elimination. | 2023 | Malaria journal | 22 | 319 | RAYYAN-INCLUSION: {"Richard"=>"Excluded"} \| RAYYAN-EXCLUSION-REASONS: wrong outcome,wrong study design \| USER-NOTES: {"Richard"=>["Ento study"]} | 10.1186/s12936-023-04754-5 | Female;Humans;Animals;Cattle;*Malaria/epidemiology/prevention & | PMC10589990 | https://pubmed.ncbi.nlm.nih.gov/37865735/ |
| rayyan-751570118 | Altare C and Matadi Basadia L and Kostandova N and Nsio Mbeta J and Bruneau S and Antoine C and Petry M | The implementation of infection prevention and control measures and health care utilisation in ACF-supported health facilities during the COVID-19 pandemic in Kinshasa, Democratic Republic of the Congo, 2020. | 2023 | Global health action | 16 | 2258711 | RAYYAN-INCLUSION: {"Richard"=>"Excluded"} \| RAYYAN-EXCLUSION-REASONS: wrong outcome,wrong study design,wrong infection / disease | 10.1080/16549716.2023.2258711 | Pregnancy;Female;Humans;*COVID-19/epidemiology/prevention & | PMC10583608 | https://pubmed.ncbi.nlm.nih.gov/37846089/ |
| rayyan-751570119 | Kihwele F and Gavana T and Makungu C and Msuya HM and Mlacha YP and Govella NJ and Chaki PP and Sunguya BF | Exploring activities and behaviours potentially increases school-age children's vulnerability to malaria infections in south-eastern Tanzania. | 2023 | Malaria journal | 22 | 293 | RAYYAN-INCLUSION: {"Richard"=>"Excluded"} \| RAYYAN-EXCLUSION-REASONS: wrong outcome,wrong study design \| USER-NOTES: {"Richard"=>["Qual study"]} | 10.1186/s12936-023-04703-2 | Male;Female;Humans;Child;Adolescent;Tanzania/epidemiology;Cross-Sectional | PMC10548596 | https://pubmed.ncbi.nlm.nih.gov/37789435/ |
| rayyan-751570120 | Moturi AK and Jalang'o R and Cherono A and Muchiri SK and Snow RW and Okiro EA | Malaria vaccine coverage estimation using age-eligible populations and service user denominators in Kenya. | 2023 | Malaria journal | 22 | 287 | RAYYAN-INCLUSION: {"Richard"=>"Excluded"} \| RAYYAN-EXCLUSION-REASONS: wrong outcome,wrong study design \| USER-NOTES: {"Richard"=>["No epi indicator; focus on coverage"]} | 10.1186/s12936-023-04721-0 | Child;Infant;Humans;*Malaria Vaccines;Kenya;Biological Transport;*Health Information | PMC10523632 | https://pubmed.ncbi.nlm.nih.gov/37759277/ |
| rayyan-751570121 | Nakijoba R and Nakayiwa Kawuma A and Ojara FW and Tabwenda JC and Kyeyune J and Turyahabwe C and Asiimwe SP and Magoola J and Banda CG and Castelnuovo B and | #NAME? | 2023 | Wellcome open research | 8 | 12 | RAYYAN-INCLUSION: {"Richard"=>"Excluded"} \| RAYYAN-EXCLUSION-REASONS: wrong outcome,wrong study design,wrong publication type \| USER-NOTES: {"Richard"=>["Protocol"]} | 10.12688/wellcomeopenres.18512.1 | Antimalarials;Infant;Breast Feeding;Malaria | PMC10514676 | https://pubmed.ncbi.nlm.nih.gov/37744730/ |
| rayyan-751570122 | Zeno EE and Obala AA and Pence B and Freedman E and Mangeni JN and Lin JT and Abel L and Edwards JK and Gower EW and Taylor SM | Risk of Malaria Following Untreated Subpatent Plasmodium falciparum Infections: Results Over 4 Years From a Cohort in a High-Transmission Area in Western Kenya. | 2024 | The Journal of infectious | 229 | 969-978 | RAYYAN-INCLUSION: {"Richard"=>"Excluded"} \| RAYYAN-EXCLUSION-REASONS: wrong outcome,wrong study design \| USER-NOTES: {"Richard"=>["Diagnostic study"]} | 10.1093/infdis/jiad398 | Humans;Plasmodium falciparum;Kenya/epidemiology;*Malaria, | PMC11011184 | https://pubmed.ncbi.nlm.nih.gov/37713614/ |
| rayyan-751570123 | Osoro CB and Dellicour S and Ochodo E and Young T and Ter Kuile FO and Gutman JR and Hill J | Perceptions and drivers of healthcare provider and drug dispenser practices for the treatment of malaria in pregnancy in the context of multiple first-line therapies in western Kenya: a qualitative study. | 2023 | Malaria journal | 22 | 274 | RAYYAN-INCLUSION: {"Richard"=>"Excluded"} \| RAYYAN-EXCLUSION-REASONS: wrong outcome,wrong study design \| USER-NOTES: {"Richard"=>["Qual study"]} | 10.1186/s12936-023-04698-w | Pregnancy;Humans;Female;*Antimalarials/therapeutic use;Kenya;Pharmaceutical | PMC10503024 | https://pubmed.ncbi.nlm.nih.gov/37710229/ |
| rayyan-751570124 | Kojom Foko LP and Jakhan J and Narang G and Singh V | Global polymorphism of Plasmodium falciparum histidine rich proteins 2/3 and impact on malaria rapid diagnostic test detection: a systematic review and meta-analysis. | 2023 | Expert review of molecular | 23 | 925-943 | RAYYAN-INCLUSION: {"Richard"=>"Excluded"} \| RAYYAN-EXCLUSION-REASONS: wrong outcome,wrong study design,wrong publication type \| USER-NOTES: {"Richard"=>["Review"]} | 10.1080/14737159.2023.2255136 | Humans;Plasmodium falciparum/genetics;Histidine;*Malaria, | | https://pubmed.ncbi.nlm.nih.gov/37698448/ |
| rayyan-751570125 | Burnett SM and Davis KM and Assefa G and Gogue C and Hinneh LD and Littrell M and Mwesigwa J and Okoko OO and Rabeherisoa S and Sillah-Kanu M and Sheahan W and Slater | Process and Methodological Considerations for Observational Analyses of Vector Control Interventions in Sub-Saharan Africa Using Routine Malaria Data. | 2023 | The American journal of | | | RAYYAN-INCLUSION: {"Richard"=>"Excluded"} \| RAYYAN-EXCLUSION-REASONS: wrong study design,wrong publication type \| USER-NOTES: {"Richard"=>["Review"]} | 10.4269/ajtmh.22-0757 | Africa South of the Sahara;Malaria;Africa | | https://pubmed.ncbi.nlm.nih.gov/37604476/ |
| rayyan-751570126 | NuÃ±ez L and Skjefte M and Asamoah OE and Owusu P and Malm KL and Miller JE | Measuring quality of facility-based ITN distribution in Ghana. | 2023 | Malaria journal | 22 | 222 | RAYYAN-INCLUSION: {"Richard"=>"Excluded"} \| RAYYAN-EXCLUSION-REASONS: wrong outcome,wrong study design \| USER-NOTES: {"Richard"=>["No epi indicator"]} | 10.1186/s12936-023-04626-y | Child;Humans;Female;Pregnancy;Ghana;*Insecticide-Treated Bednets;*Malaria/prevention | PMC10394948 | https://pubmed.ncbi.nlm.nih.gov/37533064/ |
| rayyan-751570127 | Mchenga M and Burger R and von Fintel D | Can women's reports in client exit interviews be used to measure and track progress of antenatal care services quality? Evidence from a facility assessment census in Malawi. | 2023 | PloS one | 18 | e0274650 | RAYYAN-INCLUSION: {"Richard"=>"Excluded"} \| RAYYAN-EXCLUSION-REASONS: wrong outcome,wrong study design,wrong infection / disease | 10.1371/journal.pone.0274650 | Pregnancy;Female;Humans;*Prenatal Care;Reproducibility of Results;Malawi;Censuses;Surv | PMC10389737 | https://pubmed.ncbi.nlm.nih.gov/37523376/ |
| rayyan-751570128 | Bisaccia C and De Santo LS and De Santo NG | Malaria as a Papal Disease. | 2023 | Experimental and clinical | 21 | 28-32 | RAYYAN-INCLUSION: {"Richard"=>"Excluded"} \| RAYYAN-EXCLUSION-REASONS: wrong outcome,wrong study design | 10.6002/ect.IAHNCongress.07 | Humans;Italy;Rome;*Malaria/diagnosis/epidemiology/history;Malaria | | https://pubmed.ncbi.nlm.nih.gov/37496339/ |
| rayyan-751570179 | Diallo AO and Banek K and Kashamuka MM and Bala JAM and Nkalani M and Kihuma G and Nseka TM and Atibu JL and Mahilu GE and McCormick L and White SJ and Sendor R and | Impact of malaria diagnostic choice on monitoring of Plasmodium falciparum prevalence estimates in the Democratic Republic of the Congo and relevance to control programs in high-burden countries. | 2023 | PLOS global public | 3 | e0001375 | RAYYAN-INCLUSION: {"Richard"=>"Excluded"} \| RAYYAN-EXCLUSION-REASONS: wrong outcome,wrong study design \| USER-NOTES: {"Richard"=>["Mention of routine data", "Dx study"]} | 10.1371/journal.pgph.0001375 | Prevalence;Plasmodium falciparum;Malaria;Plasmodium | PMC10370698 | https://pubmed.ncbi.nlm.nih.gov/37494361/ |
| rayyan-751570180 | Iga J and Ochaya S and Echodu R and Opiyo EA and Musiime AK and Nakamaanya A and Malinga GM | Sibling Species Composition and Susceptibility Status of Anopheles gambiae s.l. to Insecticides Used for Indoor Residual Spraying in Eastern Uganda. | 2023 | Journal of parasitology | 2023 | 2225233 | RAYYAN-INCLUSION: {"Richard"=>"Excluded"} \| RAYYAN-EXCLUSION-REASONS: wrong outcome,wrong study design \| USER-NOTES: {"Richard"=>["Ento study"]} | 10.1155/2023/2225233 | Uganda;Insecticides | PMC10352534 | https://pubmed.ncbi.nlm.nih.gov/37469527/ |
| rayyan-751570181 | Hilton ER and Rabeherisoa S and Ramandimbiarijaona H and Rajaratnam J and Belemvire A and Kapesa L and Zohdy S and Dentinger C and Gandaho T and Jacob D and Burnett S and | Using routine health data to evaluate the impact of indoor residual spraying on malaria transmission in Madagascar. | 2023 | BMJ global health | 8 | | RAYYAN-INCLUSION: {"Richard"=>"Included"} | 10.1136/bmjgh-2022-010818 | Humans;*Insecticides;Madagascar/epidemiology;Mosquito Control/methods;*Malaria/epi | PMC10357738 | https://pubmed.ncbi.nlm.nih.gov/37463785/ |

| | | | | | | | | | | | |
|---|---|---|---|---|---|---|---|---|---|---|---|
| rayyan-751570182 | Wamae K and Ndwiga L and Kharabora O and Kimenyi K and Osoti V and de Laurent Z and Wambua J and Musyoki J and Ngetsa C and Kalume P and Mwambingu G and Hamaluba M | Targeted amplicon deep sequencing of ama1 and mdr1 to track within-host P. falciparum diversity throughout treatment in a clinical drug trial. | 2022 | Wellcome open research | 7 | 95 | RAYYAN-INCLUSION: {"Richard"=>"Excluded"} \| RAYYAN-EXCLUSION-REASONS: wrong outcome,wrong study design | 10.12688/wellcomeopenres.17736.2 | | PMC10349275 | https://pubmed.ncbi.nlm.nih.gov/37456906/ |
| rayyan-751570184 | Yan H and Wei S and Sui Y and Lu S and Zhang W and Feng X and Liu Y and Zhang T and Ruan W and Xia J and Lin W and Ley B and Auburn S and Li S and Li J and Wang D | Analysis of the relapse of imported Plasmodium vivax and Plasmodium ovale in five provinces of China. | 2023 | Malaria journal | 22 | 209 | RAYYAN-INCLUSION: {"Richard"=>"Excluded"} \| RAYYAN-EXCLUSION-REASONS: wrong outcome,wrong study design | 10.1186/s12936-023-04642-y | Humans;Plasmodium vivax;*Plasmodium ovale;Case-Control Studies;Retrospective | PMC10347707 | https://pubmed.ncbi.nlm.nih.gov/37443070/ |
| rayyan-751570185 | Abong'o B and Stanton MC and Donnelly MJ and Ochomo E and Ter Kuile FO and Samuels AM and Kariuki S and Musula G and Oxborough R and Munga S and Torr SJ and Gimnig JE | Evaluation of community-based vector surveillance system for routine entomological monitoring under low malaria vector densities and high bed net coverage in western Kenya. | 2023 | Malaria journal | 22 | 203 | RAYYAN-INCLUSION: {"Richard"=>"Excluded"} \| RAYYAN-EXCLUSION-REASONS: wrong outcome,wrong study design | 10.1186/s12936-023-04629-9 | Animals;Humans;*Malaria;*Anopheles;Kenya/epidemiology;Mosquito Vectors;Feeding | PMC10318638 | https://pubmed.ncbi.nlm.nih.gov/37400805/ |
| rayyan-751570186 | Abebe A and Menard D and Dugassa S and Assefa A and Juliano JJ and Lo E and Golassa L | Significant number of Plasmodium vivax mono-infections by PCR misidentified as mixed infections (P. vivax/P. falciparum) by microscopy and rapid diagnostic tests: malaria diagnostic challenges in Ethiopia. | 2023 | Malaria journal | 22 | 201 | RAYYAN-INCLUSION: {"Richard"=>"Excluded"} \| RAYYAN-EXCLUSION-REASONS: wrong outcome,wrong study design \| USER-NOTES: {"Richard"=>["Diagnostic study"]} | 10.1186/s12936-023-04635-x | Humans;*Malaria, Vivax/diagnosis/epidemiology;Plasmodium | PMC10314452 | https://pubmed.ncbi.nlm.nih.gov/37393257/ |
| rayyan-751570187 | van Bergen KJM and Stuitje AR and Akkers RC and Vermeer HJ and Castel R and Mank TG | Performance of a novel melting curve-based qPCR assay for malaria parasites in routine clinical practice in non-endemic setting. | 2023 | Malaria journal | 22 | 191 | RAYYAN-INCLUSION: {"Richard"=>"Excluded"} \| RAYYAN-EXCLUSION-REASONS: wrong outcome,wrong study design \| USER-NOTES: {"Richard"=>["Diagnostic study"]} | 10.1186/s12936-023-04617-z | Humans;*Malaria, Falciparum/diagnosis/parasitology;*Plasmodium;*Malaria/dia | PMC10286418 | https://pubmed.ncbi.nlm.nih.gov/37349789/ |
| rayyan-751570188 | Sorano S and Gore-Langton G and Opondo C and Smith C and Matsui M and Chaponda EB and Chandramohan D and Chico RM | Coinfections of malaria and sexually transmitted and reproductive tract infections in pregnancy in sub-Saharan Africa: a systematic review and individual participant data meta-analysis protocol. | 2023 | BMJ open | 13 | e074896 | RAYYAN-INCLUSION: {"Richard"=>"Excluded"} \| RAYYAN-EXCLUSION-REASONS: wrong study design,wrong publication type \| USER-NOTES: {"Richard"=>["Review"]} | 10.1136/bmjopen-2023-074896 | Humans;Female;Pregnancy;*Reproductive Tract Infections/epidemiology;*Coin | PMC10314643 | https://pubmed.ncbi.nlm.nih.gov/37339835/ |
| rayyan-751570189 | Jones HS and Hensen B and Musemburi S and Chinyanganya L and Takaruza A and Chabata ST and Matambanadzo P and Platt L and Rice B and Cowan FM and Hargreaves JR | Temporal trends in, and risk factors for, HIV seroconversion among female sex workers accessing Zimbabwe's national sex worker programme, 2009-19: a retrospective cohort analysis of routinely collected HIV testing data. | 2023 | The lancet. HIV | 10 | e442-e452 | RAYYAN-INCLUSION: {"Richard"=>"Excluded"} \| RAYYAN-EXCLUSION-REASONS: wrong infection / disease | 10.1016/S2352-3018(23)00110-8 | Female;Humans;*HIV Infections/diagnosis/epidemiology/prevention & control;*HIV | | https://pubmed.ncbi.nlm.nih.gov/37329897/ |
| rayyan-751570191 | Angeles G and Silverstein H and Worges M and Hotchkiss DR and Wisniewski JM and Lusamba Dikassa PS and Weiss W and Ahsan KZ | Area-specific covid-19 effects on health services utilization in the Democratic Republic of the Congo using routine health information system data. | 2023 | BMC health services | 23 | 575 | RAYYAN-INCLUSION: {"Richard"=>"Maybe"} \| RAYYAN-LABELS: Excluded (C19) \| USER-NOTES: {"Richard"=>["COVID as intervention"]} | 10.1186/s12913-023-09547-9 | Child;Humans;Child, Preschool;Democratic Republic of the | PMC10239053 | https://pubmed.ncbi.nlm.nih.gov/37270545/ |
| rayyan-751570192 | Altare C and Kostandova N and Gankpe GF and Nalimo P and Almoustapha Abaradine AA and Bruneau S and Antoine C and Spiegel PB | The first year of the COVID-19 pandemic in humanitarian settings: epidemiology, health service utilization, and health care seeking behavior in Bangui and surrounding areas, Central African Republic. | 2023 | Conflict and health | 17 | 24 | RAYYAN-INCLUSION: {"Richard"=>"Excluded"} \| RAYYAN-EXCLUSION-REASONS: wrong outcome,wrong study design | 10.1186/s13031-023-00523-y | Patient Acceptance of Health Care | PMC10199293 | https://pubmed.ncbi.nlm.nih.gov/37210535/ |
| rayyan-751570194 | Augusto O and Roberton T and Fernandes Q and Chicumbe S and ManhiÃ§a I and Tembe S and Wagenaar BH and Anselmi L and Wakefield J and Sherr K | Early effects of COVID-19 on maternal and child health service disruption in Mozambique. | 2023 | Frontiers in public health | 11 | 1075691 | RAYYAN-INCLUSION: {"Richard"=>"Maybe"} \| RAYYAN-LABELS: Excluded (C19) \| USER-NOTES: {"Richard"=>["COVID19 intervention"]} | 10.3389/fpubh.2023.1075691 | Infant, Newborn;Child;Female;Humans;*COVID- | PMC10149948 | https://pubmed.ncbi.nlm.nih.gov/37139385/ |
| rayyan-751570195 | Ozodiegwu ID and Ambrose M and Galatas B and Runge M and Nandi A and Okuneye K and Dhanoa NP and Maikore I and Uhomoibhi P and Bever C and Noor A and Gerardin J | Application of mathematical modelling to inform national malaria intervention planning in Nigeria. | 2023 | Malaria journal | 22 | 137 | RAYYAN-INCLUSION: {"Richard"=>"Excluded"} \| RAYYAN-EXCLUSION-REASONS: wrong outcome,wrong study design \| USER-NOTES: {"Richard"=>["Modeling stratification"]} | 10.1186/s12936-023-04563-w | Child;Infant;Humans;Child, Preschool;Nigeria/epidemiology;*Malaria/epidemiology/prev | PMC10130303 | https://pubmed.ncbi.nlm.nih.gov/37101146/ |
| rayyan-751570196 | Azizi H and Davtalab Esmaeili E and Abbasi F | Availability of malaria diagnostic tests, anti-malarial drugs, and the correctness of treatment: a systematic review and meta-analysis. | 2023 | Malaria journal | 22 | 127 | RAYYAN-INCLUSION: {"Richard"=>"Excluded"} \| RAYYAN-EXCLUSION-REASONS: wrong study design,wrong publication type \| USER-NOTES: {"Richard"=>["REview"]} | 10.1186/s12936-023-04555-w | Humans;*Antimalarials/therapeutic use;Diagnostic Tests, Routine;*Malaria/diagnosis/dr | PMC10111310 | https://pubmed.ncbi.nlm.nih.gov/37072759/ |
| rayyan-751570197 | Srivastava B and Sharma S and Ahmed N and Kumari P and Gahtori R and Sinha S and Kumar S and Sanalkumar M and Mahale P and Swarnkar D and Anvikar AR | Quality assurance of malaria rapid diagnostic tests: An aid in malaria elimination. | 2023 | The Indian journal of | 157 | 23-29 | RAYYAN-INCLUSION: {"Richard"=>"Excluded"} \| RAYYAN-EXCLUSION-REASONS: wrong outcome,wrong study design \| USER-NOTES: {"Richard"=>["Diag study"]} | 10.4103/ijmr.ijmr_2262_21 | Humans;*Diagnostic Tests, Routine/methods;*Malaria/diagnosis;Rapid Diagnostic | PMC10284356 | https://pubmed.ncbi.nlm.nih.gov/37040223/ |
| rayyan-751570199 | Adjei MR and Amponsa-Achiano K and Okine R and Tweneboah PO and Sally ET and Dadzie JF and Osei-Sarpong F and Adjabeng MJ and Bawa JT and Bonsu G and Antwi-Agyei | Post introduction evaluation of the malaria vaccine implementation programme in Ghana, 2021. | 2023 | BMC public health | 23 | 586 | RAYYAN-INCLUSION: {"Richard"=>"Excluded"} \| RAYYAN-EXCLUSION-REASONS: wrong outcome,wrong study design \| USER-NOTES: {"Richard"=>["Not epi indicator"]} | 10.1186/s12889-023-15481-6 | Humans;*Malaria Vaccines;Ghana/epidemiology;*Malaria/prevention & | PMC10052308 | https://pubmed.ncbi.nlm.nih.gov/36991394/ |
| rayyan-751570201 | Bridges DJ and Miller JM and Chalwe V and Moonga H and Hamainza B and Steketee RW and Mambwe B and Mulube C and Wu L and Tetteh KKA and Drakeley C and Chishimba S and | Reactive focal drug administration associated with decreased malaria transmission in an elimination setting: Serological evidence from the cluster-randomized CoRE study. | 2022 | PLOS global public | 2 | e0001295 | RAYYAN-INCLUSION: {"Richard"=>"Included"} | 10.1371/journal.pgph.0001295 | Malaria | PMC10021141 | https://pubmed.ncbi.nlm.nih.gov/36962857/ |
| rayyan-751570208 | Ashton RA and Hamainza B and Lungu C and Rutagwera MI and Porter T and Bennett A and Hainsworth M and Burnett S and Silumbe K and Slater H and Eisele TP and Miller JM | Effectiveness of community case management of malaria on severe malaria and inpatient malaria deaths in Zambia: a dose-response study using routine health information system data. | 2023 | Malaria journal | 22 | 96 | RAYYAN-INCLUSION: {"Richard"=>"Included"} | 10.1186/s12936-023-04525-2 | Child;Humans;*Antimalarials/therapeutic use;Zambia/epidemiology;Cas | PMC10022244 | https://pubmed.ncbi.nlm.nih.gov/36927440/ |
| rayyan-751570212 | Semakula M and Niragire F and Faes C | Spatio-Temporal Bayesian Models for Malaria Risk Using Survey and Health Facility Routine Data in Rwanda. | 2023 | International journal of | 20 | | RAYYAN-INCLUSION: {"Richard"=>"Excluded"} \| RAYYAN-EXCLUSION-REASONS: wrong outcome,wrong study design \| USER-NOTES: {"Richard"=>["No intervnetion "]} | 10.3390/ijerph20054283 | Child;Humans;Child, Preschool;Rwanda;Bayes Theorem;*Malaria/epidemiolo | PMC10001932 | https://pubmed.ncbi.nlm.nih.gov/36901291/ |
| rayyan-751570220 | Uwimana A and Sethi R and Murindahabi M and Ntirandeka C and Piercefield E and Umulisa N and Abram A and Eckert E and Munguti K and Sullivan D and Uyizeye D and | Effectiveness of Intermittent Screening and Treatment of Malaria in Pregnancy on Maternal and Birth Outcomes in Selected Districts in Rwanda: A Cluster Randomized Controlled Trial. | 2023 | Clinical infectious diseases : | 77 | 127-134 | RAYYAN-INCLUSION: {"Richard"=>"Excluded"} \| RAYYAN-EXCLUSION-REASONS: wrong study design \| USER-NOTES: {"Richard"=>["Unclear if routine data was used"]} | 10.1093/cid/ciad128 | Infant, Newborn;Female;Pregnancy;Humans;*Antimalarials/therape | PMC10330390 | https://pubmed.ncbi.nlm.nih.gov/36896967/ |
| rayyan-751570231 | Roca A and Camara B and Bognini JD and Nakakana UN and SomÃ© AM and Beloum N and Rouamba T and Sillah F and Danso M and Jones JC and Graves S and Jagne I and Getanda P | Effect of Intrapartum Azithromycin vs Placebo on Neonatal Sepsis and Death: A Randomized Clinical Trial. | 2023 | JAMA | 329 | 716-724 | RAYYAN-INCLUSION: {"Richard"=>"Excluded"} \| RAYYAN-EXCLUSION-REASONS: wrong infection / disease | 10.1001/jama.2022.24388 | Adult;Female;Humans;Infant, Newborn;Pregnancy;*Anti-Bacterial | PMC9993186 | https://pubmed.ncbi.nlm.nih.gov/36881034/ |
| rayyan-751570237 | Ding XC and Incardona S and Serra-Casas E and Charnaud SC and Slater HC and Domingo GJ and Adams ER and Ter Kuile FO and Samuels AM and Kariuki S and Dittrich S | Malaria in pregnancy (MiP) studies assessing the clinical performance of highly sensitive rapid diagnostic tests (HS-RDT) for Plasmodium falciparum detection. | 2023 | Malaria journal | 22 | 60 | RAYYAN-INCLUSION: {"Richard"=>"Excluded"} \| RAYYAN-EXCLUSION-REASONS: wrong outcome,wrong study design \| USER-NOTES: {"Richard"=>["Diag study"]} | 10.1186/s12936-023-04445-1 | Adult;Pregnancy;Child;Humans;Female;Plasmodium falciparum;Rapid Diagnostic | PMC9942317 | https://pubmed.ncbi.nlm.nih.gov/36803858/ |
| rayyan-751570246 | Sabin S and Jones S and Patel D and Subramaniam G and Kelley J and Aidoo M and Talundzic E | Portable and cost-effective genetic detection and characterization of Plasmodium falciparum hrp2 using the MinION sequencer. | 2023 | Scientific reports | 13 | 2893 | RAYYAN-INCLUSION: {"Richard"=>"Excluded"} \| RAYYAN-EXCLUSION-REASONS: wrong outcome,wrong study design \| USER-NOTES: {"Richard"=>["Genomic surveillance"]} | 10.1038/s41598-022-26935-z | Humans;*Plasmodium falciparum/genetics;Antigens, Protozoan/genetics;Protozoan | PMC9938884 | https://pubmed.ncbi.nlm.nih.gov/36801925/ |
| rayyan-751570254 | Okereke E and Smith H and Oguoma C and Oresanya O and Maxwell K and Anikwe C and Osuji LC and Ogazi O and Musa J and Rajab A and Shekarau E and Okoh F and ViganÃ² E and | Optimizing the role of 'lead mothers' in seasonal malaria chemoprevention (SMC) campaigns: formative research in Kano State, northern Nigeria. | 2023 | Malaria journal | 22 | 13 | RAYYAN-INCLUSION: {"Richard"=>"Excluded"} \| RAYYAN-EXCLUSION-REASONS: wrong outcome,wrong study design \| USER-NOTES: {"Richard"=>["Qual study"]} | 10.1186/s12936-023-04447-z | Child;Female;Humans;Infant;Child, Preschool;Mothers;Nigeria;Sea | PMC9835293 | https://pubmed.ncbi.nlm.nih.gov/36635665/ |
| rayyan-751570258 | Kurth F and Tober-Lau P and Lingscheid T and Bardtke L and Kim J and Angheben A and Gobbi FG and Mbavu L and Stegemann MS and Heim KM and PfÃ¤fflin F and Menner N | Post-treatment haemolysis is common following oral artemisinin combination therapy of uncomplicated malaria in travellers. | 2023 | Journal of travel medicine | 30 | | RAYYAN-INCLUSION: {"Richard"=>"Excluded"} \| RAYYAN-EXCLUSION-REASONS: wrong outcome,wrong study design \| USER-NOTES: {"Richard"=>["Diag study"]} | 10.1093/jtm/taad001 | Humans;Child;*Antimalarials/adverse effects;Hemolysis;*Artemisini | PMC10198431 | https://pubmed.ncbi.nlm.nih.gov/36611010/ |
| rayyan-751570263 | Bakai TA and Thomas A and Iwaz J and Atcha-Oubou T and Tchadjobo T and Khanafer N and Rabilloud M and Voirin N | Effectiveness of seasonal malaria chemoprevention in three regions of Togo: a population-based longitudinal study from 2013 to 2020. | 2022 | Malaria journal | 21 | 400 | RAYYAN-INCLUSION: {"Richard"=>"Maybe"} \| RAYYAN-LABELS: Excluded (after full text) \| USER-NOTES: {"Richard"=>["Routine HMIS data?"]} | 10.1186/s12936-022-04434-w | Child;Humans;Infant;*Antimalarials/therapeutic use;Seasons;Togo/epidemiolo | PMC9804945 | https://pubmed.ncbi.nlm.nih.gov/36587191/ |
| rayyan-751570264 | Longchamps C and Galindo MS and Lambert Y and Sanna A and Mutricy L and Garancher L and Adenis A and Nacher M and Suarez-Mutis M and Cairo H and Hiwat H and Vreden S | Impact of Malakit intervention on perceptions, knowledge, attitudes, and practices related to malaria among workers in clandestine gold mines in French Guiana: results of multicentric cross-sectional surveys over time. | 2022 | Malaria journal | 21 | 397 | RAYYAN-INCLUSION: {"Richard"=>"Excluded"} \| RAYYAN-EXCLUSION-REASONS: wrong outcome,wrong study design \| USER-NOTES: {"Richard"=>["No epi indicator"]} | 10.1186/s12936-022-04391-4 | Humans;Cross-Sectional Studies;French Guiana/epidemiology;*Gold;H | PMC9795716 | https://pubmed.ncbi.nlm.nih.gov/36577968/ |
| rayyan-751570265 | Lambiris MJ and Venga GN and Ssempala R and Balogun V and Galactionova K and Musiitwa M and Kagwire F and Olosunde O and Emedo E and Luketa S and Sangare M and Buj V and | Health system readiness and the implementation of rectal artesunate for severe malaria in sub-Saharan Africa: an analysis of real-world costs and constraints. | 2023 | The Lancet. Global | 11 | e256-e264 | RAYYAN-INCLUSION: {"Richard"=>"Excluded"} \| RAYYAN-EXCLUSION-REASONS: wrong outcome,wrong study design \| USER-NOTES: {"Richard"=>["Costing study\n"]} | 10.1016/S2214-109X(22)00507-1 | Humans;Artesunate/therapeutic use;*Antimalarials/therapeutic | | https://pubmed.ncbi.nlm.nih.gov/36565705/ |
| rayyan-751570266 | Bazant E and McPhillips-Tangum C and Shrestha SD and G S P and Khera A and Nic Lochlainn L and Habtamu E and Patel VI and Muhire G and Saarlas KN | Promising practices for the collaborative planning of integrated health campaigns from a synthesis of case studies. | 2022 | BMJ global health | 7 | | RAYYAN-INCLUSION: {"Richard"=>"Excluded"} \| RAYYAN-EXCLUSION-REASONS: wrong study design,wrong publication type | 10.1136/bmjgh-2022-010321 | Humans;Global Health;*Health Promotion/organization & administration;Organizational | PMC9756207 | https://pubmed.ncbi.nlm.nih.gov/36517112/ |
| rayyan-751570267 | Huber JH and Elliott M and Koepfli C and Perkins TA | The Impact of Emerging Plasmodium knowlesi on Accurate Diagnosis by Light Microscopy: A Systematic Review and Modeling Analysis. | 2023 | The American journal of | 108 | 61-68 | RAYYAN-INCLUSION: {"Richard"=>"Excluded"} \| RAYYAN-EXCLUSION-REASONS: wrong outcome,wrong study design,wrong publication type \| USER-NOTES: {"Richard"=>["Review"]} | 10.4269/ajtmh.21-1155 | Humans;Bayes Theorem;*Malaria/diagnosis/epidemiology/parasitology;Mala | PMC9833074 | https://pubmed.ncbi.nlm.nih.gov/36509046/ |
| rayyan-751570268 | Martins A and Herzog SA and Mugenyi L and Faes C and Hens N and Abrams S | Modelling longitudinal binary outcomes with outcome dependent observation times: an application to a malaria cohort study. | 2022 | Malaria journal | 21 | 380 | RAYYAN-INCLUSION: {"Richard"=>"Excluded"} \| RAYYAN-EXCLUSION-REASONS: wrong outcome,wrong study design | 10.1186/s12936-022-04386-1 | Child;Humans;Cohort Studies;*Malaria/prevention & control;Parasitemia/epidemiol | PMC9741489 | https://pubmed.ncbi.nlm.nih.gov/36496382/ |
| rayyan-751570269 | Lendongo-Wombo JB and Oyegue-Liabagui SL and Biteghe-Bi-Essone JC and Ngoungou EB and Lekana-Douki JB | EpidÃ©miology of malaria from 2019 to 2021 in the southeastern city of Franceville, Gabon. | 2022 | BMC public health | 22 | 2313 | RAYYAN-INCLUSION: {"Richard"=>"Excluded"} \| RAYYAN-EXCLUSION-REASONS: wrong outcome,wrong study design | 10.1186/s12889-022-14765-7 | Humans;Retrospective Studies;Pandemics;*COVID-19/epidemiology;*Malaria/epi | PMC9739344 | https://pubmed.ncbi.nlm.nih.gov/36496354/ |
| rayyan-751570270 | Thang ND and Rovira-Vallbona E and Binh NTH and Dung DV and Ngoc NTH and Long TK and Duong TT and Martin NJ and Edgel KA | Surveillance of pfhrp2 and pfhrp3 gene deletions among symptomatic Plasmodium falciparum malaria patients in Central Vietnam. | 2022 | Malaria journal | 21 | 371 | RAYYAN-INCLUSION: {"Richard"=>"Excluded"} \| RAYYAN-EXCLUSION-REASONS: wrong outcome,wrong study design \| USER-NOTES: {"Richard"=>["Genomic study"]} | 10.1186/s12936-022-04399-w | Humans;*Plasmodium falciparum/genetics/metabolism;Protozoan | PMC9724378 | https://pubmed.ncbi.nlm.nih.gov/36471315/ |
| rayyan-751570271 | Rogier E and Bakari C and Mandara CI and Chiduo MG and Plucinski M and Nace D and Battle N and Chacky F and Rumisha SF and Molteni F and Mandike R and Mkude S and | Performance of antigen detection for HRP2-based malaria rapid diagnostic tests in community surveys: Tanzania, July-November 2017. | 2022 | Malaria journal | 21 | 361 | RAYYAN-INCLUSION: {"Richard"=>"Excluded"} \| RAYYAN-EXCLUSION-REASONS: wrong outcome,wrong study design \| USER-NOTES: {"Richard"=>["Diag study"]} | 10.1186/s12936-022-04383-4 | Humans;*Histidine;Diagnostic Tests, Routine;Cross-Sectional Studies;Tanzania/epidemiolog | PMC9714097 | https://pubmed.ncbi.nlm.nih.gov/36457087/ |

| | | | | | | | | | | | |
|---|---|---|---|---|---|---|---|---|---|---|---|
| rayyan-751570273 | Thawer SG and Golumbeanu M and Munisi K and Aaron S and Chacky F and Lazaro S and Mohamed A and Kisoka N and Lengeler C and Molteni F and Ross A and Snow RW and Pothin | The use of routine health facility data for micro-stratification of malaria risk in mainland Tanzania. | 2022 | Malaria journal | 21 | 345 | RAYYAN-INCLUSION: {"Richard"=>"Excluded"} \| RAYYAN-EXCLUSION-REASONS: wrong outcome,wrong study design \| USER-NOTES: {"Richard"=>["Stratification study"]} | 10.1186/s12936-022-04364-7 | Child;Humans;Cross-Sectional Studies;Tanzania/epidemiology;*Malaria/epidemiology/prev | PMC9675286 | https://pubmed.ncbi.nlm.nih.gov/36401310/ |
| rayyan-751570274 | Yeboah D and Owusu-Marfo J and Agyeman YN | Predictors of malaria vaccine uptake among children 6-24Â months in the Kassena Nankana Municipality in the Upper East Region of Ghana. | 2022 | Malaria journal | 21 | 339 | RAYYAN-INCLUSION: {"Richard"=>"Excluded"} \| RAYYAN-EXCLUSION-REASONS: wrong outcome,wrong study design \| USER-NOTES: {"Richard"=>["Qual / no epi indicators"]} | 10.1186/s12936-022-04378-1 | Female;Child;Humans;Infant;Young Adult;Adult;*Malaria Vaccines;Ghana/epidemiology; | PMC9666942 | https://pubmed.ncbi.nlm.nih.gov/36384655/ |
| rayyan-751570275 | Turnbull LB and Ayodo G and Knight V and John CC and McHenry MS and Tran TM | Evaluation of an ultrasensitive HRP2-based rapid diagnostic test for detection of asymptomatic Plasmodium falciparum parasitaemia among children in western Kenya. | 2022 | Malaria journal | 21 | 337 | RAYYAN-INCLUSION: {"Richard"=>"Excluded"} \| RAYYAN-EXCLUSION-REASONS: wrong outcome,wrong study design \| USER-NOTES: {"Richard"=>["Diag study"]} | 10.1186/s12936-022-04351-y | Child;Humans;Antigens, Protozoan;Asymptomatic Infections/epidemiology;Diagn | PMC9667565 | https://pubmed.ncbi.nlm.nih.gov/36380379/ |
| rayyan-751570276 | Akpa CO and Azuogu BN and Akpa WC and Umeokonkwo CD and Denwigwe CP and Azuogu VC and Agu AP and Ogah OE and Akamike IC and Ituma IB and Umeokonkwo AA and Onah | Are rural pregnant women disadvantaged in accessing intermittent preventive treatment in pregnancy in Ebonyi State, Nigeria? | 2022 | PloS one | 17 | e0269305 | RAYYAN-INCLUSION: {"Richard"=>"Excluded"} \| RAYYAN-EXCLUSION-REASONS: wrong outcome,wrong study design | 10.1371/journal.pone.0269305 | Female;Humans;Pregnancy;Young Adult;Adult;Sulfadoxine/thera | PMC9648760 | https://pubmed.ncbi.nlm.nih.gov/36355851/ |
| rayyan-751570277 | Bhavnani D and GarcÃ-a Espinosa B and Baird M and Presley N and Le Menach A and Bradley C and Outten M and GonzÃ¡lez O | Malaria surveillance and case management in remote and indigenous communities of Panama: results from a community-based health worker pilot. | 2022 | Malaria journal | 21 | 297 | RAYYAN-INCLUSION: {"Richard"=>"Maybe"} \| RAYYAN-LABELS: Include (full text) \| USER-NOTES: {"Richard"=>["Uncertain whether routine data used"]} | 10.1186/s12936-022-04318-z | Humans;*Community Health Workers;Case Management;*Malaria/diagno | PMC9587600 | https://pubmed.ncbi.nlm.nih.gov/36271383/ |
| rayyan-751570329 | Nankabirwa JI and Rek J and Arinaitwe E and Namuganga JF and Nsobya SL and Asua V and Mawejje HD and Epstein A and Greenhouse B and Rodriguez-Barraquer I and Briggs J and | East Africa International Center of Excellence for Malaria Research: Summary of Key Research Findings. | 2022 | The American journal of | 107 | 21-32 | RAYYAN-INCLUSION: {"Richard"=>"Excluded"} \| RAYYAN-EXCLUSION-REASONS: wrong study design,wrong publication type | 10.4269/ajtmh.21-1285 | Adolescent;Animals;*Antimalarials/pharmacology/therapeutic | PMC9662228 | https://pubmed.ncbi.nlm.nih.gov/36228916/ |
| rayyan-751570330 | Cheng NI and Tabong PT and Netongo PM and Mensah BA and Chu CE and Yaw EB and Enos JY and Malm K and Ahorlu CS | The impact of COVID-19 on implementation of mass testing, treatment and tracking of malaria in rural communities in Ghana: A qualitative study. | 2022 | PloS one | 17 | e0275976 | RAYYAN-INCLUSION: {"Richard"=>"Excluded"} \| RAYYAN-EXCLUSION-REASONS: wrong outcome,wrong study design \| USER-NOTES: {"Richard"=>["Qual study"]} | 10.1371/journal.pone.0275976 | *COVID-19/epidemiology;COVID-19 Testing;Child;Ghana/epidemiol | PMC9560137 | https://pubmed.ncbi.nlm.nih.gov/36227933/ |
| rayyan-751570331 | Hetzel MW and Okitawutshu J and Tshefu A and Omoluabi E and Awor P and Signorell A and Brunner NC and Kalenga JC and Akano BK and Ayodeji K and Okon C and Yusuf O and | Effectiveness of rectal artesunate as pre-referral treatment for severe malaria in children under 5 years of age: a multi-country observational study. | 2022 | BMC medicine | 20 | 343 | RAYYAN-INCLUSION: {"Richard"=>"Maybe"} \| RAYYAN-LABELS: Excluded (after full text) \| USER-NOTES: {"Richard"=>["Unclear if routine data used"]} | 10.1186/s12916-022-02541-8 | *Antimalarials/therapeutic use;*Artemisinins/therapeutic use;Artesunate/therapeutic | PMC9550309 | https://pubmed.ncbi.nlm.nih.gov/36217159/ |
| rayyan-751570332 | Molina-de la Fuente I and Yimar M and GarcÃ-a L and GonzÃ¡lez V and Amor A and Anegagrie M and Benito A and MartÃ-nez J and Moreno M and Berzosa P | Deletion patterns, genetic variability and protein structure of pfhrp2 and pfhrp3: implications for malaria rapid diagnostic test in Amhara region, Ethiopia. | 2022 | Malaria journal | 21 | 287 | RAYYAN-INCLUSION: {"Richard"=>"Excluded"} \| RAYYAN-EXCLUSION-REASONS: wrong outcome,wrong study design \| USER-NOTES: {"Richard"=>["Diag study"]} | 10.1186/s12936-022-04306-3 | Antigens, Protozoan/genetics;Diagnostic Tests, | PMC9548178 | https://pubmed.ncbi.nlm.nih.gov/36209103/ |
| rayyan-751570333 | Nkya TE and Fillinger U and Sangoro OP and Marubu R and Chanda E and Mutero CM | Six decades of malaria vector control in southern Africa: a review of the entomological evidence-base. | 2022 | Malaria journal | 21 | 279 | RAYYAN-INCLUSION: {"Richard"=>"Excluded"} \| RAYYAN-EXCLUSION-REASONS: wrong outcome,wrong study design,wrong publication type \| USER-NOTES: {"Richard"=>["Review"]} | 10.1186/s12936-022-04292-6 | Africa, Southern;Animals;*Anopheles;Humans;*Malaria/prevention | PMC9526912 | https://pubmed.ncbi.nlm.nih.gov/36184603/ |
| rayyan-751570334 | Kande Betu Kumesu V and Mutombo Kalonji W and Bardonneau C and Valverde Mordt O and Ngolo Tete D and Blesson S and Simon F and Delhomme S and Bernhard S and | Safety and efficacy of oral fexinidazole in children with gambiense human African trypanosomiasis: a multicentre, single-arm, open-label, phase 2-3 trial. | 2022 | The Lancet. Global | 10 | e1665-e1674 | RAYYAN-INCLUSION: {"Richard"=>"Excluded"} \| RAYYAN-EXCLUSION-REASONS: wrong outcome,wrong study design,wrong infection / disease | 10.1016/S2214-109X(22)00338-2 | Administration, Oral;Child;Humans;*Nitroimidazoles/administration & | PMC9554014 | https://pubmed.ncbi.nlm.nih.gov/36179736/ |
| rayyan-751570335 | SomÃ© BM and Da DF and McCabe R and DjÃ¨gbÃ¨ NDC and ParÃ© LIG and WermÃ© K and Mouline K and LefÃ¨vre T and OuÃ©draogo AG and Churcher TS and DabirÃ© RK | Adapting field-mosquito collection techniques in a perspective of near-infrared spectroscopy implementation. | 2022 | Parasites & vectors | 15 | 338 | RAYYAN-INCLUSION: {"Richard"=>"Excluded"} \| RAYYAN-EXCLUSION-REASONS: wrong outcome,wrong study design \| USER-NOTES: {"Richard"=>["Ento study"]} | 10.1186/s13071-022-05458-6 | Animals;*Anopheles;Chloroform;Ethanol;Humans;Insecticide Resistance;*Insecticides/phar | PMC9513905 | https://pubmed.ncbi.nlm.nih.gov/36163071/ |
| rayyan-751570336 | Htike W and Oo WH and Lynn T and Sovanda L and Agius PA and Oo MC and Galau NH and Thu KM and Zaw AK and Htwe EP and Cutts JC and Kearney EA and Scott N and O'Flaherty K | Reducing malaria transmission in forest-going mobile and migrant populations in Lao PDR and Cambodia: protocol for stepped-wedge cluster-randomised controlled trial. | 2022 | BMC infectious diseases | 22 | 747 | RAYYAN-INCLUSION: {"Richard"=>"Excluded"} \| RAYYAN-EXCLUSION-REASONS: wrong outcome,wrong study design,wrong publication type \| USER-NOTES: {"Richard"=>["Protocol\n"]} | 10.1186/s12879-022-07724-5 | Cambodia/epidemiology;Forests;Humans;*Insect Repellents;*Insecticides;Laos/ | PMC9509546 | https://pubmed.ncbi.nlm.nih.gov/36153501/ |
| rayyan-751570337 | Allen EN and Wiyeh AB and McCaul M | Adding rapid diagnostic tests to community-based programmes for treating malaria. | 2022 | The Cochrane database | 9 | CD009527 | RAYYAN-INCLUSION: {"Richard"=>"Excluded"} \| RAYYAN-EXCLUSION-REASONS: wrong outcome,wrong study design,wrong publication type \| USER-NOTES: {"Richard"=>["Review"]} | 10.1002/14651858.CD009527.pub3 | Anti-Bacterial Agents/therapeutic use;*Antimalarials/therapeutic | PMC9453882 | https://pubmed.ncbi.nlm.nih.gov/36073718/ |
| rayyan-751570338 | Mangeni JN and Abel L and Taylor SM and Obala A and O'Meara WP and Saran I | Experience and confidence in health technologies: evidence from malaria testing and treatment in Western Kenya. | 2022 | BMC public health | 22 | 1689 | RAYYAN-INCLUSION: {"Richard"=>"Excluded"} \| RAYYAN-EXCLUSION-REASONS: wrong outcome,wrong study design \| USER-NOTES: {"Richard"=>["Diag study"]} | 10.1186/s12889-022-14102-y | Biomedical Technology;Diagnostic Tests, Routine/methods;Family | PMC9446607 | https://pubmed.ncbi.nlm.nih.gov/36068516/ |
| rayyan-751570339 | Kagaya W and Takehara I and Kurihara K and Maina M and Chan CW and Okomo G and Kongere J and Gitaka J and Kaneko A | Potential application of the haematology analyser XN-31 prototype for field malaria surveillance in Kenya. | 2022 | Malaria journal | 21 | 252 | RAYYAN-INCLUSION: {"Richard"=>"Excluded"} \| RAYYAN-EXCLUSION-REASONS: wrong outcome,wrong study design \| USER-NOTES: {"Richard"=>["Diag study"]} | 10.1186/s12936-022-04259-7 | Diagnostic Tests, Routine/methods;*Hematology;Humans;Kenya;*Malaria/dia | PMC9434510 | https://pubmed.ncbi.nlm.nih.gov/36050757/ |
| rayyan-751570340 | Batura N and Kasteng F and Condoane J and Bagorogosa B and Castel-Branco AC and Kertho E and KÃ¤llander K and Soremekun S and Lingam R and Vassall A | Costs of treating childhood malaria, diarrhoea and pneumonia in rural Mozambique and Uganda. | 2022 | Malaria journal | 21 | 239 | RAYYAN-INCLUSION: {"Richard"=>"Excluded"} \| RAYYAN-EXCLUSION-REASONS: wrong outcome,wrong study design \| USER-NOTES: {"Richard"=>["Cost study"]} | 10.1186/s12936-022-04254-y | Child, Preschool;*Diarrhea/epidemiology/therapy;*Health Care | PMC9392282 | https://pubmed.ncbi.nlm.nih.gov/35987625/ |
| rayyan-751570341 | Elagali A and Ahmed A and Makki N and Ismail H and Ajak M and Alene KA and Weiss DJ and Mohammed AA and Abubakr M and Cameron E and Gething P and Elagali A | Spatiotemporal mapping of malaria incidence in Sudan using routine surveillance data. | 2022 | Scientific reports | 12 | 14114 | RAYYAN-INCLUSION: {"Richard"=>"Excluded"} \| RAYYAN-EXCLUSION-REASONS: wrong outcome,wrong study design \| USER-NOTES: {"Richard"=>["Modeling / projection / stratification\n"]} | 10.1038/s41598-022-16706-1 | Animals;Bayes Theorem;Humans;Incidence;*Malaria/epidemiology/preventi | PMC9387890 | https://pubmed.ncbi.nlm.nih.gov/35982088/ |
| rayyan-751570342 | Kiarie H and Temmerman M and Nyamai M and Liku N and Thuo W and Oramisi V and Nyaga L and Karimi J and Wamalwa P and Gatheca G and Mwenda V and Ombajo LA and Thumbi | The COVID-19 pandemic and disruptions to essential health services in Kenya: a retrospective time-series analysis. | 2022 | The Lancet. Global | 10 | e1257-e1267 | RAYYAN-INCLUSION: {"Richard"=>"Maybe"} \| RAYYAN-LABELS: Excluded (C19) \| USER-NOTES: {"Richard"=>["COVID-19 as intervention"]} | 10.1016/S2214-109X(22)00285-6 | *COVID-19/epidemiology;Child;Early Detection of | PMC9363041 | https://pubmed.ncbi.nlm.nih.gov/35961349/ |
| rayyan-751570343 | Mebrahtom S and Worku A and Gage DJ | Causes of infant deaths and patterns of associated factors in Eastern Ethiopia: Results of verbal autopsy (InterVA-4) study. | 2022 | PloS one | 17 | e0270245 | RAYYAN-INCLUSION: {"Richard"=>"Excluded"} \| RAYYAN-EXCLUSION-REASONS: wrong outcome,wrong study design,wrong infection / disease | 10.1371/journal.pone.0270245 | Adult;Autopsy/methods;Cause of Death;Child;Ethiopia/epidemio | PMC9352103 | https://pubmed.ncbi.nlm.nih.gov/35925957/ |
| rayyan-751570345 | Orimadegun AE and Dada-Adegbola HO and Michael OS and Adepoju AA and Funwei RE and Olusola FI and Ajayi IO and Ogunkunle OO and Ademowo OG and Jegede AS and Baba E | Non-Malaria Causes of Fever among under-5 Children with Negative Results for Malaria Rapid Diagnostic Test in South-Western Nigeria. | 2022 | Journal of tropical pediatrics | 68 | | RAYYAN-INCLUSION: {"Richard"=>"Excluded"} \| RAYYAN-EXCLUSION-REASONS: wrong outcome,wrong study design \| USER-NOTES: {"Richard"=>["Diag study"]} | 10.1093/tropej/fmac061 | *Antimalarials/therapeutic use;Child;Diagnostic Tests, Routine/methods;Fever/epide | | https://pubmed.ncbi.nlm.nih.gov/35895093/ |
| rayyan-751570347 | Kagoro FM and Allen E and Mabuza A and Workman L and Magagula R and Kok G and Davies C and Malatje G and GuÃ©rin PJ and Dhorda M and Maude RJ and Raman J and | Making data map-worthy-enhancing routine malaria data to support surveillance and mapping of Plasmodium falciparum anti-malarial resistance in a pre-elimination sub-Saharan African setting: a molecular and spatiotemporal epidemiology study. | 2022 | Malaria journal | 21 | 207 | RAYYAN-INCLUSION: {"Richard"=>"Excluded"} \| RAYYAN-EXCLUSION-REASONS: wrong outcome,wrong study design \| USER-NOTES: {"Richard"=>["Genomic surveillance"]} | 10.1186/s12936-022-04224-4 | *Antimalarials/pharmacology/therapeutic use;Drug Resistance/genetics;Humans;L | PMC9244181 | https://pubmed.ncbi.nlm.nih.gov/35768869/ |
| rayyan-751570348 | Idris IO and Ayeni GO and Iyamu IO and Sina-Odunsi AB and Adebisi YA and Obwoya JG | Factors influencing severity of recurrent malaria in a conflict-affected state of South Sudan: an unmatched case-control study. | 2022 | Conflict and health | 16 | 34 | RAYYAN-INCLUSION: {"Richard"=>"Excluded"} \| RAYYAN-EXCLUSION-REASONS: wrong outcome,wrong study design \| USER-NOTES: {"Richard"=>["No intervention"]} | 10.1186/s13031-022-00463-z | Case-Control Studies | PMC9188688 | https://pubmed.ncbi.nlm.nih.gov/35690836/ |
| rayyan-751570349 | Darkwa S and de Wildt G and Dalaba M and Vidzro E and Ansah EK | "I would have to sell things in order to get the money": A qualitative exploration of willingness to pay for the RTS,S/AS01 malaria vaccine in the Volta region, Ghana. | 2022 | PloS one | 17 | e0268009 | RAYYAN-INCLUSION: {"Richard"=>"Excluded"} \| RAYYAN-EXCLUSION-REASONS: wrong outcome,wrong study design \| USER-NOTES: {"Richard"=>["Qual study"]} | 10.1371/journal.pone.0268009 | Animals;*Anopheles;Child;Cross-Sectional Studies;Ghana;Humans;Infant; | PMC9176758 | https://pubmed.ncbi.nlm.nih.gov/35675273/ |
| rayyan-751570350 | Mooney JP and DonVito SM and Jahateh M and Bittaye H and Bottomley C and D'Alessandro U and Riley EM | Dry season prevalence of Plasmodium falciparum in asymptomatic gambian children, with a comparative evaluation of diagnostic methods. | 2022 | Malaria journal | 21 | 171 | RAYYAN-INCLUSION: {"Richard"=>"Excluded"} \| RAYYAN-EXCLUSION-REASONS: wrong outcome,wrong study design \| USER-NOTES: {"Richard"=>["Diag study"]} | 10.1186/s12936-022-04184-9 | Animals;Asymptomatic Infections/epidemiology;Child;Cross-Sectional | PMC9172138 | https://pubmed.ncbi.nlm.nih.gov/35672850/ |
| rayyan-751570351 | Poespoprodjo JR and Hafiidhaturrahmah and Sariyanti N and Indrawanti R and McLean ARD and Simpson JA and Kenangalem E and Burdam FH and Noviyanti R and Trianty L | Intermittent screening and treatment for malaria complementary to routine immunisation in the first year of life in Papua, Indonesia: a cluster randomised superiority trial. | 2022 | BMC medicine | 20 | 190 | RAYYAN-INCLUSION: {"Richard"=>"Excluded"} \| RAYYAN-EXCLUSION-REASONS: wrong outcome,wrong study design | 10.1186/s12916-022-02394-1 | *Anemia/epidemiology;*Antimalarials/therapeutic use;Child;Female;Humans;Ind | PMC9175359 | https://pubmed.ncbi.nlm.nih.gov/35672703/ |
| rayyan-751570352 | Hanpithakpong W and Day NPJ and White NJ and Tarning J | Simultaneous and enantiospecific quantification of primaquine and carboxyprimaquine in human plasma using liquid chromatography-tandem mass spectrometry. | 2022 | Malaria journal | 21 | 169 | RAYYAN-INCLUSION: {"Richard"=>"Excluded"} \| RAYYAN-EXCLUSION-REASONS: wrong outcome,wrong study design | 10.1186/s12936-022-04191-w | Acetonitriles;Chromatography, High Pressure Liquid;Chromatography, | PMC9166498 | https://pubmed.ncbi.nlm.nih.gov/35659684/ |
| rayyan-751570353 | Babalola OJ and Sesay HW and Blebo LS and Whesseh FK and Umeokonkwo CD and Adewuyi PA and Amo-Addae M | The influence of first wave of COVID-19 outbreak on routine healthcare services, Liberia, August 2020: a mixed study approach. | 2022 | BMC health services | 22 | 684 | RAYYAN-INCLUSION: {"Richard"=>"Maybe"} \| RAYYAN-LABELS: Excluded (C19) \| USER-NOTES: {"Richard"=>["Covid 19 intervention"]} | 10.1186/s12913-022-08074-3 | *COVID-19/epidemiology;Communicable Disease Control;Cross- | PMC9123861 | https://pubmed.ncbi.nlm.nih.gov/35597931/ |
| rayyan-751570354 | de Cola MA and Sawadogo B and Richardson S and Ibinaiye T and TraorÃ© A and CompaorÃ© CS and Oguoma C and Oresanya O and Tougri G and Rassi C and Roca-Feltrer A and | Impact of seasonal malaria chemoprevention on prevalence of malaria infection in malaria indicator surveys in Burkina Faso and Nigeria. | 2022 | BMJ global health | 7 | | RAYYAN-INCLUSION: {"Richard"=>"Excluded"} \| RAYYAN-EXCLUSION-REASONS: wrong study design \| USER-NOTES: {"Richard"=>["No routine data"]} | 10.1136/bmjgh-2021-008021 | *Antimalarials/therapeutic use;Burkina Faso/epidemiology;Chemopre | PMC9121431 | https://pubmed.ncbi.nlm.nih.gov/35589153/ |
| rayyan-751570355 | Rios KT and Dickson TM and Lindner SE | Standard Selection Treatments with Sulfadiazine Limit Plasmodium yoelii Host-to-Vector Transmission. | 2022 | mSphere | 7 | e0010622 | RAYYAN-INCLUSION: {"Richard"=>"Excluded"} \| RAYYAN-EXCLUSION-REASONS: wrong outcome,wrong study design | 10.1128/msphere.00106-22 | 4-Aminobenzoic Acid;Animals;*Anopheles/parasitology;*Malaria/parasitology; | PMC9241536 | https://pubmed.ncbi.nlm.nih.gov/35586987/ |
| rayyan-751570356 | Heuschen AK and Abdul-Mumin A and Adokiya M and Lu G and Jahn A and Razum O and Winkler V and MÃ¼ller O | Impact of the COVID-19 pandemic on malaria cases in health facilities in northern Ghana: a retrospective analysis of routine surveillance data. | 2022 | Malaria journal | 21 | 149 | RAYYAN-INCLUSION: {"Richard"=>"Maybe"} \| RAYYAN-LABELS: Excluded (C19) \| USER-NOTES: {"Richard"=>["COVID intervention"]} | 10.1186/s12936-022-04154-1 | Adult;*COVID-19/epidemiology;Child;Female;Ghana/epidemiology;Health | PMC9107588 | https://pubmed.ncbi.nlm.nih.gov/35570272/ |
| rayyan-751570357 | Ali PO and Okebe J and Oduwole OA | Malaria rapid diagnostic tests for the case management of febrile children in Nigerian primary healthcare settings: a cross-sectional study. | 2022 | The Journal of the | 97 | 10 | RAYYAN-INCLUSION: {"Richard"=>"Excluded"} \| RAYYAN-EXCLUSION-REASONS: wrong outcome,wrong study design | 10.1186/s42506-022-00105-5 | Cross-Sectional Studies;Cesarean Section;Case Management;Malaria | PMC9098722 | https://pubmed.ncbi.nlm.nih.gov/35551536/ |

<table>
<tr><td>rayyan-751570358</td><td>Chaudhary S and Soman B</td><td>Spatiotemporal analysis of environmental and physiographic factors related to malaria in Bareilly district, India.</td><td>2022</td><td>Osong public health</td><td>13</td><td>123-132</td><td>RAYYAN-INCLUSION: {"Richard"=>"Excluded"} | RAYYAN-EXCLUSION-REASONS: wrong outcome,wrong study design | USER-NOTES: {"Richard"=>["Spatio temporal analysis"]}</td><td>10.24171/j.phrp.2021.0304</td><td>India;Malaria</td><td>PMC9091635</td><td>https://pubmed.ncbi.nlm.nih.gov/35538684/</td></tr>
<tr><td>rayyan-751570359</td><td>Bertola M and Fornasiero D and Sgubin S and Mazzon L and Pombi M and Montarsi F</td><td>Comparative efficacy of BG-Sentinel 2 and CDC-like mosquito traps for monitoring potential malaria vectors in Europe.</td><td>2022</td><td>Parasites & vectors</td><td>15</td><td>160</td><td>RAYYAN-INCLUSION: {"Richard"=>"Excluded"} | RAYYAN-EXCLUSION-REASONS: wrong outcome,wrong study design | USER-NOTES: {"Richard"=>["Ento study"]}</td><td>10.1186/s13071-022-05285-9</td><td>*Aedes;Animals;*Anopheles;Carbon Dioxide;Centers for Disease Control and</td><td>PMC9077833</td><td>https://pubmed.ncbi.nlm.nih.gov/35526068/</td></tr>
<tr><td>rayyan-751570361</td><td>Koko VS and Warsame M and Vonhm B and Jeuronlon MK and Menard D and Ma L and Taweh F and Tehmeh L and Nyansaiye P and Pratt OJ and Parwon S and Kamara P and</td><td>Artesunate-amodiaquine and artemether-lumefantrine for the treatment of uncomplicated falciparum malaria in Liberia: in vivo efficacy and frequency of molecular markers.</td><td>2022</td><td>Malaria journal</td><td>21</td><td>134</td><td>RAYYAN-INCLUSION: {"Richard"=>"Excluded"} | RAYYAN-EXCLUSION-REASONS: wrong outcome,wrong study design</td><td>10.1186/s12936-022-04140-7</td><td>Amodiaquine/pharmacology/therapeutic use;*Antimalarials/pharmacol</td><td>PMC9044686</td><td>https://pubmed.ncbi.nlm.nih.gov/35477399/</td></tr>
<tr><td>rayyan-751570362</td><td>Praet N and Asante KP and Bozonnat MC and AkitÃ© EJ and Ansah PO and Baril L and Boahen O and Mendoza YG and Haine V and Kariuki S and Lamy M and Maleta K and</td><td>Assessing the safety, impact and effectiveness of RTS,S/AS01(E) malaria vaccine following its introduction in three sub-Saharan African countries: methodological approaches and study set-up.</td><td>2022</td><td>Malaria journal</td><td>21</td><td>132</td><td>RAYYAN-INCLUSION: {"Richard"=>"Excluded"} | RAYYAN-EXCLUSION-REASONS: wrong outcome,wrong study design</td><td>10.1186/s12936-022-04144-3</td><td>Child;Cross-Sectional Studies;Humans;Infant;Kenya;*Malaria/epidemiology/preven</td><td>PMC9036501</td><td>https://pubmed.ncbi.nlm.nih.gov/35468801/</td></tr>
<tr><td>rayyan-751570363</td><td>Aung YN and Tun STT and Vanisaveth V and Chindavongsa K and Kanya L</td><td>Cost-effectiveness analysis of G6PD diagnostic test for Plasmodium vivax radical cure in Lao PDR: An economic modelling study.</td><td>2022</td><td>PloS one</td><td>17</td><td>e0267193</td><td>RAYYAN-INCLUSION: {"Richard"=>"Excluded"} | RAYYAN-EXCLUSION-REASONS: wrong outcome,wrong study design | USER-NOTES: {"Richard"=>["Cost study"]}</td><td>10.1371/journal.pone.0267193</td><td>*Antimalarials/therapeutic use;Cost-Benefit Analysis;Diagnostic Tests,</td><td>PMC9037946</td><td>https://pubmed.ncbi.nlm.nih.gov/35468145/</td></tr>
<tr><td>rayyan-751570364</td><td>Parsons GJI and Lees RS and Balaska S and Vontas J</td><td>A Practical Insecticide Resistance Monitoring Bioassay for Orally Ingested Dinotefuran in Anopheles Malaria Vectors.</td><td>2022</td><td>Insects</td><td>13</td><td></td><td>RAYYAN-INCLUSION: {"Richard"=>"Excluded"} | RAYYAN-EXCLUSION-REASONS: wrong outcome,wrong study design | USER-NOTES: {"Richard"=>["Ento study"]}</td><td>10.3390/insects13040311</td><td>Biological Assay;Malaria</td><td>PMC9025404</td><td>https://pubmed.ncbi.nlm.nih.gov/35447753/</td></tr>
<tr><td>rayyan-751570365</td><td>Requejo J and Strong K and Agweyu A and Billah SM and Boschi-Pinto C and Horiuchi S and Jamaluddine Z and Lazzerini M and Maiga A and McKerrow N and Munos M and Park L and</td><td>Measuring and monitoring child health and wellbeing: recommendations for tracking progress with a core set of indicators in the Sustainable Development Goals era.</td><td>2022</td><td>The Lancet. Child &</td><td>6</td><td>345-352</td><td>RAYYAN-INCLUSION: {"Richard"=>"Excluded"} | RAYYAN-EXCLUSION-REASONS: wrong outcome,wrong study design,wrong infection / disease</td><td>10.1016/S2352-4642(22)00039-6</td><td>*COVID-19;Child;Child Health;Chronic Disease;Humans;Pandemics;*</td><td>PMC9764429</td><td>https://pubmed.ncbi.nlm.nih.gov/35429452/</td></tr>
<tr><td>rayyan-751570366</td><td>Dorrucci M and Boccolini D and Bella A and Lucarelli C and D'Amato S and Caraglia A and Maraglino FP and Severini C and Gradoni L and Pezzotti P</td><td>Malaria surveillance system and Hospital Discharge Records: Assessing differences in Italy, 2011-2017 database analysis.</td><td>2022</td><td>Travel medicine and</td><td>48</td><td>102322</td><td>RAYYAN-INCLUSION: {"Richard"=>"Excluded"} | RAYYAN-EXCLUSION-REASONS: wrong outcome,wrong study design</td><td>10.1016/j.tmaid.2022.102322</td><td>Adult;Databases, Factual;Hospitals;Humans;Italy/epidemiology;*Malaria/preve</td><td></td><td>https://pubmed.ncbi.nlm.nih.gov/35367383/</td></tr>
<tr><td>rayyan-751570367</td><td>Chanda-Kapata P and Ntoumi F and Kapata N and Lungu P and Mucheleng'anga LA and Chakaya J and Tembo J and Himwaze C and Ansumana R and Asogun D and Mfinanga S and Nyasulu</td><td>Tuberculosis, HIV/AIDS and Malaria Health Services in sub-Saharan Africa - A Situation Analysis of the Disruptions and Impact of the COVID-19 Pandemic.</td><td>2022</td><td>International journal of</td><td>124</td><td>S41-S46</td><td>RAYYAN-INCLUSION: {"Richard"=>"Excluded"} | RAYYAN-EXCLUSION-REASONS: wrong outcome,wrong study design</td><td>10.1016/j.ijid.2022.03.033</td><td>Humans;Pandemics/prevention & control;*COVID-19/epidemiology;*HIV</td><td>PMC8949686</td><td>https://pubmed.ncbi.nlm.nih.gov/35341998/</td></tr>
<tr><td>rayyan-751570368</td><td>Michael OS and Orimadegun AE and Falade CO</td><td>PERSISTENCE OF PLASMODIUM FALCIPARUM HRP2 ANTIGEN AFTER EFFECTIVE ANTIMALARIAL THERAPY.</td><td>2021</td><td>Annals of Ibadan postgrad</td><td>19</td><td>15-21</td><td>RAYYAN-INCLUSION: {"Richard"=>"Excluded"} | RAYYAN-EXCLUSION-REASONS: wrong outcome,wrong study design</td><td></td><td>Plasmodium falciparum</td><td>PMC8935674</td><td>https://pubmed.ncbi.nlm.nih.gov/35330886/</td></tr>
<tr><td>rayyan-751570370</td><td>Roh ME and Mpimbaza A and Oundo B and Irish A and Murphy M and Wu SL and White JS and Shiboski S and Glymour MM and Gosling R and Dorsey G and Sturrock H</td><td>Association between indoor residual spraying and pregnancy outcomes: a quasi-experimental study from Uganda.</td><td>2022</td><td>International journal of</td><td>51</td><td>1489-1501</td><td>RAYYAN-INCLUSION: {"Richard"=>"Maybe"} | RAYYAN-LABELS: Include (full text) | USER-NOTES: {"Richard"=>["IRS on pregnancy outcomes"]}</td><td>10.1093/ije/dyac043</td><td>Disease Progression;Female;Humans;*Insecticides;*Malaria/epidemi</td><td>PMC9557839</td><td>https://pubmed.ncbi.nlm.nih.gov/35301532/</td></tr>
<tr><td>rayyan-751570371</td><td>Obadia T and Nekkab N and Robinson LJ and Drakeley C and Mueller I and White MT</td><td>Developing sero-diagnostic tests to facilitate Plasmodium vivax Serological Test-and-Treat approaches: modeling the balance between public health impact and overtreatment.</td><td>2022</td><td>BMC medicine</td><td>20</td><td>98</td><td>RAYYAN-INCLUSION: {"Richard"=>"Excluded"} | RAYYAN-EXCLUSION-REASONS: wrong outcome,wrong study design | USER-NOTES: {"Richard"=>["Diag study"]}</td><td>10.1186/s12916-022-02285-5</td><td>*Diagnostic Tests, Routine;Humans;Overtreatment;*Plasmodium vivax;Public</td><td>PMC8932240</td><td>https://pubmed.ncbi.nlm.nih.gov/35300700/</td></tr>
<tr><td>rayyan-751570372</td><td>Fatunla OAT and Olatunya OS and Ogundare EO and Fatunla TO and Babatola AO and Adeniyi AT and Oyelami OA</td><td>Malaria prevention practices and malaria prevalence among children living in a rural community in Southwest Nigeria.</td><td>2022</td><td>Journal of infection in</td><td>16</td><td>352-361</td><td>RAYYAN-INCLUSION: {"Richard"=>"Excluded"} | RAYYAN-EXCLUSION-REASONS: wrong outcome,wrong study design | USER-NOTES: {"Richard"=>["No routine data"]}</td><td>10.3855/jidc.14894</td><td>Adolescent;Child;Child, Preschool;Cross-Sectional Studies;Humans;Infant;*Malar</td><td></td><td>https://pubmed.ncbi.nlm.nih.gov/35298432/</td></tr>
<tr><td>rayyan-751570373</td><td>Balla K and Malm K and Njie O and Hounto Ogouyemi A and Uhomoibhi P and Poku-Awuku A and Tchouatieu AM and Aikpon R and Bah A and Kolley O and Ogbulafor N and</td><td>Introducing field digital data collection systems into seasonal malaria chemoprevention campaigns: opportunities for robust evidence development and national e-health strategies.</td><td>2022</td><td>BMJ global health</td><td>7</td><td></td><td>RAYYAN-INCLUSION: {"Richard"=>"Excluded"} | RAYYAN-EXCLUSION-REASONS: wrong outcome,wrong study design | USER-NOTES: {"Richard"=>["No epi indictator"]}</td><td>10.1136/bmjgh-2021-007899</td><td>*Antimalarials/therapeutic use;Chemoprevention;Child;Data</td><td>PMC8928254</td><td>https://pubmed.ncbi.nlm.nih.gov/35296463/</td></tr>
<tr><td>rayyan-751570374</td><td>Karim A and de Savigny D and Ngaima JS and MÃ¤usezahl D and Cobos MuÃ±oz D and Tshefu A</td><td>Assessing Determinants of Programmatic Performance of Community Management of Malaria, Pneumonia, and Diarrhea in Children in Africa: Protocol and Data Collection for a Mixed Methods Evaluation of Integrated Community Case Management.</td><td>2022</td><td>JMIR research protocols</td><td>11</td><td>e33076</td><td>RAYYAN-INCLUSION: {"Richard"=>"Excluded"} | RAYYAN-EXCLUSION-REASONS: wrong study design,wrong publication type | USER-NOTES: {"Richard"=>["Protocol"]}</td><td>10.2196/33076</td><td>Diarrhea</td><td>PMC8961344</td><td>https://pubmed.ncbi.nlm.nih.gov/35285813/</td></tr>
<tr><td>rayyan-751570375</td><td>Marita E and Langat B and Kinyari T and Igunza P and Apat D and Kimori J and Carter J and Kiplimo R and Muhula S</td><td>Implementation of community case management of malaria in malaria endemic counties of western Kenya: are community health volunteers up to the task in diagnosing malaria?</td><td>2022</td><td>Malaria journal</td><td>21</td><td>73</td><td>RAYYAN-INCLUSION: {"Richard"=>"Excluded"} | RAYYAN-EXCLUSION-REASONS: wrong outcome,wrong study design | USER-NOTES: {"Richard"=>["Diag study"]}</td><td>10.1186/s12936-022-04094-w</td><td>Adult;Aged;*Antimalarials/therapeutic use;Artemether/therapeutic</td><td>PMC8897909</td><td>https://pubmed.ncbi.nlm.nih.gov/35248055/</td></tr>
<tr><td>rayyan-751570376</td><td>Fitri LE and Widaningrum T and Endharti AT and Prabowo MH and Winaris N and Nugraha RYB</td><td>Malaria diagnostic update: From conventional to advanced method.</td><td>2022</td><td>Journal of clinical laborator</td><td>36</td><td>e24314</td><td>RAYYAN-INCLUSION: {"Richard"=>"Excluded"} | RAYYAN-EXCLUSION-REASONS: wrong outcome,wrong study design,wrong publication type</td><td>10.1002/jcla.24314</td><td>Diagnostic Tests, Routine/methods;Humans;*Malaria/diagnosis;Microscopy/m</td><td>PMC8993657</td><td>https://pubmed.ncbi.nlm.nih.gov/35247002/</td></tr>
<tr><td>rayyan-751570377</td><td>Niba PTN and Nji AM and Ali IM and Akam LF and Dongmo CH and Chedjou JPK and Fomboh CT and Nana WD and Oben OLA and Selly-Ngaloumo AA and Moyeh MN and Ngu JA and</td><td>Effectiveness and safety of artesunate-amodiaquine versus artemether-lumefantrine for home-based treatment of uncomplicated Plasmodium falciparum malaria among children 6-120Â months in YaoundÃ©, Cameroon: a randomized trial.</td><td>2022</td><td>BMC infectious diseases</td><td>22</td><td>166</td><td>RAYYAN-INCLUSION: {"Richard"=>"Excluded"} | RAYYAN-EXCLUSION-REASONS: wrong outcome,wrong study design | USER-NOTES: {"Richard"=>["No routine data"]}</td><td>10.1186/s12879-022-07101-2</td><td>Amodiaquine/adverse effects;*Antimalarials/adverse effects;Artemether/therapeuti</td><td>PMC8862275</td><td>https://pubmed.ncbi.nlm.nih.gov/35189818/</td></tr>
<tr><td>rayyan-751570378</td><td>Kumoji E' and Awantang GN and Toso M and Kamara D and Bleu T and Lahai W and Sillah-Kanu M and Dosso A and Achu D and Babalola S</td><td>Ideational factors associated with net care behaviour: a multi-country analysis.</td><td>2022</td><td>Malaria journal</td><td>21</td><td>53</td><td>RAYYAN-INCLUSION: {"Richard"=>"Excluded"} | RAYYAN-EXCLUSION-REASONS: wrong outcome,wrong study design | USER-NOTES: {"Richard"=>["No epi indicator"]}</td><td>10.1186/s12936-022-04053-5</td><td>Cameroon;Cote d'Ivoire/epidemiology;Family Characteristics;Female;Human</td><td>PMC8851768</td><td>https://pubmed.ncbi.nlm.nih.gov/35177086/</td></tr>
<tr><td>rayyan-751570429</td><td>Kumari S and Ahmed MZ and Sharma S and Pande V and Anvikar AR</td><td>Prevalence of Pfhrp2/3 gene deletions among false negative rapid antigen test results in central India.</td><td>2021</td><td>Journal of vector borne</td><td>58</td><td>273-280</td><td>RAYYAN-INCLUSION: {"Richard"=>"Excluded"} | RAYYAN-EXCLUSION-REASONS: wrong outcome,wrong study design | USER-NOTES: {"Richard"=>["Diag study"]}</td><td>10.4103/0972-9062.328815</td><td>*Antigens, Protozoan/genetics;Diagnostic Tests, Routine;Gene</td><td></td><td>https://pubmed.ncbi.nlm.nih.gov/35170466/</td></tr>
<tr><td>rayyan-751570430</td><td>Yimam Y and Mohebali M and Abbaszadeh Afshar MJ</td><td>Comparison of diagnostic performance between conventional and ultrasensitive rapid diagnostic tests for diagnosis of malaria: A systematic review and meta-analysis.</td><td>2022</td><td>PloS one</td><td>17</td><td>e0263770</td><td>RAYYAN-INCLUSION: {"Richard"=>"Excluded"} | RAYYAN-EXCLUSION-REASONS: wrong outcome,wrong study design,wrong publication type | USER-NOTES: {"Richard"=>["Review"]}</td><td>10.1371/journal.pone.0263770</td><td>Diagnostic Tests, Routine/*methods;Humans;Malaria/*diagnosis;Reagent Kits,</td><td>PMC8830612</td><td>https://pubmed.ncbi.nlm.nih.gov/35143565/</td></tr>
<tr><td>rayyan-751570431</td><td>Vezenegho SB and Issaly J and Carinci R and Gaborit P and Girod R and Dusfour I and Briolant S</td><td>Discrimination of 15 Amazonian Anopheline Mosquito Species by Polymerase Chain Reaction-Restriction Fragment Length Polymorphism.</td><td>2022</td><td>Journal of medical entomolo</td><td>59</td><td>1060-1064</td><td>RAYYAN-INCLUSION: {"Richard"=>"Excluded"} | RAYYAN-EXCLUSION-REASONS: wrong outcome,wrong study design | USER-NOTES: {"Richard"=>["Ento study"]}</td><td>10.1093/jme/tjac008</td><td>Animals;*Anopheles/genetics;DNA, Ribosomal Spacer/genetics;*Malaria;Mos</td><td></td><td>https://pubmed.ncbi.nlm.nih.gov/35139212/</td></tr>
<tr><td>rayyan-751570432</td><td>Kamau A and Paton RS and Akech S and Mpimbaza A and Khazenzi C and Ogero M and Mumo E and Alegana VA and Agweyu A and Mturi N and Mohammed S and Bigogo G and</td><td>Malaria hospitalisation in East Africa: age, phenotype and transmission intensity.</td><td>2022</td><td>BMC medicine</td><td>20</td><td>28</td><td>RAYYAN-INCLUSION: {"Richard"=>"Excluded"} | RAYYAN-EXCLUSION-REASONS: wrong outcome,wrong study design | USER-NOTES: {"Richard"=>["No intervention data"]}</td><td>10.1186/s12916-021-02224-w</td><td>Adolescent;Africa, Eastern/epidemiology;Bayes Theorem;Child;Child,</td><td>PMC8793189</td><td>https://pubmed.ncbi.nlm.nih.gov/35081974/</td></tr>
<tr><td>rayyan-751570433</td><td>Hosch S and Yoboue CA and Donfack OT and Guirou EA and Dangy JP and Mpina M and Nyakurungu E and BlÃ¶chliger K and Guerra CA and Phiri WP and Ayekaba MO and GarcÃ-a GA</td><td>Analysis of nucleic acids extracted from rapid diagnostic tests reveals a significant proportion of false positive test results associated with recent malaria treatment.</td><td>2022</td><td>Malaria journal</td><td>21</td><td>23</td><td>RAYYAN-INCLUSION: {"Richard"=>"Excluded"} | RAYYAN-EXCLUSION-REASONS: wrong outcome,wrong study design</td><td>10.1186/s12936-022-04043-7</td><td>Antigens, Protozoan/*analysis;Coinfection/*diagnosis/epidemiology;Dia</td><td>PMC8785039</td><td>https://pubmed.ncbi.nlm.nih.gov/35073934/</td></tr>
<tr><td>rayyan-751570434</td><td>Sy M and Deme AB and Warren JL and Early A and Schaffner S and Daniels RF and Dieye B and Ndiaye IM and Diedhiou Y and Mbaye AM and Volkman SK and Hartl DL and Wirth DF and</td><td>Plasmodium falciparum genomic surveillance reveals spatial and temporal trends, association of genetic and physical distance, and household clustering.</td><td>2022</td><td>Scientific reports</td><td>12</td><td>938</td><td>RAYYAN-INCLUSION: {"Richard"=>"Excluded"} | RAYYAN-EXCLUSION-REASONS: wrong outcome,wrong study design | USER-NOTES: {"Richard"=>["Genomic study"]}</td><td>10.1038/s41598-021-04572-2</td><td>Adolescent;Animals;Child;Child, Preschool;Cluster Analysis;Cohort</td><td>PMC8766587</td><td>https://pubmed.ncbi.nlm.nih.gov/35042879/</td></tr>
<tr><td>rayyan-751570435</td><td>Oliveira KRHM and Torres MLM and Kauffmann N and de Azevedo AtaÃ-de BJ and de Souza Franco Mendes N and Dos Anjos LM and Dos Santos Borges R and Bahia CP and LeÃ£o</td><td>Euterpe oleracea fruit (AÃ§ai)-enriched diet suppresses the development of experimental cerebral malaria induced by Plasmodium berghei (ANKA) infection.</td><td>2022</td><td>BMC complementary</td><td>22</td><td>11</td><td>RAYYAN-INCLUSION: {"Richard"=>"Excluded"} | RAYYAN-EXCLUSION-REASONS: wrong outcome,wrong study design</td><td>10.1186/s12906-021-03495-9</td><td>Animal Feed;Animals;Behavioral Symptoms/etiology/preventio</td><td>PMC8751313</td><td>https://pubmed.ncbi.nlm.nih.gov/35016657/</td></tr>
<tr><td>rayyan-751570436</td><td>Hupert N and MarÃ-n-HernÃ¡ndez D and Gao B and Ãguas R and Nixon DF</td><td>Heterologous vaccination interventions to reduce pandemic morbidity and mortality: Modeling the US winter 2020 COVID-19 wave.</td><td>2022</td><td>Proceedings of the National</td><td>119</td><td></td><td>RAYYAN-INCLUSION: {"Richard"=>"Excluded"} | RAYYAN-EXCLUSION-REASONS: wrong outcome,wrong study design,wrong infection / disease</td><td>10.1073/pnas.2025448119</td><td>Algorithms;BCG Vaccine/administration & dosage/immunology;COVID-</td><td>PMC8784160</td><td>https://pubmed.ncbi.nlm.nih.gov/35012976/</td></tr>
<tr><td>rayyan-751570437</td><td>GansanÃ© A and Candrinho B and Mbituyumuremyi A and Uhomoibhi P and NFalÃ© S and Mohammed AB and Guelbeogo WM and Sanou A and Kangoye D and Debe S and</td><td>Design and methods for a quasi-experimental pilot study to evaluate the impact of dual active ingredient insecticide-treated nets on malaria burden in five regions in sub-Saharan Africa.</td><td>2022</td><td>Malaria journal</td><td>21</td><td>19</td><td>RAYYAN-INCLUSION: {"Richard"=>"Excluded"} | RAYYAN-EXCLUSION-REASONS: wrong outcome,wrong study design,wrong publication type | USER-NOTES: {"Richard"=>["Protocol"]}</td><td>10.1186/s12936-021-04026-0</td><td>Africa South of the Sahara/epidemiology;*Cost of Illness;Humans;Incidence;Inse</td><td>PMC8744060</td><td>https://pubmed.ncbi.nlm.nih.gov/35012559/</td></tr>
<tr><td>rayyan-751570438</td><td>Maher SP and Vantaux A and Cooper CA and Chasen NM and Cheng WT and Joyner CJ and Manetsch R and Witkowski B and Kyle D</td><td>A Phenotypic Screen for the Liver Stages of Plasmodium vivax.</td><td>2021</td><td>Bio-protocol</td><td>11</td><td>e4253</td><td>RAYYAN-INCLUSION: {"Richard"=>"Excluded"} | RAYYAN-EXCLUSION-REASONS: wrong outcome,wrong study design</td><td>10.21769/BioProtoc.4253</td><td>Phenotype;Liver;Plasmodium</td><td>PMC8678555</td><td>https://pubmed.ncbi.nlm.nih.gov/35005096/</td></tr>
<tr><td>rayyan-751570440</td><td>Namuganga JF and Briggs J and Roh ME and Okiring J and Kisambira Y and Sserwanga A and Kapisi JA and Arinaitwe E and Ebong C and Ssewanyana I and Maiteki-Ssebuguzi C and</td><td>Impact of COVID-19 on routine malaria indicators in rural Uganda: an interrupted time series analysis.</td><td>2021</td><td>Malaria journal</td><td>20</td><td>475</td><td>RAYYAN-INCLUSION: {"Richard"=>"Included"} | RAYYAN-LABELS: Excluded (C19) | USER-NOTES: {"Richard"=>["COVID19 intervention"]}</td><td>10.1186/s12936-021-04018-0</td><td>*Ambulatory Care;COVID-19/*epidemiology;Chronic Disease</td><td>PMC8685800</td><td>https://pubmed.ncbi.nlm.nih.gov/34930317/</td></tr>
<tr><td>rayyan-751570441</td><td>Dambach P and TraorÃ© I and Sawadogo H and ZabrÃ© P and Shukla S and Sauerborn R and Becker N and Phalkey R</td><td>Community acceptance of environmental larviciding against malaria with Bacillus thuringiensis israelensis in rural Burkina Faso - A knowledge, attitudes and practices study.</td><td>2021</td><td>Global health action</td><td>14</td><td>1988279</td><td>RAYYAN-INCLUSION: {"Richard"=>"Excluded"} | RAYYAN-EXCLUSION-REASONS: wrong outcome,wrong study design | USER-NOTES: {"Richard"=>["Qual study"]}</td><td>10.1080/16549716.2021.1988279</td><td>Animals;*Anopheles;*Bacillus thuringiensis;Burkina Faso;Cross-Sectional</td><td>PMC8725727</td><td>https://pubmed.ncbi.nlm.nih.gov/34927578/</td></tr>
<tr><td>rayyan-751570442</td><td>Leuba SI and Westreich D and Bose CL and Powers KA and Olshan A and Taylor SM and Tshefu A and Lokangaka A and Carlo WA and Chomba E and Liechty EA and Bucher SL and</td><td>Predictors of Plasmodium falciparum Infection in the First Trimester Among Nulliparous Women From Kenya, Zambia, and the Democratic Republic of the Congo.</td><td>2022</td><td>The Journal of infectious</td><td>225</td><td>2002-2010</td><td>RAYYAN-INCLUSION: {"Richard"=>"Excluded"} | RAYYAN-EXCLUSION-REASONS: wrong outcome,wrong study design</td><td>10.1093/infdis/jiab588</td><td>Aspirin/therapeutic use;Democratic Republic of the</td><td>PMC9159331</td><td>https://pubmed.ncbi.nlm.nih.gov/34888658/</td></tr>
</table>

<table>
<tr><td>rayyan-751570443</td><td>Kigozi RN and Bwanika J and Goodwin E and Thomas P and Bukoma P and Nabyonga P and Isabirye F and Oboth P and Kyozira C and Niang M and Belay K and Sebikaari G and</td><td>Determinants of malaria testing at health facilities: the case of Uganda.</td><td>2021</td><td>Malaria journal</td><td>20</td><td>456</td><td>RAYYAN-INCLUSION: {"Richard"=>"Excluded"} | RAYYAN-EXCLUSION-REASONS: wrong outcome,wrong study design</td><td>10.1186/s12936-021-03992-9</td><td>Antimalarials/*supply & distribution;Artemether, Lumefantrine Drug</td><td>PMC8645102</td><td>https://pubmed.ncbi.nlm.nih.gov/34863172/</td></tr>
<tr><td>rayyan-751570444</td><td>Marwa KJ and Konje ET and Kapesa A and Kamugisha E and Mwita S and Swedberg G</td><td>Artemether-lumefantrine and dihydroartemisinin-piperaquine treatment outcomes among children infected with uncomplicated Plasmodium falciparum malaria in Mwanza, Tanzania.</td><td>2021</td><td>Tropical medicine and</td><td>49</td><td>94</td><td>RAYYAN-INCLUSION: {"Richard"=>"Excluded"} | RAYYAN-EXCLUSION-REASONS: wrong outcome,wrong study design</td><td>10.1186/s41182-021-00383-3</td><td>Malaria, Falciparum;Plasmodium falciparum;Plasmodium</td><td>PMC8627025</td><td>https://pubmed.ncbi.nlm.nih.gov/34838151/</td></tr>
<tr><td>rayyan-751570445</td><td>Ciccone EJ and Kabugho L and Baguma E and Muhindo R and Juliano JJ and Mulogo E and Boyce RM</td><td>Rapid Diagnostic Tests to Guide Case Management of and Improve Antibiotic Stewardship for Pediatric Acute Respiratory Illnesses in Resource-Constrained Settings: a Prospective Cohort Study in Southwestern Uganda.</td><td>2021</td><td>Microbiol ogy spectrum</td><td>9</td><td>e0169421</td><td>RAYYAN-INCLUSION: {"Richard"=>"Excluded"} | RAYYAN-EXCLUSION-REASONS: wrong outcome,wrong study design</td><td>10.1128/Spectrum.01694-21</td><td>Anti-Bacterial Agents/*therapeutic use;Antimicrobial</td><td>PMC8612158</td><td>https://pubmed.ncbi.nlm.nih.gov/34817224/</td></tr>
<tr><td>rayyan-751570446</td><td>Mwangi MN and Mzembe G and Moya E and Braat S and Harding R and Robberstad B and Simpson J and Stones W and Rogerson S and Biselele K and Chinkhumba J and Larson L and</td><td>Protocol for a multicentre, parallel-group, open-label randomised controlled trial comparing ferric carboxymaltose with the standard of care in anaemic Malawian pregnant women: the REVAMP trial.</td><td>2021</td><td>BMJ open</td><td>11</td><td>e053288</td><td>RAYYAN-INCLUSION: {"Richard"=>"Excluded"} | RAYYAN-EXCLUSION-REASONS: wrong outcome,wrong study design</td><td>10.1136/bmjopen-2021-053288</td><td>*Anemia/drug therapy;*Anemia, Iron-Deficiency/drug</td><td>PMC8611444</td><td>https://pubmed.ncbi.nlm.nih.gov/34815287/</td></tr>
<tr><td>rayyan-751570447</td><td>MoukÃ©net A and de Cola MA and Ward C and BeakgoubÃ© H and Baker K and Donovan L and LaoukolÃ© J and Richardson S</td><td>Health management information system (HMIS) data quality and associated factors in Massaguet district, Chad.</td><td>2021</td><td>BMC medical informati</td><td>21</td><td>326</td><td>RAYYAN-INCLUSION: {"Richard"=>"Excluded"} | RAYYAN-EXCLUSION-REASONS: wrong outcome,wrong study design | USER-NOTES: {"Richard"=>["No intervention data"]}</td><td>10.1186/s12911-021-01684-7</td><td>Chad/epidemiology;Child;Cross-Sectional Studies;*Data Accuracy;Humans;Infant;*Man</td><td>PMC8609810</td><td>https://pubmed.ncbi.nlm.nih.gov/34809622/</td></tr>
<tr><td>rayyan-751570448</td><td>Obasohan PE and Walters SJ and Jacques R and Khatab K</td><td>Individual and Contextual Factors Associated with Malaria among Children 6-59 Months in Nigeria: A Multilevel Mixed Effect Logistic Model Approach.</td><td>2021</td><td>Internatio nal journal of</td><td>18</td><td></td><td>RAYYAN-INCLUSION: {"Richard"=>"Excluded"} | RAYYAN-EXCLUSION-REASONS: wrong outcome,wrong study design</td><td>10.3390/ijerph182111234</td><td>Child;Child, Preschool;Diagnostic Tests, Routine;Humans;Logistic</td><td>PMC8582856</td><td>https://pubmed.ncbi.nlm.nih.gov/34769754/</td></tr>
<tr><td>rayyan-751570449</td><td>Richardson S and Moukenet A and Diar MSI and de Cola MA and Rassi C and Counihan H and Roca-Feltrer A</td><td>Modeled Impact of Seasonal Malaria Chemoprevention on District-Level Suspected and Confirmed Malaria Cases in Chad Based on Routine Clinical Data (2013-2018).</td><td>2021</td><td>The American journal of</td><td>105</td><td>1712-1721</td><td>RAYYAN-INCLUSION: {"Richard"=>"Included"}</td><td>10.4269/ajtmh.21-0314</td><td>Amodiaquine/*therapeutic use;Antimalarials/*therapeutic use;Chad/epidemiology;Chem</td><td>PMC8641328</td><td>https://pubmed.ncbi.nlm.nih.gov/34662864/</td></tr>
<tr><td>rayyan-751570450</td><td>Xia J and Fan L and Si L and Lu Q and Zhao Y and Zhang M and Xu Y and Wang B</td><td>A Case Report of Serological Rapid Diagnostic Test-Negative Plasmodium malariae Malaria Imported from West Africa.</td><td>2021</td><td>Clinical laborator y</td><td>67</td><td></td><td>RAYYAN-INCLUSION: {"Richard"=>"Excluded"} | RAYYAN-EXCLUSION-REASONS: wrong outcome,wrong study design,wrong publication type | USER-NOTES: {"Richard"=>["Case report"]}</td><td>10.7754/Clin.Lab.2021.210208</td><td>Africa, Western;Diagnostic Tests, Routine;Humans;*Malaria/dia</td><td></td><td>https://pubmed.ncbi.nlm.nih.gov/34655199/</td></tr>
<tr><td>rayyan-751570451</td><td>Mosha D and Kakolwa MA and Mahende MK and Masanja H and Abdulla S and Drakeley C and Gosling R and Wamoyi J</td><td>Safety monitoring experience of single-low dose primaquine co-administered with artemether-lumefantrine among providers and patients in routine healthcare practice: a qualitative study in Eastern Tanzania.</td><td>2021</td><td>Malaria journal</td><td>20</td><td>392</td><td>RAYYAN-INCLUSION: {"Richard"=>"Excluded"} | RAYYAN-EXCLUSION-REASONS: wrong outcome,wrong study design</td><td>10.1186/s12936-021-03921-w</td><td>Adult;Antimalarials/*administration & dosage/standards;Artemether,</td><td>PMC8501629</td><td>https://pubmed.ncbi.nlm.nih.gov/34627236/</td></tr>
<tr><td>rayyan-751570452</td><td>Feleke DG and Alemu Y and Yemanebirhane N</td><td>Performance of rapid diagnostic tests, microscopy, loop-mediated isothermal amplification (LAMP) and PCR for malaria diagnosis in Ethiopia: a systematic review and meta-analysis.</td><td>2021</td><td>Malaria journal</td><td>20</td><td>384</td><td>RAYYAN-INCLUSION: {"Richard"=>"Excluded"} | RAYYAN-EXCLUSION-REASONS: wrong outcome,wrong study design,wrong publication type | USER-NOTES: {"Richard"=>["Review"]}</td><td>10.1186/s12936-021-03923-8</td><td>Diagnostic Tests, Routine/*statistics & numerical</td><td>PMC8474705</td><td>https://pubmed.ncbi.nlm.nih.gov/34579729/</td></tr>
<tr><td>rayyan-751570453</td><td>Ali IM and Nji AM and Bonkum JC and Moyeh MN and Carole GK and Efon A and Dabou S and Tchuenkam VPK and Tah C and Kengne JC and Achu DF and Bigoga JD and Mbacham WF</td><td>Diagnostic Accuracy of CareStartâ„¢ Malaria HRP2 and SD Bioline Pf/PAN for Malaria in Febrile Outpatients in Varying Malaria Transmission Settings in Cameroon.</td><td>2021</td><td>Diagnosti cs (Basel, Switzerla</td><td>11</td><td></td><td>RAYYAN-INCLUSION: {"Richard"=>"Excluded"} | RAYYAN-EXCLUSION-REASONS: wrong outcome,wrong study design | USER-NOTES: {"Richard"=>["Diag atudy"]}</td><td>10.3390/diagnostics11091556</td><td>Cameroon;Malaria</td><td>PMC8469216</td><td>https://pubmed.ncbi.nlm.nih.gov/34573898/</td></tr>
<tr><td>rayyan-751570454</td><td>Cairns M and Ceesay SJ and Sagara I and Zongo I and Kessely H and Gamougam K and Diallo A and Ogboi JS and Moroso D and Van Hulle S and Eloike T and Snell P and Scott S and Merle</td><td>Effectiveness of seasonal malaria chemoprevention (SMC) treatments when SMC is implemented at scale: Case-control studies in 5 countries.</td><td>2021</td><td>PLoS medicine</td><td>18</td><td>e1003727</td><td>RAYYAN-INCLUSION: {"Richard"=>"Maybe"} | RAYYAN-LABELS: Excluded (after full text) | USER-NOTES: {"Richard"=>["Unclear if routine data used"]}</td><td>10.1371/journal.pmed.1003727</td><td>Africa, Western/epidemiology;Age Factors;Amodiaquine/adverse</td><td>PMC8457484</td><td>https://pubmed.ncbi.nlm.nih.gov/34495978/</td></tr>
<tr><td>rayyan-751570455</td><td>Kim S and Luande VN and RocklÃ¶v J and Carlton JM and Tozan Y</td><td>A Systematic Review of the Evidence on the Effectiveness and Cost-Effectiveness of Mass Screen-and-Treat Interventions for Malaria Control.</td><td>2021</td><td>The American journal of</td><td>105</td><td>1722-1731</td><td>RAYYAN-INCLUSION: {"Richard"=>"Excluded"} | RAYYAN-EXCLUSION-REASONS: wrong study design,wrong publication type | USER-NOTES: {"Richard"=>["Review"]}</td><td>10.4269/ajtmh.21-0325</td><td>Africa South of the Sahara/epidemiology;Antimalarials/*therapeutic</td><td>PMC8641306</td><td>https://pubmed.ncbi.nlm.nih.gov/34491213/</td></tr>
<tr><td>rayyan-751570456</td><td>Francis S and Frank C and Buchanan L and Green S and Stennett-Brown R and Gordon-Strachan G and Rubio-Palis Y and Grant C and Alexander-Lindo RL and Nwokocha C and</td><td>Challenges in the control of neglected insect vector diseases of human importance in the Anglo-Caribbean.</td><td>2021</td><td>One health (Amsterd</td><td>13</td><td>100316</td><td>RAYYAN-INCLUSION: {"Richard"=>"Excluded"} | RAYYAN-EXCLUSION-REASONS: wrong infection / disease</td><td>10.1016/j.onehlt.2021.100316</td><td>Humanities;Humanism;Humans;Caribbean Region</td><td>PMC8405964</td><td>https://pubmed.ncbi.nlm.nih.gov/34485673/</td></tr>
<tr><td>rayyan-751570457</td><td>Burt JF and Ouma J and Lubyayi L and Amone A and Aol L and Sekikubo M and Nakimuli A and Nakabembe E and Mboizi R and Musoke P and Kyohere M and Namara Lugolobi E and</td><td>Indirect effects of COVID-19 on maternal, neonatal, child, sexual and reproductive health services in Kampala, Uganda.</td><td>2021</td><td>BMJ global health</td><td>6</td><td></td><td>RAYYAN-INCLUSION: {"Richard"=>"Excluded"} | RAYYAN-EXCLUSION-REASONS: wrong infection / disease</td><td>10.1136/bmjgh-2021-006102</td><td>*COVID-19;Child;Communicable Disease</td><td>PMC8406460</td><td>https://pubmed.ncbi.nlm.nih.gov/34452941/</td></tr>
<tr><td>rayyan-751570458</td><td>Chulasiri P and Ranaweera P and Sudarshan P and Jayasinghe M and Harishchandra J and Gunasekera K and Vitharana H and Silva P and Ringwald P and Fernandopulle R and Mendis K and</td><td>Transfusion-induced Plasmodium falciparum malaria in a beta thalassaemia patient during the prevention of re-establishment phase in Sri Lanka.</td><td>2021</td><td>Malaria journal</td><td>20</td><td>352</td><td>RAYYAN-INCLUSION: {"Richard"=>"Excluded"} | RAYYAN-EXCLUSION-REASONS: wrong outcome,wrong study design</td><td>10.1186/s12936-021-03881-1</td><td>Adolescent;Blood/*parasitology;*Blood Transfusion;Humans;Malaria,</td><td>PMC8390059</td><td>https://pubmed.ncbi.nlm.nih.gov/34445999/</td></tr>
<tr><td>rayyan-751570460</td><td>Ansah F and Suurbaar J and Darko D and Anabire NG and Blankson SO and Domson BKS and Soulama A and Kpasra P and Chirawurah JD and Amenga-Etego L and Kanyong P and</td><td>Development of Cooperative Primer-Based Real-Time PCR Assays for the Detection of Plasmodium malariae and Plasmodium ovale.</td><td>2021</td><td>The Journal of molecular</td><td>23</td><td>1393-1403</td><td>RAYYAN-INCLUSION: {"Richard"=>"Excluded"} | RAYYAN-EXCLUSION-REASONS: wrong outcome,wrong study design</td><td>10.1016/j.jmoldx.2021.07.022</td><td>Adolescent;Adult;Child;Child, Preschool;Coinfection/*diagnosis/epidemiology/parasitology;</td><td>PMC8591562</td><td>https://pubmed.ncbi.nlm.nih.gov/34425259/</td></tr>
<tr><td>rayyan-751570461</td><td>Omale UI and Oka OU and Okeke IM and Azuogu BN and Alo C and Madubueze UC and Eze II and Okeke KC and Utulu R and Akpan UE and Iloke CV and Nnubia AO and Ibemesi DO and</td><td>Demand for malaria rapid diagnostic test, health care-seeking behaviour, and drug use among rural community members with fever or malaria-like illness in Ebonyi state, Nigeria: a cross-sectional household survey.</td><td>2021</td><td>BMC health services</td><td>21</td><td>857</td><td>RAYYAN-INCLUSION: {"Richard"=>"Excluded"} | RAYYAN-EXCLUSION-REASONS: wrong outcome,wrong study design</td><td>10.1186/s12913-021-06865-8</td><td>Adult;Child;Child, Preschool;Cross-Sectional Studies;*Diagnostic Tests,</td><td>PMC8380369</td><td>https://pubmed.ncbi.nlm.nih.gov/34419029/</td></tr>
<tr><td>rayyan-751570462</td><td>Li W and Zhang X and Feng J and Zhang T and Xu X and Jiang J and Wang S and Lyu X and Li S and Lu M</td><td>Evaluation of the combination of rapid diagnostic tests and microscopy for imported malaria surveillance in Anhui Province, China.</td><td>2021</td><td>Acta tropica</td><td>222</td><td>106042</td><td>RAYYAN-INCLUSION: {"Richard"=>"Excluded"} | RAYYAN-EXCLUSION-REASONS: wrong outcome,wrong study design</td><td>10.1016/j.actatropica.2021.106042</td><td>China/epidemiology;Diagnostic Tests, Routine;Humans;*Malaria/dia</td><td></td><td>https://pubmed.ncbi.nlm.nih.gov/34252385/</td></tr>
<tr><td>rayyan-751570463</td><td>Hergott DEB and Balkus JE and GarcÃ-a GA and Cruz KR and Seilie AM and Masters H and Aveika AA and Iyanga MM and Mba Eyono JN and Guthrie BL and Murphy SC</td><td>Clustering of subpatent infections in households with asymptomatic rapid diagnostic test-positive cases in Bioko Island, Equatorial Guinea independent of travel to regions of higher malaria endemicity: a cross-sectional study.</td><td>2021</td><td>Malaria journal</td><td>20</td><td>313</td><td>RAYYAN-INCLUSION: {"Richard"=>"Excluded"} | RAYYAN-EXCLUSION-REASONS: wrong outcome,wrong study design</td><td>10.1186/s12936-021-03844-6</td><td>Adolescent;Adult;Aged;Aged, 80 and over;Child;Child, Preschool;Cross-Sectional</td><td>PMC8274032</td><td>https://pubmed.ncbi.nlm.nih.gov/34247643/</td></tr>
<tr><td>rayyan-751570464</td><td>Marchal B and Abejirinde IO and Sulaberidze L and Chikovani I and Uchaneishvili M and Shengelia N and Diaconu K and Vassall A and Zoidze A and Giralt AN and Witter S</td><td>How do participatory methods shape policy? Applying a realist approach to the formulation of a new tuberculosis policy in Georgia.</td><td>2021</td><td>BMJ open</td><td>11</td><td>e047948</td><td>RAYYAN-INCLUSION: {"Richard"=>"Excluded"} | RAYYAN-EXCLUSION-REASONS: wrong publication type,wrong infection / disease</td><td>10.1136/bmjopen-2020-047948</td><td>Georgia;Georgia (Republic);Government Programs;Health</td><td>PMC8245474</td><td>https://pubmed.ncbi.nlm.nih.gov/34187826/</td></tr>
<tr><td>rayyan-751570466</td><td>Hoyt J and Hill J and Achieng F and Ouma P and Kariuki S and Desai M and Webster J</td><td>Healthcare provider and pregnant women's perspectives on the implementation of intermittent screening and treatment with dihydroartemisinin-piperaquine for malaria in pregnancy in western Kenya: a qualitative study.</td><td>2021</td><td>Malaria journal</td><td>20</td><td>291</td><td>RAYYAN-INCLUSION: {"Richard"=>"Excluded"} | RAYYAN-EXCLUSION-REASONS: wrong outcome,wrong study design | USER-NOTES: {"Richard"=>["Qual study"]}</td><td>10.1186/s12936-021-03826-8</td><td>Adolescent;Adult;Antimalarials/*therapeutic use;Artemisinins/therapeutic</td><td>PMC8243500</td><td>https://pubmed.ncbi.nlm.nih.gov/34187458/</td></tr>
<tr><td>rayyan-751570467</td><td>McLean ARD and Indrasuta C and Khant ZS and Phyo AK and Maung SM and Heaton J and Aung H and Aung Y and Soe K and Swe MMM and von Seidlein L and Tun NN and Tun KM</td><td>Mass drug administration for the acceleration of malaria elimination in a region of Myanmar with artemisinin-resistant falciparum malaria: a cluster-randomised trial.</td><td>2021</td><td>The Lancet. Infectious</td><td>21</td><td>1579-1589</td><td>RAYYAN-INCLUSION: {"Richard"=>"Excluded"} | RAYYAN-EXCLUSION-REASONS: wrong study design | USER-NOTES: {"Richard"=>["No routine data"]}</td><td>10.1016/S1473-3099(20)30997-X</td><td>Adolescent;Adult;Antimalarials/administration & dosage/therapeutic</td><td>PMC7614510</td><td>https://pubmed.ncbi.nlm.nih.gov/34147154/</td></tr>
<tr><td>rayyan-751570468</td><td>Andolina C and Rek JC and Briggs J and Okoth J and Musiime A and Ramjith J and Teyssier N and Conrad M and Nankabirwa JI and Lanke K and Rodriguez-Barraquer I and Meerstein-Kessel</td><td>Sources of persistent malaria transmission in a setting with effective malaria control in eastern Uganda: a longitudinal, observational cohort study.</td><td>2021</td><td>The Lancet. Infectious</td><td>21</td><td>1568-1578</td><td>RAYYAN-INCLUSION: {"Richard"=>"Excluded"} | RAYYAN-EXCLUSION-REASONS: wrong outcome,wrong study design | USER-NOTES: {"Richard"=>["No intervention"]}</td><td>10.1016/S1473-3099(21)00072-4</td><td>Adolescent;Animals;Anopheles/*parasitology;Antimalarials/therapeutic use;Artemether,</td><td>PMC8554388</td><td>https://pubmed.ncbi.nlm.nih.gov/34146476/</td></tr>
<tr><td>rayyan-751570472</td><td>Kande Betu Ku Mesu V and Mutombo Kalonji W and Bardonneau C and Valverde Mordt O and Ngolo Tete D and Blesson S and Simon F and Delhomme S and Bernhard S and</td><td>Oral fexinidazole for stage 1 or early stage 2 African Trypanosoma brucei gambiense trypanosomiasis: a prospective, multicentre, open-label, cohort study.</td><td>2021</td><td>The Lancet. Global</td><td>9</td><td>e999-e1008</td><td>RAYYAN-INCLUSION: {"Richard"=>"Excluded"} | RAYYAN-EXCLUSION-REASONS: wrong infection / disease</td><td>10.1016/S2214-109X(21)00208-4</td><td>Administration, Oral;Adolescent;Adult;Aged;Democratic Republic of the</td><td>PMC8220131</td><td>https://pubmed.ncbi.nlm.nih.gov/34143998/</td></tr>
<tr><td>rayyan-751570484</td><td>Tetteh M and Dwomoh D and Asamoah A and Kupeh EK and Malm K and Nonvignon J</td><td>Impact of malaria diagnostic refresher training programme on competencies and skills in malaria diagnosis among medical laboratory professionals: evidence from Ghana 2015-2019.</td><td>2021</td><td>Malaria journal</td><td>20</td><td>255</td><td>RAYYAN-INCLUSION: {"Richard"=>"Excluded"} | RAYYAN-EXCLUSION-REASONS: wrong outcome,wrong study design</td><td>10.1186/s12936-021-03796-x</td><td>Clinical Competence/*statistics & numerical data;Diagnostic</td><td>PMC8186098</td><td>https://pubmed.ncbi.nlm.nih.gov/34103068/</td></tr>
<tr><td>rayyan-751570487</td><td>Danwang C and Kirakoya-Samadoulougou F and Samadoulougou S</td><td>Assessing field performance of ultrasensitive rapid diagnostic tests for malaria: a systematic review and meta-analysis.</td><td>2021</td><td>Malaria journal</td><td>20</td><td>245</td><td>RAYYAN-INCLUSION: {"Richard"=>"Excluded"} | RAYYAN-EXCLUSION-REASONS: wrong outcome,wrong study design,wrong publication type | USER-NOTES: {"Richard"=>["Review"]}</td><td>10.1186/s12936-021-03783-2</td><td>Diagnostic Tests, Routine/*methods/statistics & numerical</td><td>PMC8176703</td><td>https://pubmed.ncbi.nlm.nih.gov/34082776/</td></tr>
<tr><td>rayyan-751570491</td><td>Danwang C and Noubiap JJ and Souopgui J and Gaudart J and Yombi JC and Robert A</td><td>Accuracy of malaria diagnostic tests performed on non-invasively collected samples: a systematic review and meta-analysis.</td><td>2021</td><td>BMJ global health</td><td>6</td><td></td><td>RAYYAN-INCLUSION: {"Richard"=>"Excluded"} | RAYYAN-EXCLUSION-REASONS: wrong outcome,wrong study design,wrong publication type | USER-NOTES: {"Richard"=>["Review"]}</td><td>10.1136/bmjgh-2021-005634</td><td>*Diagnostic Tests, Routine;Humans;*Malaria/diagnosis;Microscopy;Polymerase</td><td>PMC8173286</td><td>https://pubmed.ncbi.nlm.nih.gov/34078631/</td></tr>
<tr><td>rayyan-751570494</td><td>Devine A and Battle KE and Meagher N and Howes RE and Dini S and Gething PW and Simpson JA and Price RN and Lubell Y</td><td>Global economic costs due to vivax malaria and the potential impact of its radical cure: A modelling study.</td><td>2021</td><td>PLoS medicine</td><td>18</td><td>e1003614</td><td>RAYYAN-INCLUSION: {"Richard"=>"Excluded"} | RAYYAN-EXCLUSION-REASONS: wrong outcome,wrong study design | USER-NOTES: {"Richard"=>["Cost study"]}</td><td>10.1371/journal.pmed.1003614</td><td>Adolescent;Adult;Antimalarials/adverse effects/*economics/*therape</td><td>PMC8168905</td><td>https://pubmed.ncbi.nlm.nih.gov/34061843/</td></tr>
<tr><td>rayyan-751570500</td><td>Oboh MA and Oriero EC and Ndiaye T and Badiane AS and Ndiaye D and Amambua-Ngwa A</td><td>Comparative analysis of four malaria diagnostic tools and implications for malaria treatment in southwestern Nigeria.</td><td>2021</td><td>Internatio nal journal of</td><td>108</td><td>377-381</td><td>RAYYAN-INCLUSION: {"Richard"=>"Excluded"} | RAYYAN-EXCLUSION-REASONS: wrong outcome,wrong study design | USER-NOTES: {"Richard"=>["Diag study"]}</td><td>10.1016/j.ijid.2021.05.049</td><td>Cross-Sectional Studies;Diagnostic Tests, Routine;Humans;*Malaria/dia</td><td></td><td>https://pubmed.ncbi.nlm.nih.gov/34044139/</td></tr>
<tr><td>rayyan-751570505</td><td>Arsenault C and Yakob B and Tilahun T and Nigatu TG and Dinsa G and Woldie M and Kassa M and Berman P and Kruk ME</td><td>Patient volume and quality of primary care in Ethiopia: findings from the routine health information system and the 2014 Service Provision Assessment survey.</td><td>2021</td><td>BMC health services</td><td>21</td><td>485</td><td>RAYYAN-INCLUSION: {"Richard"=>"Maybe"} | RAYYAN-LABELS: Excluded (No intervention)</td><td>10.1186/s12913-021-06524-y</td><td>Child;Cross-Sectional Studies;Ethiopia;Female;*Health Information</td><td>PMC8140434</td><td>https://pubmed.ncbi.nlm.nih.gov/34022856/</td></tr>
<tr><td>rayyan-751570509</td><td>Nolder D and Stewart L and Tucker J and Ibrahim A and Gray A and Corrah T and Gallagher C and John L and O'Brien E and Aggarwal D and Benavente ED and van Schalkwyk D and</td><td>Failure of rapid diagnostic tests in Plasmodium falciparum malaria cases among travelers to the UK and Ireland: Identification and characterisation of the parasites.</td><td>2021</td><td>Internatio nal journal of</td><td>108</td><td>137-144</td><td>RAYYAN-INCLUSION: {"Richard"=>"Excluded"} | RAYYAN-EXCLUSION-REASONS: wrong outcome,wrong study design | USER-NOTES: {"Richard"=>["Diag study"]}</td><td>10.1016/j.ijid.2021.05.008</td><td>Animals;Antigens, Protozoan/genetics;Diagnostic Tests, Routine;Gene</td><td>PMC8295040</td><td>https://pubmed.ncbi.nlm.nih.gov/33991679/</td></tr>
</table>

| | | | | | | | | | | | |
|---|---|---|---|---|---|---|---|---|---|---|---|
| rayyan-751570511 | Castellanos ME and Rodas S and JuÃ¡rez JG and Lol JC and Chanquin S and Morales Z and Vizcaino L and Smith SC and Vanden Eng J and Woldu HG and Lenhart A and Padilla N | Evaluation of the durability of long-lasting insecticidal nets in Guatemala. | 2021 | Malaria journal | 20 | 219 | RAYYAN-INCLUSION: {"Richard"=>"Excluded"} \| RAYYAN-EXCLUSION-REASONS: wrong outcome,wrong study design \| USER-NOTES: {"Richard"=>["Ento / no epi indicator"]} | 10.1186/s12936-021-03722-1 | Cross-Sectional Studies;Guatemala;Insecticide-Treated Bednets/*statistics & | PMC8120849 | https://pubmed.ncbi.nlm.nih.gov/33990197/ |
| rayyan-751570579 | Engel N and Ghergu C and Matin MA and Kibria MG and Thriemer K and Price RN and Ding XC and Howes RE and Ley B and Incardona S and Alam MS | Implementing radical cure diagnostics for malaria: user perspectives on G6PD testing in Bangladesh. | 2021 | Malaria journal | 20 | 217 | RAYYAN-INCLUSION: {"Richard"=>"Excluded"} \| RAYYAN-EXCLUSION-REASONS: wrong outcome,wrong study design \| USER-NOTES: {"Richard"=>["Diag study"]} | 10.1186/s12936-021-03743-w | Bangladesh;Diagnostic Tests, Routine/psychology/*statistics & numerical | PMC8114691 | https://pubmed.ncbi.nlm.nih.gov/33980257/ |
| rayyan-751570580 | Kong A and Wilson SA and Ah Y and Nace D and Rogier E and Aidoo M | HRP2 and HRP3 cross-reactivity and implications for HRP2-based RDT use in regions with Plasmodium falciparum hrp2 gene deletions. | 2021 | Malaria journal | 20 | 207 | RAYYAN-INCLUSION: {"Richard"=>"Excluded"} \| RAYYAN-EXCLUSION-REASONS: wrong outcome,wrong study design \| USER-NOTES: {"Richard"=>["Diag study"]} | 10.1186/s12936-021-03739-6 | Antigens, Protozoan/*genetics;Cross Reactions;Diagnostic Tests, | PMC8086288 | https://pubmed.ncbi.nlm.nih.gov/33926477/ |
| rayyan-751570581 | | Baseline incidence of meningitis, malaria, mortality and other health outcomes in infants and young sub-Saharan African children prior to the introduction of the RTS,S/AS01(E) malaria vaccine. | 2021 | Malaria journal | 20 | 197 | RAYYAN-INCLUSION: {"Richard"=>"Excluded"} \| RAYYAN-EXCLUSION-REASONS: wrong study design \| USER-NOTES: {"Richard"=>["More of a protocol study"]} | 10.1186/s12936-021-03670-w | Female;Ghana/epidemiology;Humans;Incidence;Infant;Kenya/epidemiology;Malaria | PMC8073890 | https://pubmed.ncbi.nlm.nih.gov/33902599/ |
| rayyan-751570582 | Ngo TD and Canavati SE and Dung DV and Vo TH and Tran DT and Tran LK and Whedbee RJ and Milgotina EI and Kelly GC and Edgel KA and Martin NJ | Results from a malaria indicator survey highlight the importance of routine data capture in high-risk forest and farm transmission sites in Vietnam to tailor location-specific malaria elimination interventions. | 2021 | PloS one | 16 | e0250045 | RAYYAN-INCLUSION: {"Richard"=>"Excluded"} \| RAYYAN-EXCLUSION-REASONS: wrong outcome,wrong study design \| USER-NOTES: {"Richard"=>["MIS study; qual data as well"]} | 10.1371/journal.pone.0250045 | Adult;Cross-Sectional Studies;Environmental Biomarkers;Family | PMC8051764 | https://pubmed.ncbi.nlm.nih.gov/33861798/ |
| rayyan-751570583 | Lubinda J and Haque U and Bi Y and Shad MY and Keellings D and Hamainza B and Moore AJ | Climate change and the dynamics of age-related malaria incidence in Southern Africa. | 2021 | Environmental research | 197 | 111017 | RAYYAN-INCLUSION: {"Richard"=>"Included"} | 10.1016/j.envres.2021.111017 | Africa, Southern;Aged;Child;Child, Preschool;Climate | | https://pubmed.ncbi.nlm.nih.gov/33766570/ |
| rayyan-751570584 | Laurens MB and Mungwira RG and Nampota N and Nyirenda OM and Divala TH and Kanjala M and Mkandawire FA and Galileya LT and Nyangulu W and Mwinjiwa E and Downs M | Revisiting Co-trimoxazole Prophylaxis for African Adults in the Era of Antiretroviral Therapy: A Randomized Controlled Clinical Trial. | 2021 | Clinical infectious diseases : | 73 | 1058-1065 | RAYYAN-INCLUSION: {"Richard"=>"Excluded"} \| RAYYAN-EXCLUSION-REASONS: wrong outcome,wrong study design,wrong infection / disease | 10.1093/cid/ciab252 | Adult;Anti-Retroviral Agents/therapeutic use;CD4 Lymphocyte Count;*HIV | PMC8442771 | https://pubmed.ncbi.nlm.nih.gov/33744963/ |
| rayyan-751570585 | Hu J and Zhang C and Wang S and Chen X and Hu J and Liu X | Accuracy of rapid diagnostic test to diagnose malaria in children under 5 years of age, a meta-analysis. | 2021 | Diagnostic microbiol | 100 | 115351 | RAYYAN-INCLUSION: {"Richard"=>"Excluded"} \| RAYYAN-EXCLUSION-REASONS: wrong outcome,wrong study design,wrong publication type \| USER-NOTES: {"Richard"=>["Diag study"]} | 10.1016/j.diagmicrobio.2021.115351 | Child, Preschool;*Diagnostic Tests, Routine;Humans;Malaria, | | https://pubmed.ncbi.nlm.nih.gov/33743294/ |
| rayyan-751570586 | Downey J and McKenna AH and Mendin SF and Waters A and Dunbar N and Tehmeh LG and White EE and Siedner MJ and Panjabi R and Kraemer JD and Kenny A and Ly EJ and Bass J | Measuring Knowledge of Community Health Workers at the Last Mile in Liberia: Feasibility and Results of Clinical Vignette Assessments. | 2021 | Global health, science | 9 | S111-S121 | RAYYAN-INCLUSION: {"Richard"=>"Excluded"} \| RAYYAN-EXCLUSION-REASONS: wrong outcome,wrong study design | 10.9745/GHSP-D-20-00380 | Case Management;Child;*Community Health | PMC7971375 | https://pubmed.ncbi.nlm.nih.gov/33727324/ |
| rayyan-751570587 | Napier HG and Baird M and Wong E and Walwyn-Jones E and Garcia ME and Cartagena L and Mngadi N and Vanisaveth V and Sengsavath V and Vilay P and Thongpiou K and Visser T | Evaluating Vertical Malaria Community Health Worker Programs as Malaria Declines: Learning From Program Evaluations in Honduras and Lao PDR. | 2021 | Global health, science | 9 | S98-S110 | RAYYAN-INCLUSION: {"Richard"=>"Maybe"} \| RAYYAN-LABELS: Excluded (No intervention) \| USER-NOTES: {"Richard"=>["CHW maybe"]} | 10.9745/GHSP-D-20-00379 | *Community Health Workers;Honduras/epidemiology;Humans;Laos/epidemiology | PMC7971372 | https://pubmed.ncbi.nlm.nih.gov/33727323/ |
| rayyan-751570589 | Deen J and Mukaka M and von Seidlein L | What is the yield of malaria reactive case detection in the Greater Mekong Sub-region? A review of published data and meta-analysis. | 2021 | Malaria journal | 20 | 131 | RAYYAN-INCLUSION: {"Richard"=>"Excluded"} \| RAYYAN-EXCLUSION-REASONS: wrong study design,wrong publication type \| USER-NOTES: {"Richard"=>["Review / meta-analysis"]} | 10.1186/s12936-021-03667-5 | Asia, Southeastern;Diagnostic Tests, Routine/*statistics & numerical | PMC7934542 | https://pubmed.ncbi.nlm.nih.gov/33663484/ |
| rayyan-751570590 | Rabiu OR and Dada-Adegbola H and Falade CO and Arinola OG and Odaibo AB and Ademowo OG | Malaria, Helminth Infections and Clinical Status Among HIV-Infected Pregnant Women. | 2021 | International journal of | 10 | 81-87 | RAYYAN-INCLUSION: {"Richard"=>"Excluded"} \| RAYYAN-EXCLUSION-REASONS: wrong outcome,wrong study design | 10.21106/ijma.352 | HIV Infections;Malaria | PMC7905432 | https://pubmed.ncbi.nlm.nih.gov/33659096/ |
| rayyan-751570591 | Ayemoba O and Okeji N and Hussain N and Umar T and Ajemba-Life A and Kene T and Edom U and Ogueri I and Nwagbara G and Ochai I and Adekanye U and Onoh I | Reference intervals of common clinical biochemistry analytes in young Nigerian adults. | 2021 | PloS one | 16 | e0247672 | RAYYAN-INCLUSION: {"Richard"=>"Excluded"} \| RAYYAN-EXCLUSION-REASONS: wrong outcome,wrong study design | 10.1371/journal.pone.0247672 | Adolescent;Adult;Alanine Transaminase/blood;Alkaline Phosphatase/blood;Amylases/ | PMC7920356 | https://pubmed.ncbi.nlm.nih.gov/33647019/ |
| rayyan-751570592 | Ngatu NR and Muzembo BA and Choomplang N and Kanbara S and Wumba R and Ikeda M and Mbelambela EP and Muchanga SM and Suzuki T and Wada K and Al Mahfuz H and | Malaria rapid diagnostic test (HRP2/pLDH) positivity, incidence, care accessibility and impact of community WASH Action programme in DR Congo: mixed method study involving 625 households. | 2021 | Malaria journal | 20 | 117 | RAYYAN-INCLUSION: {"Richard"=>"Excluded"} \| RAYYAN-EXCLUSION-REASONS: wrong outcome,wrong study design \| USER-NOTES: {"Richard"=>["Diag study"]} | 10.1186/s12936-021-03647-9 | Adult;Communicable Disease Control/*statistics & numerical data;Community | PMC7913406 | https://pubmed.ncbi.nlm.nih.gov/33639932/ |
| rayyan-751570593 | Ahmad A and Soni P and Kumar L and Singh MP and Verma AK and Sharma A and Das A and Bharti PK | Comparison of polymerase chain reaction, microscopy, and rapid diagnostic test in malaria detection in a high burden state (Odisha) of India. | 2021 | Pathogens and global | 115 | 267-272 | RAYYAN-INCLUSION: {"Richard"=>"Excluded"} \| RAYYAN-EXCLUSION-REASONS: wrong outcome,wrong study design \| USER-NOTES: {"Richard"=>["Diag study"]} | 10.1080/20477724.2021.1893484 | Diagnostic Tests, Routine;Humans;India/epidemiology;*Malaria/diagnosis/epid | PMC8168770 | https://pubmed.ncbi.nlm.nih.gov/33634745/ |
| rayyan-751570594 | Kim JH and Suh J and Lee WJ and Choi H and Kim JD and Kim C and Choi JY and Ko R and Kim H and Lee J and Yeom JS | Modelling the impact of rapid diagnostic tests on Plasmodium vivax malaria in South Korea: a cost-benefit analysis. | 2021 | BMJ global health | 6 | | RAYYAN-INCLUSION: {"Richard"=>"Excluded"} \| RAYYAN-EXCLUSION-REASONS: wrong outcome,wrong study design \| USER-NOTES: {"Richard"=>["Diag / costing study"]} | 10.1136/bmjgh-2020-004292 | Cost-Benefit Analysis;Diagnostic Tests, Routine;Humans;*Malaria, | PMC7888375 | https://pubmed.ncbi.nlm.nih.gov/33593755/ |
| rayyan-751570595 | Tchum SK and Arthur FK and Adu B and Sakyi SA and Abubakar LA and Atibilla D and Amenga-Etego S and Oppong FB and Dzabeng F and Amoani B and Gyan T and Arhin E and | Impact of iron fortification on anaemia and iron deficiency among pre-school children living in Rural Ghana. | 2021 | PloS one | 16 | e0246362 | RAYYAN-INCLUSION: {"Richard"=>"Excluded"} \| RAYYAN-EXCLUSION-REASONS: wrong outcome,wrong study design \| USER-NOTES: {"Richard"=>["No routine data"]} | 10.1371/journal.pone.0246362 | Anemia, Iron-Deficiency/blood/*drug therapy/epidemiology;Ascorbi | PMC7877575 | https://pubmed.ncbi.nlm.nih.gov/33571267/ |
| rayyan-751570596 | Oliphant NP and Manda S and Daniels K and Odendaal WA and Besada D and Kinney M and White Johansson E and Doherty T | Integrated community case management of childhood illness in low- and middle-income countries. | 2021 | The Cochrane database | 2 | CD012882 | RAYYAN-INCLUSION: {"Richard"=>"Excluded"} \| RAYYAN-EXCLUSION-REASONS: wrong study design,wrong publication type \| USER-NOTES: {"Richard"=>["Review"]} | 10.1002/14651858.CD012882.pub2 | Africa South of the Sahara;Asia;Bias;Case Management/*organization & | PMC8094443 | https://pubmed.ncbi.nlm.nih.gov/33565123/ |
| rayyan-751570597 | Gunasekera WMKTAW and Premaratne R and Fernando D and Munaz M and Piyasena MGY and Perera D and Wickremasinghe R and Ranaweera KDNP and Mendis K | A comparative analysis of the outcome of malaria case surveillance strategies in Sri Lanka in the prevention of re-establishment phase. | 2021 | Malaria journal | 20 | 80 | RAYYAN-INCLUSION: {"Richard"=>"Included"} | 10.1186/s12936-021-03621-5 | *Epidemiological Monitoring;Humans;Malaria/*prevention & | PMC7871399 | https://pubmed.ncbi.nlm.nih.gov/33563273/ |
| rayyan-751570598 | Kaur J and Kaura T and Sharma A and Kumar A and Pangotra MK and Upadhyay AK and Anvikar A and Grover GS and Chaturvedi HK and Sharma SK | Surveillance-based estimation of the malaria disease burden in a low endemic state of Punjab, India, targeted for malaria elimination. | 2021 | Transactions of the Royal | 115 | 512-519 | RAYYAN-INCLUSION: {"Richard"=>"Excluded"} \| RAYYAN-EXCLUSION-REASONS: wrong outcome,wrong study design \| USER-NOTES: {"Richard"=>["Stratification"]} | 10.1093/trstmh/trab005 | *Cost of Illness;Humans;Incidence;India/epidemiology;*Malaria/epide | | https://pubmed.ncbi.nlm.nih.gov/33539517/ |
| rayyan-751570599 | Muthunatarajan S and Basavaiah SH and Shenoy SM and Natarajan A and Mithra P and Suresh PK and Sreeram S | Discriminant value of automated leucocyte VCS parameters in the detection of tropical infections. | 2021 | Journal of clinical laborator | 35 | e23723 | RAYYAN-INCLUSION: {"Richard"=>"Excluded"} \| RAYYAN-EXCLUSION-REASONS: wrong outcome,wrong study design | 10.1002/jcla.23723 | Adolescent;Adult;Aged;Aged, 80 and over;Automation;Female;*He | PMC8059744 | https://pubmed.ncbi.nlm.nih.gov/33538356/ |
| rayyan-751570600 | Reyes RA and Fornace KM and Macalinao MLM and Boncayao BL and De La Fuente ES and Sabanal HM and Bareng APN and Medado IAP and Mercado ES and Baquilod MS and Luchavez | Enhanced Health Facility Surveys to Support Malaria Control and Elimination across Different Transmission Settings in the Philippines. | 2021 | The American journal of | 104 | 968-978 | RAYYAN-INCLUSION: {"Richard"=>"Excluded"} \| RAYYAN-EXCLUSION-REASONS: wrong outcome,wrong study design \| USER-NOTES: {"Richard"=>["No routine data"]} | 10.4269/ajtmh.20-0814 | Adolescent;Adult;Aged;Aged, 80 and over;Child;Child, Preschool;Cross-Sectional | PMC7941801 | https://pubmed.ncbi.nlm.nih.gov/33534761/ |
| rayyan-751570601 | Koliopoulos P and Kayange NM and Daniel T and Huth F and GrÃ¶ndahl B and Medina-MontaÃ±o GC and Pretsch L and KlÃ¼ber J and Schmidt C and ZÃ¼chner A and Ulbert S and | Multiplex-RT-PCR-ELISA panel for detecting mosquito-borne pathogens: Plasmodium sp. preserved and eluted from dried blood spots on sample cards. | 2021 | Malaria journal | 20 | 66 | RAYYAN-INCLUSION: {"Richard"=>"Excluded"} \| RAYYAN-EXCLUSION-REASONS: wrong outcome,wrong study design \| USER-NOTES: {"Richard"=>["Ento study"]} | 10.1186/s12936-021-03595-4 | Child;Child, Preschool;Diagnostic Tests, Routine/*methods;Dried | PMC7851927 | https://pubmed.ncbi.nlm.nih.gov/33526038/ |
| rayyan-751570602 | Abuaku B and Amoah LE and Peprah NY and Asamoah A and Amoako EO and Donu D and Adu GA and Malm KL | Malaria parasitaemia and mRDT diagnostic performances among symptomatic individuals in selected health care facilities across Ghana. | 2021 | BMC public health | 21 | 239 | RAYYAN-INCLUSION: {"Richard"=>"Excluded"} \| RAYYAN-EXCLUSION-REASONS: wrong outcome,wrong study design \| USER-NOTES: {"Richard"=>["Diag study"]} | 10.1186/s12889-021-10290-1 | Cross-Sectional Studies;Delivery of Health Care;Diagnostic Tests, | PMC7844948 | https://pubmed.ncbi.nlm.nih.gov/33509161/ |
| rayyan-751570603 | Oriero EC and Amenga-Etego L and Ishengoma DS and Amambua-Ngwa A | Plasmodium malariae, current knowledge and future research opportunities on a neglected malaria parasite species. | 2021 | Critical reviews in microbiol | 47 | 44-56 | RAYYAN-INCLUSION: {"Richard"=>"Excluded"} \| RAYYAN-EXCLUSION-REASONS: wrong outcome,wrong study design,wrong publication type \| USER-NOTES: {"Richard"=>["Review\n"]} | 10.1080/1040841X.2020.1838440 | Africa/epidemiology;Animals;Antibodies, Protozoan/blood;Biomedical | | https://pubmed.ncbi.nlm.nih.gov/33507842/ |
| rayyan-751570604 | Mischlinger J and Dudek V and Ramharter M | Predictive Performance of Rapid Diagnostic Tests for Falciparum Malaria and Its Modeled Impact on Integrated Community Case Management of Malaria in Sub-Saharan African Febrile Children. | 2021 | Clinical infectious diseases : | 73 | e1158-e1167 | RAYYAN-INCLUSION: {"Richard"=>"Excluded"} \| RAYYAN-EXCLUSION-REASONS: wrong outcome,wrong study design | 10.1093/cid/ciaa1942 | Antigens, Protozoan;Case Management;Child;Diagnostic Tests, | PMC8423504 | https://pubmed.ncbi.nlm.nih.gov/33506270/ |
| rayyan-751570605 | Drews SJ and Spencer BR and Wendel S and Bloch EM | Filariasis and transfusion-associated risk: a literature review. | 2021 | Vox sanguinis | 116 | 741-754 | RAYYAN-INCLUSION: {"Richard"=>"Excluded"} \| RAYYAN-EXCLUSION-REASONS: wrong infection / disease | 10.1111/vox.13073 | Animals;Blood Transfusion;*Filariasis/epidemiology/prevention & | | https://pubmed.ncbi.nlm.nih.gov/33491765/ |
| rayyan-751570606 | Manjurano A and Omolo JJ and Lyimo E and Miyaye D and Kishamawe C and Matemba LE and Massaga JJ and Changalucha J and Kazyoba PE | Performance evaluation of the highly sensitive histidine-rich protein 2 rapid test for plasmodium falciparum malaria in North-West Tanzania. | 2021 | Malaria journal | 20 | 58 | RAYYAN-INCLUSION: {"Richard"=>"Excluded"} \| RAYYAN-EXCLUSION-REASONS: wrong outcome,wrong study design \| USER-NOTES: {"Richard"=>["Diag study"]} | 10.1186/s12936-020-03568-z | Adolescent;Adult;Antigens, Protozoan/*genetics;Child;Cross-Sectional | PMC7821515 | https://pubmed.ncbi.nlm.nih.gov/33482835/ |
| rayyan-751570607 | Mekachie Sandie S and Sumbele IUN and Tasah MM and Kimbi HK | Malaria and intestinal parasite co-infection and its association with anaemia among people living with HIV in Buea, Southwest Cameroon: A community-based retrospective cohort study. | 2021 | PloS one | 16 | e0245743 | RAYYAN-INCLUSION: {"Richard"=>"Excluded"} \| RAYYAN-EXCLUSION-REASONS: wrong outcome,wrong study design | 10.1371/journal.pone.0245743 | Adolescent;Aged;Anemia/*complications;Cameroon/epidemiology;Child;Child, | PMC7822292 | https://pubmed.ncbi.nlm.nih.gov/33481933/ |
| rayyan-751570608 | Balami AD and Said SM and Zulkefli NAM and Norsa'adah B and Audu B | Improving malaria preventive practices and pregnancy outcomes through a health education intervention: A randomized controlled trial. | 2021 | Malaria journal | 20 | 55 | RAYYAN-INCLUSION: {"Richard"=>"Excluded"} \| RAYYAN-EXCLUSION-REASONS: wrong outcome,wrong study design | 10.1186/s12936-021-03586-5 | Adolescent;Adult;Female;Health Behavior;Health Education/*methods;Humans; | PMC7818731 | https://pubmed.ncbi.nlm.nih.gov/33478529/ |
| rayyan-751570610 | Nabet C and Kone AK and Dia AK and Sylla M and Gautier M and Yattara M and Thera MA and Faye O and Braack L and Manguin S and Beavogui AH and Doumbo O and Gay F and | New assessment of Anopheles vector species identification using MALDI-TOF MS. | 2021 | Malaria journal | 20 | 33 | RAYYAN-INCLUSION: {"Richard"=>"Excluded"} \| RAYYAN-EXCLUSION-REASONS: wrong outcome,wrong study design \| USER-NOTES: {"Richard"=>["Ento study"]} | 10.1186/s12936-020-03557-2 | Animals;Anopheles/*classification;Female;Guinea;Malaria/transmission;Male;Mali;Mosquito | PMC7796594 | https://pubmed.ncbi.nlm.nih.gov/33422056/ |
| rayyan-751570611 | Penjor K and Tobgyal and Zangpo T and Clements ACA and Gray DJ and Wangdi K | Has COVID19 derailed Bhutan's national malaria elimination goal? A commentary. | 2021 | Malaria journal | 20 | 20 | RAYYAN-INCLUSION: {"Richard"=>"Maybe"} \| RAYYAN-LABELS: Excluded (C19) \| USER-NOTES: {"Richard"=>["COVID19"]} | 10.1186/s12936-020-03562-5 | Bhutan/epidemiology;COVID-19/*epidemiology/virology;Goals;Humans;Malaria/*preventi | PMC7787406 | https://pubmed.ncbi.nlm.nih.gov/33407471/ |

| | | | | | | | | | | | |
|---|---|---|---|---|---|---|---|---|---|---|---|
| rayyan-751570612 | Raman J and Barnes KI and Baker L and Blaylock M and Blumberg L and Frean J and Misiani E and Ukpe IS | Maintaining focus on administering effective malaria treatment during the COVID-19 pandemic. | 2020 | South African medical | 111 | 13-16 | RAYYAN-INCLUSION: {"Richard"=>"Excluded"} \| RAYYAN-EXCLUSION-REASONS: wrong outcome,wrong study design | 10.7196/SAMJ.2020.v111i11.15289 | Administration, Intravenous;Antigens, Protozoan/blood;Antimalarials | | https://pubmed.ncbi.nlm.nih.gov/33403998/ |
| rayyan-751570613 | Kotepui M and Masangkay FR and Kotepui KU and De Jesus Milanez G | Misidentification of Plasmodium ovale as Plasmodium vivax malaria by a microscopic method: a meta-analysis of confirmed P. ovale cases. | 2020 | Scientific reports | 10 | 21807 | RAYYAN-INCLUSION: {"Richard"=>"Excluded"} \| RAYYAN-EXCLUSION-REASONS: wrong outcome,wrong study design \| USER-NOTES: {"Richard"=>["Diag study\n"]} | 10.1038/s41598-020-78691-7 | *Diagnostic Errors;Humans;Malaria, Vivax/*diagnosis;*Microscopy; | PMC7733466 | https://pubmed.ncbi.nlm.nih.gov/33311528/ |
| rayyan-751570614 | | Effectiveness of seasonal malaria chemoprevention at scale in west and central Africa: an observational study. | 2020 | Lancet (London, England) | 396 | 1829-1840 | RAYYAN-INCLUSION: {"Richard"=>"Included"} \| RAYYAN-LABELS: Include (full text) | 10.1016/S0140-6736(20)32227-3 | Adolescent;Adult;Africa, Central/epidemiology;Africa, Western/epidemiology;Amodi | PMC7718580 | https://pubmed.ncbi.nlm.nih.gov/33278936/ |
| rayyan-751570615 | Kitojo C and Chacky F and Kigadye ES and Mugasa JP and Lusasi A and Mohamed A and Walker P and Reaves EJ and Gutman JR and Ishengoma DS | Evaluation of a single screen and treat strategy to detect asymptomatic malaria among pregnant women from selected health facilities in Lindi region, Tanzania. | 2020 | Malaria journal | 19 | 438 | RAYYAN-INCLUSION: {"Richard"=>"Maybe"} \| RAYYAN-LABELS: Include (full text) | 10.1186/s12936-020-03513-0 | Antimalarials/*therapeutic use;Diagnostic Tests, Routine/*statistics & | PMC7708125 | https://pubmed.ncbi.nlm.nih.gov/33256758/ |
| rayyan-751570617 | Dharap P and Raimbault S | Performance evaluation of machine learning-based infectious screening flags on the HORIBA Medical Yumizen H550 Haematology Analyzer for vivax malaria and dengue fever. | 2020 | Malaria journal | 19 | 429 | RAYYAN-INCLUSION: {"Richard"=>"Excluded"} \| RAYYAN-EXCLUSION-REASONS: wrong outcome,wrong study design \| USER-NOTES: {"Richard"=>["Diag study"]} | 10.1186/s12936-020-03502-3 | Adolescent;Adult;Aged;Aged, 80 and over;Child;Child, Preschool;Dengue/*diagnosis; | PMC7684750 | https://pubmed.ncbi.nlm.nih.gov/33228680/ |
| rayyan-751570618 | Odhiambo JN and Sartorius B | Mapping of anaemia prevalence among pregnant women in Kenya (2016-2019). | 2020 | BMC pregnancy and | 20 | 711 | RAYYAN-INCLUSION: {"Richard"=>"Excluded"} \| RAYYAN-EXCLUSION-REASONS: wrong outcome,wrong study design \| USER-NOTES: {"Richard"=>["Strat"]} | 10.1186/s12884-020-03380-2 | Anemia/*epidemiology;Bayes Theorem;Cost of Illness;Female;Humans;Kenya/ | PMC7685542 | https://pubmed.ncbi.nlm.nih.gov/33228585/ |
| rayyan-751570619 | Zango SH and Lingani M and Valea I and Samadoulougou OS and Bihoun B and Rouamba T and Derra K and Rouamba E and Donnen P and Dramaix M and Tinto H and Robert A | Malaria and curable sexually transmitted infections in pregnant women: A two-years observational study in rural Burkina Faso. | 2020 | PloS one | 15 | e0242368 | RAYYAN-INCLUSION: {"Richard"=>"Excluded"} \| RAYYAN-EXCLUSION-REASONS: wrong outcome,wrong study design | 10.1371/journal.pone.0242368 | Adult;Antimalarials/therapeutic use;Burkina Faso/epidemiology;Coinfectio | PMC7668607 | https://pubmed.ncbi.nlm.nih.gov/33196665/ |
| rayyan-751570620 | Restrepo-Posada DC and Carmona-Fonseca J and Cardona-Arias JA | Cost-effectiveness of rapid diagnostic tests, compared to microscopic tests, for the diagnosis and treatment of gestational malaria in Colombia from an institutional perspective. | 2020 | Malaria journal | 19 | 400 | RAYYAN-INCLUSION: {"Richard"=>"Excluded"} \| RAYYAN-EXCLUSION-REASONS: wrong outcome,wrong study design \| USER-NOTES: {"Richard"=>["Diag costing study"]} | 10.1186/s12936-020-03472-6 | Adolescent;Adult;Child;Child, Preschool;Colombia;Cost-Benefit Analysis/*statistics & | PMC7653853 | https://pubmed.ncbi.nlm.nih.gov/33172472/ |
| rayyan-751570621 | Gatton ML and Chaudhry A and Glenn J and Wilson S and Ah Y and Kong A and Ord RL and Rees-Channer RR and Chiodini P and Incardona S and Cheng Q and Aidoo M and Cunningham J | Impact of Plasmodium falciparum gene deletions on malaria rapid diagnostic test performance. | 2020 | Malaria journal | 19 | 392 | RAYYAN-INCLUSION: {"Richard"=>"Excluded"} \| RAYYAN-EXCLUSION-REASONS: wrong outcome,wrong study design \| USER-NOTES: {"Richard"=>["Genomic surveillance"]} | 10.1186/s12936-020-03460-w | Antigens, Protozoan/*genetics;Diagnostic Tests, Routine/*statistics & | PMC7640408 | https://pubmed.ncbi.nlm.nih.gov/33148265/ |
| rayyan-751570622 | Bakari C and Jones S and Subramaniam G and Mandara CI and Chiduo MG and Rumisha S and Chacky F and Molteni F and Mandike R and Mkude S and Njau R and Herman C and Nace | Community-based surveys for Plasmodium falciparum pfhrp2 and pfhrp3 gene deletions in selected regions of mainland Tanzania. | 2020 | Malaria journal | 19 | 391 | RAYYAN-INCLUSION: {"Richard"=>"Excluded"} \| RAYYAN-EXCLUSION-REASONS: wrong outcome,wrong study design \| USER-NOTES: {"Richard"=>["Diag / genomic surveilance"]} | 10.1186/s12936-020-03459-3 | Adolescent;Adult;Antigens, Protozoan/*genetics;Child;Child, Preschool;Cross-Sectional | PMC7640459 | https://pubmed.ncbi.nlm.nih.gov/33148255/ |
| rayyan-751570623 | Utzinger KS and Held U and Hanscheid T and GÃ¼ltekin N and Fehr J and Grobusch MP and Schlagenhauf P | Self-diagnosis and self-treatment of Plasmodium spp. infection by travellers (1989-2019): A systematic review and meta-analysis. | 2020 | Travel medicine and | 38 | 101902 | RAYYAN-INCLUSION: {"Richard"=>"Excluded"} \| RAYYAN-EXCLUSION-REASONS: wrong outcome,wrong study design,wrong publication type \| USER-NOTES: {"Richard"=>["Review"]} | 10.1016/j.tmaid.2020.101902 | Antimalarials/*therapeutic use;Diagnostic Tests, Routine/methods;Emergency | | https://pubmed.ncbi.nlm.nih.gov/33132136/ |
| rayyan-751570624 | Ojo AA and Maxwell K and Oresanya O and Adaji J and Hamade P and Tibenderana JK and Abubakar SS and Audu BM and Njidda A and Gubio AB and Snow RW and Zurovac D | Health systems readiness and quality of inpatient malaria case-management in Kano State, Nigeria. | 2020 | Malaria journal | 19 | 384 | RAYYAN-INCLUSION: {"Richard"=>"Excluded"} \| RAYYAN-EXCLUSION-REASONS: wrong outcome,wrong study design | 10.1186/s12936-020-03449-5 | Antimalarials/*therapeutic use;*Case Management;Child;Child, | PMC7602350 | https://pubmed.ncbi.nlm.nih.gov/33126886/ |
| rayyan-751570625 | Stresman G and Whittaker C and Slater HC and Bousema T and Cook J | Quantifying Plasmodium falciparum infections clustering within households to inform household-based intervention strategies for malaria control programs: An observational study and meta-analysis from 41 malaria-endemic countries. | 2020 | PLoS medicine | 17 | e1003370 | RAYYAN-INCLUSION: {"Richard"=>"Excluded"} \| RAYYAN-EXCLUSION-REASONS: wrong publication type \| USER-NOTES: {"Richard"=>["Review"]} | 10.1371/journal.pmed.1003370 | Africa/epidemiology;Child;Child, Preschool;Cluster Analysis;Cross-Sectional | PMC7595326 | https://pubmed.ncbi.nlm.nih.gov/33119589/ |
| rayyan-751570626 | Nice J and Nahusenay H and Eckert E and Eisele TP and Ashton RA | Estimating malaria chemoprevention and vector control coverage using program and campaign data: A scoping review of current practices and opportunities. | 2020 | Journal of global health | 10 | 020413 | RAYYAN-INCLUSION: {"Richard"=>"Excluded"} \| RAYYAN-EXCLUSION-REASONS: wrong publication type \| USER-NOTES: {"Richard"=>["Review"]} | 10.7189/jogh.10.020413 | Chemoprevention;Cross-Sectional Studies;Female;Humans;*Inse | PMC7568932 | https://pubmed.ncbi.nlm.nih.gov/33110575/ |
| rayyan-751570627 | Opoku MM and Bonful HA and Koram KA | Antibiotic prescription for febrile outpatients: a health facility-based secondary data analysis for the Greater Accra region of Ghana. | 2020 | BMC health services | 20 | 978 | RAYYAN-INCLUSION: {"Richard"=>"Excluded"} \| RAYYAN-EXCLUSION-REASONS: wrong outcome,wrong study design,wrong infection / disease | 10.1186/s12913-020-05771-9 | Adolescent;Adult;*Ambulatory Care Facilities;Anti-Bacterial Agents/*therapeutic | PMC7590657 | https://pubmed.ncbi.nlm.nih.gov/33109158/ |
| rayyan-751570628 | Rathmes G and Rumisha SF and Lucas TCD and Twohig KA and Python A and Nguyen M and Nandi AK and Keddie SH and Collins EL and Rozier JA and Gibson HS and Chestnutt EG and | Global estimation of anti-malarial drug effectiveness for the treatment of uncomplicated Plasmodium falciparum malaria 1991-2019. | 2020 | Malaria journal | 19 | 374 | RAYYAN-INCLUSION: {"Richard"=>"Excluded"} \| RAYYAN-EXCLUSION-REASONS: wrong outcome,wrong study design,wrong publication type \| USER-NOTES: {"Richard"=>["Review Rx"]} | 10.1186/s12936-020-03446-8 | Antimalarials/*therapeutic use;Artemisinins/*therapeutic use;*Drug | PMC7573874 | https://pubmed.ncbi.nlm.nih.gov/33081784/ |
| rayyan-751570730 | Toya Y and Tougan T and Horii T and Uchihashi K | Lysercell M enhances the detection of stage-specific Plasmodium-infected red blood cells in the automated hematology analyzer XN-31 prototype. | 2021 | Parasitology internatio | 80 | 102206 | RAYYAN-INCLUSION: {"Richard"=>"Excluded"} \| RAYYAN-EXCLUSION-REASONS: wrong outcome,wrong study design \| USER-NOTES: {"Richard"=>["Diag study"]} | 10.1016/j.parint.2020.102206 | Automation, Laboratory;Diagnostic Tests, Routine/instrumentation/*me | | https://pubmed.ncbi.nlm.nih.gov/33049417/ |
| rayyan-751570732 | Nfor NO and Senyuy DT | Malaria among febrile neonates attending the neonatology unit of the Bamenda regional hospital. | 2020 | Parasite epidemiology and | 11 | e00184 | RAYYAN-INCLUSION: {"Richard"=>"Excluded"} \| RAYYAN-EXCLUSION-REASONS: wrong outcome,wrong study design \| USER-NOTES: {"Richard"=>["Epi study"]} | 10.1016/j.parepi.2020.e00184 | Infant, Newborn;Malaria | PMC7527715 | https://pubmed.ncbi.nlm.nih.gov/33024846/ |
| rayyan-751570733 | Rouamba T and Samadoulougou S and Kirakoya-Samadoulougou F | Addressing challenges in routine health data reporting in Burkina Faso through Bayesian spatiotemporal prediction of weekly clinical malaria incidence. | 2020 | Scientific reports | 10 | 16568 | RAYYAN-INCLUSION: {"Richard"=>"Excluded"} \| RAYYAN-EXCLUSION-REASONS: wrong outcome,wrong study design \| USER-NOTES: {"Richard"=>["Strat / modeling"]} | 10.1038/s41598-020-73601-3 | Bayes Theorem;Burkina Faso/epidemiology;Communicable Disease | PMC7538437 | https://pubmed.ncbi.nlm.nih.gov/33024162/ |
| rayyan-751570734 | Zhong D and Hemming-Schroeder E and Wang X and Kibret S and Zhou G and Atieli H and Lee MC and Afrane YA and Githeko AK and Yan G | Extensive new Anopheles cryptic species involved in human malaria transmission in western Kenya. | 2020 | Scientific reports | 10 | 16139 | RAYYAN-INCLUSION: {"Richard"=>"Excluded"} \| RAYYAN-EXCLUSION-REASONS: wrong outcome,wrong study design \| USER-NOTES: {"Richard"=>["Ento study"]} | 10.1038/s41598-020-73073-5 | Animals;Anopheles/*genetics;Demography;Insect Vectors/classification;Kenya/e | PMC7527330 | https://pubmed.ncbi.nlm.nih.gov/32999365/ |
| rayyan-751570735 | Du YQ and Ling XX and Jin JJ and Zhou HY and Zhu S and Zhu GD and Wang W and Cao J and Huang JY | Cost-effectiveness analysis of malaria rapid diagnostic test in the elimination setting. | 2020 | Infectious diseases of | 9 | 135 | RAYYAN-INCLUSION: {"Richard"=>"Excluded"} \| RAYYAN-EXCLUSION-REASONS: wrong outcome,wrong study design,wrong infection / disease \| USER-NOTES: {"Richard"=>["Diag / costing study"]} | 10.1186/s40249-020-00745-9 | Cost-Benefit Analysis;Decision Making;Diagnostic Tests, Routine/*economics;Evidence- | PMC7523355 | https://pubmed.ncbi.nlm.nih.gov/32993762/ |
| rayyan-751570736 | Weiss DJ and Bertozzi-Villa A and Rumisha SF and Amratia P and Arambepola R and Battle KE and Cameron E and Chestnutt E and Gibson HS and Harris J and Keddie S and | Indirect effects of the COVID-19 pandemic on malaria intervention coverage, morbidity, and mortality in Africa: a geospatial modelling analysis. | 2021 | The Lancet. Infectious | 21 | 59-69 | RAYYAN-INCLUSION: {"Richard"=>"Maybe"} \| RAYYAN-LABELS: Excluded (C19) | 10.1016/S1473-3099(20)30700-3 | Africa/epidemiology;Antimalarials/therapeutic use;Bayes Theorem;COVID- | PMC7505634 | https://pubmed.ncbi.nlm.nih.gov/32971006/ |
| rayyan-751570737 | Wagman J and CissÃ© I and Kone D and Fomba S and Eckert E and Mihigo J and Bankineza E and Bah M and Diallo D and Gogue C and Tynuv K and Saibu A and Richardson JH and | Rapid reduction of malaria transmission following the introduction of indoor residual spraying in previously unsprayed districts: an observational analysis of Mopti Region, Mali, in 2017. | 2020 | Malaria journal | 19 | 340 | RAYYAN-INCLUSION: {"Richard"=>"Included"} | 10.1186/s12936-020-03414-2 | Communicable Disease Control/*statistics & numerical data;Disease | PMC7501620 | https://pubmed.ncbi.nlm.nih.gov/32950056/ |
| rayyan-751570738 | Semakula M and Niragire FI and Faes C | Bayesian spatio-temporal modeling of malaria risk in Rwanda. | 2020 | PloS one | 15 | e0238504 | RAYYAN-INCLUSION: {"Richard"=>"Excluded"} \| RAYYAN-EXCLUSION-REASONS: wrong outcome,wrong study design \| USER-NOTES: {"Richard"=>["Strat"]} | 10.1371/journal.pone.0238504 | Bayes Theorem;Health Policy;Humans;Incidence;Malaria/*epidemiology;Probability; | PMC7482939 | https://pubmed.ncbi.nlm.nih.gov/32911503/ |
| rayyan-751570739 | Gachugia J and Chebore W and Otieno K and Ngugi CW and Godana A and Kariuki S | Evaluation of the colorimetric malachite green loop-mediated isothermal amplification (MG-LAMP) assay for the detection of malaria species at two different health facilities in a malaria endemic area of western Kenya. | 2020 | Malaria journal | 19 | 329 | RAYYAN-INCLUSION: {"Richard"=>"Excluded"} \| RAYYAN-EXCLUSION-REASONS: wrong outcome,wrong study design \| USER-NOTES: {"Richard"=>["Diag study"]} | 10.1186/s12936-020-03397-0 | Colorimetry/*methods;Diagnostic Tests, Routine/*methods;Humans;Ke | PMC7487890 | https://pubmed.ncbi.nlm.nih.gov/32907582/ |
| rayyan-751570741 | Tadesse G and Kamaliddin C and Doolan C and Amarasekara R and Legese R and Mohon AN and Cheaveau J and Yewhalaw D and Pillai DR | Active case detection of malaria in pregnancy using loop-mediated amplification (LAMP): a pilot outcomes study in South West Ethiopia. | 2020 | Malaria journal | 19 | 305 | RAYYAN-INCLUSION: {"Richard"=>"Excluded"} \| RAYYAN-EXCLUSION-REASONS: wrong outcome,wrong study design \| USER-NOTES: {"Richard"=>["Diag study"]} | 10.1186/s12936-020-03380-9 | Adult;Diagnostic Tests, Routine;Ethiopia/epidemiology;Female;Humans;Longitudinal | PMC7457308 | https://pubmed.ncbi.nlm.nih.gov/32854715/ |
| rayyan-751570742 | Haanshuus CG and MÃ¸rch K | Detection of remaining Plasmodium DNA and gametocytes during follow up after curative malaria treatment among returned travellers in Norway. | 2020 | Malaria journal | 19 | 296 | RAYYAN-INCLUSION: {"Richard"=>"Excluded"} \| RAYYAN-EXCLUSION-REASONS: wrong outcome,wrong study design \| USER-NOTES: {"Richard"=>["Diag / Rx study"]} | 10.1186/s12936-020-03367-6 | Communicable Diseases, Imported/*prevention & control;DNA, | PMC7436973 | https://pubmed.ncbi.nlm.nih.gov/32814587/ |
| rayyan-751570743 | Babalola OJ and Ajumobi O and Ajayi IO | Rural-urban disparities and factors associated with delayed care-seeking and testing for malaria before medication use by mothers of under-five children, Igabi LGA, Kaduna Nigeria. | 2020 | Malaria journal | 19 | 294 | RAYYAN-INCLUSION: {"Richard"=>"Excluded"} \| RAYYAN-EXCLUSION-REASONS: wrong outcome,wrong study design \| USER-NOTES: {"Richard"=>["Epi study"]} | 10.1186/s12936-020-03371-w | Adolescent;Adult;Antimalarials/*therapeutic use;Child, Preschool;Cross-Sectional | PMC7436948 | https://pubmed.ncbi.nlm.nih.gov/32811529/ |
| rayyan-751570744 | Kakuru A and Jagannathan P and Kajubi R and Ochieng T and Ochokoru H and Nakalembe M and Clark TD and Ruel T and Staedke SG and Chandramohan D and Havlir DV and Kamya | Impact of intermittent preventive treatment of malaria in pregnancy with dihydroartemisinin-piperaquine versus sulfadoxine-pyrimethamine on the incidence of malaria in infancy: a randomized controlled trial. | 2020 | BMC medicine | 18 | 207 | RAYYAN-INCLUSION: {"Richard"=>"Maybe"} \| RAYYAN-LABELS: Excluded (after full text) | 10.1186/s12916-020-01675-x | Adult;Antimalarials/pharmacology/*therapeutic use;Artesunate/pharmacology | PMC7416391 | https://pubmed.ncbi.nlm.nih.gov/32772921/ |
| rayyan-751570745 | Varo R and Balanza N and Mayor A and Bassat Q | Diagnosis of clinical malaria in endemic settings. | 2021 | Expert review of anti- | 19 | 79-92 | RAYYAN-INCLUSION: {"Richard"=>"Excluded"} \| RAYYAN-EXCLUSION-REASONS: wrong outcome,wrong study design,wrong publication type | 10.1080/14787210.2020.1807940 | Child;Diagnostic Tests, Routine/*methods;Humans;Malaria/*diagnosis/epidemiolog | | https://pubmed.ncbi.nlm.nih.gov/32772759/ |
| rayyan-751570746 | Gontie GB and Wolde HF and Baraki AG | Prevalence and associated factors of malaria among pregnant women in Sherkole district, Benishangul Gumuz regional state, West Ethiopia. | 2020 | BMC infectious diseases | 20 | 573 | RAYYAN-INCLUSION: {"Richard"=>"Excluded"} \| RAYYAN-EXCLUSION-REASONS: wrong outcome,wrong study design | 10.1186/s12879-020-05289-9 | Adolescent;Adult;Antigens, Protozoan/blood;Asymptomatic Infections;Cross-Sectional | PMC7405459 | https://pubmed.ncbi.nlm.nih.gov/32758164/ |
| rayyan-751570747 | Rocha D and de Melo GC and Carneiro JMH and Ribeiro M and Ribeiro S and de Godoy DT and Costa E and de Almeida ACG and de Andrade EF and Abrahim CMM and Fraiji NA and | Use of a NAT-based assay to improve the surveillance system and prevent transfusion-transmitted malaria in blood banks. | 2020 | Malaria journal | 19 | 275 | RAYYAN-INCLUSION: {"Richard"=>"Excluded"} \| RAYYAN-EXCLUSION-REASONS: wrong outcome,wrong study design | 10.1186/s12936-020-03345-y | Adolescent;Adult;Blood Banks;Blood Transfusion;Brazil;Epidemiolog | PMC7395345 | https://pubmed.ncbi.nlm.nih.gov/32736625/ |

| | | | | | | | | | | | |
|---|---|---|---|---|---|---|---|---|---|---|---|
| rayyan-751570748 | Fraser M and Miller JM and Silumbe K and Hainsworth M and Mudenda M and Hamainza B and Moonga H and Chizema Kawesha E and Mercer LD and Bennett A and Schneider K and | Evaluating the Impact of Programmatic Mass Drug Administration for Malaria in Zambia Using Routine Incidence Data. | 2022 | The Journal of infectious | 225 | 1415-1423 | RAYYAN-INCLUSION: {"Richard"=>"Included"} | 10.1093/infdis/jiaa434 | *Antimalarials/therapeutic use;Humans;Incidence;*Malaria/drug | PMC9016426 | https://pubmed.ncbi.nlm.nih.gov/32691047/ |
| rayyan-751570749 | Conner RO and Dieye Y and Hainsworth M and Tall A and CissÃ© B and Faye F and Sy MD and Ba A and Sene D and Ba S and DoucourÃ© E and Thiam T and Diop M and Schneider K | Mass testing and treatment for malaria followed by weekly fever screening, testing and treatment in Northern Senegal: feasibility, cost and impact. | 2020 | Malaria journal | 19 | 252 | RAYYAN-INCLUSION: {"Richard"=>"Maybe"} \| RAYYAN-LABELS: Include (full text) | 10.1186/s12936-020-03313-6 | Adolescent;Adult;Aged;Antimalarials/*therapeutic use;Artemisinins/*therapeutic | PMC7362450 | https://pubmed.ncbi.nlm.nih.gov/32664939/ |
| rayyan-751570750 | Picot S and Cucherat M and Bienvenu AL | Systematic review and meta-analysis of diagnostic accuracy of loop-mediated isothermal amplification (LAMP) methods compared with microscopy, polymerase chain reaction and rapid diagnostic tests for malaria diagnosis. | 2020 | International journal of | 98 | 408-419 | RAYYAN-INCLUSION: {"Richard"=>"Excluded"} \| RAYYAN-EXCLUSION-REASONS: wrong outcome,wrong study design,wrong publication type \| USER-NOTES: {"Richard"=>["Review"]} | 10.1016/j.ijid.2020.07.009 | Diagnostic Tests, Routine/methods/*standards; Humans;Malaria/*diagnosis/p | | https://pubmed.ncbi.nlm.nih.gov/32659450/ |
| rayyan-751570751 | Gogue C and Wagman J and Tynuv K and Saibu A and Yihdego Y and Malm K and Mohamed W and Akplu W and Tagoe T and Ofosu A and Williams I and Asiedu S and Richardson J and | An observational analysis of the impact of indoor residual spraying in Northern, Upper East, and Upper West Regions of Ghana: 2014 through 2017. | 2020 | Malaria journal | 19 | 242 | RAYYAN-INCLUSION: {"Richard"=>"Included"} | 10.1186/s12936-020-03318-1 | Ghana/epidemiology;Humans;Incidence;Insecticides/*administration & | PMC7353711 | https://pubmed.ncbi.nlm.nih.gov/32652994/ |
| rayyan-751570752 | Liang D and Jin JJ and Wang WM and Cao YY and Zhu GD and Zhou HY and Cao J and Huang JY | Evaluating the implementation of rapid diagnostic tests in a malaria elimination setting. | 2020 | Infectious diseases of | 9 | 84 | RAYYAN-INCLUSION: {"Richard"=>"Excluded"} \| RAYYAN-EXCLUSION-REASONS: wrong outcome,wrong study design \| USER-NOTES: {"Richard"=>["Diag study"]} | 10.1186/s40249-020-00702-6 | China;Communicable Diseases, Imported/*diagnosis;Diagnostic Tests, Routine/*statistics & | PMC7346650 | https://pubmed.ncbi.nlm.nih.gov/32641123/ |
| rayyan-751570753 | Cirera L and Galatas B and Alonso S and Paaijmans K and Mamuquele M and MartÃ--Soler H and Guinovart C and Munguambe H and Luis F and Nhantumbo H and MontaÃ±Ã J | Moving towards malaria elimination in southern Mozambique: Cost and cost-effectiveness of mass drug administration combined with intensified malaria control. | 2020 | PloS one | 15 | e0235631 | RAYYAN-INCLUSION: {"Richard"=>"Excluded"} \| RAYYAN-EXCLUSION-REASONS: wrong outcome,wrong study design \| USER-NOTES: {"Richard"=>["Costing study"]} | 10.1371/journal.pone.0235631 | *Cost-Benefit Analysis;Humans;Malaria/*economics/*prevention & | PMC7337313 | https://pubmed.ncbi.nlm.nih.gov/32628741/ |
| rayyan-751570754 | Kumar R and Farzeen M and Hafeez A and Achakzai BK and Vankwani M and Lal M and Iqbal R and Somrongthong R | Effectiveness of a health education intervention on the use of long-lasting insecticidal nets for the prevention of malaria in pregnant women of Pakistan: a quasi-experimental study. | 2020 | Malaria journal | 19 | 232 | RAYYAN-INCLUSION: {"Richard"=>"Excluded"} \| RAYYAN-EXCLUSION-REASONS: wrong outcome,wrong study design | 10.1186/s12936-020-03298-2 | Adult;Female;Health Education/*statistics & numerical data;*Health | PMC7325083 | https://pubmed.ncbi.nlm.nih.gov/32600347/ |
| rayyan-751570755 | Tran EE and Cheeks ML and Kakuru A and Muhindo MK and Natureeba P and Nakalembe M and Ategeka J and Nayebare P and Kamya M and Havlir D and Feeney ME and Dorsey G and | The impact of gravidity, symptomatology and timing of infection on placental malaria. | 2020 | Malaria journal | 19 | 227 | RAYYAN-INCLUSION: {"Richard"=>"Excluded"} \| RAYYAN-EXCLUSION-REASONS: wrong outcome,wrong study design | 10.1186/s12936-020-03297-3 | Adolescent;Adult;Double-Blind Method;Female;Gravidity;Humans;Malaria, | PMC7315526 | https://pubmed.ncbi.nlm.nih.gov/32580739/ |
| rayyan-751570757 | Selvarajah D and Naing C and Htet NH and Mak JW | Loop-mediated isothermal amplification (LAMP) test for diagnosis of uncomplicated malaria in endemic areas: a meta-analysis of diagnostic test accuracy. | 2020 | Malaria journal | 19 | 211 | RAYYAN-INCLUSION: {"Richard"=>"Excluded"} \| RAYYAN-EXCLUSION-REASONS: wrong outcome,wrong study design,wrong publication type \| USER-NOTES: {"Richard"=>["Diag study"]} | 10.1186/s12936-020-03283-9 | Diagnostic Tests, Routine/*methods;Malaria/*diagnosis;Molecular Diagnostic | PMC7305603 | https://pubmed.ncbi.nlm.nih.gov/32560728/ |
| rayyan-751570758 | Millar J and Toh KB and Valle D | To screen or not to screen: an interactive framework for comparing costs of mass malaria treatment interventions. | 2020 | BMC medicine | 18 | 149 | RAYYAN-INCLUSION: {"Richard"=>"Excluded"} \| RAYYAN-EXCLUSION-REASONS: wrong outcome,wrong study design,wrong publication type | 10.1186/s12916-020-01609-7 | Cost-Benefit Analysis/*methods;Diagnostic Tests, | PMC7304173 | https://pubmed.ncbi.nlm.nih.gov/32552743/ |
| rayyan-751570759 | Khan AQ and Pernaute-Lau L and Khattak AA and Luijcx S and Aydin-Schmidt B and Hussain M and Khan TA and Mufti FU and Morris U | Surveillance of genetic markers associated with Plasmodium falciparum resistance to artemisinin-based combination therapy in Pakistan, 2018-2019. | 2020 | Malaria journal | 19 | 206 | RAYYAN-INCLUSION: {"Richard"=>"Excluded"} \| RAYYAN-EXCLUSION-REASONS: wrong outcome,wrong study design \| USER-NOTES: {"Richard"=>["Diag / genomic surveillance study"]} | 10.1186/s12936-020-03276-8 | Antimalarials/*therapeutic use;Artemether, Lumefantrine Drug | PMC7282094 | https://pubmed.ncbi.nlm.nih.gov/32513171/ |
| rayyan-751570760 | Diengou NH and Cumber SN and Nkfusai CN and Mbinyui MS and Viyoff VZ and Bede F and Akwah L and Tsoka-Gwegweni JM and Judith AK | Factors associated with the uptake of intermittent preventive treatment of malaria in pregnancy in the Bamenda health districts, Cameroon. | 2020 | The Pan African medical | 35 | 42 | RAYYAN-INCLUSION: {"Richard"=>"Excluded"} \| RAYYAN-EXCLUSION-REASONS: wrong outcome,wrong study design | 10.11604/pamj.2020.35.42.17600 | Adolescent;Adult;Cameroon/epidemiology;Cross-Sectional Studies;Directly Observed | PMC7245972 | https://pubmed.ncbi.nlm.nih.gov/32499857/ |
| rayyan-751570761 | Briand V and Cottrell G and Tuike Ndam N and MartiÃ¡Ã±ez-Vendrell X and Vianou B and Mama A and Kouwaye B and HouzÃ© S and Bailly J and Gbaguidi E and Sossou D and | Prevalence and clinical impact of malaria infections detected with a highly sensitive HRP2 rapid diagnostic test in Beninese pregnant women. | 2020 | Malaria journal | 19 | 188 | RAYYAN-INCLUSION: {"Richard"=>"Excluded"} \| RAYYAN-EXCLUSION-REASONS: wrong outcome,wrong study design \| USER-NOTES: {"Richard"=>["Diag study"]} | 10.1186/s12936-020-03261-1 | Adult;Antigens, Protozoan/*analysis;Diagnostic Tests, Routine/*statistics & | PMC7247134 | https://pubmed.ncbi.nlm.nih.gov/32448310/ |
| rayyan-751570762 | Bath D and Goodman C and Yeung S | Modelling the cost-effectiveness of introducing subsidised malaria rapid diagnostic tests in the private retail sector in sub-Saharan Africa. | 2020 | BMJ global health | 5 | | RAYYAN-INCLUSION: {"Richard"=>"Excluded"} \| RAYYAN-EXCLUSION-REASONS: wrong outcome,wrong study design \| USER-NOTES: {"Richard"=>["Costing study"]} | 10.1136/bmjgh-2019-002138 | Africa South of the Sahara;Cost-Benefit Analysis;*Diagnostic Tests, | PMC7247415 | https://pubmed.ncbi.nlm.nih.gov/32439690/ |
| rayyan-751570763 | Palmer T and Aiyenigba AO and Bates I and Okyere DD and Tagbor H and Ampofo GD | Improving the effectiveness of point of care tests for malaria and anaemia: a qualitative study across three Ghanaian antenatal clinics. | 2020 | BMC health services | 20 | 444 | RAYYAN-INCLUSION: {"Richard"=>"Excluded"} \| RAYYAN-EXCLUSION-REASONS: wrong outcome,wrong study design \| USER-NOTES: {"Richard"=>["Qual studt"]} | 10.1186/s12913-020-05274-7 | Adolescent;Adult;Ambulatory Care Facilities;Anemia/*diagnosis;Di | PMC7238731 | https://pubmed.ncbi.nlm.nih.gov/32429903/ |
| rayyan-751570764 | Brabin L and Roberts SA and Tinto H and Gies S and Diallo S and Brabin B | Iron Status of BurkinabÃ© Adolescent Girls Predicts Malaria Risk in the Following Rainy Season. | 2020 | Nutrients | 12 | | RAYYAN-INCLUSION: {"Richard"=>"Excluded"} \| RAYYAN-EXCLUSION-REASONS: wrong outcome,wrong study design | 10.3390/nu12051446 | Adolescent;Burkina Faso;Double-Blind Method;Female;Humans;Iron/ | PMC7284973 | https://pubmed.ncbi.nlm.nih.gov/32429481/ |
| rayyan-751570765 | Magnus D and Bhatta S and Mytton J and Joshi E and Bird EL and Bhatta S and Manandhar SR and Joshi SK | Establishing injury surveillance in emergency departments in Nepal: protocol for mixed methods prospective study. | 2020 | BMC health services | 20 | 433 | RAYYAN-INCLUSION: {"Richard"=>"Excluded"} \| RAYYAN-EXCLUSION-REASONS: wrong outcome,wrong study design,wrong infection / disease | 10.1186/s12913-020-05280-9 | Adolescent;Adult;Child;Child, Preschool;Emergency Service, Hospital/*statistics & | PMC7236178 | https://pubmed.ncbi.nlm.nih.gov/32423459/ |
| rayyan-751570766 | Addisu A and Tegegne Y and Mihiret Y and Setegn A and Zeleke AJ | A 7-Year Trend of Malaria at Primary Health Facilities in Northwest Ethiopia. | 2020 | Journal of parasitology | 2020 | 4204987 | RAYYAN-INCLUSION: {"Richard"=>"Excluded"} \| RAYYAN-EXCLUSION-REASONS: wrong outcome,wrong study design | 10.1155/2020/4204987 | Northwestern United States;Health Facilities;Malaria | PMC7204144 | https://pubmed.ncbi.nlm.nih.gov/32411421/ |
| rayyan-751570767 | Thawer SG and Chacky F and Runge M and Reaves E and Mandike R and Lazaro S and Mkude S and Rumisha SF and Kumalija C and Lengeler C and Mohamed A and Pothin E and | Sub-national stratification of malaria risk in mainland Tanzania: a simplified assembly of survey and routine data. | 2020 | Malaria journal | 19 | 177 | RAYYAN-INCLUSION: {"Richard"=>"Excluded"} \| RAYYAN-EXCLUSION-REASONS: wrong outcome,wrong study design \| USER-NOTES: {"Richard"=>["Strat study"]} | 10.1186/s12936-020-03250-4 | Adolescent;Adult;Aged;Aged, 80 and over;Child;Child, Preschool;Female;Humans;Inf | PMC7206674 | https://pubmed.ncbi.nlm.nih.gov/32384923/ |
| rayyan-751570768 | Zhu W and Ling X and Shang W and Du Y and Liu J and Cao Y and Yang M and Zhu G and Cao J and Huang J | High value of rapid diagnostic tests to diagnose malaria within children: A systematic review and meta-analysis. | 2020 | Journal of global health | 10 | 010411 | RAYYAN-INCLUSION: {"Richard"=>"Excluded"} \| RAYYAN-EXCLUSION-REASONS: wrong outcome,wrong study design,wrong publication type \| USER-NOTES: {"Richard"=>["Review"]} | 10.7189/jogh.10.010411 | Child, Preschool;China;*Diagnostic Tests, | PMC7182354 | https://pubmed.ncbi.nlm.nih.gov/32373330/ |
| rayyan-751570769 | Gaye S and Kibler J and Ndiaye JL and Diouf MB and Linn A and Gueye AB and Fall FB and Ndiop M and Diallo I and Cisse M and Ba M and Thwing J | Proactive community case management in Senegal 2014-2016: a case study in maximizing the impact of community case management of malaria. | 2020 | Malaria journal | 19 | 166 | RAYYAN-INCLUSION: {"Richard"=>"Maybe"} \| RAYYAN-LABELS: Excluded (after full text) | 10.1186/s12936-020-03238-0 | Adolescent;Adult;Aged;Aged, 80 and over;Case Management/*statistics & | PMC7183580 | https://pubmed.ncbi.nlm.nih.gov/32334581/ |
| rayyan-751570770 | Tilahun A and Yimer M and Gelaye W and Tegegne B | Prevalence of asymptomatic Plasmodium species infection and associated factors among pregnant women attending antenatal care at Fendeka town health facilities, Jawi District, North west Ethiopia: A cross-sectional study. | 2020 | PloS one | 15 | e0231477 | RAYYAN-INCLUSION: {"Richard"=>"Excluded"} \| RAYYAN-EXCLUSION-REASONS: wrong outcome,wrong study design | 10.1371/journal.pone.0231477 | Adult;Asymptomatic Infections/*epidemiology;Cross-Sectional Studies;Diagnostic | PMC7173768 | https://pubmed.ncbi.nlm.nih.gov/32315341/ |
| rayyan-751570771 | Lek D and Callery JJ and Nguon C and Debackere M and Sovannaroth S and Tripura R and Wojnarski M and Piola P and Khean ST and Manion K and Nguon S and Kunkel A and | Tools to accelerate falciparum malaria elimination in Cambodia: a meeting report. | 2020 | Malaria journal | 19 | 151 | RAYYAN-INCLUSION: {"Richard"=>"Excluded"} \| RAYYAN-EXCLUSION-REASONS: wrong outcome,wrong study design,wrong publication type | 10.1186/s12936-020-03197-6 | Antimalarials/therapeutic use;Cambodia;*Diagnostic Tests, Routine;Disease | PMC7161105 | https://pubmed.ncbi.nlm.nih.gov/32293452/ |
| rayyan-751570772 | Tangena JA and Hendriks CMJ and Devine M and Tammaro M and Trett AE and Williams I and DePina AJ and Sisay A and Herizo R and Kafy HT and Chizema E and Were A and Rozier J | Indoor residual spraying for malaria control in sub-Saharan Africa 1997 to 2017: an adjusted retrospective analysis. | 2020 | Malaria journal | 19 | 150 | RAYYAN-INCLUSION: {"Richard"=>"Excluded"} \| RAYYAN-EXCLUSION-REASONS: wrong outcome,wrong study design | 10.1186/s12936-020-03216-6 | Africa South of the Sahara;Communicable Disease Control/*statistics & numerical | PMC7149868 | https://pubmed.ncbi.nlm.nih.gov/32276585/ |
| rayyan-751570773 | Kigozi SP and Giorgi E and Mpimbaza A and Kigozi RN and Bousema T and Arinaitwe E and Nankabirwa JI and Sebuguzi CM and Kamya MR and Staedke SG and Dorsey G and Pullan | Practical Implications of a Relationship between Health Management Information System and Community Cohort-Based Malaria Incidence Rates. | 2020 | The American journal of | 103 | 404-414 | RAYYAN-INCLUSION: {"Richard"=>"Excluded"} \| RAYYAN-EXCLUSION-REASONS: wrong outcome,wrong study design | 10.4269/ajtmh.19-0950 | Ambulatory Care;Child;Child, Preschool;Cohort Studies;*Communicable | PMC7356469 | https://pubmed.ncbi.nlm.nih.gov/32274990/ |
| rayyan-751570774 | Sahu M and Tediosi F and Noor AM and Aponte JJ and Fink G | Health systems and global progress towards malaria elimination, 2000-2016. | 2020 | Malaria journal | 19 | 141 | RAYYAN-INCLUSION: {"Richard"=>"Excluded"} \| RAYYAN-EXCLUSION-REASONS: wrong outcome,wrong study design | 10.1186/s12936-020-03208-6 | Disease Eradication/*statistics & numerical data;Global Health/*statistics & numerical | PMC7140365 | https://pubmed.ncbi.nlm.nih.gov/32268917/ |
| rayyan-751570775 | Nakakana UN and Mohammed IA and Onankpa BO and Jega RM and Jiya NM | A validation of the Malaria Atlas Project maps and development of a new map of malaria transmission in Sokoto, Nigeria: a cross-sectional study using geographic information systems. | 2020 | Malaria journal | 19 | 149 | RAYYAN-INCLUSION: {"Richard"=>"Excluded"} \| RAYYAN-EXCLUSION-REASONS: wrong outcome,wrong study design | 10.1186/s12936-020-03214-8 | Child;Child, Preschool;Cross-Sectional Studies;Female;*Geographic | PMC7140379 | https://pubmed.ncbi.nlm.nih.gov/32268904/ |
| rayyan-751570776 | KonatÃ© D and Diawara SI and TourÃ© M and DiakitÃ© SAS and Guindo A and TraorÃ© K and Diarra A and Keita B and Thiam S and Keita M and Sissoko I and Sogoba N and TraorÃ© | Effect of routine seasonal malaria chemoprevention on malaria trends in children under 5Â years in Dangassa, Mali. | 2020 | Malaria journal | 19 | 137 | RAYYAN-INCLUSION: {"Richard"=>"Maybe"} \| RAYYAN-LABELS: Excluded (after full text) | 10.1186/s12936-020-03202-y | Antimalarials/*administration & dosage;Chemoprevention/*sta | PMC7137428 | https://pubmed.ncbi.nlm.nih.gov/32252774/ |
| rayyan-751570777 | Cunningham JA and Thomson RM and Murphy SC and de la Paz Ade M and Ding XC and Incardona S and Legrand E and Lucchi NW and Menard D and Nsobya SL and Saez AC and | WHO malaria nucleic acid amplification test external quality assessment scheme: results of distribution programmes one to three. | 2020 | Malaria journal | 19 | 129 | RAYYAN-INCLUSION: {"Richard"=>"Excluded"} \| RAYYAN-EXCLUSION-REASONS: wrong outcome,wrong study design | 10.1186/s12936-020-03200-0 | Diagnostic Tests, Routine/*statistics & numerical | PMC7106789 | https://pubmed.ncbi.nlm.nih.gov/32228615/ |
| rayyan-751570778 | Guiguemde KT and Dieye Y and LÃ´ AC and Ndiaye M and Lam A and Manga IA and Sow GD and Diop M and SouanÃ© T and Diouf MP and Tine RCK and Faye B | Molecular detection and quantification of Plasmodium falciparum gametocytes carriage in used RDTs in malaria elimination settings in northern Senegal. | 2020 | Malaria journal | 19 | 123 | RAYYAN-INCLUSION: {"Richard"=>"Excluded"} \| RAYYAN-EXCLUSION-REASONS: wrong outcome,wrong study design | 10.1186/s12936-020-03204-w | DNA, Protozoan/*isolation & purification;*Diagnostic Tests, Routine;Epidemiological | PMC7106854 | https://pubmed.ncbi.nlm.nih.gov/32228599/ |
| rayyan-751570851 | Kigozi SP and Kigozi RN and Epstein A and Mpimbaza A and Sserwanga A and Yeka A and Nankabirwa JI and Halliday K and Pullan RL and Rutazaana D and Sebuguzi CM and Opigo J and | Rapid shifts in the age-specific burden of malaria following successful control interventions in four regions of Uganda. | 2020 | Malaria journal | 19 | 128 | RAYYAN-INCLUSION: {"Richard"=>"Maybe"} \| RAYYAN-LABELS: Include (full text) | 10.1186/s12936-020-03196-7 | Adolescent;Adult;Age Distribution;Age Factors;Aged;Child;Child, | PMC7106889 | https://pubmed.ncbi.nlm.nih.gov/32228584/ |
| rayyan-751570852 | Gmanyami JM and Ameko A and Ahiafe SS and Bosoka SA and Kweku M and Ansah EK | Effect of pre-consultation testing on clinicians' adherence to malaria test results and waiting time among children under 5Â years in the Northern Zone of Volta Region of Ghana. | 2020 | Malaria journal | 19 | 120 | RAYYAN-INCLUSION: {"Richard"=>"Excluded"} \| RAYYAN-EXCLUSION-REASONS: wrong outcome,wrong study design | 10.1186/s12936-020-03189-6 | Antimalarials/administration & dosage;Child, Preschool;*Delivery of Health | PMC7085133 | https://pubmed.ncbi.nlm.nih.gov/32197616/ |

| | | | | | | | | | | | |
|---|---|---|---|---|---|---|---|---|---|---|---|
| rayyan-751570853 | L'Episcopia M and Kelley J and Patel D and Schmedes S and Ravishankar S and Menegon M and Perrotti E and Nurahmed AM and Talha AA and Nour BY and Lucchi N and Severini C | Targeted deep amplicon sequencing of kelch 13 and cytochrome b in Plasmodium falciparum isolates from an endemic African country using the Malaria Resistance Surveillance (MaRS) protocol. | 2020 | Parasites & vectors | 13 | 137 | RAYYAN-INCLUSION: {"Richard"=>"Excluded"} \| RAYYAN-EXCLUSION-REASONS: wrong outcome,wrong study design | 10.1186/s13071-020-4005-7 | Africa;Anti-Infective Agents/pharmacology;Antimalarials/pharmacology;Artemisin | PMC7071742 | https://pubmed.ncbi.nlm.nih.gov/32171330/ |
| rayyan-751570854 | Corder RM and Ferreira MU and Gomes MGM | Modelling the epidemiology of residual Plasmodium vivax malaria in a heterogeneous host population: A case study in the Amazon Basin. | 2020 | PLoS computational | 16 | e1007377 | RAYYAN-INCLUSION: {"Richard"=>"Excluded"} \| RAYYAN-EXCLUSION-REASONS: wrong outcome,wrong study design | 10.1371/journal.pcbi.1007377 | Brazil/epidemiology;Computer Simulation;Female;Humans;Incidence;Malaria/*epidemiolog | PMC7108741 | https://pubmed.ncbi.nlm.nih.gov/32168349/ |
| rayyan-751570855 | Mayengue PI and Kouhounina Batsimba D and Niama RF and Ibara Ottia R and Malonga-Massanga A and Fila-Fila GPU and Ahombo G and Kobawila SC and Parra HJ | Variation of prevalence of malaria, parasite density and the multiplicity of Plasmodium falciparum infection throughout the year at three different health centers in Brazzaville, Republic of Congo. | 2020 | BMC infectious diseases | 20 | 190 | RAYYAN-INCLUSION: {"Richard"=>"Excluded"} \| RAYYAN-EXCLUSION-REASONS: wrong outcome,wrong study design | 10.1186/s12879-020-4913-3 | Animals;Antigens, Protozoan/genetics;Child;Child , | PMC7057455 | https://pubmed.ncbi.nlm.nih.gov/32131754/ |
| rayyan-751570856 | Maffioli EM and Mohanan M and Saran I and O'Meara WP | Does improving appropriate use of malaria medicines change population beliefs in testing and treatment? Evidence from a randomized controlled trial. | 2020 | Health policy and | 35 | 556-566 | RAYYAN-INCLUSION: {"Richard"=>"Excluded"} \| RAYYAN-EXCLUSION-REASONS: wrong outcome,wrong study design | 10.1093/heapol/czaa010 | Antimalarials/*therapeutic use;Artemether, Lumefantrine Drug | | https://pubmed.ncbi.nlm.nih.gov/32129851/ |
| rayyan-751570857 | Lover AA and Dantzer E and Hocini S and Estera R and Rerolle F and Smith JL and Hwang J and Gosling R and Yukich J and Greenhouse B and Jacobson J and Phetsouvanh R and | Study protocol for a cluster-randomized split-plot design trial to assess the effectiveness of targeted active malaria case detection among high-risk populations in Southern Lao PDR (the AcME-Lao study). | 2019 | Gates open research | 3 | 1730 | RAYYAN-INCLUSION: {"Richard"=>"Excluded"} \| RAYYAN-EXCLUSION-REASONS: wrong publication type \| USER-NOTES: {"Richard"=>["Protocol"]} | 10.12688/gatesopenres.13088.1 | Malaria | PMC7019195 | https://pubmed.ncbi.nlm.nih.gov/32118199/ |
| rayyan-751570858 | Jegede A and Willey B and Hamade P and Oshiname F and Chandramohan D and Ajayi I and Falade C and Baba E and Webster J | Evaluation of a capacity building intervention on malaria treatment for under-fives in rural health facilities in Niger State, Nigeria. | 2020 | Malaria journal | 19 | 90 | RAYYAN-INCLUSION: {"Richard"=>"Excluded"} \| RAYYAN-EXCLUSION-REASONS: wrong outcome,wrong study design | 10.1186/s12936-020-03167-y | Antimalarials/*therapeutic use;Capacity Building/*statistics & | PMC7041190 | https://pubmed.ncbi.nlm.nih.gov/32093679/ |
| rayyan-751570859 | Dini S and Douglas NM and Poespoprodjo JR and Kenangalem E and Sugiarto P and Plumb ID and Price RN and Simpson JA | The risk of morbidity and mortality following recurrent malaria in Papua, Indonesia: a retrospective cohort study. | 2020 | BMC medicine | 18 | 28 | RAYYAN-INCLUSION: {"Richard"=>"Excluded"} \| RAYYAN-EXCLUSION-REASONS: wrong outcome,wrong study design | 10.1186/s12916-020-1497-0 | Adolescent;Adult;Child;Female;Humans;Indonesia;Malaria/*epidemiology/*mortality;Male; | PMC7031957 | https://pubmed.ncbi.nlm.nih.gov/32075649/ |
| rayyan-751570861 | Rouamba T and Samadoulougou S and Tinto H and Alegana VA and Kirakoya-Samadoulougou F | Bayesian Spatiotemporal Modeling of Routinely Collected Data to Assess the Effect of Health Programs in Malaria Incidence During Pregnancy in Burkina Faso. | 2020 | Scientific reports | 10 | 2618 | RAYYAN-INCLUSION: {"Richard"=>"Included"} | 10.1038/s41598-020-58899-3 | Adult;*Bayes Theorem;Burkina Faso/epidemiology;Female;Health | PMC7021681 | https://pubmed.ncbi.nlm.nih.gov/32060297/ |
| rayyan-751570862 | Krampa FD and Aniweh Y and Kanyong P and Awandare GA | Recent Advances in the Development of Biosensors for Malaria Diagnosis. | 2020 | Sensors (Basel, Switzerla | 20 | | RAYYAN-INCLUSION: {"Richard"=>"Excluded"} \| RAYYAN-EXCLUSION-REASONS: wrong outcome,wrong study design | 10.3390/s20030799 | Antigens, Protozoan/immunology/isolation & purification;*Biosensing | PMC7038750 | https://pubmed.ncbi.nlm.nih.gov/32024098/ |
| rayyan-751570863 | Hawaria D and Demissew A and Kibret S and Lee MC and Yewhalaw D and Yan G | Effects of environmental modification on the diversity and positivity of anopheline mosquito aquatic habitats at Arjo-Dedessa irrigation development site, Southwest Ethiopia. | 2020 | Infectious diseases of | 9 | 9 | RAYYAN-INCLUSION: {"Richard"=>"Excluded"} \| RAYYAN-EXCLUSION-REASONS: wrong outcome,wrong study design | 10.1186/s40249-019-0620-y | *Agricultural Irrigation;*Animal Distribution;Animals;Anophele | PMC6986026 | https://pubmed.ncbi.nlm.nih.gov/31987056/ |
| rayyan-751570864 | Surendra H and Supargiyono and Ahmad RA and Kusumasari RA and Rahayujati TB and Damayanti SY and Tetteh KKA and Chitnis C and Stresman G and Cook J and Drakeley C | Using health facility-based serological surveillance to predict receptive areas at risk of malaria outbreaks in elimination areas. | 2020 | BMC medicine | 18 | 9 | RAYYAN-INCLUSION: {"Richard"=>"Excluded"} \| RAYYAN-EXCLUSION-REASONS: wrong outcome,wrong study design | 10.1186/s12916-019-1482-7 | Adult;Cluster Analysis;Cross-Sectional Studies;Diagnostic Tests, Routine;*Disease | PMC6986103 | https://pubmed.ncbi.nlm.nih.gov/31987052/ |
| rayyan-751570865 | Mbonye AK and Buregyeya E and Rutebemberwa E and Lal S and Clarke SE and Hansen KS and Magnussen P and LaRussa P | Treatment of Sick Children Seeking Care in the Private Health Sector in Uganda: A Cluster Randomized Trial. | 2020 | The American journal of | 102 | 658-666 | RAYYAN-INCLUSION: {"Richard"=>"Excluded"} \| RAYYAN-EXCLUSION-REASONS: wrong outcome,wrong study design | 10.4269/ajtmh.19-0367 | *Child Health;Child, Preschool;Community Health Services/organization & | PMC7056412 | https://pubmed.ncbi.nlm.nih.gov/31971139/ |
| rayyan-751570866 | OuÃ©draogo M and Rouamba T and Samadoulougou S and Kirakoya-Samadoulougou F | Effect of Free Healthcare Policy for Children under Five Years Old on the Incidence of Reported Malaria Cases in Burkina Faso by Bayesian Modelling: "Not only the Ears but also the Head of the Hippopotamus". | 2020 | International journal of | 17 | | RAYYAN-INCLUSION: {"Richard"=>"Maybe"} \| RAYYAN-LABELS: Excluded (after full text) \| RAYYAN-EXCLUSION-REASONS: wrong outcome | 10.3390/ijerph17020417 | Animals;Bayes Theorem;Burkina Faso/epidemiology;Child, | PMC7014427 | https://pubmed.ncbi.nlm.nih.gov/31936308/ |
| rayyan-751570867 | Jones S and Plucinski M and Kay K and Hodel EM and Hastings IM | A Computer Modelling Approach To Evaluate the Accuracy of Microsatellite Markers for Classification of Recurrent Infections during Routine Monitoring of Antimalarial Drug Efficacy. | 2020 | Antimicrobial agents | 64 | | RAYYAN-INCLUSION: {"Richard"=>"Excluded"} \| RAYYAN-EXCLUSION-REASONS: wrong outcome,wrong study design | 10.1128/AAC.01517-19 | Algorithms;Antimalarials/*therapeutic use;Artemether, Lumefantrine Drug | PMC7179279 | https://pubmed.ncbi.nlm.nih.gov/31932376/ |
| rayyan-751570868 | Challenger JD and GonÃ§alves BP and Bradley J and Bruxvoort K and Tiono AB and Drakeley C and Bousema T and Ghani AC and Okell LC | How delayed and non-adherent treatment contribute to onward transmission of malaria: a modelling study. | 2019 | BMJ global health | 4 | e001856 | RAYYAN-INCLUSION: {"Richard"=>"Excluded"} \| RAYYAN-EXCLUSION-REASONS: wrong outcome,wrong study design | 10.1136/bmjgh-2019-001856 | Malaria | PMC6936434 | https://pubmed.ncbi.nlm.nih.gov/31908862/ |
| rayyan-751570870 | Sicuri E and Yaya Bocoum F and Nonvignon J and Alonso S and Fakih B and Bonsu G and Kariuki S and Leeuwenkamp O and Munguambe K and Mrisho M and Were V and Sauboin C | The Costs of Implementing Vaccination With the RTS,S Malaria Vaccine in Five Sub-Saharan African Countries. | 2019 | MDM policy & practice | 4 | 23814683 19896280 | RAYYAN-INCLUSION: {"Richard"=>"Excluded"} \| RAYYAN-EXCLUSION-REASONS: wrong outcome,wrong study design | 10.1177/2381468319896280 | Vaccination;Malaria;Vaccines | PMC6923697 | https://pubmed.ncbi.nlm.nih.gov/31903424/ |
| rayyan-751570871 | Enoch AJ and English M and McGivern G and Shepperd S | Variability in the use of pulse oximeters with children in Kenyan hospitals: A mixed-methods analysis. | 2019 | PLoS medicine | 16 | e1002987 | RAYYAN-INCLUSION: {"Richard"=>"Excluded"} \| RAYYAN-EXCLUSION-REASONS: wrong outcome,wrong study design,wrong infection / disease | 10.1371/journal.pmed.1002987 | Child;Child, Preschool;Female;Health Personnel/statistics & | PMC6938307 | https://pubmed.ncbi.nlm.nih.gov/31891572/ |
| rayyan-751570872 | Ling XX and Jin JJ and Zhu GD and Wang WM and Cao YY and Yang MM and Zhou HY and Cao J and Huang JY | Cost-effectiveness analysis of malaria rapid diagnostic tests: a systematic review. | 2019 | Infectious diseases of | 8 | 104 | RAYYAN-INCLUSION: {"Richard"=>"Excluded"} \| RAYYAN-EXCLUSION-REASONS: wrong outcome,wrong study design \| USER-NOTES: {"Richard"=>["Costing study"]} | 10.1186/s40249-019-0615-8 | *Cost-Benefit Analysis;Diagnostic Tests, Routine/*economics;Humans; | PMC6937952 | https://pubmed.ncbi.nlm.nih.gov/31888731/ |
| rayyan-751570873 | Pull L and Lupoglazoff JM and Beardmore M and Michel JF and Buffet P and Bouchaud O and Siriez JY | Artenimol-piperaquine in children with uncomplicated imported falciparum malaria: experience from a prospective cohort. | 2019 | Malaria journal | 18 | 419 | RAYYAN-INCLUSION: {"Richard"=>"Excluded"} \| RAYYAN-EXCLUSION-REASONS: wrong outcome,wrong study design | 10.1186/s12936-019-3047-9 | Adolescent;Artemisinins/*therapeutic use;Child;Child, Preschool;Communicable | PMC6915931 | https://pubmed.ncbi.nlm.nih.gov/31843017/ |
| rayyan-751570874 | Ley B and Winasti Satyagraha A and Rahmat H and von Fricken ME and Douglas NM and Pfeffer DA and Espino F and von Seidlein L and Henriques G and Oo NN and Menard D and | Performance of the Access Bio/CareStart rapid diagnostic test for the detection of glucose-6-phosphate dehydrogenase deficiency: AÂ systematic review and meta-analysis. | 2019 | PLoS medicine | 16 | e1002992 | RAYYAN-INCLUSION: {"Richard"=>"Excluded"} \| RAYYAN-EXCLUSION-REASONS: wrong outcome,wrong study design,wrong publication type \| USER-NOTES: {"Richard"=>["Review"]} | 10.1371/journal.pmed.1002992 | *Diagnostic Tests, Routine/methods;Endemic Diseases;Female;Glucosephos | PMC6910667 | https://pubmed.ncbi.nlm.nih.gov/31834890/ |
| rayyan-751570875 | Devine A and Pasaribu AP and Teferi T and Pham HT and Awab GR and Contantia F and Nguyen TN and Ngo VT and Tran TH and Hailu A and Gilchrist K and Green JA and Koh GC | Provider and household costs of Plasmodium vivax malaria episodes: a multicountry comparative analysis of primary trial data. | 2019 | Bulletin of the World | 97 | 828-836 | RAYYAN-INCLUSION: {"Richard"=>"Excluded"} \| RAYYAN-EXCLUSION-REASONS: wrong outcome,wrong study design | 10.2471/BLT.18.226688 | Absenteeism;Adolescent;Adult;Aged;Aminoquinolines/economics/*therapeutic | PMC6883272 | https://pubmed.ncbi.nlm.nih.gov/31819291/ |
| rayyan-751570876 | Ome-Kaius M and Kattenberg JH and Zaloumis S and Siba M and Kiniboro B and Jally S and Razook Z and Mantila D and Sui D and Ginny J and Rosanas-Urgell A and Karl S and Obadia T | Differential impact of malaria control interventions on P. falciparum and P. vivax infections in young Papua New Guinean children. | 2019 | BMC medicine | 17 | 220 | RAYYAN-INCLUSION: {"Richard"=>"Excluded"} \| RAYYAN-EXCLUSION-REASONS: wrong outcome,wrong study design,wrong study duration | 10.1186/s12916-019-1456-9 | Animals;Child, Preschool;Female;Humans;Incidence;Infant;Longitudinal | PMC6900859 | https://pubmed.ncbi.nlm.nih.gov/31813381/ |
| rayyan-751570877 | Smith TA and Pemberton-Ross P and Penny MA and Chitnis N | Resurgence of malaria infection after mass treatment: a simulation study. | 2019 | Malaria journal | 18 | 409 | RAYYAN-INCLUSION: {"Richard"=>"Excluded"} \| RAYYAN-EXCLUSION-REASONS: wrong outcome,wrong study design | 10.1186/s12936-019-3019-0 | Animals;*Anopheles;Antimalarials/*administration & dosage;Incidence;Malaria/*epi | PMC6896478 | https://pubmed.ncbi.nlm.nih.gov/31805947/ |
| rayyan-751570878 | Dowden J and Mushamiri I and McFeely E and Apat D and Sacks J and Ben Amor Y | The impact of "male clinics" on health-seeking behaviors of adult men in rural Kenya. | 2019 | PloS one | 14 | e0224749 | RAYYAN-INCLUSION: {"Richard"=>"Excluded"} \| RAYYAN-EXCLUSION-REASONS: wrong outcome,wrong study design | 10.1371/journal.pone.0224749 | Adolescent;Adult;Aged;Aged, 80 and over;Ambulatory Care Facilities/economics/organizati | PMC6872147 | https://pubmed.ncbi.nlm.nih.gov/31751377/ |
| rayyan-751570879 | Rodriguez-Rodriguez D and Maraga S and Lorry L and Robinson LJ and Siba PM and Mueller I and Pulford J and Ross A and Hetzel MW | Repeated mosquito net distributions, improved treatment, and trends in malaria cases in sentinel health facilities in Papua New Guinea. | 2019 | Malaria journal | 18 | 364 | RAYYAN-INCLUSION: {"Richard"=>"Maybe"} \| RAYYAN-LABELS: Include (full text) | 10.1186/s12936-019-2993-6 | Artemisinins/*therapeutic use;Communicable Disease Control/*statistics & numerical | PMC6852945 | https://pubmed.ncbi.nlm.nih.gov/31718659/ |
| rayyan-751570880 | Kitojo C and Gutman JR and Chacky F and Kigadye E and Mkude S and Mandike R and Mohamed A and Reaves EJ and Walker P and Ishengoma DS | Estimating malaria burden among pregnant women using data from antenatal care centres in Tanzania: a population-based study. | 2019 | The Lancet. Global | 7 | e1695-e1705 | RAYYAN-INCLUSION: {"Richard"=>"Maybe"} \| RAYYAN-LABELS: Excluded (No intervention) | 10.1016/S2214-109X(19)30405-X | Cross-Sectional Studies;Female;Health Facilities;Humans;Malaria/*epi | PMC8752309 | https://pubmed.ncbi.nlm.nih.gov/31708150/ |
| rayyan-751570881 | Armstrong JF and Faccenda E and Harding SD and Pawson AJ and Southan C and Sharman JL and Campo B and Cavanagh DR and Alexander SPH and Davenport AP and Spedding M and | The IUPHAR/BPS Guide to PHARMACOLOGY in 2020: extending immunopharmacology content and introducing the IUPHAR/MMV Guide to MALARIA PHARMACOLOGY. | 2020 | Nucleic acids research | 48 | D1006-D1021 | RAYYAN-INCLUSION: {"Richard"=>"Excluded"} \| RAYYAN-EXCLUSION-REASONS: wrong outcome,wrong study design | 10.1093/nar/gkz951 | Antimalarials/*pharmacology/therapeutic use;*Databases, Factual;*Databases, | PMC7145572 | https://pubmed.ncbi.nlm.nih.gov/31691834/ |
| rayyan-751570882 | Ferdinand DY and Nadlaou B and Samuel N and Oscar BY and Raphael M and Christophe N and Lopes S and Allan R | Evaluation of the effectiveness of community health workers in the fight against malaria in the Central African Republic (2012-2017). | 2020 | Transactions of the Royal | 114 | 173-184 | RAYYAN-INCLUSION: {"Richard"=>"Excluded"} \| RAYYAN-EXCLUSION-REASONS: wrong study design | 10.1093/trstmh/trz104 | Central African Republic;*Community Health Workers;Diagnostic Tests, | | https://pubmed.ncbi.nlm.nih.gov/31687768/ |
| rayyan-751570883 | Oniyangi O and Omari AA | Malaria chemoprophylaxis in sickle cell disease. | 2019 | The Cochrane database | 2019 | | RAYYAN-INCLUSION: {"Richard"=>"Excluded"} \| RAYYAN-EXCLUSION-REASONS: wrong outcome,wrong study design | 10.1002/14651858.CD003489.pub2 | Anemia, Sickle Cell/*complications;Antimalarials/*therapeutic | | https://pubmed.ncbi.nlm.nih.gov/31681984/ |
| rayyan-751570884 | Kubalalika PJ | Malaria Intermittent Preventive Treatment (IPTi) pharmacovigilance in Malawi: A case of Lilongwe district. | 2019 | Online journal of public | 11 | e9 | RAYYAN-INCLUSION: {"Richard"=>"Excluded"} \| RAYYAN-EXCLUSION-REASONS: wrong outcome,wrong study design | 10.5210/ojphi.v11i2.9956 | Malaria | PMC6788894 | https://pubmed.ncbi.nlm.nih.gov/31632603/ |
| rayyan-751570885 | Heraud P and Chatchawal P and Wongwattanakul M and Tippayawat P and Doerig C and Jearanaikoon P and Perez-Guaita D and Wood BR | Infrared spectroscopy coupled to cloud-based data management as a tool to diagnose malaria: a pilot study in a malaria-endemic country. | 2019 | Malaria journal | 18 | 348 | RAYYAN-INCLUSION: {"Richard"=>"Excluded"} \| RAYYAN-EXCLUSION-REASONS: wrong outcome,wrong study design | 10.1186/s12936-019-2945-1 | Adolescent;Adult;Aged;Aged, 80 and over;Child;Child, Preschool;*Cloud | PMC6794904 | https://pubmed.ncbi.nlm.nih.gov/31619246/ |
| rayyan-751570886 | Li J and Shi Y and Zhang W and Yan H and Lin K and Wei S and Wei H and Yang Y and Huang S and Lu Y and Ma A and Qin J | K13-propeller gene polymorphisms of Plasmodium falciparum and the therapeutic effect of artesunate among migrant workers returning to Guangxi, China (2014-2017). | 2019 | Malaria journal | 18 | 349 | RAYYAN-INCLUSION: {"Richard"=>"Excluded"} \| RAYYAN-EXCLUSION-REASONS: wrong outcome,wrong study design | 10.1186/s12936-019-2984-7 | Antimalarials/*therapeutic use;Artemisinins/*therapeutic use;Artesunate/*therapeutic | PMC6794752 | https://pubmed.ncbi.nlm.nih.gov/31619243/ |

| | | | | | | | | | | | |
|---|---|---|---|---|---|---|---|---|---|---|---|
| rayyan-751570887 | Jacobs J and Hardy L and Semret M and Lunguya O and Phe T and Affolabi D and Yansouni C and Vandenberg O | Diagnostic Bacteriology in District Hospitals in Sub-Saharan Africa: At the Forefront of the Containment of Antimicrobial Resistance. | 2019 | Frontiers in medicine | 6 | 205 | RAYYAN-INCLUSION: {"Richard"=>"Excluded"} \| RAYYAN-EXCLUSION-REASONS: wrong outcome,wrong study design,wrong infection / disease | 10.3389/fmed.2019.00205 | Africa South of the Sahara | PMC6771306 | https://pubmed.ncbi.nlm.nih.gov/31608280/ |
| rayyan-751570888 | Felger I and Snounou G and Hastings I and Moehrle JJ and Beck HP | PCR correction strategies for malaria drug trials: updates and clarifications. | 2020 | The Lancet. Infectious | 20 | e20-e25 | RAYYAN-INCLUSION: {"Richard"=>"Excluded"} \| RAYYAN-EXCLUSION-REASONS: wrong outcome,wrong study design | 10.1016/S1473-3099(19)30426-8 | Antimalarials/*therapeutic use;*Clinical Trials as Topic;Genotype;Genotyping | | https://pubmed.ncbi.nlm.nih.gov/31540841/ |
| rayyan-751570889 | Collins KA and Ouedraogo A and Guelbeogo WM and Awandu SS and Stone W and Soulama I and Ouattara MS and Nombre A and Diarra A and Bradley J and Selvaraj P and Gerardin J and | Investigating the impact of enhanced community case management and monthly screening and treatment on the transmissibility of malaria infections in Burkina Faso: study protocol for a cluster-randomised trial. | 2019 | BMJ open | 9 | e030598 | RAYYAN-INCLUSION: {"Richard"=>"Excluded"} \| RAYYAN-EXCLUSION-REASONS: wrong publication type \| USER-NOTES: {"Richard"=>["Protocol"]} | 10.1136/bmjopen-2019-030598 | Adult;*Asymptomatic Infections/epidemiology/therapy;Burkina | PMC6747640 | https://pubmed.ncbi.nlm.nih.gov/31519680/ |
| rayyan-751570890 | Unger HW and Rosanas-Urgell A and Robinson LJ and Ome-Kaius M and Jally S and Umbers AJ and Pomat W and Mueller I and Kattenberg E and Rogerson SJ | Microscopic and submicroscopic Plasmodium falciparum infection, maternal anaemia and adverse pregnancy outcomes in Papua New Guinea: a cohort study. | 2019 | Malaria journal | 18 | 302 | RAYYAN-INCLUSION: {"Richard"=>"Excluded"} \| RAYYAN-EXCLUSION-REASONS: wrong study design | 10.1186/s12936-019-2931-7 | Adult;Anemia/*parasitology;Anti-Bacterial Agents/administration & | PMC6720091 | https://pubmed.ncbi.nlm.nih.gov/31477117/ |
| rayyan-751570891 | Mwesigwa J and Slater H and Bradley J and Saidy B and Ceesay F and Whittaker C and Kandeh B and Nkwakamna D and Drakeley C and Van Geertruyden JP and Bousema T and Achan | Field performance of the malaria highly sensitive rapid diagnostic test in a setting of varying malaria transmission. | 2019 | Malaria journal | 18 | 288 | RAYYAN-INCLUSION: {"Richard"=>"Excluded"} \| RAYYAN-EXCLUSION-REASONS: wrong outcome,wrong study design \| USER-NOTES: {"Richard"=>["Diag study"]} | 10.1186/s12936-019-2929-1 | Adolescent;Adult;Aged;Aged, 80 and over;Child;Child, Preschool;Cross-Sectional | PMC6712604 | https://pubmed.ncbi.nlm.nih.gov/31455349/ |
| rayyan-751570892 | Raman J and Kagoro FM and Mabuza A and Malatje G and Reid A and Frean J and Barnes KI | Absence of kelch13 artemisinin resistance markers but strong selection for lumefantrine-tolerance molecular markers following 18Â years of artemisinin-based combination therapy use in Mpumalanga Province, South Africa (2001-2018). | 2019 | Malaria journal | 18 | 280 | RAYYAN-INCLUSION: {"Richard"=>"Excluded"} \| RAYYAN-EXCLUSION-REASONS: wrong outcome,wrong study design \| USER-NOTES: {"Richard"=>["Rx study"]} | 10.1186/s12936-019-2911-y | Antimalarials/*pharmacology;Artemisinins/*pharmacology;Diagnostic Tests, Routine;Drug | PMC6704579 | https://pubmed.ncbi.nlm.nih.gov/31438951/ |
| rayyan-751570893 | LaupÃ¨ze B and HervÃ© C and Di Pasquale A and Tavares Da Silva F | Adjuvant Systems for vaccines: 13â€¯years of post-licensure experience in diverse populations have progressed the way adjuvanted vaccine safety is investigated and understood. | 2019 | Vaccine | 37 | 5670-5680 | RAYYAN-INCLUSION: {"Richard"=>"Excluded"} \| RAYYAN-EXCLUSION-REASONS: wrong outcome,wrong study design \| USER-NOTES: {"Richard"=>["Vx study"]} | 10.1016/j.vaccine.2019.07.098 | *Adjuvants, Immunologic;Animals;Drug Compounding;Humans;Licens | | https://pubmed.ncbi.nlm.nih.gov/31420171/ |
| rayyan-751570894 | DobaÃ±o C and Ubillos I and Jairoce C and Gyan B and Vidal M and JimÃ©nez A and Santano R and Dosoo D and Nhabomba AJ and Ayestaran A and Aguilar R and Williams NA and DÃez- | RTS,S/AS01E immunization increases antibody responses to vaccine-unrelated Plasmodium falciparum antigens associated with protection against clinical malaria in African children: a case-control study. | 2019 | BMC medicine | 17 | 157 | RAYYAN-INCLUSION: {"Richard"=>"Excluded"} \| RAYYAN-EXCLUSION-REASONS: wrong outcome,wrong study design \| USER-NOTES: {"Richard"=>["Vx study"]} | 10.1186/s12916-019-1378-6 | Antibodies, Protozoan/*immunology;Antibody Formation;Antigens, | PMC6693200 | https://pubmed.ncbi.nlm.nih.gov/31409398/ |
| rayyan-751570895 | Fang Y and Shi WQ and Wu JT and Li YY and Xue JB and Zhang Y | Resistance to pyrethroid and organophosphate insecticides, and the geographical distribution and polymorphisms of target-site mutations in voltage-gated sodium channel and acetylcholinesterase 1 genes in Anopheles sinensis populations in Shanghai, China. | 2019 | Parasites & vectors | 12 | 396 | RAYYAN-INCLUSION: {"Richard"=>"Excluded"} \| RAYYAN-EXCLUSION-REASONS: wrong outcome,wrong study design \| USER-NOTES: {"Richard"=>["Ento study"]} | 10.1186/s13071-019-3657-7 | Acetylcholinesterase/*genetics;Alleles;Animals;Anopheles/enzymology/*genetics;Biologica | PMC6688361 | https://pubmed.ncbi.nlm.nih.gov/31399130/ |
| rayyan-751570896 | Corder RM and Paula GA and Pincelli A and Ferreira MU | Statistical modeling of surveillance data to identify correlates of urban malaria risk: A population-based study in the Amazon Basin. | 2019 | PloS one | 14 | e0220980 | RAYYAN-INCLUSION: {"Richard"=>"Excluded"} \| RAYYAN-EXCLUSION-REASONS: wrong outcome,wrong study design \| USER-NOTES: {"Richard"=>["Epi / strat study"]} | 10.1371/journal.pone.0220980 | Adolescent;Adult;Aged;Aged, 80 and over;Brazil/epidemiology;Child | PMC6688813 | https://pubmed.ncbi.nlm.nih.gov/31398228/ |
| rayyan-751570897 | Serra-Casas E and Guetens P and Chiheb D and Gamboa D and Rosanas-Urgell A | A pilot evaluation of alternative procedures to simplify LAMP-based malaria diagnosis in field conditions. | 2019 | Acta tropica | 200 | 105125 | RAYYAN-INCLUSION: {"Richard"=>"Excluded"} \| RAYYAN-EXCLUSION-REASONS: wrong outcome,wrong study design \| USER-NOTES: {"Richard"=>["Dx study"]} | 10.1016/j.actatropica.2019.105125 | Diagnostic Tests, Routine/*methods;Humans;Malaria/*diagnosis;Mass | | https://pubmed.ncbi.nlm.nih.gov/31394079/ |
| rayyan-751570899 | Sun Y and Xiang D and Chen C and He S and Qi H and Wang C | Infected RBC flag/parameter provided by Mindray BC-6800 haematology analyzer aid the diagnosis of malaria. | 2019 | Malaria journal | 18 | 262 | RAYYAN-INCLUSION: {"Richard"=>"Excluded"} \| RAYYAN-EXCLUSION-REASONS: wrong population | 10.1186/s12936-019-2890-z | Adolescent;Adult;Aged;Blood/*parasitology;Blood Chemical Analysis/instrumentation/*me | PMC6668162 | https://pubmed.ncbi.nlm.nih.gov/31366365/ |
| rayyan-751570900 | Reiker T and Chitnis N and Smith T | Modelling reactive case detection strategies for interrupting transmission of Plasmodium falciparum malaria. | 2019 | Malaria journal | 18 | 259 | RAYYAN-INCLUSION: {"Richard"=>"Excluded"} \| RAYYAN-EXCLUSION-REASONS: wrong outcome,wrong study design \| USER-NOTES: {"Richard"=>["Modeling study"]} | 10.1186/s12936-019-2893-9 | Disease Eradication/*methods;Humans;Incidence;Malaria, | PMC6668148 | https://pubmed.ncbi.nlm.nih.gov/31362768/ |
| rayyan-751570901 | Nag S and Ursing J and Rodrigues A and Crespo M and Krogsgaard C and Lund O and Aarestrup FM and Alifrangis M and Kofoed PE | Proof of concept: used malaria rapid diagnostic tests applied for parallel sequencing for surveillance of molecular markers of anti-malarial resistance in Bissau, Guinea-Bissau during 2014-2017. | 2019 | Malaria journal | 18 | 252 | RAYYAN-INCLUSION: {"Richard"=>"Excluded"} \| RAYYAN-EXCLUSION-REASONS: wrong outcome,wrong study design \| USER-NOTES: {"Richard"=>["Diag study"]} | 10.1186/s12936-019-2894-8 | Adolescent;Adult;Child;Child, Preschool;Diagnostic Tests, Routine/*statistics & | PMC6660714 | https://pubmed.ncbi.nlm.nih.gov/31349834/ |
| rayyan-751570902 | Lechthaler F and Matthys B and Lechthaler-Felber G and Likwela JL and Mavoko HM and Rika JM and Mutombo MM and Ruckstuhl L and Barczyk J and Shargie E and Prytherch H | Trends in reported malaria cases and the effects of malaria control in the Democratic Republic of the Congo. | 2019 | PloS one | 14 | e0219853 | RAYYAN-INCLUSION: {"Richard"=>"Excluded"} \| RAYYAN-EXCLUSION-REASONS: wrong outcome,wrong study design \| USER-NOTES: {"Richard"=>["Epi study"]} | 10.1371/journal.pone.0219853 | Adolescent;Child;Child, Preschool;Democratic Republic of the | PMC6658057 | https://pubmed.ncbi.nlm.nih.gov/31344062/ |
| rayyan-751570903 | Smith S and Koech R and Nzorubara D and Otieno M and Wong L and Bhat G and van den Bogaart E and Thuranira M and Onchonga D and Rinke de Wit TF | Connected diagnostics: linking digital rapid diagnostic tests and mobile health wallets to diagnose and treat brucellosis in Samburu, Kenya. | 2019 | BMC medical informati | 19 | 139 | RAYYAN-INCLUSION: {"Richard"=>"Excluded"} \| RAYYAN-EXCLUSION-REASONS: wrong outcome,wrong study design \| USER-NOTES: {"Richard"=>["Diag study"]} | 10.1186/s12911-019-0854-4 | Adolescent;Adult;Aged;Aged, 80 and over;Brucellosis/*diagnosis/ep | PMC6647279 | https://pubmed.ncbi.nlm.nih.gov/31331394/ |
| rayyan-751570904 | Ajonina MU and Ware KB and Ngonga DB and Abong RA and Nfor CK and Apinjoh TO | Malaria Perceptions among Medicine Vendors in Buea Community: An Assessment of Knowledge of Malaria and Conditions of Antimalarial Drug Dispensing. | 2019 | Innovations in pharmacy | 10 | | RAYYAN-INCLUSION: {"Richard"=>"Excluded"} \| RAYYAN-EXCLUSION-REASONS: wrong outcome,wrong study design \| USER-NOTES: {"Richard"=>["QUal study"]} | 10.24926/iip.v10i3.1381 | Antimalarials | PMC8127083 | https://pubmed.ncbi.nlm.nih.gov/34007559/ |
| rayyan-751570905 | Olasehinde GI and Diji-Geske RI and Fadina I and Arogundade D and Darby P and Adeleke A and Dokunmu TM and Adebayo AH and Oyelade J | Epidemiology of Plasmodium falciparum infection and drug resistance markers in Ota Area, Southwestern Nigeria. | 2019 | Infection and drug resistanc | 12 | 1941-1949 | RAYYAN-INCLUSION: {"Richard"=>"Excluded"} \| RAYYAN-EXCLUSION-REASONS: wrong outcome,wrong study design \| USER-NOTES: {"Richard"=>["Rx study"]} | 10.2147/IDR.S190386 | Plasmodium falciparum;Drug Resistance;Nigeria;Plasmodium | PMC6616117 | https://pubmed.ncbi.nlm.nih.gov/31308714/ |
| rayyan-751570906 | Erhabor O and Abdulrahaman A and Erhabor T | Effects of malaria parasitaemia on some haemostatic parameters among pregnant women of African descent in Specialist Hospital Sokoto, Nigeria. | 2020 | Human antibodies | 28 | 21-28 | RAYYAN-INCLUSION: {"Richard"=>"Excluded"} \| RAYYAN-EXCLUSION-REASONS: wrong outcome,wrong study design \| USER-NOTES: {"Richard"=>["Dx study"]} | 10.3233/HAB-190389 | Adolescent;Adult;Blood Coagulation/*physiology;Blood Platelets/physiology;Female;H | | https://pubmed.ncbi.nlm.nih.gov/31282411/ |
| rayyan-751570907 | Prosnitz D and Herrera S and Coelho H and Moonzwe Davis L and Zalisk K and Yourkavitch J | Evidence of Impact: iCCM as a strategy to save lives of children under five. | 2019 | Journal of global health | 9 | 010801 | RAYYAN-INCLUSION: {"Richard"=>"Maybe"} \| RAYYAN-LABELS: Excluded (after full text) | 10.7189/jogh.09.010801 | Case Management/*organization & administration;Child | PMC6594661 | https://pubmed.ncbi.nlm.nih.gov/31263547/ |
| rayyan-751570908 | Monroe A and Mihayo K and Okumu F and Finda M and Moore S and Koenker H and Lynch M and Haji K and Abbas F and Ali A and Greer G and Harvey S | Human behaviour and residual malaria transmission in Zanzibar: findings from in-depth interviews and direct observation of community events. | 2019 | Malaria journal | 18 | 220 | RAYYAN-INCLUSION: {"Richard"=>"Excluded"} \| RAYYAN-EXCLUSION-REASONS: wrong outcome,wrong study design \| USER-NOTES: {"Richard"=>["SBC study"]} | 10.1186/s12936-019-2855-2 | Adult;Aged;Aged, 80 and over;Circadian Rhythm;Communicable | PMC6604484 | https://pubmed.ncbi.nlm.nih.gov/31262306/ |
| rayyan-751570909 | Battle KE and Lucas TCD and Nguyen M and Howes RE and Nandi AK and Twohig KA and Pfeffer DA and Cameron E and Rao PC and Casey D and Gibson HS and Rozier JA and | Mapping the global endemicity and clinical burden of Plasmodium vivax, 2000-17: a spatial and temporal modelling study. | 2019 | Lancet (London, England) | 394 | 332-343 | RAYYAN-INCLUSION: {"Richard"=>"Excluded"} \| RAYYAN-EXCLUSION-REASONS: wrong outcome,wrong study design \| USER-NOTES: {"Richard"=>["Epi study"]} | 10.1016/S0140-6736(19)31096-7 | Africa/epidemiology;Americas/epidemiology;Asia, Southeastern/epidemiology;Ba | PMC6675736 | https://pubmed.ncbi.nlm.nih.gov/31229233/ |
| rayyan-751570910 | Mussa A and Talib M and Mohamed Z and Hajissa K | Genetic diversity of Plasmodium falciparum histidine-rich protein 2 (PfHRP2) and its effect on the performance of PfHRP2-based rapid diagnostic tests. | 2019 | BMC research notes | 12 | 334 | RAYYAN-INCLUSION: {"Richard"=>"Excluded"} \| RAYYAN-EXCLUSION-REASONS: wrong outcome,wrong study design \| USER-NOTES: {"Richard"=>["Dx / Rx study"]} | 10.1186/s13104-019-4361-6 | Amino Acid Sequence;Antigens, Protozoan/*genetics;Diagnosis | PMC6560730 | https://pubmed.ncbi.nlm.nih.gov/31186056/ |
| rayyan-751570911 | Zainabadi K and Nyunt MM and Plowe CV | An improved nucleic acid extraction method from dried blood spots for amplification of Plasmodium falciparum kelch13 for detection of artemisinin resistance. | 2019 | Malaria journal | 18 | 192 | RAYYAN-INCLUSION: {"Richard"=>"Excluded"} \| RAYYAN-EXCLUSION-REASONS: wrong outcome,wrong study design \| USER-NOTES: {"Richard"=>["Diag study"]} | 10.1186/s12936-019-2817-8 | Antimalarials/pharmacology;Artemisinins/*pharmacology;Asia, | PMC6558694 | https://pubmed.ncbi.nlm.nih.gov/31185976/ |
| rayyan-751570912 | Theiss-Nyland K and Lines J and Fine P | Can ITN distribution policies increase children's ITN use? A DHS analysis. | 2019 | Malaria journal | 18 | 191 | RAYYAN-INCLUSION: {"Richard"=>"Excluded"} \| RAYYAN-EXCLUSION-REASONS: wrong outcome,wrong study design | 10.1186/s12936-019-2824-9 | Adolescent;Adult;Africa;Child, Preschool;*Equipment and Supplies | PMC6555912 | https://pubmed.ncbi.nlm.nih.gov/31176365/ |
| rayyan-751570914 | Andrada A and Herrera S and Inyang U and Mohammed AB and Uhomoibhi P and YÃ© Y | A subnational profiling analysis reveals regional differences as the main predictor of ITN ownership and use in Nigeria. | 2019 | Malaria journal | 18 | 185 | RAYYAN-INCLUSION: {"Richard"=>"Excluded"} \| RAYYAN-EXCLUSION-REASONS: wrong outcome,wrong study design \| USER-NOTES: {"Richard"=>["No routine data used"]} | 10.1186/s12936-019-2816-9 | Cross-Sectional Studies;*Family Characteristics;Geography;Hu | PMC6540480 | https://pubmed.ncbi.nlm.nih.gov/31138216/ |
| rayyan-751570915 | Decuyper II and Van Damme P and Soentjens P and Wojciechowski M | Low adherence with national travel medicine recommendations in Belgian expatriate children: A retrospective analysis. | 2019 | Travel medicine and | 32 | 101424 | RAYYAN-INCLUSION: {"Richard"=>"Excluded"} \| RAYYAN-EXCLUSION-REASONS: wrong outcome,wrong study design | 10.1016/j.tmaid.2019.05.010 | Only Child;Child;Travel | | https://pubmed.ncbi.nlm.nih.gov/31085332/ |
| rayyan-751570916 | Fuseini G and Phiri WP and von Fricken ME and Smith J and Garcia GA | Evaluation of the residual effectiveness of Fludoraâ„¢ fusion WP-SB, a combination of clothianidin and deltamethrin, for the control of pyrethroid-resistant malaria vectors on Bioko Island, Equatorial Guinea. | 2019 | Acta tropica | 196 | 42-47 | RAYYAN-INCLUSION: {"Richard"=>"Excluded"} \| RAYYAN-EXCLUSION-REASONS: wrong outcome,wrong study design \| USER-NOTES: {"Richard"=>["Ento study"]} | 10.1016/j.actatropica.2019.05.006 | Animals;Anopheles/*drug effects;Equatorial Guinea/epidemiology;Guanidin | | https://pubmed.ncbi.nlm.nih.gov/31077641/ |
| rayyan-751570917 | Kifle MM and Teklemariam TT and Teweldeberhan AM and Tesfamariam EH and Andegiorgish AK and Azaria Kidane E | Malaria Risk Stratification and Modeling the Effect of Rainfall on Malaria Incidence in Eritrea. | 2019 | Journal of environmental and | 2019 | 7314129 | RAYYAN-INCLUSION: {"Richard"=>"Excluded"} \| RAYYAN-EXCLUSION-REASONS: wrong outcome,wrong study design \| USER-NOTES: {"Richard"=>["Epi / strat study"]} | 10.1155/2019/7314129 | Bayes Theorem;Eritrea/epidemiology;Forecasting/*methods;Huma | PMC6466923 | https://pubmed.ncbi.nlm.nih.gov/31061663/ |
| rayyan-751570918 | Candrinho B and Plucinski MM and Colborn JM and da Silva M and Mathe G and Dimene M and Chico AR and Castel-Branco AC and Brito F and Andela M and Ponce de Leon G and | Quality of malaria services offered in public health facilities in three provinces of Mozambique: a cross-sectional study. | 2019 | Malaria journal | 18 | 162 | RAYYAN-INCLUSION: {"Richard"=>"Excluded"} \| RAYYAN-EXCLUSION-REASONS: wrong outcome,wrong study design | 10.1186/s12936-019-2796-9 | Adolescent;Adult;Ambulatory Care;Antimalarials/therapeutic use;Child;Child, | PMC6503352 | https://pubmed.ncbi.nlm.nih.gov/31060605/ |
| rayyan-751570919 | Watson OJ and Verity R and Ghani AC and Garske T and Cunningham J and Tshefu A and Mwandagalirwa MK and Meshnick SR and Parr JB and Slater HC | Impact of seasonal variations in Plasmodium falciparum malaria transmission on the surveillance of pfhrp2 gene deletions. | 2019 | eLife | 8 | | RAYYAN-INCLUSION: {"Richard"=>"Excluded"} \| RAYYAN-EXCLUSION-REASONS: wrong outcome,wrong study design \| USER-NOTES: {"Richard"=>["Genomic surveillance"]} | 10.7554/eLife.40339 | Africa;Antigens, Protozoan/*genetics;Diagnostic Errors;Diagnostic Tests, | PMC6533063 | https://pubmed.ncbi.nlm.nih.gov/31045490/ |
| rayyan-751570921 | Nambuusi BB and Ssempiira J and Makumbi FE and Kasasa S and Vounatsou P | The effects and contribution of childhood diseases on the geographical distribution of all-cause under-five mortality in Uganda. | 2019 | Parasite epidemiology and | 5 | e00089 | RAYYAN-INCLUSION: {"Richard"=>"Excluded"} \| RAYYAN-EXCLUSION-REASONS: wrong outcome,wrong study design \| USER-NOTES: {"Richard"=>["No routine data "]} | 10.1016/j.parepi.2019.e00089 | Uganda | PMC6424012 | https://pubmed.ncbi.nlm.nih.gov/30923753/ |

| | | | | | | | | | | | |
|---|---|---|---|---|---|---|---|---|---|---|---|
| rayyan-751570922 | Jahan F and Khan NH and Wahid S and Ullah Z and Kausar A and Ali N | Malaria epidemiology and comparative reliability of diagnostic tools in Bannu; an endemic malaria focus in south of Khyber Pakhtunkhwa, Pakistan. | 2019 | Pathogens and global | 113 | 75-85 | RAYYAN-INCLUSION: {"Richard"=>"Excluded"} \| RAYYAN-EXCLUSION-REASONS: wrong outcome,wrong study design \| USER-NOTES: {"Richard"=>["Diag study"]} | 10.1080/20477724.2019.1595904 | Diagnostic Tests, Routine/*methods;Endemic Diseases;Humans;Immunoassa | PMC6493316 | https://pubmed.ncbi.nlm.nih.gov/30894081/ |
| rayyan-751570923 | Mfuh KO and Achonduh-Atijegbe OA and Bekindaka ON and Esemu LF and Mbakop CD and Gandhi K and Leke RGF and Taylor DW and Nerurkar VR | A comparison of thick-film microscopy, rapid diagnostic test, and polymerase chain reaction for accurate diagnosis of Plasmodium falciparum malaria. | 2019 | Malaria journal | 18 | 73 | RAYYAN-INCLUSION: {"Richard"=>"Excluded"} \| RAYYAN-EXCLUSION-REASONS: wrong outcome,wrong study design \| USER-NOTES: {"Richard"=>["Diag study"]} | 10.1186/s12936-019-2711-4 | Adolescent;Adult;Aged;Aged, 80 and over;Blood/*parasitology;Cam | PMC6416847 | https://pubmed.ncbi.nlm.nih.gov/30866947/ |
| rayyan-751570924 | Baia-da-Silva DC and Brito-Sousa JD and Rodovalho SR and Peterka C and Moresco G and Lapouble OMM and Melo GC and Sampaio VS and Alecrim MDGC and Pimenta P and Lima | Current vector control challenges in the fight against malaria in Brazil. | 2019 | Revista da Sociedad | 52 | e20180542 | RAYYAN-INCLUSION: {"Richard"=>"Excluded"} \| RAYYAN-EXCLUSION-REASONS: wrong outcome,wrong study design \| USER-NOTES: {"Richard"=>["Ento study"]} | 10.1590/0037-8682-0542-2018 | Animals;*Anopheles;Brazil/epidemiology;Humans;Insecticide Resistance;Insecticides/pharm | | https://pubmed.ncbi.nlm.nih.gov/30843971/ |
| rayyan-751570925 | Balami AD and Said SM and Zulkefli NAM and Bachok N and Audu B | Effects of a health educational intervention on malaria knowledge, motivation, and behavioural skills: a randomized controlled trial. | 2019 | Malaria journal | 18 | 41 | RAYYAN-INCLUSION: {"Richard"=>"Excluded"} \| RAYYAN-EXCLUSION-REASONS: wrong outcome,wrong study design \| USER-NOTES: {"Richard"=>["SBC study"]} | 10.1186/s12936-019-2676-3 | Adolescent;Adult;Antimalarials/*therapeutic use;Behavior Therapy/*methods;Communic | PMC6381678 | https://pubmed.ncbi.nlm.nih.gov/30786906/ |
| rayyan-751570926 | Bajaj S and Wakode SR and Khan W and Manchanda S and Kumar S | Simultaneous HPTLC analysis and in vitro antileishmanic activity of various secondary metabolites in extract of the traditional medicinal herb Artabotrys hexapetalus (L.f.). | 2018 | Ayu | 39 | 92-100 | RAYYAN-INCLUSION: {"Richard"=>"Excluded"} \| RAYYAN-EXCLUSION-REASONS: wrong outcome,wrong study design \| USER-NOTES: {"Richard"=>["Diag / immunol study"]} | 10.4103/ayu.AYU_158_17 | Plants, Medicinal;Medicine, Traditional | PMC6369602 | https://pubmed.ncbi.nlm.nih.gov/30783364/ |
| rayyan-751570927 | Fontecha G and Pinto A and Escobar D and Matamoros G and Ortiz B | Genetic variability of Plasmodium falciparum histidine-rich proteins 2 and 3 in Central America. | 2019 | Malaria journal | 18 | 31 | RAYYAN-INCLUSION: {"Richard"=>"Excluded"} \| RAYYAN-EXCLUSION-REASONS: wrong outcome,wrong study design \| USER-NOTES: {"Richard"=>["Genomic surveillance"]} | 10.1186/s12936-019-2668-3 | Antigens, Protozoan/chemistry/*genetics;Diagnostic Tests, | PMC6357481 | https://pubmed.ncbi.nlm.nih.gov/30704496/ |
| rayyan-751570928 | Bonko MDA and Kiemde F and Tahita MC and Lompo P and Some AM and Tinto H and van Hensbroek MB and Mens PF and Schallig HDFH | The effect of malaria rapid diagnostic tests results on antimicrobial prescription practices of health care workers in Burkina Faso. | 2019 | Annals of clinical microbiol | 18 | 5 | RAYYAN-INCLUSION: {"Richard"=>"Excluded"} \| RAYYAN-EXCLUSION-REASONS: wrong outcome,wrong study design \| USER-NOTES: {"Richard"=>["Diag study"]} | 10.1186/s12941-019-0304-2 | Antigens, Protozoan;Antimalarials/*therapeutic use;Burkina | PMC6348635 | https://pubmed.ncbi.nlm.nih.gov/30691506/ |
| rayyan-751570929 | Daher A and Silva JCAL and Stevens A and Marchesini P and Fontes CJ and Ter Kuile FO and Lalloo DG | Evaluation of Plasmodium vivax malaria recurrence in Brazil. | 2019 | Malaria journal | 18 | 18 | RAYYAN-INCLUSION: {"Richard"=>"Excluded"} \| RAYYAN-EXCLUSION-REASONS: wrong outcome,wrong study design \| USER-NOTES: {"Richard"=>["Epi study"]} | 10.1186/s12936-019-2644-y | Adolescent;Adult;Aged;Aged, 80 and over;Antimalarials/*therapeuti | PMC6343355 | https://pubmed.ncbi.nlm.nih.gov/30670020/ |
| rayyan-751570930 | BjÃ¶rkman A and Shakely D and Ali AS and Morris U and Mkali H and Abbas AK and Al-Mafazy AW and Haji KA and Mcha J and Omar R and Cook J and Elfving K and Petzold M and Sachs | From high to low malaria transmission in Zanzibar-challenges and opportunities to achieve elimination. | 2019 | BMC medicine | 17 | 14 | RAYYAN-INCLUSION: {"Richard"=>"Included"} | 10.1186/s12916-018-1243-z | Adolescent;Child;Child, Preschool;Cross-Sectional Studies;Female;Humans;Incide | PMC6341737 | https://pubmed.ncbi.nlm.nih.gov/30665398/ |
| rayyan-751570931 | Orish V and Amegan Aho K and Ofori-Amoah J and Osei-Yobah J and Jamfaru I and Afeke I and Mac-Ankrah L and Adzaku F | Asymptomatic Plasmodium falciparum infection and poor school performance in primary school children in the Volta Region of Ghana. | 2018 | Ethiopian journal of health | 28 | 749-758 | RAYYAN-INCLUSION: {"Richard"=>"Excluded"} \| RAYYAN-EXCLUSION-REASONS: wrong outcome,wrong study design | 10.4314/ejhs.v28i6.10 | *Academic Performance;Adolescent;Child;Cross-Sectional | PMC6308746 | https://pubmed.ncbi.nlm.nih.gov/30607092/ |
| rayyan-751570932 | Tao D and McGill B and Hamerly T and Kobayashi T and Khare P and Dziedzic A and Leski T and Holtz A and Shull B and Jedlicka AE and Walzer A and Slowey PD and Slowey CC and | A saliva-based rapid test to quantify the infectious subclinical malaria parasite reservoir. | 2019 | Science translational | 11 | | RAYYAN-INCLUSION: {"Richard"=>"Excluded"} \| RAYYAN-EXCLUSION-REASONS: wrong outcome,wrong study design | 10.1126/scitranslmed.aan4479 | Adolescent;Animals;*Asymptomatic Infections;Biomarkers/metabo | PMC6441545 | https://pubmed.ncbi.nlm.nih.gov/30602535/ |
| rayyan-751570934 | Dimala CA and Kika BT and Kadia BM and Blencowe H | Current challenges and proposed solutions to the effective implementation of the RTS, S/AS01 Malaria Vaccine Program in sub-Saharan Africa: A systematic review. | 2018 | PloS one | 13 | e0209744 | RAYYAN-INCLUSION: {"Richard"=>"Excluded"} \| RAYYAN-EXCLUSION-REASONS: wrong outcome,wrong study design,wrong publication type \| USER-NOTES: {"Richard"=>["Review"]} | 10.1371/journal.pone.0209744 | Africa South of the Sahara;Humans;Immunization Programs/methods;Immunizat | PMC6312235 | https://pubmed.ncbi.nlm.nih.gov/30596732/ |
| rayyan-751570935 | Onono M and Abdi M and Opondo I and Okung'u J and Asadhi E and Nyamai R and Karimurio L and Okoth P and Qazi SA | Using the RE-AIM framework to evaluate the implementation of integrated community case management in Kenya. | 2018 | Acta paediatrica (Oslo, | 107 | 53-62 | RAYYAN-INCLUSION: {"Richard"=>"Excluded"} \| RAYYAN-EXCLUSION-REASONS: wrong outcome,wrong study design \| USER-NOTES: {"Richard"=>["No epi indicators"]} | 10.1111/apa.14662 | Child Health Services/standards/*statistics & numerical data;Child, | | https://pubmed.ncbi.nlm.nih.gov/30570791/ |
| rayyan-751570936 | Usman R and Umar AA and Gidado S and Gobir AA and Obi IF and Ajayi I and Ajumobi O | Predictors of malaria Rapid Diagnostic Tests' utilisation among healthcare workers in Zamfara State. | 2018 | PloS one | 13 | e0200856 | RAYYAN-INCLUSION: {"Richard"=>"Excluded"} \| RAYYAN-EXCLUSION-REASONS: wrong outcome,wrong study design \| USER-NOTES: {"Richard"=>["Diag study"]} | 10.1371/journal.pone.0200856 | Adult;Cross-Sectional Studies;Female;*Health Personnel;Humans;*Knowledg | PMC6294357 | https://pubmed.ncbi.nlm.nih.gov/30550562/ |
| rayyan-751570937 | Win Han Oo and Cutts JC and Agius PA and Kyaw Zayar Aung and Poe Poe Aung and Aung Thi and Nyi Nyi Zaw and Htin Kyaw Thu and Wai Yan Min Htay and Ataide R and O'Flaherty | Effectiveness of repellent delivered through village health volunteers on malaria incidence in villages in South-East Myanmar: a stepped-wedge cluster-randomised controlled trial protocol. | 2018 | BMC infectious diseases | 18 | 663 | RAYYAN-INCLUSION: {"Richard"=>"Excluded"} \| RAYYAN-EXCLUSION-REASONS: wrong outcome,wrong study design \| USER-NOTES: {"Richard"=>["Protocol"]} | 10.1186/s12879-018-3566-y | Community Health Workers;Humans;Incidence;Insect | PMC6295052 | https://pubmed.ncbi.nlm.nih.gov/30547749/ |
| rayyan-751570938 | Bawa S and Shuaib F and Saidu M and Ningi A and Abdullahi S and Abba B and Idowu A and Alkasim J and Hammanyero K and Warigon C and Tegegne SG and Banda R and Korir C and | Conduct of vaccination in hard-to-reach areas to address potential polio reservoir areas, 2014-2015. | 2018 | BMC public health | 18 | 1312 | RAYYAN-INCLUSION: {"Richard"=>"Excluded"} \| RAYYAN-EXCLUSION-REASONS: wrong outcome,wrong study design,wrong infection / disease | 10.1186/s12889-018-6194-y | Child, Preschool;*Health Services Accessibility;Humans;Immuniz | PMC6291919 | https://pubmed.ncbi.nlm.nih.gov/30541501/ |
| rayyan-751570939 | Thielemans L and Hashmi A and Priscilla DD and Kho Paw M and Pimolsorntong T and Ngerseng T and Van Overmeire B and Proux S and Nosten F and McGready R and Carrara VI and | Laboratory validation and field usability assessment of a point-of-care test for serum bilirubin levels in neonates in a tropical setting. | 2018 | Wellcome open research | 3 | 110 | RAYYAN-INCLUSION: {"Richard"=>"Excluded"} \| RAYYAN-EXCLUSION-REASONS: wrong outcome,wrong study design | 10.12688/wellcomeopenres.14767.2 | Infant, Newborn | PMC6137410 | https://pubmed.ncbi.nlm.nih.gov/30271889/ |
| rayyan-751570940 | Hailu A and LindtjÃ¸rn B and Deressa W and Gari T and Loha E and Robberstad B | Cost-effectiveness of a combined intervention of long lasting insecticidal nets and indoor residual spraying compared with each intervention alone for malaria prevention in Ethiopia. | 2018 | Cost effectiveness and | 16 | 61 | RAYYAN-INCLUSION: {"Richard"=>"Excluded"} \| RAYYAN-EXCLUSION-REASONS: wrong outcome,wrong study design \| USER-NOTES: {"Richard"=>["Costing study"]} | 10.1186/s12962-018-0164-1 | Cost-Benefit Analysis;Malaria;Insecticides | PMC6251210 | https://pubmed.ncbi.nlm.nih.gov/30498400/ |
| rayyan-751570941 | Coghlan R and Stephens P and Mwale B and Siyanga M | A new approach to gathering pharmaceutical market data to support policy implementation and access to medicines: as demonstrated by malaria medicines in Zambia. | 2018 | Malaria journal | 17 | 444 | RAYYAN-INCLUSION: {"Richard"=>"Excluded"} \| RAYYAN-EXCLUSION-REASONS: wrong outcome,wrong study design \| USER-NOTES: {"Richard"=>["Diag / Rx study", "Rx study"]} | 10.1186/s12936-018-2594-9 | Antimalarials/*supply & distribution;*Databases, Factual;*Drug | PMC6267849 | https://pubmed.ncbi.nlm.nih.gov/30497504/ |
| rayyan-751570942 | Kessler A and van Eijk AM and Jamir L and Walton C and Carlton JM and Albert S | Malaria in Meghalaya: a systematic literature review and analysis of data from the National Vector-Borne Disease Control Programme. | 2018 | Malaria journal | 17 | 411 | RAYYAN-INCLUSION: {"Richard"=>"Excluded"} \| RAYYAN-EXCLUSION-REASONS: wrong outcome,wrong study design,wrong publication type \| USER-NOTES: {"Richard"=>["Review"]} | 10.1186/s12936-018-2563-3 | Animals;Anopheles/*physiology;Humans;Incidence;India/epidemiology;*Malaria, | PMC6219117 | https://pubmed.ncbi.nlm.nih.gov/30400879/ |
| rayyan-751570943 | Jawara M and Jatta E and Bell D and Burkot TR and Bradley J and Hunt V and Kandeh B and Jones C and Manjang AM and Pinder M and Stone S and D'Alessandro U and Knudsen J and | New Prototype Screened Doors and Windows for Excluding Mosquitoes from Houses: A Pilot Study in Rural Gambia. | 2018 | The American journal of | 99 | 1475-1484 | RAYYAN-INCLUSION: {"Richard"=>"Excluded"} \| RAYYAN-EXCLUSION-REASONS: wrong outcome,wrong study design \| USER-NOTES: {"Richard"=>["Ento study"]} | 10.4269/ajtmh.18-0660 | Adult;Animals;Anopheles/*parasitology/physiology;Child;Family | PMC6283509 | https://pubmed.ncbi.nlm.nih.gov/30350770/ |
| rayyan-751570944 | Wen-Ji Q and Jia-Jie J and Yuan-Yuan C and Wei-Ming W and Meng-Meng Y and Jun C and Yu X and Jia-Yan H and Guo-Ding Z | [Evaluation of an optimization strategy for use and management of malaria rapid diagnostic testing (RDT) in Jiangsu Province]. | 2018 | Zhongguo xue xi chong | 30 | 383-389 | RAYYAN-INCLUSION: {"Richard"=>"Excluded"} \| RAYYAN-EXCLUSION-REASONS: wrong outcome,wrong study design | 10.16250/j.32.1374.2018141 | China;*Diagnostic Tests, Routine/methods/standards;Humans;*Malaria/diagnosis;Mal | | https://pubmed.ncbi.nlm.nih.gov/30350500/ |
| rayyan-751570945 | Jia-Jie J and Wei-Ming W and Yuan-Yuan C and Guo-Ding Z and Hua-Yun Z and Jia-Yan H and Jun C | [Investigation on malaria rapid diagnostic testing (RDT) - related knowledge, attitude and practice in primary healthcare professionals of Jiangsu Province]. | 2018 | Zhongguo xue xi chong | 30 | 378-382 | RAYYAN-INCLUSION: {"Richard"=>"Excluded"} \| RAYYAN-EXCLUSION-REASONS: wrong outcome,wrong study design \| USER-NOTES: {"Richard"=>["Qual study"]} | 10.16250/j.32.1374.2018004 | China;*Diagnostic Tests, Routine/standards/statistics & numerical data;*Health | | https://pubmed.ncbi.nlm.nih.gov/30350499/ |
| rayyan-751570946 | McLean ARD and Wai HP and Thu AM and Khant ZS and Indrasuta C and Ashley EA and Kyaw TT and Day NPJ and Dondorp A and White NJ and Smithuis FM | Malaria elimination in remote communities requires integration of malaria control activities into general health care: an observational study and interrupted time series analysis in Myanmar. | 2018 | BMC medicine | 16 | 183 | RAYYAN-INCLUSION: {"Richard"=>"Maybe"} \| RAYYAN-LABELS: Include (full text) | 10.1186/s12916-018-1172-x | Community Health Services/methods/*organization & administration;Delivery | PMC6196466 | https://pubmed.ncbi.nlm.nih.gov/30343666/ |
| rayyan-751570947 | Henry M and Florey L and Youll S and Gutman JR | An analysis of country adoption and implementation of the 2012 WHO recommendations for intermittent preventive treatment for pregnant women in sub-Saharan Africa. | 2018 | Malaria journal | 17 | 364 | RAYYAN-INCLUSION: {"Richard"=>"Excluded"} \| RAYYAN-EXCLUSION-REASONS: wrong outcome,wrong study design,wrong publication type | 10.1186/s12936-018-2512-1 | Adolescent;Adult;Africa South of the Sahara;Antimalarials/*therape | PMC6192297 | https://pubmed.ncbi.nlm.nih.gov/30326904/ |
| rayyan-751570948 | Chinula D and Hamainza B and Chizema E and Kavishe DR and Sikaala CH and Killeen GF | Proportional decline of Anopheles quadriannulatus and increased contribution of An. arabiensis to the An. gambiae complex following introduction of indoor residual spraying with pirimiphos-methyl: an observational, retrospective secondary analysis of pre-existing data from south-east Zambia. | 2018 | Parasites & vectors | 11 | 544 | RAYYAN-INCLUSION: {"Richard"=>"Excluded"} \| RAYYAN-EXCLUSION-REASONS: wrong outcome,wrong study design \| USER-NOTES: {"Richard"=>["Ento study"]} | 10.1186/s13071-018-3121-0 | Animals;Anopheles/classification/*drug effects;Environment;Feeding | PMC6180389 | https://pubmed.ncbi.nlm.nih.gov/30305147/ |
| rayyan-751570949 | Batista-Dos-Santos SA and Freitas DRC and Raiol M and Cabral GF and Feio AC and PÃ³voa MM and Cunha MG and Ribeiro-Dos-Santos Ã, | Strategy to improve malaria surveillance system preventing transfusion-transmitted malaria in blood banks using molecular diagnostic. | 2018 | Malaria journal | 17 | 344 | RAYYAN-INCLUSION: {"Richard"=>"Excluded"} \| RAYYAN-EXCLUSION-REASONS: wrong outcome,wrong study design | 10.1186/s12936-018-2486-z | Blood/parasitology;*Blood Banks;Blood Transfusion;Brazil/epidemiolog | PMC6167815 | https://pubmed.ncbi.nlm.nih.gov/30285750/ |
| rayyan-751570950 | Liverani M and Teng S and Le MS and Coker R | Sharing public health data and information across borders: lessons from Southeast Asia. | 2018 | Globalization and health | 14 | 94 | RAYYAN-INCLUSION: {"Richard"=>"Excluded"} \| RAYYAN-EXCLUSION-REASONS: wrong outcome,wrong study design \| USER-NOTES: {"Richard"=>["Qual study"]} | 10.1186/s12992-018-0415-0 | Asia, Southeastern;Communicable Disease | PMC6162912 | https://pubmed.ncbi.nlm.nih.gov/30268139/ |
| rayyan-751570951 | Ferrara P and Masuet-Aumatell C and AgÃ¼ero F and Ramon-Torrell JM | The use of stand-by emergency treatment (SBET) for malaria in travellers: A systematic review and meta-analysis of observational studies. | 2018 | The Journal of infection | 77 | 455-462 | RAYYAN-INCLUSION: {"Richard"=>"Excluded"} \| RAYYAN-EXCLUSION-REASONS: wrong outcome,wrong study design,wrong publication type \| USER-NOTES: {"Richard"=>["Review"]} | 10.1016/j.jinf.2018.09.007 | Antimalarials/*therapeutic use;Diagnostic Tests, Routine;Emergency | | https://pubmed.ncbi.nlm.nih.gov/30267799/ |
| rayyan-751570952 | Pradhan N and Hazra RK | Can a non-invasive urine-based test become the next-generation diagnostic tool for malaria? | 2018 | Le infezioni in | 26 | 199-203 | RAYYAN-INCLUSION: {"Richard"=>"Excluded"} \| RAYYAN-EXCLUSION-REASONS: wrong outcome,wrong study design | | Chromatography, Affinity/methods;Colorimetry/methods;Diagnostic Tests, | | https://pubmed.ncbi.nlm.nih.gov/30246761/ |
| rayyan-751570953 | Berzosa P and de Lucio A and Romay-Barja M and Herrador Z and GonzÃ¡lez V and GarcÃa L and FernÃ¡ndez-MartÃnez A and Santana-Morales M and Ncogo P and Valladares B and | Comparison of three diagnostic methods (microscopy, RDT, and PCR) for the detection of malaria parasites in representative samples from Equatorial Guinea. | 2018 | Malaria journal | 17 | 333 | RAYYAN-INCLUSION: {"Richard"=>"Excluded"} \| RAYYAN-EXCLUSION-REASONS: wrong outcome,wrong study design \| USER-NOTES: {"Richard"=>["Diag study"]} | 10.1186/s12936-018-2481-4 | Adolescent;Adult;Aged;Aged, 80 and over;Animals;Child;Child, | PMC6142353 | https://pubmed.ncbi.nlm.nih.gov/30223852/ |
| rayyan-751570954 | Uwimana A and Penkunas MJ and Nisingizwe MP and Uyizeye D and Hakizimana D and Musanabaganwa C and Musabyimana JP and Ngwije A and Turate I and | Expanding home-based management of malaria to all age groups in Rwanda: analysis of acceptability and facility-level time-series data. | 2018 | Transactions of the Royal | 112 | 513-521 | RAYYAN-INCLUSION: {"Richard"=>"Included"} | 10.1093/trstmh/try093 | Adolescent;Adult;Antimalarials/*therapeutic use;Child;Community Health | | https://pubmed.ncbi.nlm.nih.gov/30184186/ |

| | | | | | | | | | | | |
|---|---|---|---|---|---|---|---|---|---|---|---|
| rayyan-751570955 | Marcos-Marcos J and Olry de Labry-Lima A and Toro-Cardenas S and LacasaÃ±a M and Degroote S and Ridde V and Bermudez-Tamayo C | Impact, economic evaluation, and sustainability of integrated vector management in urban settings to prevent vector-borne diseases: a scoping review. | 2018 | Infectious diseases of | 7 | 83 | RAYYAN-INCLUSION: {"Richard"=>"Excluded"} \| RAYYAN-EXCLUSION-REASONS: wrong outcome,wrong study design,wrong publication type \| USER-NOTES: {"Richard"=>["Review"]} | 10.1186/s40249-018-0464-x | Animals;Dengue/*economics/ prevention & control/transmission;Disease | PMC6120095 | https://pubmed.ncbi.nlm.nih.gov/30173675/ |
| rayyan-751570956 | Osorio L and Garcia JA and Parra LG and Garcia V and Torres L and Degroote S and Ridde V | A scoping review on the field validation and implementation of rapid diagnostic tests for vector-borne and other infectious diseases of poverty in urban areas. | 2018 | Infectious diseases of | 7 | 87 | RAYYAN-INCLUSION: {"Richard"=>"Excluded"} \| RAYYAN-EXCLUSION-REASONS: wrong outcome,wrong study design,wrong publication type \| USER-NOTES: {"Richard"=>["Review"]} | 10.1186/s40249-018-0474-8 | Animals;Communicable Diseases/*diagnosis/economic s/transmission;Diagnostic | PMC6120097 | https://pubmed.ncbi.nlm.nih.gov/30173662/ |
| rayyan-751570957 | Hellewell J and Walker P and Ghani A and Rao B and Churcher TS | Using ante-natal clinic prevalence data to monitor temporal changes in malaria incidence in a humanitarian setting in the Democratic Republic of Congo. | 2018 | Malaria journal | 17 | 312 | RAYYAN-INCLUSION: {"Richard"=>"Excluded"} \| RAYYAN-EXCLUSION-REASONS: wrong outcome,wrong study design | 10.1186/s12936-018-2460-9 | Child, Preschool;Democratic Republic of the Congo/epidemiology;Humans;I | PMC6114784 | https://pubmed.ncbi.nlm.nih.gov/30157850/ |
| rayyan-751570958 | Nderu D and Kimani F and Thiong'o K and Akinyi M and Karanja E and Meyer CG and Velavan TP | PfHRP2-PfHRP3 diversity among Kenyan isolates and comparative evaluation of PfHRP2/pLDH malaria RDT with microscopy and nested PCR methodologies. | 2018 | Parasitology internatio | 67 | 793-799 | RAYYAN-INCLUSION: {"Richard"=>"Excluded"} \| RAYYAN-EXCLUSION-REASONS: wrong outcome,wrong study design | 10.1016/j.parint.2018.08.007 | Antigens, Protozoan/*genetics;Cross-Sectional Studies;Diagnostic | | https://pubmed.ncbi.nlm.nih.gov/30138695/ |
| rayyan-751570959 | Dharmawardena P and Premaratne R and Mendis K and Wickemasinghe R and Rodrigo C and Harintheran A and Fernando D | Effectiveness of passive case detection for imported malaria in a hospital setting in Sri Lanka during the prevention of re-introduction phase of malaria. | 2019 | International health | 11 | 64-70 | RAYYAN-INCLUSION: {"Richard"=>"Excluded"} \| RAYYAN-EXCLUSION-REASONS: wrong outcome,wrong study design | 10.1093/inthealth/ihy061 | Adult;Communicable Diseases, Imported/*diagnosis;Fever;Health Knowledge, Attitudes, | | https://pubmed.ncbi.nlm.nih.gov/30137418/ |
| rayyan-751570960 | Desalegn Z and Wegayehu T and Massebo F | Wall-type and indoor residual spraying application quality affect the residual efficacy of indoor residual spray against wild malaria vector in southwest Ethiopia. | 2018 | Malaria journal | 17 | 300 | RAYYAN-INCLUSION: {"Richard"=>"Excluded"} \| RAYYAN-EXCLUSION-REASONS: wrong outcome,wrong study design \| USER-NOTES: {"Richard"=>["Ento study"]} | 10.1186/s12936-018-2458-3 | Aerosols/*administration & dosage;Animals;Anopheles/*drug | PMC6102920 | https://pubmed.ncbi.nlm.nih.gov/30126433/ |
| rayyan-751570961 | Pham NM and Karlen W and Beck HP and Delamarche E | Malaria and the 'last' parasite: how can technology help? | 2018 | Malaria journal | 17 | 260 | RAYYAN-INCLUSION: {"Richard"=>"Excluded"} \| RAYYAN-EXCLUSION-REASONS: wrong outcome,wrong study design,wrong publication type | 10.1186/s12936-018-2408-0 | Diagnostic Tests, Routine/*methods;Disease Eradication/*methods;Human | PMC6042346 | https://pubmed.ncbi.nlm.nih.gov/29996831/ |
| rayyan-751570962 | Kinyoki DK and Moloney GM and Uthman OA and Odundo EO and Kandala NB and Noor AM and Snow RW and Berkley JA | Co-morbidity of malnutrition with falciparum malaria parasitaemia among children under the aged 6-59Â months in Somalia: a geostatistical analysis. | 2018 | Infectious diseases of | 7 | 72 | RAYYAN-INCLUSION: {"Richard"=>"Excluded"} \| RAYYAN-EXCLUSION-REASONS: wrong study design,wrong study duration \| USER-NOTES: {"Richard"=>["Period 2010"]} | 10.1186/s40249-018-0449-9 | Bayes Theorem;Child, Preschool;Comorbidity;Cross-Sectional | PMC6036667 | https://pubmed.ncbi.nlm.nih.gov/29986753/ |
| rayyan-751570963 | Ergete S and Sorsa S and Loha E and Asnake S | Trend of Malaria Cases in Hana and Keyafer Health Centers, South Omo Zone, Southern Ethiopia. | 2018 | Ethiopian journal of health | 28 | 277-286 | RAYYAN-INCLUSION: {"Richard"=>"Excluded"} \| RAYYAN-EXCLUSION-REASONS: wrong outcome,wrong study design \| USER-NOTES: {"Richard"=>["No epi / intervention study"]} | 10.4314/ejhs.v28i3.5 | Adolescent;Adult;Age Factors;Aged;Child;Child, Preschool;Ethiopia/epidemiolo | PMC6016364 | https://pubmed.ncbi.nlm.nih.gov/29983527/ |
| rayyan-751570964 | Esu E and Berens-Riha N and Pritsch M and Nwachuku N and Loescher T and Meremikwu M | Intermittent screening and treatment with artemether-lumefantrine versus intermittent preventive treatment with sulfadoxine-pyrimethamine for malaria in pregnancy: a facility-based, open-label, non-inferiority trial in Nigeria. | 2018 | Malaria journal | 17 | 251 | RAYYAN-INCLUSION: {"Richard"=>"Excluded"} \| RAYYAN-EXCLUSION-REASONS: wrong outcome,wrong study design \| USER-NOTES: {"Richard"=>["Rx study"]} | 10.1186/s12936-018-2394-2 | Adolescent;Adult;Antimalarials /*therapeutic use;Artemether, Lumefantrine Drug | PMC6034215 | https://pubmed.ncbi.nlm.nih.gov/29976228/ |
| rayyan-751570965 | Henriques G and Phommasone K and Tripura R and Peto TJ and Raut S and Snethlage C and Sambo I and Sanann N and Nguon C and Adhikari B and Pongvongsa T and Imwong M and | Comparison of glucose-6 phosphate dehydrogenase status by fluorescent spot test and rapid diagnostic test in Lao PDR and Cambodia. | 2018 | Malaria journal | 17 | 243 | RAYYAN-INCLUSION: {"Richard"=>"Excluded"} \| RAYYAN-EXCLUSION-REASONS: wrong outcome,wrong study design \| USER-NOTES: {"Richard"=>["Diag study"]} | 10.1186/s12936-018-2390-6 | Adolescent;Adult;Aged;Aged, 80 and over;Cambodia;Child;Child, | PMC6013858 | https://pubmed.ncbi.nlm.nih.gov/29929514/ |
| rayyan-751570966 | Dombrowski JG and de Souza RM and Silva NRM and Barateiro A and Epiphanio S and GonÃ§alves LA and Marinho CRF | Malaria during pregnancy and newborn outcome in an unstable transmission area in Brazil: A population-based record linkage study. | 2018 | PloS one | 13 | e0199415 | RAYYAN-INCLUSION: {"Richard"=>"Excluded"} \| RAYYAN-EXCLUSION-REASONS: wrong outcome,wrong study design \| USER-NOTES: {"Richard"=>["Epi study"]} | 10.1371/journal.pone.0199415 | Birth Weight;Brazil/epidemiology;Female;Geography;Humans;Infa | PMC6013245 | https://pubmed.ncbi.nlm.nih.gov/29928025/ |
| rayyan-751570967 | Thriemer K and Bobogare A and Ley B and Gudo CS and Alam MS and Anstey NM and Ashley E and Baird JK and Gryseels C and Jambert E and Lacerda M and Laihad F and Marfurt J and | Quantifying primaquine effectiveness and improving adherence: a round table discussion of the APMEN Vivax Working Group. | 2018 | Malaria journal | 17 | 241 | RAYYAN-INCLUSION: {"Richard"=>"Excluded"} \| RAYYAN-EXCLUSION-REASONS: wrong outcome,wrong study design,wrong publication type | 10.1186/s12936-018-2380-8 | Antimalarials/*therapeutic use;Asia;Humans;Malaria, Vivax/*prevention & | PMC6011582 | https://pubmed.ncbi.nlm.nih.gov/29925430/ |
| rayyan-751570968 | Linn NYY and Kathirvel S and Das M and Thapa B and Rahman MM and Maung TM and Kyaw AMM and Thi A and Lin Z | Are village health volunteers as good as basic health staffs in providing malaria care? A country wide analysis from Myanmar, 2015. | 2018 | Malaria journal | 17 | 242 | RAYYAN-INCLUSION: {"Richard"=>"Maybe"} \| RAYYAN-LABELS: Include (full text) | 10.1186/s12936-018-2384-4 | Cohort Studies;Community Health Workers/organization & administration/*statistics & | PMC6011412 | https://pubmed.ncbi.nlm.nih.gov/29925375/ |
| rayyan-751570969 | Ampofo GD and Tagbor H and Bates I | Effectiveness of pregnant women's active participation in their antenatal care for the control of malaria and anaemia in pregnancy in Ghana: a cluster randomized controlled trial. | 2018 | Malaria journal | 17 | 238 | RAYYAN-INCLUSION: {"Richard"=>"Excluded"} \| RAYYAN-EXCLUSION-REASONS: wrong outcome,wrong study design | 10.1186/s12936-018-2387-1 | Adult;Anemia/*prevention & control;Antimalarials/*administration & dosage;Cluster | PMC6009977 | https://pubmed.ncbi.nlm.nih.gov/29921302/ |
| rayyan-751570970 | Fornace KM and Surendra H and Abidin TR and Reyes R and Macalinao MLM and Stresman G and Luchavez J and Ahmad RA and Supargiyono S and Espino F and Drakeley CJ and Cook | Use of mobile technology-based participatory mapping approaches to geolocate health facility attendees for disease surveillance in low resource settings. | 2018 | International journal of | 17 | 21 | RAYYAN-INCLUSION: {"Richard"=>"Excluded"} \| RAYYAN-EXCLUSION-REASONS: wrong outcome,wrong study design | 10.1186/s12942-018-0141-0 | *Computers, Handheld;*Geographic Information Systems/statistics | PMC6006992 | https://pubmed.ncbi.nlm.nih.gov/29914506/ |
| rayyan-751570971 | Al Manir MS and Brenas JH and Baker CJ and Shaban-Nejad A | A Surveillance Infrastructure for Malaria Analytics: Provisioning Data Access and Preservation of Interoperability. | 2018 | JMIR public health | 4 | e10218 | RAYYAN-INCLUSION: {"Richard"=>"Excluded"} \| RAYYAN-EXCLUSION-REASONS: wrong outcome,wrong study design \| USER-NOTES: {"Richard"=>["Strat / surveillance platform"]} | 10.2196/10218 | Malaria | PMC6026300 | https://pubmed.ncbi.nlm.nih.gov/29907554/ |
| rayyan-751570972 | Dalrymple U and Arambepola R and Gething PW and Cameron E | How long do rapid diagnostic tests remain positive after anti-malarial treatment? | 2018 | Malaria journal | 17 | 228 | RAYYAN-INCLUSION: {"Richard"=>"Excluded"} \| RAYYAN-EXCLUSION-REASONS: wrong outcome,wrong study design | 10.1186/s12936-018-2371-9 | Antigens, Protozoan/*blood/classification;Antimalarials/*administratio | PMC5994115 | https://pubmed.ncbi.nlm.nih.gov/29884184/ |
| rayyan-751570973 | Kheang ST and Lin MA and Lwin S and Naing YH and Yarzar P and Kak N and Price T | Malaria Case Detection Among Mobile Populations and Migrant Workers in Myanmar: Comparison of 3 Service Delivery Approaches. | 2018 | Global health, science | 6 | 384-389 | RAYYAN-INCLUSION: {"Richard"=>"Maybe"} \| RAYYAN-LABELS: Excluded (after full text) | 10.9745/GHSP-D-17-00318 | Delivery of Health Care/*methods;Health Services | PMC6024619 | https://pubmed.ncbi.nlm.nih.gov/29875157/ |
| rayyan-751570975 | Druetz T | Evaluation of direct and indirect effects of seasonal malaria chemoprevention in Mali. | 2018 | Scientific reports | 8 | 8104 | RAYYAN-INCLUSION: {"Richard"=>"Excluded"} \| RAYYAN-EXCLUSION-REASONS: wrong outcome,wrong study design | 10.1038/s41598-018-26474-6 | Anemia/complications;Antimalarials/pharmacology;Chemoprevention/*statistics & | PMC5970148 | https://pubmed.ncbi.nlm.nih.gov/29802375/ |
| rayyan-751570976 | Ugele M and Weniger M and Leidenberger M and Huang Y and Bassler M and Friedrich O and Kappes B and Hayden O and Richter L | Label-free, high-throughput detection of P. falciparum infection in sphered erythrocytes with digital holographic microscopy. | 2018 | Lab on a chip | 18 | 1704-1712 | RAYYAN-INCLUSION: {"Richard"=>"Excluded"} \| RAYYAN-EXCLUSION-REASONS: wrong outcome,wrong study design | 10.1039/c8lc00350e | Erythrocytes/*parasitology;Holography/*methods;Humans;Life Cycle | | https://pubmed.ncbi.nlm.nih.gov/29796511/ |
| rayyan-751570977 | Kang SY and Battle KE and Gibson HS and Ratsimbasoa A and Randrianarivelojosia M and Ramboarina S and Zimmerman PA and Weiss DJ and Cameron E and Gething PW and Howes RE | Spatio-temporal mapping of Madagascar's Malaria Indicator Survey results to assess Plasmodium falciparum endemicity trends between 2011 and 2016. | 2018 | BMC medicine | 16 | 71 | RAYYAN-INCLUSION: {"Richard"=>"Excluded"} \| RAYYAN-EXCLUSION-REASONS: wrong outcome,wrong study design \| USER-NOTES: {"Richard"=>["No routine data"]} | 10.1186/s12916-018-1060-4 | Child, Preschool;Female;History, 21st Century;Humans;Infant;Madag | PMC5964908 | https://pubmed.ncbi.nlm.nih.gov/29788968/ |
| rayyan-751570978 | Gallay J and Mosha D and Lutahakana E and Mazuguni F and Zuakulu M and Decosterd LA and Genton B and Pothin E | Appropriateness of malaria diagnosis and treatment for fever episodes according to patient history and anti-malarial blood measurement: a cross-sectional survey from Tanzania. | 2018 | Malaria journal | 17 | 209 | RAYYAN-INCLUSION: {"Richard"=>"Excluded"} \| RAYYAN-EXCLUSION-REASONS: wrong outcome,wrong study design \| USER-NOTES: {"Richard"=>["DIag study"]} | 10.1186/s12936-018-2357-7 | Adolescent;Adult;Aged;Aged, 80 and over;Antimalarials/*blood;Cas | PMC5963060 | https://pubmed.ncbi.nlm.nih.gov/29784001/ |
| rayyan-751570979 | Boyce MR and Menya D and Turner EL and Laktabai J and Prudhomme-O'Meara W | Evaluation of malaria rapid diagnostic test (RDT) use by community health workers: a longitudinal study in western Kenya. | 2018 | Malaria journal | 17 | 206 | RAYYAN-INCLUSION: {"Richard"=>"Excluded"} \| RAYYAN-EXCLUSION-REASONS: wrong outcome,wrong study design | 10.1186/s12936-018-2358-6 | Adult;Age Factors;Aged;Clinical Competence/*statistics & | PMC5960182 | https://pubmed.ncbi.nlm.nih.gov/29776359/ |
| rayyan-751570980 | Pulford J and Kurumop S and Mueller I and Siba PM and Hetzel MW | The impact of the scale-up of malaria rapid diagnostic tests on the routine clinical diagnosis procedures for febrile illness: a series of repeated cross-sectional studies in Papua New Guinea. | 2018 | Malaria journal | 17 | 202 | RAYYAN-INCLUSION: {"Richard"=>"Excluded"} \| RAYYAN-EXCLUSION-REASONS: wrong outcome,wrong study design | 10.1186/s12936-018-2351-0 | Case Management/statistics & numerical data;Cross-Sectional Studies;Diagnostic Tests, | PMC5956836 | https://pubmed.ncbi.nlm.nih.gov/29769128/ |
| rayyan-751570981 | Masaninga F and Mukumbuta N and Ndhlovu K and Hamainza B and Wamulume P and Chanda E and Banda J and Mwanza-Ingwe M and Miller JM and Ameneshewa B and Mnzava A and | Insecticide-treated nets mass distribution campaign: benefits and lessons in Zambia. | 2018 | Malaria journal | 17 | 173 | RAYYAN-INCLUSION: {"Richard"=>"Excluded"} \| RAYYAN-EXCLUSION-REASONS: wrong outcome,wrong study design | 10.1186/s12936-018-2314-5 | Family Characteristics;Humans;Insecticide-Treated | PMC5916719 | https://pubmed.ncbi.nlm.nih.gov/29690873/ |
| rayyan-751570982 | Kibret S and Wilson GG and Ryder D and Tekie H and Petros B | Can water-level management reduce malaria mosquito abundance around large dams in sub-Saharan Africa? | 2018 | PloS one | 13 | e0196064 | RAYYAN-INCLUSION: {"Richard"=>"Excluded"} \| RAYYAN-EXCLUSION-REASONS: wrong outcome,wrong study design \| USER-NOTES: {"Richard"=>["Ento study"]} | 10.1371/journal.pone.0196064 | Africa South of the Sahara/epidemiology;Animals;Anopheles;Disease | PMC5909510 | https://pubmed.ncbi.nlm.nih.gov/29672560/ |
| rayyan-751570983 | Ssempiira J and Kissa J and Nambuusi B and Kyozira C and Rutazaana D and Mukooyo E and Opigo J and Makumbi F and Kasasa S and Vounatsou P | The effect of case management and vector-control interventions on space-time patterns of malaria incidence in Uganda. | 2018 | Malaria journal | 17 | 162 | RAYYAN-INCLUSION: {"Richard"=>"Included"} | 10.1186/s12936-018-2312-7 | Adolescent;Adult;Aged;Aged, 80 and over;Antimalarials/*therapeuti | PMC5898071 | https://pubmed.ncbi.nlm.nih.gov/29650005/ |
| rayyan-751570984 | Douine M and Sanna A and Galindo M and Musset L and Pommier de Santi V and Marchesini P and Magalhaes ED and Suarez-Mutis M and Hiwat H and Nacher M and Vreden S and | Malakit: an innovative pilot project to self-diagnose and self-treat malaria among illegal gold miners in the Guiana Shield. | 2018 | Malaria journal | 17 | 158 | RAYYAN-INCLUSION: {"Richard"=>"Excluded"} \| RAYYAN-EXCLUSION-REASONS: wrong outcome,wrong study design | 10.1186/s12936-018-2306-5 | Diagnostic Tests, Routine/instrumentation/*methods;French | PMC5892004 | https://pubmed.ncbi.nlm.nih.gov/29631588/ |
| rayyan-751570985 | Maung TM and Oo T and Wai KT and Hlaing T and Owiti P and Kumar B and Shewade HD and Zachariah R and Thi A | Assessment of household ownership of bed nets in areas with and without artemisinin resistance containment measures in Myanmar. | 2018 | Infectious diseases of | 7 | 19 | RAYYAN-INCLUSION: {"Richard"=>"Excluded"} \| RAYYAN-EXCLUSION-REASONS: wrong outcome,wrong study design \| USER-NOTES: {"Richard"=>["No routine data"]} | 10.1186/s40249-018-0399-2 | Adolescent;Adult;Antimalarials /pharmacology/*therapeutic use;Artemisinins/pharmacolog | PMC5865351 | https://pubmed.ncbi.nlm.nih.gov/29571301/ |
| rayyan-751570986 | Hamid MMA and Thriemer K and Elobied ME and Mahgoub NS and Boshara SA and Elsafi HMH and Gumaa SA and Hamid T and Abdelbagi H and Basheir HM and Marfurt J and Chen I | Low risk of recurrence following artesunate-Sulphadoxine-pyrimethamine plus primaquine for uncomplicated Plasmodium falciparum and Plasmodium vivax infections in the Republic of the Sudan. | 2018 | Malaria journal | 17 | 117 | RAYYAN-INCLUSION: {"Richard"=>"Excluded"} \| RAYYAN-EXCLUSION-REASONS: wrong outcome,wrong study design | 10.1186/s12936-018-2266-9 | Adolescent;Adult;Antimalarials /administration & dosage/therapeutic | PMC5857106 | https://pubmed.ncbi.nlm.nih.gov/29548285/ |
| rayyan-751570987 | Arnaldo P and Rovira-Vallbona E and Langa JS and Salvador C and Guetens P and Chiheb D and Xavier B and Kestens L and Enosse SM and Rosanas-Urgell A | Uptake of intermittent preventive treatment and pregnancy outcomes: health facilities and community surveys in ChÃ³kwÃ¨ district, southern Mozambique. | 2018 | Malaria journal | 17 | 109 | RAYYAN-INCLUSION: {"Richard"=>"Excluded"} \| RAYYAN-EXCLUSION-REASONS: wrong outcome,wrong study design \| USER-NOTES: {"Richard"=>["No routine data"]} | 10.1186/s12936-018-2255-z | Antimalarials/administration & dosage/*therapeutic use;Drug Combinations;Female;*Health | PMC5848514 | https://pubmed.ncbi.nlm.nih.gov/29530044/ |

| | | | | | | | | | | | |
|---|---|---|---|---|---|---|---|---|---|---|---|
| rayyan-751570988 | Riley C and Dellicour S and Ouma P and Kioko U and Omar A and Kariuki S and Ng'ang'a Z and Desai M and Buff AM and Gutman JR | Knowledge and Adherence to the National Guidelines for Malaria Diagnosis in Pregnancy among Health-Care Providers and Drug-Outlet Dispensers in Rural Western Kenya. | 2018 | The American journal of | 98 | 1367-1373 | RAYYAN-INCLUSION: {"Richard"=>"Excluded"} \| RAYYAN-EXCLUSION-REASONS: wrong outcome,wrong study design | 10.4269/ajtmh.17-0594 | Adult;Antimalarials/administration & dosage/*therapeutic use;Diagnostic Tests, | PMC5953364 | https://pubmed.ncbi.nlm.nih.gov/29512480/ |
| rayyan-751570990 | Winn LK and Lesser A and Menya D and Baumgartner JN and Kipkoech Kirui J and Saran I and Prudhomme-O'Meara W | Motivation and satisfaction among community health workers administering rapid diagnostic tests for malaria in Western Kenya. | 2018 | Journal of global health | 8 | 010401 | RAYYAN-INCLUSION: {"Richard"=>"Excluded"} \| RAYYAN-EXCLUSION-REASONS: wrong outcome,wrong study design | 10.7189/jogh.08.010401 | Adult;Aged;*Attitude of Health Personnel;Case Management;Community | PMC5823030 | https://pubmed.ncbi.nlm.nih.gov/29497500/ |
| rayyan-751570991 | Kolie D and Camara BS and Delamou A and BÃ©avogui AH and Hermans V and Edwards JK and Benedetti G and Muller CP and Griensven JV and Zachariah R | The Ebola-effect in Guinea 2014-15: Tangled trends of malaria care in children under-five. | 2018 | PloS one | 13 | e0192798 | RAYYAN-INCLUSION: {"Richard"=>"Excluded"} \| RAYYAN-EXCLUSION-REASONS: wrong outcome,wrong study design | 10.1371/journal.pone.0192798 | Administration, Oral;Antimalarials/administration & dosage/therapeutic | PMC5830300 | https://pubmed.ncbi.nlm.nih.gov/29489836/ |
| rayyan-751570992 | Wagenaar BH and Augusto O and Beste J and Toomay SJ and Wickett E and Dunbar N and Bawo L and Wesseh CS | The 2014-2015 Ebola virus disease outbreak and primary healthcare delivery in Liberia: Time-series analyses for 2010-2016. | 2018 | PLoS medicine | 15 | e1002508 | RAYYAN-INCLUSION: {"Richard"=>"Excluded"} \| RAYYAN-EXCLUSION-REASONS: wrong outcome,wrong study design,wrong infection / disease | 10.1371/journal.pmed.1002508 | Adolescent;Adult;Delivery of Health Care/organization & administration/standards/*sta | PMC5819774 | https://pubmed.ncbi.nlm.nih.gov/29462138/ |
| rayyan-751570993 | Talundzic E and Ravishankar S and Kelley J and Patel D and Plucinski M and Schmedes S and Ljolje D and Clemons B and Madison-Antenucci S and Arguin PM and Lucchi NW and | Next-Generation Sequencing and Bioinformatics Protocol for Malaria Drug Resistance Marker Surveillance. | 2018 | Antimicrobial agents | 62 | | RAYYAN-INCLUSION: {"Richard"=>"Excluded"} \| RAYYAN-EXCLUSION-REASONS: wrong outcome,wrong study design \| USER-NOTES: {"Richard"=>["Genomic surveillance"]} | 10.1128/AAC.02474-17 | Antimalarials/*pharmacology; Computational Biology/*methods;Drug | PMC5913988 | https://pubmed.ncbi.nlm.nih.gov/29439965/ |
| rayyan-751570994 | D'Alessandro U and Hill J and Tarning J and Pell C and Webster J and Gutman J and Sevene E | Treatment of uncomplicated and severe malaria during pregnancy. | 2018 | The Lancet. Infectious | 18 | e133-e146 | RAYYAN-INCLUSION: {"Richard"=>"Excluded"} \| RAYYAN-EXCLUSION-REASONS: wrong outcome,wrong study design | 10.1016/S1473-3099(18)30065-3 | Antimalarials/adverse effects/pharmacokinetics/*therapeutic | PMC6590069 | https://pubmed.ncbi.nlm.nih.gov/29395998/ |
| rayyan-751570995 | Awine T and Malm K and Peprah NY and Silal SP | Spatio-temporal heterogeneity of malaria morbidity in Ghana: Analysis of routine health facility data. | 2018 | PloS one | 13 | e0191707 | RAYYAN-INCLUSION: {"Richard"=>"Excluded"} \| RAYYAN-EXCLUSION-REASONS: wrong outcome \| USER-NOTES: {"Richard"=>["Epi / strat study"]} | 10.1371/journal.pone.0191707 | Climate;Ghana/epidemiology;*Health Facilities;Humans;Malaria/*epi | PMC5788359 | https://pubmed.ncbi.nlm.nih.gov/29377908/ |
| rayyan-751570996 | Varo R and Crowley VM and Sitoe A and Madrid L and Serghides L and Kain KC and Bassat Q | Adjunctive therapy for severe malaria: a review and critical appraisal. | 2018 | Malaria journal | 17 | 47 | RAYYAN-INCLUSION: {"Richard"=>"Excluded"} \| RAYYAN-EXCLUSION-REASONS: wrong outcome,wrong study design,wrong publication type \| USER-NOTES: {"Richard"=>["Review"]} | 10.1186/s12936-018-2195-7 | Adolescent;Adult;Aged;Aged, 80 and over;Animals;Antimalarials/*th | PMC5781278 | https://pubmed.ncbi.nlm.nih.gov/29361945/ |
| rayyan-751570997 | Wagman J and Gogue C and Tynuv K and Mihigo J and Bankineza E and Bah M and Diallo D and Saibu A and Richardson JH and Kone D and Fomba S and Bernson J and | An observational analysis of the impact of indoor residual spraying with non-pyrethroid insecticides on the incidence of malaria in SÃ©gou Region, Mali: 2012-2015. | 2018 | Malaria journal | 17 | 19 | RAYYAN-INCLUSION: {"Richard"=>"Included"} | 10.1186/s12936-017-2168-2 | Adolescent;Adult;Aged;Animals;Child;Child, Preschool;Disease | PMC5761159 | https://pubmed.ncbi.nlm.nih.gov/29316917/ |
| rayyan-751570998 | Morang'a C and Ayieko C and Awinda G and Achilla R and Moseti C and Ogutu B and Waitumbi J and Wanja E | Stabilization of RDT target antigens present in dried Plasmodium falciparum-infected samples for validating malaria rapid diagnostic tests at the point of care. | 2018 | Malaria journal | 17 | 10 | RAYYAN-INCLUSION: {"Richard"=>"Excluded"} \| RAYYAN-EXCLUSION-REASONS: wrong outcome,wrong study design | 10.1186/s12936-017-2155-7 | Antigens, Protozoan/immunology;Chromatography, | PMC5759799 | https://pubmed.ncbi.nlm.nih.gov/29310651/ |
| rayyan-751570999 | Leong FJ and Jain JP and Feng Y and Goswami B and Stein DS | A phase 1 evaluation of the pharmacokinetic/pharmacodynamic interaction of the anti-malarial agents KAF156 and piperaquine. | 2018 | Malaria journal | 17 | 7 | RAYYAN-INCLUSION: {"Richard"=>"Excluded"} \| RAYYAN-EXCLUSION-REASONS: wrong outcome,wrong study design \| USER-NOTES: {"Richard"=>["Rx study"]} | 10.1186/s12936-017-2162-8 | Adolescent;Adult;Antimalarials/administration & dosage/adverse | PMC5756412 | https://pubmed.ncbi.nlm.nih.gov/29304859/ |
| rayyan-751571000 | Jones C and Talisuna AO and Snow RW and Zurovac D | "We were being treated like the Queen": understanding trial factors influencing high paediatric malaria treatment adherence in western Kenya. | 2018 | Malaria journal | 17 | 8 | RAYYAN-INCLUSION: {"Richard"=>"Excluded"} \| RAYYAN-EXCLUSION-REASONS: wrong outcome,wrong study design | 10.1186/s12936-017-2164-6 | Adult;Aged;Antiviral Agents/*therapeutic use;Child, Preschool;Female;Humans;Inf | PMC5756360 | https://pubmed.ncbi.nlm.nih.gov/29304798/ |
| rayyan-751571001 | Janet S and Carrara VI and Simpson JA and Thin NWW and Say WW and Paw NTM and Chotivanich K and Turner C and Crawley J and McGready R | Early neonatal mortality and neurological outcomes of neonatal resuscitation in a resource-limited setting on the Thailand-Myanmar border: A descriptive study. | 2018 | PloS one | 13 | e0190419 | RAYYAN-INCLUSION: {"Richard"=>"Excluded"} \| RAYYAN-EXCLUSION-REASONS: wrong outcome,wrong study design,wrong infection / disease | 10.1371/journal.pone.0190419 | Cohort Studies;*Health Care Rationing;Humans;Infant;*Infant Mortality;Infant, | PMC5755780 | https://pubmed.ncbi.nlm.nih.gov/29304139/ |
| rayyan-751571002 | Druetz T and Corneau-Tremblay N and Millogo T and Kouanda S and Ly A and Bicaba A and Haddad S | Impact Evaluation of Seasonal Malaria Chemoprevention under Routine Program Implementation: A Quasi-Experimental Study in Burkina Faso. | 2018 | The American journal of | 98 | 524-533 | RAYYAN-INCLUSION: {"Richard"=>"Maybe"} \| RAYYAN-LABELS: Excluded (after full text) | 10.4269/ajtmh.17-0599 | Adolescent;Adult;Antimalarials/*administration & dosage/pharmacology/therape | PMC5929206 | https://pubmed.ncbi.nlm.nih.gov/29260654/ |
| rayyan-751571003 | Hansen KS and Clarke SE and Lal S and Magnussen P and Mbonye AK | Cost-effectiveness analysis of introducing malaria diagnostic testing in drug shops: A cluster-randomised trial in Uganda. | 2017 | PloS one | 12 | e0189758 | RAYYAN-INCLUSION: {"Richard"=>"Excluded"} \| RAYYAN-EXCLUSION-REASONS: wrong outcome,wrong study design \| USER-NOTES: {"Richard"=>["Costing study"]} | 10.1371/journal.pone.0189758 | Antimalarials/*standards/therapeutic use;Artemisinins/*standards/t | PMC5731679 | https://pubmed.ncbi.nlm.nih.gov/29244829/ |
| rayyan-751571004 | Bakr S and Edris S and Abdel Fattah NS and Ibrahim NM and El-Khadragy MF | Molecular Screening for Malaria among Blood Donors in a WHO Claimed Region of Egypt, Fayoum Governorate. | 2017 | Mediterranean journal of | 9 | e2017065 | RAYYAN-INCLUSION: {"Richard"=>"Excluded"} \| RAYYAN-EXCLUSION-REASONS: wrong outcome,wrong study design \| USER-NOTES: {"Richard"=>["Epi study"]} | 10.4084/MJHID.2017.065 | Egypt;Blood Donors;Malaria;Mass Screening | PMC5667524 | https://pubmed.ncbi.nlm.nih.gov/29181142/ |
| rayyan-751571005 | Brabin L and Roberts SA and Gies S and Nelson A and Diallo S and Stewart CJ and Kazienga A and Birtles J and Ouedraogo S and Claeys Y and Tinto H and d'Alessandro U and Faragher EB | Effects of long-term weekly iron and folic acid supplementation on lower genital tract infection - a double blind, randomised controlled trial in Burkina Faso. | 2017 | BMC medicine | 15 | 206 | RAYYAN-INCLUSION: {"Richard"=>"Excluded"} \| RAYYAN-EXCLUSION-REASONS: wrong outcome,wrong study design,wrong infection / disease | 10.1186/s12916-017-0967-5 | Adolescent;Anemia/prevention & control;Burkina Faso;Dietary | PMC5700548 | https://pubmed.ncbi.nlm.nih.gov/29166928/ |
| rayyan-751571006 | Kanyangarara M and Chou VB | Linking household surveys and health facility assessments to estimate intervention coverage for the Lives Saved Tool (LiST). | 2017 | BMC public health | 17 | 780 | RAYYAN-INCLUSION: {"Richard"=>"Excluded"} \| RAYYAN-EXCLUSION-REASONS: wrong outcome,wrong study design,wrong infection / disease | 10.1186/s12889-017-4743-4 | Computer Simulation;Family Characteristics;Female;*Health Care Surveys;Health | PMC5688485 | https://pubmed.ncbi.nlm.nih.gov/29143639/ |
| rayyan-751571007 | Kamau A and Nyaga V and Bauni E and Tsofa B and Noor AM and Bejon P and Scott JAG and Hammitt LL | Trends in bednet ownership and usage, and the effect of bednets on malaria hospitalization in the Kilifi Health and Demographic Surveillance System (KHDSS): 2008-2015. | 2017 | BMC infectious diseases | 17 | 720 | RAYYAN-INCLUSION: {"Richard"=>"Maybe"} \| RAYYAN-LABELS: Include (full text) | 10.1186/s12879-017-2822-x | Adolescent;Child;Child, Preschool;Female;Health Surveys;Hospitalization;Huma | PMC5688631 | https://pubmed.ncbi.nlm.nih.gov/29141606/ |
| rayyan-751571008 | Oyet C and Roh ME and Kiwanuka GN and Orikiriza P and Wade M and Parikh S and Mwanga-Amumpaire J and Boum Y 2nd | Evaluation of the Deki Readerâ„¢, an automated RDT reader and data management device, in a household survey setting in low malaria endemic southwestern Uganda. | 2017 | Malaria journal | 16 | 449 | RAYYAN-INCLUSION: {"Richard"=>"Excluded"} \| RAYYAN-EXCLUSION-REASONS: wrong outcome,wrong study design | 10.1186/s12936-017-2094-3 | Child, Preschool;Cross-Sectional Studies;Diagnostic Tests, | PMC5678817 | https://pubmed.ncbi.nlm.nih.gov/29115991/ |
| rayyan-751571009 | Burnett JL and Carns JL and Richards-Kortum R | Towards a needle-free diagnosis of malaria: in vivo identification and classification of red and white blood cells containing haemozoin. | 2017 | Malaria journal | 16 | 447 | RAYYAN-INCLUSION: {"Richard"=>"Excluded"} \| RAYYAN-EXCLUSION-REASONS: wrong outcome,wrong study design | 10.1186/s12936-017-2096-1 | Diagnostic Tests, Routine/*methods;Erythrocytes/*parasitology;Hemeprotein | PMC5678583 | https://pubmed.ncbi.nlm.nih.gov/29115957/ |
| rayyan-751571010 | Bridges DJ and Miller JM and Chalwe V and Moonga H and Hamainza B and Steketee R and Silumbe K and Nyangu J and Larsen DA | Community-led Responses for Elimination (CoRE): a study protocol for a community randomized controlled trial assessing the effectiveness of community-level, reactive focal drug administration for reducing Plasmodium falciparum infection prevalence and incidence in Southern Province, Zambia. | 2017 | Trials | 18 | 511 | RAYYAN-INCLUSION: {"Richard"=>"Excluded"} \| RAYYAN-EXCLUSION-REASONS: wrong publication type \| USER-NOTES: {"Richard"=>["Protocol"]} | 10.1186/s13063-017-2249-0 | Adolescent;Adult;Antimalarials/*administration & dosage/adverse | PMC5667476 | https://pubmed.ncbi.nlm.nih.gov/29096671/ |
| rayyan-751571011 | Mfuh KO and Tassi Yunga S and Esemu LF and Bekindaka ON and Yonga J and Djontu JC and Mbakop CD and Taylor DW and Nerurkar VR and Leke RGF | Detection of Plasmodium falciparum DNA in saliva samples stored at room temperature: potential for a non-invasive saliva-based diagnostic test for malaria. | 2017 | Malaria journal | 16 | 434 | RAYYAN-INCLUSION: {"Richard"=>"Excluded"} \| RAYYAN-EXCLUSION-REASONS: wrong outcome,wrong study design \| USER-NOTES: {"Richard"=>["Diag study"]} | 10.1186/s12936-017-2084-5 | Adolescent;Adult;Aged;Cameroon/epidemiology;Child;Child, Preschool;Diagnostic Tests, | PMC5658920 | https://pubmed.ncbi.nlm.nih.gov/29078786/ |
| rayyan-751571012 | Kitutu FE and Kalyango JN and Mayora C and Selling KE and Peterson S and Wamani H | Integrated community case management by drug sellers influences appropriate treatment of paediatric febrile illness in South Western Uganda: a quasi-experimental study. | 2017 | Malaria journal | 16 | 425 | RAYYAN-INCLUSION: {"Richard"=>"Excluded"} \| RAYYAN-EXCLUSION-REASONS: wrong outcome,wrong study design | 10.1186/s12936-017-2072-9 | Antidiarrheals/administration & dosage;Antimalarials/administ | PMC5654057 | https://pubmed.ncbi.nlm.nih.gov/29061148/ |
| rayyan-751571014 | Junaid QO and Khaw LT and Mahmud R and Ong KC and Lau YL and Borade PU and Liew JWK and Sivanandam S and Wong KT and Vythilingam I | Pathogenesis of Plasmodium berghei ANKA infection in the gerbil (Meriones unguiculatus) as an experimental model for severe malaria. | 2017 | Parasite (Paris, France) | 24 | 38 | RAYYAN-INCLUSION: {"Richard"=>"Excluded"} \| RAYYAN-EXCLUSION-REASONS: wrong outcome,wrong study design \| USER-NOTES: {"Richard"=>["Experimental model"]} | 10.1051/parasite/2017040 | Animals;Body Temperature;Body Weight;Brain/immunology/pat | PMC5642054 | https://pubmed.ncbi.nlm.nih.gov/29034874/ |
| rayyan-751571015 | Okami S and Kohtake N | Spatiotemporal Modeling for Fine-Scale Maps of Regional Malaria Endemicity and Its Implications for Transitional Complexities in a Routine Surveillance Network in Western Cambodia. | 2017 | Frontiers in public health | 5 | 262 | RAYYAN-INCLUSION: {"Richard"=>"Excluded"} \| RAYYAN-EXCLUSION-REASONS: wrong outcome,wrong study design \| USER-NOTES: {"Richard"=>["Epi / strat study"]} | 10.3389/fpubh.2017.00262 | Malaria | PMC5627027 | https://pubmed.ncbi.nlm.nih.gov/29034229/ |
| rayyan-751571017 | Ruckstuhl L and Lengeler C and Moyen JM and Garro H and Allan R | Malaria case management by community health workers in the Central African Republic from 2009-2014: overcoming challenges of access and instability due to conflict. | 2017 | Malaria journal | 16 | 388 | RAYYAN-INCLUSION: {"Richard"=>"Excluded"} \| RAYYAN-EXCLUSION-REASONS: wrong outcome,wrong study design | 10.1186/s12936-017-2005-7 | Case Management/organization & administration/*statistics & | PMC5622524 | https://pubmed.ncbi.nlm.nih.gov/28962622/ |
| rayyan-751571018 | Hooft AM and Ripp K and Ndenga B and Mutuku F and Vu D and Baltzell K and Masese LN and Vulule J and Mukoko D and LaBeaud AD | Principles, practices and knowledge of clinicians when assessing febrile children: a qualitative study in Kenya. | 2017 | Malaria journal | 16 | 381 | RAYYAN-INCLUSION: {"Richard"=>"Excluded"} \| RAYYAN-EXCLUSION-REASONS: wrong outcome,wrong study design | 10.1186/s12936-017-2021-7 | Adult;Child;Child, Preschool;Clinical Competence/*statistics & | PMC5607512 | https://pubmed.ncbi.nlm.nih.gov/28931399/ |
| rayyan-751571019 | Krampa FD and Aniweh Y and Awandare GA and Kanyong P | Recent Progress in the Development of Diagnostic Tests for Malaria. | 2017 | Diagnostics (Basel, Switzerla | 7 | | RAYYAN-INCLUSION: {"Richard"=>"Excluded"} \| RAYYAN-EXCLUSION-REASONS: wrong outcome,wrong study design,wrong publication type \| USER-NOTES: {"Richard"=>["Review / opinion piece"]} | 10.3390/diagnostics7030054 | Malaria | PMC5617953 | https://pubmed.ncbi.nlm.nih.gov/28925968/ |
| rayyan-751571021 | Pantoja T and Opiyo N and Lewin S and Paulsen E and Ciapponi A and Wiysonge CS and Herrera CA and Rada G and PeÃ±aloza B and Dudley L and Gagnon MP and Garcia Marti S | Implementation strategies for health systems in low-income countries: an overview of systematic reviews. | 2017 | The Cochrane database | 9 | CD011086 | RAYYAN-INCLUSION: {"Richard"=>"Excluded"} \| RAYYAN-EXCLUSION-REASONS: wrong outcome,wrong study design,wrong publication type \| USER-NOTES: {"Richard"=>["Review"]} | 10.1002/14651858.CD011086.pub2 | *Developing Countries;Evidence-Based Practice;Health | PMC5621088 | https://pubmed.ncbi.nlm.nih.gov/28895659/ |
| rayyan-751571022 | Blessborn D and Kaewkhao K and Song L and White NJ and Day NPJ and Tarning J | Quantification of the antimalarial drug pyronaridine in whole blood using LC-MS/MS - Increased sensitivity resulting from reduced non-specific binding. | 2017 | Journal of pharmaceutical and | 146 | 214-219 | RAYYAN-INCLUSION: {"Richard"=>"Excluded"} \| RAYYAN-EXCLUSION-REASONS: wrong outcome,wrong study design \| USER-NOTES: {"Richard"=>["Rx study"]} | 10.1016/j.jpba.2017.08.023 | Antimalarials/*blood/*chemistry/therapeutic use;Artemisinins/chemistry;Ch | PMC5637160 | https://pubmed.ncbi.nlm.nih.gov/28886522/ |
| rayyan-751571023 | Ladner J and Davis B and Audureau E and Saba J | Treatment-seeking patterns for malaria in pharmacies in five sub-Saharan African countries. | 2017 | Malaria journal | 16 | 353 | RAYYAN-INCLUSION: {"Richard"=>"Excluded"} \| RAYYAN-EXCLUSION-REASONS: wrong outcome,wrong study design | 10.1186/s12936-017-1997-3 | Adolescent;Adult;Africa South of the Sahara;Antimalarials/*econom | PMC5574241 | https://pubmed.ncbi.nlm.nih.gov/28851358/ |

| | | | | | | | | | | | |
|---|---|---|---|---|---|---|---|---|---|---|---|
| rayyan-751571024 | Douglas NM and Poespoprodjo JR and Patriani D and Malloy MJ and Kenangalem E and Sugiarto P and Simpson JA and Soenarto Y and Anstey NM and Price RN | Unsupervised primaquine for the treatment of Plasmodium vivax malaria relapses in southern Papua: A hospital-based cohort study. | 2017 | PLoS medicine | 14 | e1002379 | RAYYAN-INCLUSION: {"Richard"=>"Excluded"} \| RAYYAN-EXCLUSION-REASONS: wrong outcome,wrong study design | 10.1371/journal.pmed.1002379 | Adolescent;Adult;Aged;Aged, 80 and over;Antimalarials/*therapeuti | PMC5574534 | https://pubmed.ncbi.nlm.nih.gov/28850568/ |
| rayyan-751571025 | Gibson LE and Markwalter CF and Kimmel DW and Mudenda L and Mbambara S and Thuma PE and Wright DW | Plasmodium falciparum HRP2 ELISA for analysis of dried blood spot samples in rural Zambia. | 2017 | Malaria journal | 16 | 350 | RAYYAN-INCLUSION: {"Richard"=>"Excluded"} \| RAYYAN-EXCLUSION-REASONS: wrong outcome,wrong study design \| USER-NOTES: {"Richard"=>["Diag study"]} | 10.1186/s12936-017-1996-4 | Antigens, Protozoan/*blood;Artemisinins/therapeutic | PMC5569460 | https://pubmed.ncbi.nlm.nih.gov/28835253/ |
| rayyan-751571026 | Cotter C and Sudathip P and Herdiana H and Cao Y and Liu Y and Luo A and Ranasinghe N and Bennett A and Cao J and Gosling RD | Piloting a programme tool to evaluate malaria case investigation and reactive case detection activities: results from 3 settings in the Asia Pacific. | 2017 | Malaria journal | 16 | 347 | RAYYAN-INCLUSION: {"Richard"=>"Excluded"} \| RAYYAN-EXCLUSION-REASONS: wrong outcome,wrong study design | 10.1186/s12936-017-1991-9 | China/epidemiology;*Epidemiological Monitoring;Humans;Indonesia | PMC5568298 | https://pubmed.ncbi.nlm.nih.gov/28830519/ |
| rayyan-751571027 | Saran I and Maffioli EM and Menya D and O'Meara WP | Household beliefs about malaria testing and treatment in Western Kenya: the role of health worker adherence to malaria test results. | 2017 | Malaria journal | 16 | 349 | RAYYAN-INCLUSION: {"Richard"=>"Excluded"} \| RAYYAN-EXCLUSION-REASONS: wrong outcome,wrong study design \| USER-NOTES: {"Richard"=>["Qual study"]} | 10.1186/s12936-017-1993-7 | Adult;*Attitude of Health Personnel;Case Management/*statistics & | PMC5568326 | https://pubmed.ncbi.nlm.nih.gov/28830439/ |
| rayyan-751571028 | Bruxvoort KJ and Leurent B and Chandler CIR and Ansah EK and Baiden F and BjÃ¶rkman A and Burchett HED and Clarke SE and Cundill B and DiLiberto DD and Elfving K and Goodman | The Impact of Introducing Malaria Rapid Diagnostic Tests on Fever Case Management: A Synthesis of Ten Studies from the ACT Consortium. | 2017 | The American journal of | 97 | 1170-1179 | RAYYAN-INCLUSION: {"Richard"=>"Excluded"} \| RAYYAN-EXCLUSION-REASONS: wrong outcome,wrong study design \| USER-NOTES: {"Richard"=>["No epi study"]} | 10.4269/ajtmh.16-0955 | Afghanistan/epidemiology;Africa South of the Sahara/epidemiology;Antimala | PMC5637593 | https://pubmed.ncbi.nlm.nih.gov/28820705/ |
| rayyan-751571029 | Bretscher MT and Griffin JT and Ghani AC and Okell LC | Modelling the benefits of long-acting or transmission-blocking drugs for reducing Plasmodium falciparum transmission by case management or by mass treatment. | 2017 | Malaria journal | 16 | 341 | RAYYAN-INCLUSION: {"Richard"=>"Excluded"} \| RAYYAN-EXCLUSION-REASONS: wrong outcome,wrong study design \| USER-NOTES: {"Richard"=>["Modeling study"]} | 10.1186/s12936-017-1988-4 | Antimalarials/*therapeutic use;Case Management/*statistics & | PMC5559805 | https://pubmed.ncbi.nlm.nih.gov/28814310/ |
| rayyan-751571030 | Diawara F and Steinhardt LC and Mahamar A and Traore T and Kone DT and Diawara H and Kamate B and Kone D and Diallo M and Sadou A and Mihigo J and Sagara I and Djimde AA and | Measuring the impact of seasonal malaria chemoprevention as part of routine malaria control in Kita, Mali. | 2017 | Malaria journal | 16 | 325 | RAYYAN-INCLUSION: {"Richard"=>"Excluded"} \| RAYYAN-EXCLUSION-REASONS: wrong outcome,wrong study design | 10.1186/s12936-017-1974-x | Amodiaquine/therapeutic use;Anemia/epidemiology;Antimalarials/*therapeutic | PMC5553795 | https://pubmed.ncbi.nlm.nih.gov/28797263/ |
| rayyan-751571031 | Ley B and Thriemer K and Jaswal J and Poirot E and Alam MS and Phru CS and Khan WA and Dysoley L and Qi G and Kheong CC and Shamsudin UK and Chen I and Hwang J and Gosling R | Barriers to routine G6PD testing prior to treatment with primaquine. | 2017 | Malaria journal | 16 | 329 | RAYYAN-INCLUSION: {"Richard"=>"Excluded"} \| RAYYAN-EXCLUSION-REASONS: wrong outcome,wrong study design | 10.1186/s12936-017-1981-y | Administrative Personnel/psychology;Bangladesh;Cambodia;China;Diagnosti | PMC5553859 | https://pubmed.ncbi.nlm.nih.gov/28797255/ |
| rayyan-751571033 | Yukich J and Bennett A and Yukich R and Stuck L and Hamainza B and Silumbe K and Smith T and Chitnis N and Steketee RW and Finn T and Eisele TP and Miller JM | Estimation of malaria parasite reservoir coverage using reactive case detection and active community fever screening from census data with rapid diagnostic tests in southern Zambia: a re-sampling approach. | 2017 | Malaria journal | 16 | 317 | RAYYAN-INCLUSION: {"Richard"=>"Excluded"} \| USER-NOTES: {"Richard"=>["No routine data used"]} | 10.1186/s12936-017-1962-1 | Antimalarials/*therapeutic use;Diagnostic Tests, Routine/*statistics & | PMC5547485 | https://pubmed.ncbi.nlm.nih.gov/28784122/ |
| rayyan-751571034 | Rosas-Aguirre A and Guzman-Guzman M and Gamboa D and Chuquiyauri R and Ramirez R and Manrique P and Carrasco-Escobar G and Puemape C and Llanos-Cuentas A and Vinetz | Micro-heterogeneity of malaria transmission in the Peruvian Amazon: a baseline assessment underlying a population-based cohort study. | 2017 | Malaria journal | 16 | 312 | RAYYAN-INCLUSION: {"Richard"=>"Excluded"} \| RAYYAN-EXCLUSION-REASONS: wrong outcome,wrong study design \| USER-NOTES: {"Richard"=>["Epi study"]} | 10.1186/s12936-017-1957-y | Adolescent;Adult;Aged;Child;Child, Preschool;Cohort Studies;Ecosystem;Female;Hu | PMC5544973 | https://pubmed.ncbi.nlm.nih.gov/28778210/ |
| rayyan-751571035 | Golassa L and White MT | Population-level estimates of the proportion of Plasmodium vivax blood-stage infections attributable to relapses among febrile patients attending Adama Malaria Diagnostic Centre, East Shoa Zone, Oromia, Ethiopia. | 2017 | Malaria journal | 16 | 301 | RAYYAN-INCLUSION: {"Richard"=>"Excluded"} \| RAYYAN-EXCLUSION-REASONS: wrong outcome,wrong study design | 10.1186/s12936-017-1944-3 | Adolescent;Adult;Aged;Aged, 80 and over;Child;Child, Preschool;Cross-Sectional | PMC5530918 | https://pubmed.ncbi.nlm.nih.gov/28750669/ |
| rayyan-751571036 | Khagayi S and Amek N and Bigogo G and Odhiambo F and Vounatsou P | Bayesian spatio-temporal modeling of mortality in relation to malaria incidence in Western Kenya. | 2017 | PloS one | 12 | e0180516 | RAYYAN-INCLUSION: {"Richard"=>"Excluded"} \| RAYYAN-EXCLUSION-REASONS: wrong outcome,wrong study design | 10.1371/journal.pone.0180516 | Adolescent;Bayes Theorem;Child;Child, Preschool;Female;Humans;Inci | PMC5509217 | https://pubmed.ncbi.nlm.nih.gov/28704417/ |
| rayyan-751571037 | Chourasia MK and Raghavendra K and Bhatt RM and Swain DK and Dutta GDP and Kleinschmidt I | Involvement of Mitanins (female health volunteers) in active malaria surveillance, determinants and challenges in tribal populated malaria endemic villages of Chhattisgarh, India. | 2017 | BMC public health | 18 | 9 | RAYYAN-INCLUSION: {"Richard"=>"Excluded"} \| RAYYAN-EXCLUSION-REASONS: wrong outcome,wrong study design | 10.1186/s12889-017-4565-4 | Adult;*Community Health Workers/education;Education, Nonprofessional;Endemic | PMC5504842 | https://pubmed.ncbi.nlm.nih.gov/28693465/ |
| rayyan-751571038 | CorrÃªa G and Das M and Kovelamudi R and Jaladi N and Pignon C and Vysyaraju K and Yedla U and Laxmi V and Vemula P and Gowthami V and Sharma H and Remartinez D | High burden of malaria and anemia among tribal pregnant women in a chronic conflict corridor in India. | 2017 | Conflict and health | 11 | 10 | RAYYAN-INCLUSION: {"Richard"=>"Excluded"} \| RAYYAN-EXCLUSION-REASONS: wrong outcome,wrong study design \| USER-NOTES: {"Richard"=>["Epi study"]} | 10.1186/s13031-017-0113-1 | Anemia;India;Malaria | PMC5477337 | https://pubmed.ncbi.nlm.nih.gov/28649273/ |
| rayyan-751571039 | Stobaugh HC and Bollinger LB and Adams SE and Crocker AH and Grise JB and Kennedy JA and Thakwalakwa C and Maleta KM and Dietzen DJ and Manary MJ and Trehan I | Effect of a package of health and nutrition services on sustained recovery in children after moderate acute malnutrition and factors related to sustaining recovery: a cluster-randomized trial. | 2017 | The American journal of | 106 | 657-666 | RAYYAN-INCLUSION: {"Richard"=>"Excluded"} \| RAYYAN-EXCLUSION-REASONS: wrong outcome,wrong study design,wrong infection / disease | 10.3945/ajcn.116.149799 | Child;Child Nutrition Disorders/diet therapy/*therapy;Child | PMC6482975 | https://pubmed.ncbi.nlm.nih.gov/28615258/ |
| rayyan-751571040 | Samuels AM and Awino N and Odongo W and Abong'o B and Gimnig J and Otieno K and Shi YP and Were V and Allen DR and Were F and Sang T and Obor D and Williamson J and | Community-based intermittent mass testing and treatment for malaria in an area of high transmission intensity, western Kenya: study design and methodology for a cluster randomized controlled trial. | 2017 | Malaria journal | 16 | 240 | RAYYAN-INCLUSION: {"Richard"=>"Excluded"} \| RAYYAN-EXCLUSION-REASONS: wrong outcome,wrong study design,wrong publication type \| USER-NOTES: {"Richard"=>["Protocol"]} | 10.1186/s12936-017-1883-z | Adolescent;Adult;Aged;Aged, 80 and over;Antimalarials/*therapeuti | PMC5463392 | https://pubmed.ncbi.nlm.nih.gov/28592250/ |
| rayyan-751571041 | Jephcott FL and Wood JLN and Cunningham AA | Facility-based surveillance for emerging infectious diseases; diagnostic practices in rural West African hospital settings: observations from Ghana. | 2017 | Philosophical transactio | 372 | | RAYYAN-INCLUSION: {"Richard"=>"Excluded"} \| RAYYAN-EXCLUSION-REASONS: wrong study duration,wrong infection / disease | 10.1098/rstb.2016.0544 | Communicable Diseases, Emerging/*diagnosis;Ghana;Hospitals, Rural/*statistics & | PMC5468698 | https://pubmed.ncbi.nlm.nih.gov/28584181/ |
| rayyan-751571042 | Wanja E and Achilla R and Obare P and Adeny R and Moseti C and Otieno V and Morang'a C and Murigi E and Nyamuni J and Monthei DR and Ogutu B and Buff AM | Evaluation of a laboratory quality assurance pilot programme for malaria diagnostics in low-transmission areas of Kenya, 2013. | 2017 | Malaria journal | 16 | 221 | RAYYAN-INCLUSION: {"Richard"=>"Excluded"} \| RAYYAN-EXCLUSION-REASONS: wrong outcome,wrong study design | 10.1186/s12936-017-1856-2 | Diagnostic Tests, Routine/*methods;Health Facilities/statistics & numerical | PMC5445328 | https://pubmed.ncbi.nlm.nih.gov/28545579/ |
| rayyan-751571043 | Devine A and Parmiter M and Chu CS and Bancone G and Nosten F and Price RN and Lubell Y and Yeung S | Using G6PD tests to enable the safe treatment of Plasmodium vivax infections with primaquine on the Thailand-Myanmar border: A cost-effectiveness analysis. | 2017 | PLoS neglected tropical | 11 | e0005602 | RAYYAN-INCLUSION: {"Richard"=>"Excluded"} \| RAYYAN-EXCLUSION-REASONS: wrong outcome,wrong study design \| USER-NOTES: {"Richard"=>["Costing study"]} | 10.1371/journal.pntd.0005602 | Adult;Antimalarials/*adverse effects/*therapeutic use;Cost-Benefit Analysis;Diagnostic | PMC5460895 | https://pubmed.ncbi.nlm.nih.gov/28542194/ |
| rayyan-751571044 | Michael D and Mkunde SP | The malaria testing and treatment landscape in mainland Tanzania, 2016. | 2017 | Malaria journal | 16 | 202 | RAYYAN-INCLUSION: {"Richard"=>"Excluded"} \| RAYYAN-EXCLUSION-REASONS: wrong outcome,wrong study design | 10.1186/s12936-017-1819-7 | Antimalarials/*therapeutic use;Artemisinins/*therapeutic use;Commerce/statistics & | PMC5437635 | https://pubmed.ncbi.nlm.nih.gov/28521811/ |
| rayyan-751571045 | Boyce MR and O'Meara WP | Use of malaria RDTs in various health contexts across sub-Saharan Africa: a systematic review. | 2017 | BMC public health | 17 | 470 | RAYYAN-INCLUSION: {"Richard"=>"Excluded"} \| RAYYAN-EXCLUSION-REASONS: wrong outcome,wrong study design,wrong publication type \| USER-NOTES: {"Richard"=>["Review"]} | 10.1186/s12889-017-4398-1 | Africa South of the Sahara;Community Health Workers/statistics & numerical | PMC5437623 | https://pubmed.ncbi.nlm.nih.gov/28521798/ |
| rayyan-751571046 | Maung CN and Sein TT and Hlaing T and Okanurak K and Silawan T and Kaewkungwal J | Promoting community malaria control in rural Myanmar through an active community participation program using the participatory learning approach. | 2017 | Rural and remote health | 17 | 4130 | RAYYAN-INCLUSION: {"Richard"=>"Excluded"} \| RAYYAN-EXCLUSION-REASONS: wrong outcome,wrong study design \| USER-NOTES: {"Richard"=>["SBC study"]} | 10.22605/RRH4130 | Adolescent;Adult;Aged;Aged, 80 and over;Animals;Antimalarials/the | | https://pubmed.ncbi.nlm.nih.gov/28502184/ |
| rayyan-751571047 | Moore KA and Fowkes FJI and Wiladphaingern J and Wai NS and Paw MK and Pimanpanarak M and Carrara VI and Raksuansak J and Simpson JA and White NJ and Nosten F and | Mediation of the effect of malaria in pregnancy on stillbirth and neonatal death in an area of low transmission: observational data analysis. | 2017 | BMC medicine | 15 | 98 | RAYYAN-INCLUSION: {"Richard"=>"Excluded"} \| RAYYAN-EXCLUSION-REASONS: wrong outcome,wrong study design \| USER-NOTES: {"Richard"=>["Epi study"]} | 10.1186/s12916-017-0863-z | Adolescent;Adult;Anemia;Female;Gestational Age;Humans;Infant;Infant | PMC5424335 | https://pubmed.ncbi.nlm.nih.gov/28486979/ |
| rayyan-751571048 | Ugah UI and Alo MN and Owolabi JO and Okata-Nwali OD and Ekejindu IM and Ibeh N and Elom MO | Evaluation of the utility value of three diagnostic methods in the detection of malaria parasites in endemic area. | 2017 | Malaria journal | 16 | 189 | RAYYAN-INCLUSION: {"Richard"=>"Excluded"} \| RAYYAN-EXCLUSION-REASONS: wrong outcome,wrong study design | 10.1186/s12936-017-1838-4 | Adolescent;Adult;Aged;Child;Child, Preschool;Coinfection/epidemi | PMC5420404 | https://pubmed.ncbi.nlm.nih.gov/28477621/ |
| rayyan-751571049 | Leidenberger M and VoigtlÃ¤nder C and Simon N and Kappes B | SYBR(Â®) Green I-Based Fluorescence Assay to Assess Cell Viability of Malaria Parasites for Routine Use in Compound Screening. | 2017 | Methods in molecular | 1601 | 97-110 | RAYYAN-INCLUSION: {"Richard"=>"Excluded"} \| RAYYAN-EXCLUSION-REASONS: wrong outcome,wrong study design | 10.1007/978-1-4939-6960-9_9 | Antimalarials/*pharmacology; Benzothiazoles;Cell Survival/*drug | | https://pubmed.ncbi.nlm.nih.gov/28470521/ |
| rayyan-751571050 | Hansen KS and Ndyomugyenyi R and Magnussen P and Lal S and Clarke SE | Cost-effectiveness analysis of malaria rapid diagnostic tests for appropriate treatment of malaria at the community level in Uganda. | 2017 | Health policy and | 32 | 676-689 | RAYYAN-INCLUSION: {"Richard"=>"Excluded"} \| RAYYAN-EXCLUSION-REASONS: wrong outcome,wrong study design \| USER-NOTES: {"Richard"=>["Costing study\n"]} | 10.1093/heapol/czw171 | Antimalarials/economics/*therapeutic use;Artemisinins/economics/* | PMC5406761 | https://pubmed.ncbi.nlm.nih.gov/28453718/ |
| rayyan-751571051 | Zinsou C and Cherifath AB | The malaria testing and treatment landscape in Benin. | 2017 | Malaria journal | 16 | 174 | RAYYAN-INCLUSION: {"Richard"=>"Excluded"} \| RAYYAN-EXCLUSION-REASONS: wrong outcome,wrong study design \| USER-NOTES: {"Richard"=>["Rx study"]} | 10.1186/s12936-017-1808-x | Antimalarials/economics/supply & distribution/*therapeutic use;Benin;Cross-Sectional | PMC5405537 | https://pubmed.ncbi.nlm.nih.gov/28446236/ |
| rayyan-751571052 | Phanalasy S | The malaria testing and treatment landscape in the southern Lao People's Democratic Republic (PDR). | 2017 | Malaria journal | 16 | 169 | RAYYAN-INCLUSION: {"Richard"=>"Excluded"} \| RAYYAN-EXCLUSION-REASONS: wrong outcome,wrong study design | 10.1186/s12936-017-1769-0 | Antimalarials/*supply & distribution;Case Management;Clinical | PMC5404290 | https://pubmed.ncbi.nlm.nih.gov/28438155/ |
| rayyan-751571053 | BjÃ¶rkman A and Cook J and Sturrock H and Msellem M and Ali A and Xu W and Molteni F and Gosling R and Drakeley C and MÃ¥rtensson A | Spatial Distribution of Falciparum Malaria Infections in Zanzibar: Implications for Focal Drug Administration Strategies Targeting Asymptomatic Parasite Carriers. | 2017 | Clinical infectious diseases : | 64 | 1236-1243 | RAYYAN-INCLUSION: {"Richard"=>"Excluded"} \| RAYYAN-EXCLUSION-REASONS: wrong outcome,wrong study design \| USER-NOTES: {"Richard"=>["No routine data"]} | 10.1093/cid/cix136 | Adolescent;Adult;Animals;Antimalarials/*therapeutic use;Blood/parasitology;Carrier | PMC5399945 | https://pubmed.ncbi.nlm.nih.gov/28431115/ |
| rayyan-751571054 | Alegana VA and Wright J and Pezzulo C and Tatem AJ and Atkinson PM | Treatment-seeking behaviour in low- and middle-income countries estimated using a Bayesian model. | 2017 | BMC medical research | 17 | 67 | RAYYAN-INCLUSION: {"Richard"=>"Excluded"} \| RAYYAN-EXCLUSION-REASONS: wrong outcome,wrong study design | 10.1186/s12874-017-0346-0 | Bayes Theorem;Developing Countries;Fees and Charges/*statistics & | PMC5397699 | https://pubmed.ncbi.nlm.nih.gov/28427337/ |
| rayyan-751571055 | Ogoma SB and Mmando AS and Swai JK and Horstmann S and Malone D and Killeen GF | A low technology emanator treated with the volatile pyrethroid transfluthrin confers long term protection against outdoor biting vectors of lymphatic filariasis, arboviruses and malaria. | 2017 | PLoS neglected tropical | 11 | e0005455 | RAYYAN-INCLUSION: {"Richard"=>"Excluded"} \| RAYYAN-EXCLUSION-REASONS: wrong outcome,wrong study design \| USER-NOTES: {"Richard"=>["Ento study"]} | 10.1371/journal.pntd.0005455 | Animals;Anopheles;Culex;*Cyclopropanes;Elephantiasis, Filarial/prevention & | PMC5384659 | https://pubmed.ncbi.nlm.nih.gov/28388682/ |
| rayyan-751571057 | Kozycki CT and Umulisa N and Rulisa S and Mwikarago EI and Musabyimana JP and Habimana JP and Karema C and Krogstad DJ | False-negative malaria rapid diagnostic tests in Rwanda: impact of Plasmodium falciparum isolates lacking hrp2 and declining malaria transmission. | 2017 | Malaria journal | 16 | 123 | RAYYAN-INCLUSION: {"Richard"=>"Excluded"} \| RAYYAN-EXCLUSION-REASONS: wrong outcome,wrong study design | 10.1186/s12936-017-1768-1 | Adolescent;Adult;Antigens, Protozoan/*genetics;Child;Child, Preschool;Cross-Sectional | PMC5359811 | https://pubmed.ncbi.nlm.nih.gov/28320390/ |

| | | | | | | | | | | | |
|---|---|---|---|---|---|---|---|---|---|---|---|
| rayyan-751571058 | Chao CC and Zhangm Z and Weissenberger G and Chen HW and Ching WM | Lateral Flow Rapid Test for Accurate and Early Diagnosis of Scrub Typhus: A Febrile Illness of Historically Military Importance in the Pacific Rim. | 2017 | Military medicine | 182 | 369-375 | RAYYAN-INCLUSION: {"Richard"=>"Excluded"} \| RAYYAN-EXCLUSION-REASONS: wrong outcome,wrong study design | 10.7205/MILMED-D-16-00091 | Asia, Southeastern;Diagnostic Tests, Routine/instrumentation/met | | https://pubmed.ncbi.nlm.nih.gov/28291501/ |
| rayyan-751571059 | Bennett A and Bisanzio D and Yukich JO and Mappin B and Fergus CA and Lynch M and Cibulskis RE and Bhatt S and Weiss DJ and Cameron E and Gething PW and Eisele TP | Population coverage of artemisinin-based combination treatment in children younger than 5 years with fever and Plasmodium falciparum infection in Africa, 2003-2015: a modelling study using data from national surveys. | 2017 | The Lancet. Global | 5 | e418-e427 | RAYYAN-INCLUSION: {"Richard"=>"Excluded"} \| RAYYAN-EXCLUSION-REASONS: wrong outcome,wrong study design \| USER-NOTES: {"Richard"=>["Modeling study"]} | 10.1016/S2214-109X(17)30076-1 | Africa South of the Sahara;Antimalarials/*therapeutic | PMC5450656 | https://pubmed.ncbi.nlm.nih.gov/28288746/ |
| rayyan-751571060 | Chandler CI and Webb EL and Maiteki-Sebuguzi C and Nayiga S and Nabirye C and DiLiberto DD and Ssemmondo E and Dorsey G and Kamya MR and Staedke SG | The impact of an intervention to introduce malaria rapid diagnostic tests on fever case management in a high transmission setting in Uganda: A mixed-methods cluster-randomized trial (PRIME). | 2017 | PloS one | 12 | e0170998 | RAYYAN-INCLUSION: {"Richard"=>"Excluded"} \| RAYYAN-EXCLUSION-REASONS: wrong outcome,wrong study design | 10.1371/journal.pone.0170998 | *Case Management;Case-Control Studies;Child, Preschool;Cluster | PMC5347994 | https://pubmed.ncbi.nlm.nih.gov/28288172/ |
| rayyan-751571061 | Hathiwala R and Mehta PR and Nataraj G and Hathiwala S | LED fluorescence microscopy: Novel method for malaria diagnosis compared with routine methods. | 2017 | Journal of infection and | 10 | 824-828 | RAYYAN-INCLUSION: {"Richard"=>"Excluded"} \| RAYYAN-EXCLUSION-REASONS: wrong outcome,wrong study design | 10.1016/j.jiph.2017.01.001 | Cross-Sectional Studies;Diagnostic Tests, Routine/*methods;Hospitals, | | https://pubmed.ncbi.nlm.nih.gov/28279619/ |
| rayyan-751571062 | Murungi M and Fulton T and Reyes R and Matte M and Ntaro M and Mulogo E and Nyehangane D and Juliano JJ and Siedner MJ and Boum Y 2nd and Boyce RM | Improving the Specificity of Plasmodium falciparum Malaria Diagnosis in High-Transmission Settings with a Two-Step Rapid Diagnostic Test and Microscopy Algorithm. | 2017 | Journal of clinical microbiol | 55 | 1540-1549 | RAYYAN-INCLUSION: {"Richard"=>"Excluded"} \| RAYYAN-EXCLUSION-REASONS: wrong outcome,wrong study design \| USER-NOTES: {"Richard"=>["Diag study"]} | 10.1128/JCM.00130-17 | Adolescent;Adult;Aged;Algorithms;Antigens, Protozoan/*analysis;Antimalar | PMC5405272 | https://pubmed.ncbi.nlm.nih.gov/28275077/ |
| rayyan-751571063 | Darkoh EL and Larbi JA and Lawer EA | A Weather-Based Prediction Model of Malaria Prevalence in Amenfi West District, Ghana. | 2017 | Malaria research and | 2017 | 7820454 | RAYYAN-INCLUSION: {"Richard"=>"Excluded"} \| RAYYAN-EXCLUSION-REASONS: wrong outcome,wrong study design \| USER-NOTES: {"Richard"=>["Epi / strat study"]} | 10.1155/2017/7820454 | Prevalence;Ghana;Malaria | PMC5307250 | https://pubmed.ncbi.nlm.nih.gov/28255497/ |
| rayyan-751571064 | Mpanya G and Tshefu A and Likwela JL | The malaria testing and treatment market in Kinshasa, Democratic Republic of the Congo, 2013. | 2017 | Malaria journal | 16 | 94 | RAYYAN-INCLUSION: {"Richard"=>"Excluded"} \| RAYYAN-EXCLUSION-REASONS: wrong outcome,wrong study design | 10.1186/s12936-016-1659-x | Antimalarials/*supply & distribution;Democratic Republic of the | PMC5330009 | https://pubmed.ncbi.nlm.nih.gov/28241832/ |
| rayyan-751571065 | Malima R and Emidi B and Messenger LA and Oxborough RM and Batengana B and Sudi W and Weston S and Mtove G and Mugasa JP and Mosha FW and Rowland MW and Kisinza W | Experimental hut evaluation of a novel long-lasting non-pyrethroid durable wall lining for control of pyrethroid-resistant Anopheles gambiae and Anopheles funestus in Tanzania. | 2017 | Malaria journal | 16 | 82 | RAYYAN-INCLUSION: {"Richard"=>"Excluded"} \| RAYYAN-EXCLUSION-REASONS: wrong outcome,wrong study design \| USER-NOTES: {"Richard"=>["Experimental study"]} | 10.1186/s12936-017-1710-6 | Animals;*Anopheles;*Benzoates;Humans;*Insecticide Resistance;*Insecticides;Iverm | PMC5316163 | https://pubmed.ncbi.nlm.nih.gov/28212636/ |
| rayyan-751571066 | Rogier E and Plucinski M and Lucchi N and Mace K and Chang M and Lemoine JF and Candrinho B and Colborn J and Dimbu R and Fortes F and Udhayakumar V and Barnwell J | Bead-based immunoassay allows sub-picogram detection of histidine-rich protein 2 from Plasmodium falciparum and estimates reliability of malaria rapid diagnostic tests. | 2017 | PloS one | 12 | e0172139 | RAYYAN-INCLUSION: {"Richard"=>"Excluded"} \| RAYYAN-EXCLUSION-REASONS: wrong outcome,wrong study design \| USER-NOTES: {"Richard"=>["Diag study"]} | 10.1371/journal.pone.0172139 | Adolescent;Adult;Aged;Aged, 80 and over;Amino Acid Sequence;Angola/epidemiolog | PMC5305216 | https://pubmed.ncbi.nlm.nih.gov/28192523/ |
| rayyan-751571067 | Nash S and Mentzer AJ and Lule SA and Kizito D and Smits G and van der Klis FR and Elliott AM | The impact of prenatal exposure to parasitic infections and to anthelminthic treatment on antibody responses to routine immunisations given in infancy: Secondary analysis of a randomised controlled trial. | 2017 | PLoS neglected tropical | 11 | e0005213 | RAYYAN-INCLUSION: {"Richard"=>"Excluded"} \| RAYYAN-EXCLUSION-REASONS: wrong outcome,wrong study design,wrong infection / disease | 10.1371/journal.pntd.0005213 | Adult;Albendazole/administration & dosage;Animals;Anthelmintics | PMC5298230 | https://pubmed.ncbi.nlm.nih.gov/28178298/ |
| rayyan-751571068 | Cowell AN and Loy DE and Sundararaman SA and Valdivia H and Fisch K and Lescano AG and Baldeviano GC and Durand S and Gerbasi V and Sutherland CJ and Nolder D and Vinetz JM | Selective Whole-Genome Amplification Is a Robust Method That Enables Scalable Whole-Genome Sequencing of Plasmodium vivax from Unprocessed Clinical Samples. | 2017 | mBio | 8 | | RAYYAN-INCLUSION: {"Richard"=>"Excluded"} \| RAYYAN-EXCLUSION-REASONS: wrong outcome,wrong study design \| USER-NOTES: {"Richard"=>["Genomic surveillance"]} | 10.1128/mBio.02257-16 | Blood/*parasitology;Humans;Malaria, Vivax/*parasitology;Nucleic | PMC5296604 | https://pubmed.ncbi.nlm.nih.gov/28174312/ |
| rayyan-751571069 | Biggs J and Raman J and Cook J and Hlongwana K and Drakeley C and Morris N and Serocharan I and Agubuzo E and Kruger P and Mabuza A and Zitha A and Machaba E and Coetzee M and | Serology reveals heterogeneity of Plasmodium falciparum transmission in northeastern South Africa: implications for malaria elimination. | 2017 | Malaria journal | 16 | 48 | RAYYAN-INCLUSION: {"Richard"=>"Excluded"} \| RAYYAN-EXCLUSION-REASONS: wrong outcome,wrong study design \| USER-NOTES: {"Richard"=>["Epi study"]} | 10.1186/s12936-017-1701-7 | Adolescent;Adult;Age Factors;Child;Child, Preschool;Cluster | PMC5270351 | https://pubmed.ncbi.nlm.nih.gov/28126001/ |
| rayyan-751571070 | Mamai W and Bimbile-Somda NS and Maiga H and Juarez JG and Muosa ZA and Ali AB and Lees RS and Gilles JR | Optimization of mosquito egg production under mass rearing setting: effects of cage volume, blood meal source and adult population density for the malaria vector, Anopheles arabiensis. | 2017 | Malaria journal | 16 | 41 | RAYYAN-INCLUSION: {"Richard"=>"Excluded"} \| RAYYAN-EXCLUSION-REASONS: wrong outcome,wrong study design \| USER-NOTES: {"Richard"=>["Ento study"]} | 10.1186/s12936-017-1685-3 | Animal Feed;Animals;Anopheles/*growth & | PMC5260048 | https://pubmed.ncbi.nlm.nih.gov/28118825/ |
| rayyan-751571071 | Njoroge MM and Tirados I and Lindsay SW and Vale GA and Torr SJ and Fillinger U | Exploring the potential of using cattle for malaria vector surveillance and control: a pilot study in western Kenya. | 2017 | Parasites & vectors | 10 | 18 | RAYYAN-INCLUSION: {"Richard"=>"Excluded"} \| RAYYAN-EXCLUSION-REASONS: wrong outcome,wrong study design \| USER-NOTES: {"Richard"=>["Ento study"]} | 10.1186/s13071-016-1957-8 | Animal Husbandry;Animals;Anopheles/drug | PMC5223359 | https://pubmed.ncbi.nlm.nih.gov/28069065/ |
| rayyan-751571072 | Kulkarni MA and Desrochers RE and Kajeguka DC and Kaaya RD and Tomayer A and Kweka EJ and Protopopoff N and Mosha FW | 10 Years of Environmental Change on the Slopes of Mount Kilimanjaro and Its Associated Shift in Malaria Vector Distributions. | 2016 | Frontiers in public health | 4 | 281 | RAYYAN-INCLUSION: {"Richard"=>"Excluded"} \| RAYYAN-EXCLUSION-REASONS: wrong outcome,wrong study design \| USER-NOTES: {"Richard"=>["Ento study"]} | 10.3389/fpubh.2016.00281 | Malaria | PMC5174083 | https://pubmed.ncbi.nlm.nih.gov/28066759/ |
| rayyan-751571073 | Tambo E and Ugwu CE and Guan Y and Wei D and Xiao-Ning and Xiao-Nong Z | China-Africa Health Development Initiatives: Benefits and Implications for Shaping Innovative and Evidence-informed National Health Policies and Programs in Sub-saharan African Countries. | 2016 | International journal of | 5 | 119-133 | RAYYAN-INCLUSION: {"Richard"=>"Excluded"} \| RAYYAN-EXCLUSION-REASONS: wrong outcome,wrong study design | 10.21106/ijma.100 | Health Policy;China | PMC5187644 | https://pubmed.ncbi.nlm.nih.gov/28058199/ |
| rayyan-751571074 | Comfort A and Leegwater A and Nakhimovsky S and Kansembe H and Hamainza B and Bwalya B and Alilio M and Johns B and Olsho L | Exploring the use of routinely-available, retrospective data to study the association between malaria control scale-up and micro-economic outcomes in Zambia. | 2017 | Malaria journal | 16 | 15 | RAYYAN-INCLUSION: {"Richard"=>"Excluded"} \| RAYYAN-EXCLUSION-REASONS: wrong outcome,wrong study design,wrong study duration \| USER-NOTES: {"Richard"=>["Survey 2010"]} | 10.1186/s12936-016-1665-z | Adolescent;Adult;Aged;Aged, 80 and over;Agriculture/*economics/ | PMC5209918 | https://pubmed.ncbi.nlm.nih.gov/28052759/ |
| rayyan-751571075 | Tukei BB and Beke A and Lamadrid-Figueroa H | Assessing the effect of indoor residual spraying (IRS) on malaria morbidity in Northern Uganda: a before and after study. | 2017 | Malaria journal | 16 | 4 | RAYYAN-INCLUSION: {"Richard"=>"Included"} \| RAYYAN-LABELS: Excluded (outside study period) | 10.1186/s12936-016-1652-4 | Adolescent;Adult;Aged;Aged, 80 and over;Child;Child, Preschool;Disease | PMC5209922 | https://pubmed.ncbi.nlm.nih.gov/28049475/ |
| rayyan-751571076 | Johns B and Yihdego YY and Kolyada L and Dengela D and Chibsa S and Dissanayake G and George K and Taffese HS and Lucas B | Indoor Residual Spraying Delivery Models to Prevent Malaria: Comparison of Community- and District-Based Approaches in Ethiopia. | 2016 | Global health, science | 4 | 529-541 | RAYYAN-INCLUSION: {"Richard"=>"Excluded"} \| RAYYAN-EXCLUSION-REASONS: wrong outcome,wrong study design \| USER-NOTES: {"Richard"=>["No epi inidcators"]} | 10.9745/GHSP-D-16-00165 | Community Health Services/*methods;Ethiopia;Humans;Insecticides/*administr | PMC5199172 | https://pubmed.ncbi.nlm.nih.gov/27965266/ |
| rayyan-751571077 | Jegede AS and Oshiname FO and Sanou AK and Nsungwa-Sabiiti J and Ajayi IO and SiribiÃ© M and Afonne C and SermÃ© L and Falade CO | Assessing Acceptability of a Diagnostic and Malaria Treatment Package Delivered by Community Health Workers in Malaria-Endemic Settings of Burkina Faso, Nigeria, and Uganda. | 2016 | Clinical infectious diseases : | 63 | S306-S311 | RAYYAN-INCLUSION: {"Richard"=>"Excluded"} \| RAYYAN-EXCLUSION-REASONS: wrong outcome,wrong study design | 10.1093/cid/ciw630 | Antimalarials/*therapeutic use;Artemisinins/therapeutic use;Artesunate;Burkina | PMC5146702 | https://pubmed.ncbi.nlm.nih.gov/27941109/ |
| rayyan-751571078 | Falade CO and Ajayi IO and Nsungwa-Sabiiti J and SiribiÃ© M and Diarra A and SermÃ© L and Afonne C and Yusuf OB and Gansane Z and Jegede AS and Singlovic J and Gomes M | Malaria Rapid Diagnostic Tests and Malaria Microscopy for Guiding Malaria Treatment of Uncomplicated Fevers in Nigeria and Prereferral Cases in 3 African Countries. | 2016 | Clinical infectious diseases : | 63 | S290-S297 | RAYYAN-INCLUSION: {"Richard"=>"Excluded"} \| RAYYAN-EXCLUSION-REASONS: wrong outcome,wrong study design | 10.1093/cid/ciw628 | Artemisinins/administration & dosage/therapeutic use;Burkina | PMC5146700 | https://pubmed.ncbi.nlm.nih.gov/27941107/ |
| rayyan-751571079 | Sanford C and McConnell A and Osborn J | The Pretravel Consultation. | 2016 | American family physician | 94 | 620-627 | RAYYAN-INCLUSION: {"Richard"=>"Excluded"} \| RAYYAN-EXCLUSION-REASONS: wrong outcome,wrong study design,wrong publication type | | Accidents/psychology;Directive Counseling/methods/organizat | | https://pubmed.ncbi.nlm.nih.gov/27929232/ |
| rayyan-751571080 | Donald W and Pasay C and Guintran JO and Iata H and Anderson K and Nausien J and Gresty KJ and Waters NC and Vestergaard LS and Taleo G and Cheng Q | The Utility of Malaria Rapid Diagnostic Tests as a Tool in Enhanced Surveillance for Malaria Elimination in Vanuatu. | 2016 | PloS one | 11 | e0167136 | RAYYAN-INCLUSION: {"Richard"=>"Maybe"} \| RAYYAN-LABELS: Excluded (after full text) \| USER-NOTES: {"Richard"=>["Unclear if rHMIS used"]} | 10.1371/journal.pone.0167136 | Diagnostic Tests, Routine/*statistics & numerical | PMC5130254 | https://pubmed.ncbi.nlm.nih.gov/27902755/ |
| rayyan-751571081 | Antwi GD and Bates LA and King R and Mahama PR and Tagbor H and Cairns M and Newell JN | Facilitators and Barriers to Uptake of an Extended Seasonal Malaria Chemoprevention Programme in Ghana: A Qualitative Study of Caregivers and Community Health Workers. | 2016 | PloS one | 11 | e0166951 | RAYYAN-INCLUSION: {"Richard"=>"Excluded"} \| RAYYAN-EXCLUSION-REASONS: wrong outcome,wrong study design \| USER-NOTES: {"Richard"=>["Qual study"]} | 10.1371/journal.pone.0166951 | Antimalarials/*therapeutic use;Attitude of Health Personnel;*Caregivers;*Case | PMC5127521 | https://pubmed.ncbi.nlm.nih.gov/27898699/ |
| rayyan-751571082 | Jain S and Zorzi N | Investing for Impact: The Global Fund Approach to Measurement of AIDS Response. | 2017 | AIDS and behavior | 21 | 91-100 | RAYYAN-INCLUSION: {"Richard"=>"Excluded"} \| RAYYAN-EXCLUSION-REASONS: wrong infection / disease | 10.1007/s10461-016-1620-6 | AIDS Serodiagnosis/*economics;Acquired Immunodeficiency | PMC5515961 | https://pubmed.ncbi.nlm.nih.gov/27896551/ |
| rayyan-751571083 | Almond D and Madanitsa M and Mwapasa V and Kalilani-Phiri L and Webster J and Ter Kuile F and Paintain L | Provider and user acceptability of intermittent screening and treatment for the control of malaria in pregnancy in Malawi. | 2016 | Malaria journal | 15 | 574 | RAYYAN-INCLUSION: {"Richard"=>"Excluded"} \| RAYYAN-EXCLUSION-REASONS: wrong outcome,wrong study design \| USER-NOTES: {"Richard"=>["Qual study"]} | 10.1186/s12936-016-1627-5 | Adolescent;Adult;Antimalarials/therapeutic use;Female;*Health | PMC5126811 | https://pubmed.ncbi.nlm.nih.gov/27894353/ |
| rayyan-751571084 | Galactionova K and Tediosi F and Camponovo F and Smith TA and Gething PW and Penny MA | Country specific predictions of the cost-effectiveness of malaria vaccine RTS,S/AS01 in endemic Africa. | 2017 | Vaccine | 35 | 53-60 | RAYYAN-INCLUSION: {"Richard"=>"Excluded"} \| RAYYAN-EXCLUSION-REASONS: wrong outcome,wrong study design \| USER-NOTES: {"Richard"=>["Costing study"]} | 10.1016/j.vaccine.2016.11.042 | Africa/epidemiology;*Cost-Benefit Analysis;Humans;Malaria | | https://pubmed.ncbi.nlm.nih.gov/27890400/ |
| rayyan-751571085 | Vandoolaeghe P and Schuerman L | The RTS,S/AS01 malaria vaccine in children 5 to 17 months of age at first vaccination. | 2016 | Expert review of vaccines | 15 | 1481-1493 | RAYYAN-INCLUSION: {"Richard"=>"Excluded"} \| RAYYAN-EXCLUSION-REASONS: wrong outcome,wrong study design,wrong publication type | 10.1080/14760584.2016.1236689 | Humans;*Immunization Schedule;Infant;Malaria/*prevention & control;Malaria | | https://pubmed.ncbi.nlm.nih.gov/27841689/ |
| rayyan-751571087 | Orobaton N and Austin AM and Abegunde D and Ibrahim M and Mohammed Z and Abdul-Azeez J and Ganiyu H and Nanbol Z and Fapohunda B and Beal K | Scaling-up the use of sulfadoxine-pyrimethamine for the preventive treatment of malaria in pregnancy: results and lessons on scalability, costs and programme impact from three local government areas in Sokoto State, Nigeria. | 2016 | Malaria journal | 15 | 533 | RAYYAN-INCLUSION: {"Richard"=>"Excluded"} \| RAYYAN-EXCLUSION-REASONS: wrong outcome,wrong study design \| USER-NOTES: {"Richard"=>["Rx study"]} | 10.1186/s12936-016-1578-x | Adolescent;Adult;Antimalarials/*administration & dosage/*economics;Drug | PMC5097385 | https://pubmed.ncbi.nlm.nih.gov/27814763/ |
| rayyan-751571088 | Liu JX and Modrek S | Evaluation of SMS reminder messages for altering treatment adherence and health seeking perceptions among malaria care-seekers in Nigeria. | 2016 | Health policy and | 31 | 1374-1383 | RAYYAN-INCLUSION: {"Richard"=>"Excluded"} \| RAYYAN-EXCLUSION-REASONS: wrong outcome,wrong study design | 10.1093/heapol/czw076 | Adult;Antimalarials/therapeutic use;Diagnostic Tests, Routine/*methods;Female;*H | PMC5091342 | https://pubmed.ncbi.nlm.nih.gov/27315831/ |
| rayyan-751571089 | Larocca A and Moro Visconti R and Marconi M | Malaria diagnosis and mapping with m-Health and geographic information systems (GIS): evidence from Uganda. | 2016 | Malaria journal | 15 | 520 | RAYYAN-INCLUSION: {"Richard"=>"Excluded"} \| RAYYAN-EXCLUSION-REASONS: wrong outcome,wrong study design | 10.1186/s12936-016-1546-5 | Adolescent;Adult;Aged;Aged, 80 and over;Child;Child, Preschool;Diagnostic Tests, | PMC5075756 | https://pubmed.ncbi.nlm.nih.gov/27776516/ |
| rayyan-751571090 | Peixoto HM and Brito MA and Romero GA and Monteiro WM and de Lacerda MV and de Oliveira MR | Rapid diagnostic test for G6PD deficiency in Plasmodium vivax-infected men: a budget impact analysis based in Brazilian Amazon. | 2017 | Tropical medicine & | 22 | 21-31 | RAYYAN-INCLUSION: {"Richard"=>"Excluded"} \| RAYYAN-EXCLUSION-REASONS: wrong outcome,wrong study design | 10.1111/tmi.12800 | Antimalarials/therapeutic use;Brazil/epidemiology;Budgets;Decision Support | | https://pubmed.ncbi.nlm.nih.gov/27770602/ |

| | | | | | | | | | | | |
|---|---|---|---|---|---|---|---|---|---|---|---|
| rayyan-751571091 | Valle D and Millar J and Amratia P | Spatial heterogeneity can undermine the effectiveness of country-wide test and treat policy for malaria: a case study from Burkina Faso. | 2016 | Malaria journal | 15 | 513 | RAYYAN-INCLUSION: {"Richard"=>"Excluded"} \| RAYYAN-EXCLUSION-REASONS: wrong outcome,wrong study design | 10.1186/s12936-016-1565-2 | Antimalarials/*therapeutic use;Burkina Faso/epidemiology;Child, | PMC5070201 | https://pubmed.ncbi.nlm.nih.gov/27760546/ |
| rayyan-751571092 | Amboko BI and Ayieko P and Ogero M and Julius T and Irimu G and English M | Malaria investigation and treatment of children admitted to county hospitals in western Kenya. | 2016 | Malaria journal | 15 | 506 | RAYYAN-INCLUSION: {"Richard"=>"Excluded"} \| RAYYAN-EXCLUSION-REASONS: wrong outcome,wrong study design \| USER-NOTES: {"Richard"=>["No routine date; Rx study"]} | 10.1186/s12936-016-1553-6 | Antimalarials/*therapeutic use;Case Management/*organization & | PMC5069818 | https://pubmed.ncbi.nlm.nih.gov/27756388/ |
| rayyan-751571093 | Burnett SM and Mbonye MK and Martin R and Ronald A and Zawedde-Muyanja S and Willis KS and Colebunders R and Manabe YC and Weaver MR | Effect of On-Site Support on Laboratory Practice for Human Immunodeficiency Virus, Tuberculosis, and Malaria Testing. | 2016 | American journal of clinical | 146 | 469-77 | RAYYAN-INCLUSION: {"Richard"=>"Excluded"} \| RAYYAN-EXCLUSION-REASONS: wrong outcome,wrong study design,wrong study duration \| USER-NOTES: {"Richard"=>["2010 study period"]} | 10.1093/ajcp/aqw138 | Diagnostic Tests, Routine;HIV Infections/*diagnosis;Humans;*Laboratories;Malaria/*diagno | | https://pubmed.ncbi.nlm.nih.gov/27686173/ |
| rayyan-751571095 | Mohapatra S and Ghosh A and Singh R and Singh DP and Sharma B and Samantaray JC and Deb M and Gaind R | Hemozoin Pigment: An Important Tool for Low Parasitemic Malarial Diagnosis. | 2016 | The Korean journal of | 54 | 393-7 | RAYYAN-INCLUSION: {"Richard"=>"Excluded"} \| RAYYAN-EXCLUSION-REASONS: wrong outcome,wrong study design | 10.3347/kjp.2016.54.4.393 | Blood Buffy Coat/*chemistry/*parasitology;Chromatography, | PMC5040090 | https://pubmed.ncbi.nlm.nih.gov/27658589/ |
| rayyan-751571096 | Ghai RR and Thurber MI and El Bakry A and Chapman CA and Goldberg TL | Multi-method assessment of patients with febrile illness reveals over-diagnosis of malaria in rural Uganda. | 2016 | Malaria journal | 15 | 460 | RAYYAN-INCLUSION: {"Richard"=>"Excluded"} \| RAYYAN-EXCLUSION-REASONS: wrong outcome,wrong study design \| USER-NOTES: {"Richard"=>["Diag study"]} | 10.1186/s12936-016-1502-4 | Adolescent;Adult;Africa;Aged;Aged, 80 and over;Blood/parasitology;Child; | PMC5015337 | https://pubmed.ncbi.nlm.nih.gov/27604542/ |
| rayyan-751571097 | Beyene HB and Beyene MB and Ebstie YA and Desalegn Z | Efficacy of Chloroquine for the Treatment of Vivax malaria in Northwest Ethiopia. | 2016 | PloS one | 11 | e0161483 | RAYYAN-INCLUSION: {"Richard"=>"Excluded"} \| RAYYAN-EXCLUSION-REASONS: wrong outcome,wrong study design \| USER-NOTES: {"Richard"=>["Rx study"]} | 10.1371/journal.pone.0161483 | Adolescent;Adult;Aged;Antimalarials/*administration & dosage/adverse | PMC5007045 | https://pubmed.ncbi.nlm.nih.gov/27579480/ |
| rayyan-751571098 | Muriuki JM and Kitala P and Muchemi G and Njeru I and Karanja J and Bett B | A comparison of malaria prevalence, control and management strategies in irrigated and non-irrigated areas in eastern Kenya. | 2016 | Malaria journal | 15 | 402 | RAYYAN-INCLUSION: {"Richard"=>"Excluded"} \| RAYYAN-EXCLUSION-REASONS: wrong outcome,wrong study design \| USER-NOTES: {"Richard"=>["No routine data"]} | 10.1186/s12936-016-1458-4 | Adolescent;Adult;Aged;Aged, 80 and over;*Agricultural Irrigation;Animals;Child;Child, | PMC4982001 | https://pubmed.ncbi.nlm.nih.gov/27515696/ |
| rayyan-751571099 | Ricks KM and Adams NM and Scherr TF and Haselton FR and Wright DW | Direct transfer of HRPII-magnetic bead complexes to malaria rapid diagnostic tests significantly improves test sensitivity. | 2016 | Malaria journal | 15 | 399 | RAYYAN-INCLUSION: {"Richard"=>"Excluded"} \| RAYYAN-EXCLUSION-REASONS: wrong outcome,wrong study design \| USER-NOTES: {"Richard"=>["Diag study"]} | 10.1186/s12936-016-1448-6 | Antigens, Protozoan/*blood;Biomarkers/*blood;Chromatography, | PMC4975893 | https://pubmed.ncbi.nlm.nih.gov/27495329/ |
| rayyan-751571100 | Tusting LS and Rek J and Arinaitwe E and Staedke SG and Kamya MR and Cano J and Bottomley C and Johnston D and Dorsey G and Lindsay SW and Lines J | Why is malaria associated with poverty? Findings from a cohort study in rural Uganda. | 2016 | Infectious diseases of | 5 | 78 | RAYYAN-INCLUSION: {"Richard"=>"Excluded"} \| RAYYAN-EXCLUSION-REASONS: wrong outcome,wrong study design \| USER-NOTES: {"Richard"=>["Epi study"]} | 10.1186/s40249-016-0164-3 | Child;Child, Preschool;Cohort Studies;Humans;Incidence;Infant;Infant, Newborn;Malaria, | PMC4972958 | https://pubmed.ncbi.nlm.nih.gov/27488674/ |
| rayyan-751571101 | Willilo RA and Molteni F and Mandike R and Mugalura FE and Mutafungwa A and Thadeo A and Benedictor E and Kafuko JM and Kaspar N and Ramsan MM and Mwaipape O and McElroy | Pregnant women and infants as sentinel populations to monitor prevalence of malaria: results of pilot study in Lake Zone of Tanzania. | 2016 | Malaria journal | 15 | 392 | RAYYAN-INCLUSION: {"Richard"=>"Excluded"} \| RAYYAN-EXCLUSION-REASONS: wrong outcome,wrong study design \| USER-NOTES: {"Richard"=>["Epi study"]} | 10.1186/s12936-016-1441-0 | Diagnostic Tests, Routine;Female;Humans;Immunization | PMC4966757 | https://pubmed.ncbi.nlm.nih.gov/27473039/ |
| rayyan-751571102 | Kasetsirikul S and Buranapong J and Srituravanich W and Kaewthamasorn M and Pimpin A | The development of malaria diagnostic techniques: a review of the approaches with focus on dielectrophoretic and magnetophoretic methods. | 2016 | Malaria journal | 15 | 358 | RAYYAN-INCLUSION: {"Richard"=>"Excluded"} \| RAYYAN-EXCLUSION-REASONS: wrong outcome,wrong study design \| USER-NOTES: {"Richard"=>["Diag study"]} | 10.1186/s12936-016-1400-9 | Diagnostic Tests, Routine/*methods;Erythrocytes/*parasitology;Humans;Mala | PMC4942956 | https://pubmed.ncbi.nlm.nih.gov/27405995/ |
| rayyan-751571103 | Oyo-Ita A and Wiysonge CS and Oringanje C and Nwachukwu CE and Oduwole O and Meremikwu MM | Interventions for improving coverage of childhood immunisation in low- and middle-income countries. | 2016 | The Cochrane database | 7 | CD008145 | RAYYAN-INCLUSION: {"Richard"=>"Excluded"} \| RAYYAN-EXCLUSION-REASONS: wrong outcome,wrong study design | 10.1002/14651858.CD008145.pub3 | *Developing Countries;*Health Education;Humans;Immunization/*statistics & numerical | PMC4981642 | https://pubmed.ncbi.nlm.nih.gov/27394698/ |
| rayyan-751571104 | Ndyomugyenyi R and Magnussen P and Lal S and Hansen K and Clarke SE | Appropriate targeting of artemisinin-based combination therapy by community health workers using malaria rapid diagnostic tests: findings from randomized trials in two contrasting areas of high and low malaria transmission in south-western Uganda. | 2016 | Tropical medicine & | 21 | 1157-70 | RAYYAN-INCLUSION: {"Richard"=>"Excluded"} \| RAYYAN-EXCLUSION-REASONS: wrong outcome,wrong study design \| USER-NOTES: {"Richard"=>["Diag study"]} | 10.1111/tmi.12748 | Adult;Antimalarials/*therapeutic use;Artemisinins/*therapeutic | PMC5031222 | https://pubmed.ncbi.nlm.nih.gov/27383558/ |
| rayyan-751571105 | Okeibunor J and Nsubuga P and Salla M and Mihigo R and Mkanda P | Coordination as a best practice from the polio eradication initiative: Experiences from five member states in the African region of the World Health Organization. | 2016 | Vaccine | 34 | 5203-5207 | RAYYAN-INCLUSION: {"Richard"=>"Excluded"} \| RAYYAN-EXCLUSION-REASONS: wrong outcome,wrong study design | 10.1016/j.vaccine.2016.05.066 | Advisory Committees;Africa/epidemiology;Chad/epidemiology;Child;D | | https://pubmed.ncbi.nlm.nih.gov/27381643/ |
| rayyan-751571106 | Onwuka JU and Akinyemi JO and Ajayi IO | Household ownership and use of insecticide-treated bednets among school children in Ibadan, Oyo State, Nigeria. | 2016 | MalariaWorld journal | 7 | 9 | RAYYAN-INCLUSION: {"Richard"=>"Excluded"} \| RAYYAN-EXCLUSION-REASONS: wrong outcome,wrong study design \| USER-NOTES: {"Richard"=>["Qual study"]} | 10.5281/zenodo.10818068 | Only Child;Child;Nigeria | PMC11003202 | https://pubmed.ncbi.nlm.nih.gov/38601354/ |
| rayyan-751571107 | Cho SJ and Lee J and Lee HJ and Jo HY and Sinniah M and Kim HY and Chong CK and Song HO | A Novel Malaria Pf/Pv Ab Rapid Diagnostic Test Using a Differential Diagnostic Marker Identified by Network Biology. | 2016 | International journal of | 12 | 824-35 | RAYYAN-INCLUSION: {"Richard"=>"Excluded"} \| RAYYAN-EXCLUSION-REASONS: wrong outcome,wrong study design \| USER-NOTES: {"Richard"=>["Dx Study"]} | 10.7150/ijbs.14408 | Antigens, Protozoan/immunology;Diagnostic Tests, | PMC4910601 | https://pubmed.ncbi.nlm.nih.gov/27313496/ |
| rayyan-751571108 | Kesteman T and Randrianarivelojosia M and Piola P and Rogier C | Post-deployment effectiveness of malaria control interventions on Plasmodium infections in Madagascar: a comprehensive phase IV assessment. | 2016 | Malaria journal | 15 | 322 | RAYYAN-INCLUSION: {"Richard"=>"Excluded"} \| RAYYAN-EXCLUSION-REASONS: wrong outcome,wrong study design \| USER-NOTES: {"Richard"=>["No routine data"]} | 10.1186/s12936-016-1376-5 | Adolescent;Adult;Aged;Aged, 80 and over;Child;Communicable | PMC4910239 | https://pubmed.ncbi.nlm.nih.gov/27306378/ |
| rayyan-751571109 | Pinchoff J and Chaponda M and Shields TM and Sichivula J and Muleba M and Mulenga M and Kobayashi T and Curriero FC and Moss WJ | Individual and Household Level Risk Factors Associated with Malaria in Nchelenge District, a Region with Perennial Transmission: A Serial Cross-Sectional Study from 2012 to 2015. | 2016 | PloS one | 11 | e0156717 | RAYYAN-INCLUSION: {"Richard"=>"Excluded"} \| RAYYAN-EXCLUSION-REASONS: wrong outcome,wrong study design \| USER-NOTES: {"Richard"=>["No routine data used"]} | 10.1371/journal.pone.0156717 | Adolescent;Child;Child, Preschool;Cross-Sectional Studies;Diagnostic Tests, | PMC4900528 | https://pubmed.ncbi.nlm.nih.gov/27281028/ |
| rayyan-751571110 | Staedke SG and Maiteki-Sebuguzi C and DiLiberto DD and Webb EL and Mugenyi L and Mbabazi E and Gonahasa S and Kigozi SP and Willey BA and Dorsey G and Kamya MR and | The Impact of an Intervention to Improve Malaria Care in Public Health Centers on Health Indicators of Children in Tororo, Uganda (PRIME): A Cluster-Randomized Trial. | 2016 | The American journal of | 95 | 358-367 | RAYYAN-INCLUSION: {"Richard"=>"Excluded"} \| RAYYAN-EXCLUSION-REASONS: wrong outcome,wrong study design | 10.4269/ajtmh.16-0103 | Anemia/*diagnosis/drug therapy/epidemiology/parasitology;Antimalarials/*therapeu | PMC4973182 | https://pubmed.ncbi.nlm.nih.gov/27273646/ |
| rayyan-751571111 | Kaingona-Daniel EP and Gomes LR and Gama BE and Almeida-de-Oliveira NK and Fortes F and MÃ©nard D and Daniel-Ribeiro CT and Ferreira-da-Cruz Mde F | Low-grade sulfadoxine-pyrimethamine resistance in Plasmodium falciparum parasites from Lubango, Angola. | 2016 | Malaria journal | 15 | 309 | RAYYAN-INCLUSION: {"Richard"=>"Excluded"} \| RAYYAN-EXCLUSION-REASONS: wrong outcome,wrong study design | 10.1186/s12936-016-1358-7 | Angola;Antimalarials/*pharmacology;Dihydropteroate Synthase/*genetics;Drug | PMC4895897 | https://pubmed.ncbi.nlm.nih.gov/27267365/ |
| rayyan-751571112 | Kuijpers LM and Maltha J and Guiraud I and KaborÃ© B and Lompo P and Devlieger H and Van Geet C and Tinto H and Jacobs J | Severe anaemia associated with Plasmodium falciparum infection in children: consequences for additional blood sampling for research. | 2016 | Malaria journal | 15 | 304 | RAYYAN-INCLUSION: {"Richard"=>"Excluded"} \| RAYYAN-EXCLUSION-REASONS: wrong outcome,wrong study design | 10.1186/s12936-016-1356-9 | Anemia/*diagnosis;Biomedical Research/*methods;Burkina Faso;Child;Child, | PMC4890332 | https://pubmed.ncbi.nlm.nih.gov/27251128/ |
| rayyan-751571113 | Garley A and Eckert E and Sie A and Ye M and Malm K and Afari EA and Sawadogo M and Herrera S and Ivanovich E and Ye Y | Strengthening individual capacity in monitoring and evaluation of malaria control programmes to streamline M&E systems and enhance information use in malaria endemic countries. | 2016 | Malaria journal | 15 | 300 | RAYYAN-INCLUSION: {"Richard"=>"Excluded"} \| RAYYAN-EXCLUSION-REASONS: wrong outcome,wrong study design | 10.1186/s12936-016-1354-y | Burkina Faso;Communicable Disease Control/*organization & administration;Disease | PMC4884432 | https://pubmed.ncbi.nlm.nih.gov/27233243/ |
| rayyan-751571114 | Vontas J and Mitsakakis K and Zengerle R and Yewhalaw D and Sikaala CH and Etang J and Fallani M and Carman B and MÃ¼ller P and ChouaÃ¯bou M and Coleman M and Coleman | Automated innovative diagnostic, data management and communication tool, for improving malaria vector control in endemic settings. | 2016 | Studies in health technolog | 224 | 54-60 | RAYYAN-INCLUSION: {"Richard"=>"Excluded"} \| RAYYAN-EXCLUSION-REASONS: wrong outcome,wrong study design | | Africa South of the Sahara/epidemiology;Animals;Culicidae/*classification/*para | | https://pubmed.ncbi.nlm.nih.gov/27225553/ |
| rayyan-751571115 | Wen S and Harvard KE and Gueye CS and Canavati SE and Chancellor A and Ahmed BN and Leaburi J and Lek D and Namgay R and Surya A and Thakur GD and Whittaker MA and | Targeting populations at higher risk for malaria: a survey of national malaria elimination programmes in the Asia Pacific. | 2016 | Malaria journal | 15 | 271 | RAYYAN-INCLUSION: {"Richard"=>"Excluded"} \| RAYYAN-EXCLUSION-REASONS: wrong outcome,wrong study design | 10.1186/s12936-016-1319-1 | Asia/epidemiology;Communicable Disease Control/*organization & | PMC4863339 | https://pubmed.ncbi.nlm.nih.gov/27165296/ |
| rayyan-751571116 | Theiss-Nyland K and Lynch M and Lines J | Assessing the availability of LLINs for continuous distribution through routine antenatal care and the Expanded Programme on Immunizations in sub-Saharan Africa. | 2016 | Malaria journal | 15 | 255 | RAYYAN-INCLUSION: {"Richard"=>"Excluded"} \| RAYYAN-EXCLUSION-REASONS: wrong outcome,wrong study design,wrong publication type \| USER-NOTES: {"Richard"=>["Review"]} | 10.1186/s12936-016-1309-3 | Africa South of the Sahara;Female;Health Policy;*Health Services | PMC4857284 | https://pubmed.ncbi.nlm.nih.gov/27146406/ |
| rayyan-751571117 | Low M and Tsegaye AT and Ignell R and Hill S and Elleby R and Feltelius V and Hopkins R | The importance of accounting for larval detectability in mosquito habitat-association studies. | 2016 | Malaria journal | 15 | 253 | RAYYAN-INCLUSION: {"Richard"=>"Excluded"} \| RAYYAN-EXCLUSION-REASONS: wrong outcome,wrong study design \| USER-NOTES: {"Richard"=>["Ento study"]} | 10.1186/s12936-016-1308-4 | Animals;Anopheles/*growth & development;*Ecosystem;Entomology/methods;Environme | PMC4855760 | https://pubmed.ncbi.nlm.nih.gov/27142303/ |
| rayyan-751571118 | Graham HR and Ayede AI and Bakare AA and Oyewole OB and Peel D and Falade AG and Duke T | Oxygen for children and newborns in non-tertiary hospitals in South-west Nigeria: A needs assessment. | 2016 | African journal of medicine | 45 | 31-49 | RAYYAN-INCLUSION: {"Richard"=>"Excluded"} \| RAYYAN-EXCLUSION-REASONS: wrong outcome,wrong study design | | Child;Cross-Sectional Studies;Health Services Accessibility/standards/statisti | | https://pubmed.ncbi.nlm.nih.gov/28686826/ |
| rayyan-751571119 | English L and Miller JS and Mbusa R and Matte M and Kenney J and Bwambale S and Ntaro M and Patel P and Mulogo E and Stone GS | Monitoring iCCM referral systems: Bugoye Integrated Community Case Management Initiative (BIMI) in Uganda. | 2016 | Malaria journal | 15 | 247 | RAYYAN-INCLUSION: {"Richard"=>"Excluded"} \| RAYYAN-EXCLUSION-REASONS: wrong outcome,wrong study design \| USER-NOTES: {"Richard"=>["No epi indicators"]} | 10.1186/s12936-016-1300-z | *Case Management/organization & administration/standards;Child | PMC4850682 | https://pubmed.ncbi.nlm.nih.gov/27129920/ |
| rayyan-751571120 | Izadi S | The effects of electricity network development besides routine malaria control measures in an underdeveloped region in the pre-elimination phase. | 2016 | Malaria journal | 15 | 222 | RAYYAN-INCLUSION: {"Richard"=>"Excluded"} \| RAYYAN-EXCLUSION-REASONS: wrong outcome,wrong study design,wrong study duration \| USER-NOTES: {"Richard"=>["2010 study period"]} | 10.1186/s12936-016-1273-y | Animals;Anopheles/drug effects;*Electric Power Supplies/statistics & numerical | PMC4835824 | https://pubmed.ncbi.nlm.nih.gov/27091331/ |
| rayyan-751571121 | Feikin DR and Flannery B and Hamel MJ and Stack M and Hansen PM | Vaccines for Children in Low- and Middle-Income Countries. | 2016 | | | | RAYYAN-INCLUSION: {"Richard"=>"Excluded"} \| RAYYAN-EXCLUSION-REASONS: wrong outcome,wrong study design,wrong publication type | 10.1596/978-1-4648-0348-2_ch10 | Only Child;Child;Vaccination;Vaccines | | https://pubmed.ncbi.nlm.nih.gov/27227236/ |
| rayyan-751571122 | Dawam JA and Madaki JKA and Gambazai AA and Okpe ES and Lar-ndam N and Onu A and Gyang M | Monthly sulphadoxine-pyrimethamine combination versus daily proguanil for malaria chemoprophylaxis in sickle cell disease: a randomized controlled study at the Jos University Teaching Hospital. | 2016 | Nigerian journal of medicine | 25 | 119-27 | RAYYAN-INCLUSION: {"Richard"=>"Excluded"} \| RAYYAN-EXCLUSION-REASONS: wrong outcome,wrong study design | | Adult;Anemia, Sickle Cell/complications/*drug therapy;Antimalarials/*admini | | https://pubmed.ncbi.nlm.nih.gov/29944308/ |
| rayyan-751571123 | Dixit A and Lee MC and Goettsch B and Afrane Y and Githeko AK and Yan G | Discovering the cost of care: consumer, provider, and retailer surveys shed light on the determinants of malaria health-seeking behaviours. | 2016 | Malaria journal | 15 | 179 | RAYYAN-INCLUSION: {"Richard"=>"Excluded"} \| RAYYAN-EXCLUSION-REASONS: wrong outcome,wrong study design \| USER-NOTES: {"Richard"=>["Qual study"]} | 10.1186/s12936-016-1232-7 | Antimalarials/*economics;Cross-Sectional Studies;Diagnostic Tests, | PMC4802645 | https://pubmed.ncbi.nlm.nih.gov/27006074/ |

| | | | | | | | | | | | |
|---|---|---|---|---|---|---|---|---|---|---|---|
| rayyan-751571124 | Beyene BB and Yalew WG and Demilew E and Abie G and Tewabe T and Abera B | Performance evaluation of rapid diagnostic test for malaria in high malarious districts of Amhara region, Ethiopia. | 2016 | Journal of vector borne | 53 | 63-9 | RAYYAN-INCLUSION: {"Richard"=>"Excluded"} \| RAYYAN-EXCLUSION-REASONS: wrong outcome,wrong study design | | Adolescent;Adult;Aged;Aged, 80 and over;Child;Child, Preschool;Chromatography, | | https://pubmed.ncbi.nlm.nih.gov/27004580/ |
| rayyan-751571125 | Wangdi K and Banwell C and Gatton ML and Kelly GC and Namgay R and Clements AC | Development and evaluation of a spatial decision support system for malaria elimination in Bhutan. | 2016 | Malaria journal | 15 | 180 | RAYYAN-INCLUSION: {"Richard"=>"Maybe"} \| RAYYAN-LABELS: Excluded (after full text) | 10.1186/s12936-016-1235-4 | Bhutan;Child, Preschool;*Decision Support Systems, | PMC4804570 | https://pubmed.ncbi.nlm.nih.gov/27004465/ |
| rayyan-751571126 | Phiri TB and Kaunda-Khangamwa BN and Bauleni A and Chimuna T and Melody D and Kalengamaliro H and Sande JH and Nsona HK and Mathanga DP | Feasibility, acceptability and impact of integrating malaria rapid diagnostic tests and pre-referral rectal artesunate into the integrated community case management programme. A pilot study in Mchinji district, Malawi. | 2016 | Malaria journal | 15 | 177 | RAYYAN-INCLUSION: {"Richard"=>"Excluded"} \| RAYYAN-EXCLUSION-REASONS: wrong outcome,wrong study design | 10.1186/s12936-016-1237-2 | Administration, Rectal;Adult;Antimalarials/*administration & | PMC4802711 | https://pubmed.ncbi.nlm.nih.gov/27000034/ |
| rayyan-751571127 | Fokam EB and Ngimuh L and Anchang-Kimbi JK and Wanji S | Assessment of the usage and effectiveness of intermittent preventive treatment and insecticide-treated nets on the indicators of malaria among pregnant women attending antenatal care in the Buea Health District, Cameroon. | 2016 | Malaria journal | 15 | 172 | RAYYAN-INCLUSION: {"Richard"=>"Excluded"} \| RAYYAN-EXCLUSION-REASONS: wrong outcome,wrong study design | 10.1186/s12936-016-1228-3 | Adolescent;Adult;Anemia/epidemiology;Antimalarials/*administration & | PMC4794838 | https://pubmed.ncbi.nlm.nih.gov/26987387/ |
| rayyan-751571128 | Hill J and Hoyt J and Achieng F and Ouma P and L'lanziva A and Kariuki S and Desai M and Webster J | User and Provider Acceptability of Intermittent Screening and Treatment and Intermittent Preventive Treatment with Dihydroartemisinin-Piperaquine to Prevent Malaria in Pregnancy in Western Kenya. | 2016 | PloS one | 11 | e0150259 | RAYYAN-INCLUSION: {"Richard"=>"Excluded"} \| RAYYAN-EXCLUSION-REASONS: wrong outcome,wrong study design \| USER-NOTES: {"Richard"=>["Qual study"]} | 10.1371/journal.pone.0150259 | Adolescent;Adult;Antimalarials/administration & dosage/*therapeutic | PMC4795545 | https://pubmed.ncbi.nlm.nih.gov/26986471/ |
| rayyan-751571129 | Kabaghe AN and Visser BJ and Spijker R and Phiri KS and Grobusch MP and van Vugt M | Health workers' compliance to rapid diagnostic tests (RDTs) to guide malaria treatment: a systematic review and meta-analysis. | 2016 | Malaria journal | 15 | 163 | RAYYAN-INCLUSION: {"Richard"=>"Excluded"} \| RAYYAN-EXCLUSION-REASONS: wrong outcome,wrong study design,wrong publication type \| USER-NOTES: {"Richard"=>["Review"]} | 10.1186/s12936-016-1218-5 | Africa South of the Sahara;Antimalarials/*therapeutic use;*Attitude of Health | PMC4791859 | https://pubmed.ncbi.nlm.nih.gov/26979286/ |
| rayyan-751571130 | Mssusa AK and Fimbo AM and Nkayamba AF and Irunde HF and Sillo HB and Shewiyo DH and Hill G and Minzi OM | Safety Profile of Artemether-Lumefantrine: A Cohort Event Monitoring Study in Public Health Facilities in Tanzania. | 2016 | Clinical drug investigat | 36 | 401-11 | RAYYAN-INCLUSION: {"Richard"=>"Excluded"} \| RAYYAN-EXCLUSION-REASONS: wrong outcome,wrong study design \| USER-NOTES: {"Richard"=>["Rx study"]} | 10.1007/s40261-016-0385-z | Adolescent;Adult;Antimalarials/*administration & dosage/*adverse | | https://pubmed.ncbi.nlm.nih.gov/26951203/ |
| rayyan-751571131 | Amegah AK and Rezza G and Jaakkola JJ | Temperature-related morbidity and mortality in Sub-Saharan Africa: A systematic review of the empirical evidence. | 2016 | Environment internatio | 91 | 133-49 | RAYYAN-INCLUSION: {"Richard"=>"Excluded"} \| RAYYAN-EXCLUSION-REASONS: wrong outcome,wrong study design,wrong publication type \| USER-NOTES: {"Richard"=>["Review"]} | 10.1016/j.envint.2016.02.027 | Africa South of the Sahara/epidemiology;Cardiovascular | | https://pubmed.ncbi.nlm.nih.gov/26949867/ |
| rayyan-751571132 | Decuypere S and Maltha J and Deborggraeve S and Rattray NJ and Issa G and BÃ©renger K and Lompo P and Tahita MC and Ruspasinghe T and McConville M and Goodacre R and Tinto H | Towards Improving Point-of-Care Diagnosis of Non-malaria Febrile Illness: A Metabolomics Approach. | 2016 | PLoS neglected tropical | 10 | e0004480 | RAYYAN-INCLUSION: {"Richard"=>"Excluded"} \| RAYYAN-EXCLUSION-REASONS: wrong outcome,wrong study design \| USER-NOTES: {"Richard"=>["Dx / immunological"]} | 10.1371/journal.pntd.0004480 | Adolescent;Africa;Child;Child, Preschool;Diagnostic Tests, Routine/*methods;Female;Fev | PMC4778767 | https://pubmed.ncbi.nlm.nih.gov/26943791/ |
| rayyan-751571133 | Wirjanata G and Handayuni I and Zaloumis SG and Chalfein F and Prayoga P and Kenangalem E and Poespoprodjo JR and Noviyanti R and Simpson JA and Price RN and Marfurt J | Analysis of ex vivo drug response data of Plasmodium clinical isolates: the pros and cons of different computer programs and online platforms. | 2016 | Malaria journal | 15 | 137 | RAYYAN-INCLUSION: {"Richard"=>"Excluded"} \| RAYYAN-EXCLUSION-REASONS: wrong outcome,wrong study design | 10.1186/s12936-016-1173-1 | Antimalarials/*pharmacology; Computational Biology;*Computer | PMC4776429 | https://pubmed.ncbi.nlm.nih.gov/26935745/ |
| rayyan-751571135 | Buchwald AG and Walldorf JA and Cohee LM and Coalson JE and Chimbiya N and Bauleni A and Nkanaunena K and Ngwira A and Kapito-Tembo A and Mathanga DP and Taylor TE and | Bed net use among school-aged children after a universal bed net campaign in Malawi. | 2016 | Malaria journal | 15 | 127 | RAYYAN-INCLUSION: {"Richard"=>"Excluded"} \| RAYYAN-EXCLUSION-REASONS: wrong outcome,wrong study design \| USER-NOTES: {"Richard"=>["No routine data / no epi outcome"]} | 10.1186/s12936-016-1178-9 | Adolescent;Child;Child, Preschool;Cross-Sectional Studies;Female;*Health | PMC4770676 | https://pubmed.ncbi.nlm.nih.gov/26928321/ |
| rayyan-751571136 | Boadu NY and Amuasi J and Ansong D and Einsiedel E and Menon D and Yanow SK | Challenges with implementing malaria rapid diagnostic tests at primary care facilities in a Ghanaian district: a qualitative study. | 2016 | Malaria journal | 15 | 126 | RAYYAN-INCLUSION: {"Richard"=>"Excluded"} \| RAYYAN-EXCLUSION-REASONS: wrong outcome,wrong study design \| USER-NOTES: {"Richard"=>["QUal study"]} | 10.1186/s12936-016-1174-0 | Adult;Aged;*Diagnostic Tests, Routine;Female;Ghana/epidemiology;*Guideline | PMC4769585 | https://pubmed.ncbi.nlm.nih.gov/26921263/ |
| rayyan-751571137 | Ng M and Ellicott Colson K and Fullman N and Dwyer-Lindgren L and Achoki T and Schneider MT and Mulenga P and Hangoma P and Masiye F and Gakidou E | Assessing the Contribution of Malaria Vector Control and Other Maternal and Child Health Interventions in Reducing All-Cause Under-Five Mortality in Zambia, 1990-2010. | 2017 | The American journal of | 97 | 58-64 | RAYYAN-INCLUSION: {"Richard"=>"Excluded"} \| RAYYAN-EXCLUSION-REASONS: wrong study duration \| USER-NOTES: {"Richard"=>["2010"]} | 10.4269/ajtmh.15-0315 | Cause of Death;Child Health;*Child Mortality;Child, Preschool;Health Impact | PMC5619928 | https://pubmed.ncbi.nlm.nih.gov/26880778/ |
| rayyan-751571138 | Peixoto HM and Brito MA and Romero GA and Monteiro WM and de Lacerda MV and de Oliveira MR | Cost-effectiveness analysis of rapid diagnostic tests for G6PD deficiency in patients with Plasmodium vivax malaria in the Brazilian Amazon. | 2016 | Malaria journal | 15 | 82 | RAYYAN-INCLUSION: {"Richard"=>"Excluded"} \| RAYYAN-EXCLUSION-REASONS: wrong outcome,wrong study design \| USER-NOTES: {"Richard"=>["Costing study"]} | 10.1186/s12936-016-1140-x | Brazil/epidemiology;Cost-Benefit Analysis;Diagnostic Tests, | PMC4750282 | https://pubmed.ncbi.nlm.nih.gov/26864333/ |
| rayyan-751571140 | van der Kam S and Roll S and Swarthout T and Edyegu-Otelu G and Matsumoto A and Kasujja FX and Casademont C and Shanks L and Salse-Ubach N | Effect of Short-Term Supplementation with Ready-to-Use Therapeutic Food or Micronutrients for Children after Illness for Prevention of Malnutrition: A Randomised Controlled Trial in Uganda. | 2016 | PLoS medicine | 13 | e1001951 | RAYYAN-INCLUSION: {"Richard"=>"Excluded"} \| RAYYAN-EXCLUSION-REASONS: wrong outcome,wrong study design,wrong infection / disease | 10.1371/journal.pmed.1001951 | Body Weight;Child, Preschool;Chronic Disease;*Dietary | PMC4747529 | https://pubmed.ncbi.nlm.nih.gov/26859481/ |
| rayyan-751571141 | Beisel U and Umlauf R and Hutchinson E and Chandler CI | The complexities of simple technologies: re-imagining the role of rapid diagnostic tests in malaria control efforts. | 2016 | Malaria journal | 15 | 64 | RAYYAN-INCLUSION: {"Richard"=>"Excluded"} \| RAYYAN-EXCLUSION-REASONS: wrong outcome,wrong study design \| USER-NOTES: {"Richard"=>["Dx study"]} | 10.1186/s12936-016-1083-2 | Diagnostic Tests, Routine/*methods;Humans;Malaria/*diagnosis;Sierra | PMC4743404 | https://pubmed.ncbi.nlm.nih.gov/26850000/ |
| rayyan-751571142 | Leurent B and Reyburn H and Muro F and Mbakilwa H and Schellenberg D | Monitoring patient care through health facility exit interviews: an assessment of the Hawthorne effect in a trial of adherence to malaria treatment guidelines in Tanzania. | 2016 | BMC infectious diseases | 16 | 59 | RAYYAN-INCLUSION: {"Richard"=>"Excluded"} \| RAYYAN-EXCLUSION-REASONS: wrong outcome,wrong study design | 10.1186/s12879-016-1362-0 | Adolescent;Adult;Antimalarials/*therapeutic use;Awareness;Child;Child, | PMC4739341 | https://pubmed.ncbi.nlm.nih.gov/26842751/ |
| rayyan-751571143 | Pinchoff J and Larsen DA and Renn S and Pollard D and Fornadel C and Maire M and Sikaala C and Sinyangwe C and Winters B and Bridges DJ and Winters AM | Targeting indoor residual spraying for malaria using epidemiological data: a case study of the Zambia experience. | 2016 | Malaria journal | 15 | 11 | RAYYAN-INCLUSION: {"Richard"=>"Excluded"} \| RAYYAN-EXCLUSION-REASONS: wrong outcome,wrong study design \| USER-NOTES: {"Richard"=>["Not clear whether this is a protocol"]} | 10.1186/s12936-015-1073-9 | Animals;Anopheles/drug effects/pathogenicity;Geographic Information | PMC4704423 | https://pubmed.ncbi.nlm.nih.gov/26738936/ |
| rayyan-751571144 | Oyeyemi OT and Sode OJ and Adebayo OD and Mensah-Agyei GO | Reliability of rapid diagnostic tests in diagnosing pregnancy and infant-associated malaria in Nigeria. | 2016 | Journal of infection and | 9 | 471-7 | RAYYAN-INCLUSION: {"Richard"=>"Excluded"} \| RAYYAN-EXCLUSION-REASONS: wrong outcome,wrong study design | 10.1016/j.jiph.2015.11.012 | Adolescent;Adult;Child, Preschool;Chromatography, Affinity/*methods;Diagnostic | | https://pubmed.ncbi.nlm.nih.gov/26738923/ |
| rayyan-751571145 | Gandapur AS and Nadeem S and Riaz M and Modood-ul-Mannan | DIAGNOSTIC IMPORTANCE OF BONE MARROW EXAMINATION IN HAEMATOLOGICAL MALIGNANT AND NON-MALIGNANT DISORDERS. | 2015 | Journal of Ayub Medical | 27 | 692-4 | RAYYAN-INCLUSION: {"Richard"=>"Excluded"} \| RAYYAN-EXCLUSION-REASONS: wrong outcome,wrong study design,wrong infection / disease | | Adolescent;Adult;Aged;Bone Marrow/*pathology;Bone Marrow | | https://pubmed.ncbi.nlm.nih.gov/26721042/ |
| rayyan-751571146 | Theiss-Nyland K and KonÃ© D and Karema C and Ejersa W and Webster J and Lines J | The relative roles of ANC and EPI in the continuous distribution of LLINs: a qualitative study in four countries. | 2017 | Health policy and | 32 | 467-475 | RAYYAN-INCLUSION: {"Richard"=>"Excluded"} \| RAYYAN-EXCLUSION-REASONS: wrong outcome,wrong study design \| USER-NOTES: {"Richard"=>["Qual study"]} | 10.1093/heapol/czw158 | Africa;*Data Collection;Female;Health Policy;Humans;Immunization | | https://pubmed.ncbi.nlm.nih.gov/28334799/ |
| rayyan-751571147 | Srivastava B and Anvikar AR and Ghosh SK and Mishra N and Kumar N and Houri-Yafin A and Pollak JJ and Salpeter SJ and Valecha N | Computer-vision-based technology for fast, accurate and cost effective diagnosis of malaria. | 2015 | Malaria journal | 14 | 526 | RAYYAN-INCLUSION: {"Richard"=>"Excluded"} \| RAYYAN-EXCLUSION-REASONS: wrong outcome,wrong study design \| USER-NOTES: {"Richard"=>["Diag study"]} | 10.1186/s12936-015-1060-1 | Costs and Cost Analysis;Diagnostic Tests, Routine/economics/instrumen | PMC4696165 | https://pubmed.ncbi.nlm.nih.gov/26714633/ |
| rayyan-751571148 | Nankabirwa JI and Yeka A and Arinaitwe E and Kigozi R and Drakeley C and Kamya MR and Greenhouse B and Rosenthal PJ and Dorsey G and Staedke SG | Estimating malaria parasite prevalence from community surveys in Uganda: a comparison of microscopy, rapid diagnostic tests and polymerase chain reaction. | 2015 | Malaria journal | 14 | 528 | RAYYAN-INCLUSION: {"Richard"=>"Excluded"} \| RAYYAN-EXCLUSION-REASONS: wrong outcome,wrong study design | 10.1186/s12936-015-1056-x | Adolescent;Adult;Age Factors;Aged;Aged, 80 and over;Child;Child, | PMC4696244 | https://pubmed.ncbi.nlm.nih.gov/26714465/ |
| rayyan-751571149 | Ursing J and Rombo L and Bergqvist Y and Rodrigues A and Kofoed PE | High-Dose Chloroquine for Treatment of Chloroquine-Resistant Plasmodium falciparum Malaria. | 2016 | The Journal of infectious | 213 | 1315-21 | RAYYAN-INCLUSION: {"Richard"=>"Excluded"} \| RAYYAN-EXCLUSION-REASONS: wrong outcome,wrong study design | 10.1093/infdis/jiv590 | Antimalarials/*administration & dosage/pharmacology/*therap | | https://pubmed.ncbi.nlm.nih.gov/26656124/ |
| rayyan-751571150 | Slater HC and Ross A and OuÃ©draogo AL and White LJ and Nguon C and Walker PG and Ngor P and Aguas R and Silal SP and Dondorp AM and La Barre P and Burton R and Sauerwein | Assessing the impact of next-generation rapid diagnostic tests on Plasmodium falciparum malaria elimination strategies. | 2015 | Nature | 528 | S94-101 | RAYYAN-INCLUSION: {"Richard"=>"Excluded"} \| RAYYAN-EXCLUSION-REASONS: wrong outcome,wrong study design \| USER-NOTES: {"Richard"=>["Diag study"]} | 10.1038/nature16040 | Adolescent;Adult;Animals;Child;Child, Preschool;*Diagnostic Tests, | | https://pubmed.ncbi.nlm.nih.gov/26633771/ |
| rayyan-751571151 | Bisanzio D and Mutuku F and LaBeaud AD and Mungai PL and Muinde J and Busaidy H and Mukoko D and King CH and Kitron U | Use of prospective hospital surveillance data to define spatiotemporal heterogeneity of malaria risk in coastal Kenya. | 2015 | Malaria journal | 14 | 482 | RAYYAN-INCLUSION: {"Richard"=>"Maybe"} \| RAYYAN-LABELS: Include (full text) | 10.1186/s12936-015-1006-7 | Adolescent;Adult;Aged;Aged, 80 and over;Child;Child, Preschool;*Epidemiological | PMC4665820 | https://pubmed.ncbi.nlm.nih.gov/26625721/ |
| rayyan-751571152 | Gebreslasie MT | A review of spatial technologies with applications for malaria transmission modelling and control in Africa. | 2015 | Geospatial health | 10 | 328 | RAYYAN-INCLUSION: {"Richard"=>"Excluded"} \| RAYYAN-EXCLUSION-REASONS: wrong outcome,wrong study design \| USER-NOTES: {"Richard"=>["Strat study"]} | 10.4081/gh.2015.328 | Africa/epidemiology;Communicable Disease Control/*trends;*Geographic | | https://pubmed.ncbi.nlm.nih.gov/26618308/ |
| rayyan-751571153 | Sahu PK and Satpathi S and Behera PK and Mishra SK and Mohanty S and Wassmer SC | Pathogenesis of cerebral malaria: new diagnostic tools, biomarkers, and therapeutic approaches. | 2015 | Frontiers in cellular and | 5 | 75 | RAYYAN-INCLUSION: {"Richard"=>"Excluded"} \| RAYYAN-EXCLUSION-REASONS: wrong outcome,wrong study design | 10.3389/fcimb.2015.00075 | Animals;Biomarkers/*analysis; Diagnostic Tests, Routine/*methods;Drug | PMC4621481 | https://pubmed.ncbi.nlm.nih.gov/26579500/ |
| rayyan-751571154 | Mappin B and Cameron E and Dalrymple U and Weiss DJ and Bisanzio D and Bhatt S and Gething PW | Standardizing Plasmodium falciparum infection prevalence measured via microscopy versus rapid diagnostic test. | 2015 | Malaria journal | 14 | 460 | RAYYAN-INCLUSION: {"Richard"=>"Excluded"} \| RAYYAN-EXCLUSION-REASONS: wrong outcome,wrong study design | 10.1186/s12936-015-0984-9 | Africa South of the Sahara/epidemiology;Child;Child, Preschool;Chromatography, | PMC4650290 | https://pubmed.ncbi.nlm.nih.gov/26577805/ |
| rayyan-751571156 | Okombo J and Chibale K | Antiplasmodial drug targets: a patent review (2000 - 2013). | 2016 | Expert opinion on | 26 | 107-30 | RAYYAN-INCLUSION: {"Richard"=>"Excluded"} \| RAYYAN-EXCLUSION-REASONS: wrong outcome,wrong study design,wrong publication type \| USER-NOTES: {"Richard"=>["Review"]} | 10.1517/13543776.2016.1113258 | Animals;Antimalarials/*pharmacology;Drug Design;Drug Resistance;High-Throughput | | https://pubmed.ncbi.nlm.nih.gov/26566576/ |
| rayyan-751571157 | Penny MA and Verity R and Bever CA and Sauboin C and Galactionova K and Flasche S and White MT and Wenger EA and Van de Velde N and Pemberton-Ross P and Griffin JT and | Public health impact and cost-effectiveness of the RTS,S/AS01 malaria vaccine: a systematic comparison of predictions from four mathematical models. | 2016 | Lancet (London, England) | 387 | 367-375 | RAYYAN-INCLUSION: {"Richard"=>"Excluded"} \| RAYYAN-EXCLUSION-REASONS: wrong outcome,wrong study design,wrong study duration \| USER-NOTES: {"Richard"=>["Modeling"]} | 10.1016/S0140-6736(15)00725-4 | Africa/epidemiology;Clinical Trials, Phase III as Topic;Cost-Benefit | PMC4723722 | https://pubmed.ncbi.nlm.nih.gov/26549466/ |
| rayyan-751571158 | Valle D and Lima JM and Millar J and Amratia P and Haque U | Bias in logistic regression due to imperfect diagnostic test results and practical correction approaches. | 2015 | Malaria journal | 14 | 434 | RAYYAN-INCLUSION: {"Richard"=>"Excluded"} \| RAYYAN-EXCLUSION-REASONS: wrong outcome,wrong study design | 10.1186/s12936-015-0966-y | Brazil/epidemiology;*Diagnostic Errors;Diagnostic Tests, Routine/*methods;Humans;M | PMC4634725 | https://pubmed.ncbi.nlm.nih.gov/26537373/ |

| | | | | | | | | | | | |
|---|---|---|---|---|---|---|---|---|---|---|---|
| rayyan-751571159 | Gonzalez-Ceron L and Rodriguez MH and Sandoval MA and Santillan F and Galindo-Virgen S and Betanzos AF and Rosales AF and Palomeque OL | Effectiveness of combined chloroquine and primaquine treatment in 14 days versus intermittent single dose regimen, in an open, non-randomized, clinical trial, to eliminate Plasmodium vivax in southern Mexico. | 2015 | Malaria journal | 14 | 426 | RAYYAN-INCLUSION: {"Richard"=>"Excluded"} \| RAYYAN-EXCLUSION-REASONS: wrong outcome,wrong study design,wrong study duration | 10.1186/s12936-015-0938-2 | Adolescent;Adult;Aged;Aged, 80 and over;Antimalarials/*administra | PMC4628368 | https://pubmed.ncbi.nlm.nih.gov/26518132/ |
| rayyan-751571160 | Stevenson JC and Stresman GH and Baidjoe A and Okoth A and Oriango R and Owaga C and Marube E and Bousema T and Cox J and Drakeley C | Use of different transmission metrics to describe malaria epidemiology in the highlands of western Kenya. | 2015 | Malaria journal | 14 | 418 | RAYYAN-INCLUSION: {"Richard"=>"Excluded"} \| RAYYAN-EXCLUSION-REASONS: wrong outcome,wrong study design,wrong study duration | 10.1186/s12936-015-0944-4 | Adolescent;Adult;Child;Child, Preschool;Chromatography, Affinity;Clinical Laboratory | PMC4624380 | https://pubmed.ncbi.nlm.nih.gov/26502920/ |
| rayyan-751571161 | Umbers AJ and Unger HW and Rosanas-Urgell A and Wangnapi RA and Kattenberg JH and Jally S and Silim S and Lufele E and Karl S and Ome-Kaius M and Robinson LJ and | Accuracy of an HRP-2/panLDH rapid diagnostic test to detect peripheral and placental Plasmodium falciparum infection in Papua New Guinean women with anaemia or suspected malaria. | 2015 | Malaria journal | 14 | 412 | RAYYAN-INCLUSION: {"Richard"=>"Excluded"} \| RAYYAN-EXCLUSION-REASONS: wrong outcome,wrong study design \| USER-NOTES: {"Richard"=>["Diag study"]} | 10.1186/s12936-015-0927-5 | Adolescent;Adult;Anemia/*diagnosis;Antigens, Protozoan/blood;Chromatogra | PMC4617889 | https://pubmed.ncbi.nlm.nih.gov/26480941/ |
| rayyan-751571162 | Chaponda EB and Chandramohan D and Michelo C and Mharakurwa S and Chipeta J and Chico RM | High burden of malaria infection in pregnant women in a rural district of Zambia: a cross-sectional study. | 2015 | Malaria journal | 14 | 380 | RAYYAN-INCLUSION: {"Richard"=>"Excluded"} \| RAYYAN-EXCLUSION-REASONS: wrong outcome,wrong study design | 10.1186/s12936-015-0866-1 | Adult;Cross-Sectional Studies;Female;HIV Infections/complications/epid | PMC4590700 | https://pubmed.ncbi.nlm.nih.gov/26423819/ |
| rayyan-751571163 | Sharma D and Lather M and Dykes CL and Dang AS and Adak T and Singh OP | Disagreement in genotyping results of drug resistance alleles of the Plasmodium falciparum dihydrofolate reductase (Pfdhfr) gene by allele-specific PCR (ASPCR) assays and Sanger sequencing. | 2016 | Parasitology research | 115 | 323-8 | RAYYAN-INCLUSION: {"Richard"=>"Excluded"} \| RAYYAN-EXCLUSION-REASONS: wrong outcome,wrong study design \| USER-NOTES: {"Richard"=>["Rx study / genomic surveillance"]} | 10.1007/s00436-015-4750-2 | Alleles;Antimalarials/*pharmacology/therapeutic use;Artemisinins/pharmacolog | | https://pubmed.ncbi.nlm.nih.gov/26407876/ |
| rayyan-751571164 | Homan T and Di Pasquale A and Kiche I and Onoka K and Hiscox A and Mweresa C and Mukabana WR and Takken W and Maire N | Innovative tools and OpenHDS for health and demographic surveillance on Rusinga Island, Kenya. | 2015 | BMC research notes | 8 | 397 | RAYYAN-INCLUSION: {"Richard"=>"Maybe"} \| RAYYAN-LABELS: Excluded (No intervention) | 10.1186/s13104-015-1373-8 | *Demography;Geographic Information Systems;*Health Status | PMC4556052 | https://pubmed.ncbi.nlm.nih.gov/26323664/ |
| rayyan-751571165 | Njuguna J and Menge D and Nzou J and Chege C | Impact of an Intervention to Minimize Overdiagnosis of Malaria Cases in a Low Risk Kenyan sub-County. | 2015 | Journal of health care for | 26 | 802-10 | RAYYAN-INCLUSION: {"Richard"=>"Excluded"} \| RAYYAN-EXCLUSION-REASONS: wrong study duration | 10.1353/hpu.2015.0101 | Case Management;Diagnostic Tests, Routine;Health Personnel/*education;Human | | https://pubmed.ncbi.nlm.nih.gov/26320915/ |
| rayyan-751571166 | Yoder PS and Nsabagasani X and Eckert E and Moran A and YÃ© Y | Perspectives of health care providers on the provision of intermittent preventive treatment in pregnancy in health facilities in Malawi. | 2015 | BMC health services | 15 | 354 | RAYYAN-INCLUSION: {"Richard"=>"Excluded"} \| RAYYAN-EXCLUSION-REASONS: wrong outcome,wrong study design | 10.1186/s12913-015-0986-x | Adult;Ambulatory Care Facilities;Antimalarials/therapeutic use;Drug | PMC4552981 | https://pubmed.ncbi.nlm.nih.gov/26318623/ |
| rayyan-751571167 | Shah JA and Emina JB and Eckert E and Ye Y | Prompt access to effective malaria treatment among children under five in sub-Saharan Africa: a multi-country analysis of national household survey data. | 2015 | Malaria journal | 14 | 329 | RAYYAN-INCLUSION: {"Richard"=>"Excluded"} \| RAYYAN-EXCLUSION-REASONS: wrong outcome,wrong study design | 10.1186/s12936-015-0844-7 | Africa South of the Sahara/epidemiology;Antimalarials/*therapeutic | PMC4549012 | https://pubmed.ncbi.nlm.nih.gov/26303581/ |
| rayyan-751571168 | Cisse MB and Keita C and Dicko A and Dengela D and Coleman J and Lucas B and Mihigo J and Sadou A and Belemvire A and George K and Fornadel C and Beach R | Characterizing the insecticide resistance of Anopheles gambiae in Mali. | 2015 | Malaria journal | 14 | 327 | RAYYAN-INCLUSION: {"Richard"=>"Excluded"} \| RAYYAN-EXCLUSION-REASONS: wrong outcome,wrong study design \| USER-NOTES: {"Richard"=>["Ento study"]} | 10.1186/s12936-015-0847-4 | Animals;Anopheles/*drug effects/enzymology/genetics;Biological | PMC4546276 | https://pubmed.ncbi.nlm.nih.gov/26296644/ |
| rayyan-751571169 | Sondo P and Derra K and Diallo-Nakanabo S and Tarnagda Z and Zampa O and Kazienga A and Valea I and Sorgho H and Owusu-Dabo E and Ouedraogo JB and Guiguemde TR and | Effectiveness and safety of artemether-lumefantrine versus artesunate-amodiaquine for unsupervised treatment of uncomplicated falciparum malaria in patients of all age groups in Nanoro, Burkina Faso: a randomized open label trial. | 2015 | Malaria journal | 14 | 325 | RAYYAN-INCLUSION: {"Richard"=>"Excluded"} \| RAYYAN-EXCLUSION-REASONS: wrong outcome,wrong study design | 10.1186/s12936-015-0843-8 | Amodiaquine/adverse effects/*therapeutic use;Antimalarials/adverse | PMC4545998 | https://pubmed.ncbi.nlm.nih.gov/26289949/ |
| rayyan-751571170 | Tagbor H and Cairns M and Bojang K and Coulibaly SO and Kayentao K and Williams J and Abubakar I and Akor F and Mohammed K and Bationo R and Dabira E and Soulama A and | A Non-Inferiority, Individually Randomized Trial of Intermittent Screening and Treatment versus Intermittent Preventive Treatment in the Control of Malaria in Pregnancy. | 2015 | PloS one | 10 | e0132247 | RAYYAN-INCLUSION: {"Richard"=>"Excluded"} \| RAYYAN-EXCLUSION-REASONS: wrong outcome,wrong study design | 10.1371/journal.pone.0132247 | Adolescent;Adult;Africa;Antimalarials/*therapeutic use;Birth Weight;Burkina Faso;Drug | PMC4530893 | https://pubmed.ncbi.nlm.nih.gov/26258474/ |
| rayyan-751571171 | GÃ³mez-PÃ©rez GP and Legarda A and MuÃ±oz J and Sim BK and Ballester MR and DobaÃ±o C and Moncunill G and Campo JJ and CisterÃ³ P and Jimenez A and Barrios D and | Controlled human malaria infection by intramuscular and direct venous inoculation of cryopreserved Plasmodium falciparum sporozoites in malaria-naÃ¯ve volunteers: effect of injection volume and dose on infectivity rates. | 2015 | Malaria journal | 14 | 306 | RAYYAN-INCLUSION: {"Richard"=>"Excluded"} \| RAYYAN-EXCLUSION-REASONS: wrong outcome,wrong study design | 10.1186/s12936-015-0817-x | Adolescent;Adult;Animals;Dose-Response Relationship, Drug;Female;Humans;Injectio | PMC4527105 | https://pubmed.ncbi.nlm.nih.gov/26245196/ |
| rayyan-751571172 | Quive IM and Candrinho B and Geelhoed D | Household survey of availability of long-lasting insecticide-treated nets and its determinants in rural Mozambique. | 2015 | Malaria journal | 14 | 304 | RAYYAN-INCLUSION: {"Richard"=>"Excluded"} \| RAYYAN-EXCLUSION-REASONS: wrong outcome,wrong study duration | 10.1186/s12936-015-0811-3 | Child, Preschool;Female;Health Services | PMC4526174 | https://pubmed.ncbi.nlm.nih.gov/26243141/ |
| rayyan-751571173 | Schroeder LF and Elbireer A and Jackson JB and Amukele TK | Laboratory Diagnostics Market in East Africa: A Survey of Test Types, Test Availability, and Test Prices in Kampala, Uganda. | 2015 | PloS one | 10 | e0134578 | RAYYAN-INCLUSION: {"Richard"=>"Excluded"} \| RAYYAN-EXCLUSION-REASONS: wrong outcome,wrong study design | 10.1371/journal.pone.0134578 | Clinical Laboratory Techniques/economics/*statistics & numerical | PMC4520457 | https://pubmed.ncbi.nlm.nih.gov/26226183/ |
| rayyan-751571174 | Mbonye AK and Magnussen P and Lal S and Hansen KS and Cundill B and Chandler C and Clarke SE | A Cluster Randomised Trial Introducing Rapid Diagnostic Tests into Registered Drug Shops in Uganda: Impact on Appropriate Treatment of Malaria. | 2015 | PloS one | 10 | e0129545 | RAYYAN-INCLUSION: {"Richard"=>"Excluded"} \| RAYYAN-EXCLUSION-REASONS: wrong outcome,wrong study design | 10.1371/journal.pone.0129545 | Adolescent;Adult;Aged;Antimalarials/economics/*therapeutic use;Child;Child, | PMC4511673 | https://pubmed.ncbi.nlm.nih.gov/26200467/ |
| rayyan-751571175 | Atroosh WM and Al-Mekhlafi HM and Al-Jasari A and Sady H and Al-Delaimy AK and Nasr NA and Dawaki S and Abdulsalam AM and Ithoi I and Lau YL and Fong MY and Surin J | Genetic variation of pfhrp2 in Plasmodium falciparum isolates from Yemen and the performance of HRP2-based malaria rapid diagnostic test. | 2015 | Parasites & vectors | 8 | 388 | RAYYAN-INCLUSION: {"Richard"=>"Excluded"} \| RAYYAN-EXCLUSION-REASONS: wrong outcome,wrong study design \| USER-NOTES: {"Richard"=>["Diag study"]} | 10.1186/s13071-015-1008-x | Antigens, Protozoan/*genetics;Diagnostic Tests, | PMC4511234 | https://pubmed.ncbi.nlm.nih.gov/26198252/ |
| rayyan-751571176 | Linn AM and Ndiaye Y and Hennessee I and Gaye S and Linn P and Nordstrom K and McLaughlin M | Reduction in symptomatic malaria prevalence through proactive community treatment in rural Senegal. | 2015 | Tropical medicine & | 20 | 1438-1446 | RAYYAN-INCLUSION: {"Richard"=>"Excluded"} \| RAYYAN-EXCLUSION-REASONS: wrong outcome,wrong study design | 10.1111/tmi.12564 | Senegal;Prevalence;Malaria | | https://pubmed.ncbi.nlm.nih.gov/26171642/ |
| rayyan-751571177 | Franke J and Gebreslasie M and Bauwens I and Deleu J and Siegert F | Earth observation in support of malaria control and epidemiology: MALAREO monitoring approaches. | 2015 | Geospatial health | 10 | 335 | RAYYAN-INCLUSION: {"Richard"=>"Excluded"} \| RAYYAN-EXCLUSION-REASONS: wrong outcome,wrong study design | 10.4081/gh.2015.335 | Africa South of the Sahara/epidemiology;Animals;Anopheles/parasitology;Breedi | | https://pubmed.ncbi.nlm.nih.gov/26054520/ |
| rayyan-751571178 | Hansen KS and Grieve E and Mikhail A and Mayan I and Mohammed N and Anwar M and Baktash SH and Drake TL and Whitty CJ and Rowland MW and Leslie TJ | Cost-effectiveness of malaria diagnosis using rapid diagnostic tests compared to microscopy or clinical symptoms alone in Afghanistan. | 2015 | Malaria journal | 14 | 217 | RAYYAN-INCLUSION: {"Richard"=>"Excluded"} \| RAYYAN-EXCLUSION-REASONS: wrong outcome,wrong study design \| USER-NOTES: {"Richard"=>["Costing study"]} | 10.1186/s12936-015-0696-1 | Adolescent;Adult;Afghanistan;Aged;Animals;Antimalarials/therapeutic | PMC4450447 | https://pubmed.ncbi.nlm.nih.gov/26016871/ |
| rayyan-751571179 | Mookherji S and Ski S and Huntington D | Tracking Global Fund HIV/AIDS resources used for sexual and reproductive health service integration: case study from Ethiopia. | 2015 | Globalization and health | 11 | 21 | RAYYAN-INCLUSION: {"Richard"=>"Excluded"} \| RAYYAN-EXCLUSION-REASONS: wrong outcome,wrong study design,wrong infection / disease | 10.1186/s12992-015-0106-z | Acquired Immunodeficiency Syndrome/*economics;*Delivery of Health Care, | PMC4453032 | https://pubmed.ncbi.nlm.nih.gov/26013060/ |
| rayyan-751571180 | Reader J and Botha M and Theron A and Lauterbach SB and Rossouw C and Engelbrecht D and Wepener M and Smit A and Leroy D and Mancama D and Coetzer TL and Birkholtz LM | Nowhere to hide: interrogating different metabolic parameters of Plasmodium falciparum gametocytes in a transmission blocking drug discovery pipeline towards malaria elimination. | 2015 | Malaria journal | 14 | 213 | RAYYAN-INCLUSION: {"Richard"=>"Excluded"} \| RAYYAN-EXCLUSION-REASONS: wrong outcome,wrong study design | 10.1186/s12936-015-0718-z | Antimalarials/*pharmacology;Disease Eradication;*Drug Discovery;Malaria/*prevention | PMC4449569 | https://pubmed.ncbi.nlm.nih.gov/25994518/ |
| rayyan-751571181 | Silumbe K and Yukich JO and Hamainza B and Bennett A and Earle D and Kamuliwo M and Steketee RW and Eisele TP and Miller JM | Costs and cost-effectiveness of a large-scale mass testing and treatment intervention for malaria in Southern Province, Zambia. | 2015 | Malaria journal | 14 | 211 | RAYYAN-INCLUSION: {"Richard"=>"Excluded"} \| RAYYAN-EXCLUSION-REASONS: wrong outcome,wrong study design \| USER-NOTES: {"Richard"=>["Costing study"]} | 10.1186/s12936-015-0722-3 | Antimalarials/*therapeutic use;Artemether, Lumefantrine Drug | PMC4490652 | https://pubmed.ncbi.nlm.nih.gov/25985992/ |
| rayyan-751571182 | Morris U and Khamis M and Aydin-Schmidt B and Abass AK and Msellem MI and Nassor MH and GonzÃ¡lez IJ and MÃ¥rtensson A and Ali AS and BjÃ¶rkman A and Cook J | Field deployment of loop-mediated isothermal amplification for centralized mass-screening of asymptomatic malaria in Zanzibar: a pre-elimination setting. | 2015 | Malaria journal | 14 | 205 | RAYYAN-INCLUSION: {"Richard"=>"Excluded"} \| RAYYAN-EXCLUSION-REASONS: wrong outcome,wrong study design | 10.1186/s12936-015-0731-2 | Adolescent;Adult;Aged;Aged, 80 and over;Asymptomatic Infections/*epidemiology;Chil | PMC4440539 | https://pubmed.ncbi.nlm.nih.gov/25982190/ |
| rayyan-751571183 | Cundill B and Mbakilwa H and Chandler CI and Mtove G and Mtei F and Willetts A and Foster E and Muro F and Mwinyishehe R and Mandike R and Olomi R and Whitty CJ and | Prescriber and patient-oriented behavioural interventions to improve use of malaria rapid diagnostic tests in Tanzania: facility-based cluster randomised trial. | 2015 | BMC medicine | 13 | 118 | RAYYAN-INCLUSION: {"Richard"=>"Excluded"} \| RAYYAN-EXCLUSION-REASONS: wrong outcome,wrong study design | 10.1186/s12916-015-0346-z | Adolescent;Adult;Africa;Child;Child, Preschool;Diagnostic Tests, | PMC4445498 | https://pubmed.ncbi.nlm.nih.gov/25980737/ |
| rayyan-751571184 | Boyce RM and Muiru A and Reyes R and Ntaro M and Mulogo E and Matte M and Siedner MJ | Impact of rapid diagnostic tests for the diagnosis and treatment of malaria at a peripheral health facility in Western Uganda: an interrupted time series analysis. | 2015 | Malaria journal | 14 | 203 | RAYYAN-INCLUSION: {"Richard"=>"Maybe"} \| RAYYAN-LABELS: Include (full text) | 10.1186/s12936-015-0725-0 | Adolescent;Adult;Aged;Aged, 80 and over;Antimalarials/*therapeuti | PMC4435913 | https://pubmed.ncbi.nlm.nih.gov/25971788/ |
| rayyan-751571185 | Chima CC and Homedes N | Impact of global health governance on country health systems: the case of HIV initiatives in Nigeria. | 2015 | Journal of global health | 5 | 010407 | RAYYAN-INCLUSION: {"Richard"=>"Excluded"} \| RAYYAN-EXCLUSION-REASONS: wrong infection / disease | 10.7189/jogh.05.010407 | Nigeria | PMC4416331 | https://pubmed.ncbi.nlm.nih.gov/25969731/ |
| rayyan-751571186 | Johansson EW and Gething PW and Hildenwall H and Mappin B and Petzold M and Peterson SS and Selling KE | Effect of diagnostic testing on medicines used by febrile children less than five years in 12 malaria-endemic African countries: a mixed-methods study. | 2015 | Malaria journal | 14 | 194 | RAYYAN-INCLUSION: {"Richard"=>"Excluded"} \| RAYYAN-EXCLUSION-REASONS: wrong outcome,wrong study design \| USER-NOTES: {"Richard"=>["Diag study"]} | 10.1186/s12936-015-0709-0 | Africa South of the Sahara/epidemiology;Anti-Bacterial Agents/*therapeutic | PMC4432948 | https://pubmed.ncbi.nlm.nih.gov/25957881/ |
| rayyan-751571187 | Angwenyi V and Asante KP and TraorÃ© A and Febir LG and Tawiah C and Kwarteng A and OuÃ©draogo A and Sirima SB and Owusu-Agyei S and Imoukhuede EB and Webster J and | Health providers' perceptions of clinical trials: lessons from Ghana, Kenya and Burkina Faso. | 2015 | PloS one | 10 | e0124554 | RAYYAN-INCLUSION: {"Richard"=>"Excluded"} \| RAYYAN-EXCLUSION-REASONS: wrong outcome,wrong study design | 10.1371/journal.pone.0124554 | Burkina Faso;*Clinical Trials as Topic;Delivery of Health Care;Ghana;Health | PMC4416706 | https://pubmed.ncbi.nlm.nih.gov/25933429/ |
| rayyan-751571188 | Sagnon N and Pinder M and Tchicaya EF and Tiono AB and Faragher B and Ranson H and Lindsay SW | To assess whether addition of pyriproxyfen to long-lasting insecticidal mosquito nets increases their durability compared to standard long-lasting insecticidal mosquito nets: study protocol for a randomised controlled trial. | 2015 | Trials | 16 | 195 | RAYYAN-INCLUSION: {"Richard"=>"Excluded"} \| RAYYAN-EXCLUSION-REASONS: wrong publication type \| USER-NOTES: {"Richard"=>["Protocol"]} | 10.1186/s13063-015-0700-7 | Animals;Anopheles/*parasitology;Burkina Faso;*Disease Vectors;Equipment | PMC4423132 | https://pubmed.ncbi.nlm.nih.gov/25927378/ |
| rayyan-751571189 | Silumbe K and Chiyende E and Finn TP and Desmond M and Puta C and Hamainza B and Kamuliwo M and Larsen DA and Eisele TP and Miller J and Bennett A | A qualitative study of perceptions of a mass test and treat campaign in Southern Zambia and potential barriers to effectiveness. | 2015 | Malaria journal | 14 | 171 | RAYYAN-INCLUSION: {"Richard"=>"Excluded"} \| RAYYAN-EXCLUSION-REASONS: wrong outcome,wrong study design \| USER-NOTES: {"Richard"=>["QUal study"]} | 10.1186/s12936-015-0686-3 | *Attitude of Health Personnel;Community Health Workers/psychology;Diagnosti | PMC4426174 | https://pubmed.ncbi.nlm.nih.gov/25896068/ |
| rayyan-751571190 | Yapabandara MA and Sarmento R and de Fatima Mota Mdo R and don Bosco J and Martins N and Wickremasinghe AR | Evidence-based malaria control in Timor Leste from 2006 to 2012. | 2015 | Malaria journal | 14 | 109 | RAYYAN-INCLUSION: {"Richard"=>"Excluded"} \| RAYYAN-EXCLUSION-REASONS: wrong outcome,wrong study design,wrong study duration | 10.1186/s12936-015-0614-6 | Animals;Anopheles/parasitology/*physiology;Antimalarials/*therapeutic use;*Evidence- | PMC4363051 | https://pubmed.ncbi.nlm.nih.gov/25890294/ |

| | | | | | | | | | | | |
|---|---|---|---|---|---|---|---|---|---|---|---|
| rayyan-751571191 | Nega D and Dana D and Tefera T and Eshetu T | Anemia associated with asymptomatic malaria among pregnant women in the rural surroundings of Arba Minch Town, South Ethiopia. | 2015 | BMC research notes | 8 | 110 | RAYYAN-INCLUSION: {"Richard"=>"Excluded"} \| RAYYAN-EXCLUSION-REASONS: wrong outcome,wrong study design | 10.1186/s13104-015-1081-4 | Adult;Anemia/complications/*epidemiology;Ethiopia/epidemiology;Female;Humans;Malaria | PMC4392875 | https://pubmed.ncbi.nlm.nih.gov/25889703/ |
| rayyan-751571192 | Sibley CH | Observing in real time the evolution of artemisinin resistance in Plasmodium falciparum. | 2015 | BMC medicine | 13 | 67 | RAYYAN-INCLUSION: {"Richard"=>"Excluded"} \| RAYYAN-EXCLUSION-REASONS: wrong outcome,wrong study design | 10.1186/s12916-015-0316-5 | Antimalarials/*therapeutic use;Artemisinins/*therapeutic use;Asia, | PMC4379603 | https://pubmed.ncbi.nlm.nih.gov/25889405/ |
| rayyan-751571193 | Dassah ET and Adu-Sarkodie Y and Mayaud P | Factors associated with failure to screen for syphilis during antenatal care in Ghana: a case control study. | 2015 | BMC infectious diseases | 15 | 125 | RAYYAN-INCLUSION: {"Richard"=>"Excluded"} \| RAYYAN-EXCLUSION-REASONS: wrong infection / disease | 10.1186/s12879-015-0868-1 | Adolescent;Adult;Case-Control Studies;Female;Ghana/epidemiology;Humans;Infant, | PMC4364573 | https://pubmed.ncbi.nlm.nih.gov/25888254/ |
| rayyan-751571194 | Ayele D and Zewotir T and Mwambi H | Multiple correspondence analysis as a tool for analysis of large health surveys in African settings. | 2014 | African health sciences | 14 | 1036-45 | RAYYAN-INCLUSION: {"Richard"=>"Excluded"} \| RAYYAN-EXCLUSION-REASONS: wrong outcome,wrong study design \| USER-NOTES: {"Richard"=>["Diag study"]} | 10.4314/ahs.v14i4.35 | Demography;Diagnostic Tests, Routine;Ethiopia/epidemiology ;Family | PMC4386317 | https://pubmed.ncbi.nlm.nih.gov/25873942/ |
| rayyan-751571195 | Lee Y and Weakley AM and Nieman CC and Malvick J and Lanzaro GC | A multi-detection assay for malaria transmitting mosquitoes. | 2015 | Journal of visualized experime | | e52385 | RAYYAN-INCLUSION: {"Richard"=>"Excluded"} \| RAYYAN-EXCLUSION-REASONS: wrong outcome,wrong study design | 10.3791/52385 | Animals;Anopheles/*classification/genetics/*parasitology;DNA, | PMC4401152 | https://pubmed.ncbi.nlm.nih.gov/25867057/ |
| rayyan-751571196 | Lussiana C | Towards subsidized malaria rapid diagnostic tests. Lessons learned from programmes to subsidise artemisinin-based combination therapies in the private sector: a review. | 2016 | Health policy and | 31 | 928-39 | RAYYAN-INCLUSION: {"Richard"=>"Excluded"} \| RAYYAN-EXCLUSION-REASONS: wrong outcome,wrong study design,wrong publication type \| USER-NOTES: {"Richard"=>["Review"]} | 10.1093/heapol/czv028 | Antimalarials/supply & distribution/*therapeutic use;Artemisinins/supply & | PMC4977424 | https://pubmed.ncbi.nlm.nih.gov/25862732/ |
| rayyan-751571197 | Larsen DA and Bennett A and Silumbe K and Hamainza B and Yukich JO and Keating J and Littrell M and Miller JM and Steketee RW and Eisele TP | Population-wide malaria testing and treatment with rapid diagnostic tests and artemether-lumefantrine in southern Zambia: a community randomized step-wedge control trial design. | 2015 | The American journal of | 92 | 913-921 | RAYYAN-INCLUSION: {"Richard"=>"Maybe"} \| RAYYAN-LABELS: Excluded (after full text),Excluded (outside study period) \| USER-NOTES: {"Richard"=>["Unclear is rHMIS used; no date mentioned"]} | 10.4269/ajtmh.14-0347 | Antimalarials/*therapeutic use;Artemether, Lumefantrine Drug | PMC4426577 | https://pubmed.ncbi.nlm.nih.gov/25802434/ |
| rayyan-751571198 | Subramaniam K and Plank RM and Lin N and Goldman-Yassen A and Ivan E and Becerril C and Kemal K and Heo M and Keller MJ and Mutimura E and Anastos K and Daily JP | Plasmodium falciparum Infection Does Not Affect Human Immunodeficiency Virus Viral Load in Coinfected Rwandan Adults. | 2014 | Open forum infectious | 1 | ofu066 | RAYYAN-INCLUSION: {"Richard"=>"Excluded"} \| RAYYAN-EXCLUSION-REASONS: wrong outcome,wrong study design,wrong infection / disease | 10.1093/ofid/ofu066 | Humanities;Humanism;Humans;Plasmodium falciparum;Viral Load | PMC4281786 | https://pubmed.ncbi.nlm.nih.gov/25734136/ |
| rayyan-751571199 | Olaosebikan R and Ernest K and Bojang K and Mokuolu O and Rehman AM and Affara M and Nwakanma D and Kiechel JR and Ogunkunle T and Olagunju T and Murtala R and Omefe P | A Randomized Trial to Compare the Safety, Tolerability, and Effectiveness of 3 Antimalarial Regimens for the Prevention of Malaria in Nigerian Patients With Sickle Cell Disease. | 2015 | The Journal of infectious | 212 | 617-25 | RAYYAN-INCLUSION: {"Richard"=>"Excluded"} \| RAYYAN-EXCLUSION-REASONS: wrong outcome,wrong study design | 10.1093/infdis/jiv093 | Adolescent;Adult;Anemia, Sickle Cell/*complications;Antimalari | PMC4512609 | https://pubmed.ncbi.nlm.nih.gov/25701866/ |
| rayyan-751571200 | Cottrell G and Moussiliou A and Luty AJ and Cot M and Fievet N and Massougbodji A and Deloron P and Tuikue Ndam N | Submicroscopic Plasmodium falciparum Infections Are Associated With Maternal Anemia, Premature Births, and Low Birth Weight. | 2015 | Clinical infectious diseases : | 60 | 1481-8 | RAYYAN-INCLUSION: {"Richard"=>"Excluded"} \| RAYYAN-EXCLUSION-REASONS: wrong outcome,wrong study design | 10.1093/cid/civ122 | Adolescent;Adult;Anemia/epidemiology/*etiology;Benin/epidemiology;Cohort | | https://pubmed.ncbi.nlm.nih.gov/25694651/ |
| rayyan-751571201 | Luo Z and Sullivan SA and Carlton JM | The biology of Plasmodium vivax explored through genomics. | 2015 | Annals of the New York | 1342 | 53-61 | RAYYAN-INCLUSION: {"Richard"=>"Excluded"} \| RAYYAN-EXCLUSION-REASONS: wrong outcome,wrong study design | 10.1111/nyas.12708 | Animals;Genomics/*methods/trends;Humans;Malaria, Vivax/diagnosis/*genetics;Phyl | PMC4405435 | https://pubmed.ncbi.nlm.nih.gov/25693446/ |
| rayyan-751571202 | Lynch CA and Bruce J and Bhasin A and Roper C and Cox J and Abeku TA | Association between recent internal travel and malaria in Ugandan highland and highland fringe areas. | 2015 | Tropical medicine & | 20 | 773-80 | RAYYAN-INCLUSION: {"Richard"=>"Excluded"} \| RAYYAN-EXCLUSION-REASONS: wrong outcome,wrong study design | 10.1111/tmi.12480 | Adolescent;Adult;Case-Control Studies;Female;Geography;Humans;Malaria/*epidemiology/t | PMC5006858 | https://pubmed.ncbi.nlm.nih.gov/25689689/ |
| rayyan-751571203 | Andreoli A and Giorgetti PF and Pietra V and Melzani A and Seni W and Castelli F and Simpore J | Evaluation of a PfHRP-2 based rapid diagnostic test versus microscopy method among HIV-positive and unknown serology patients in Ouagadougou, Burkina Faso. | 2015 | The American journal of | 92 | 834-7 | RAYYAN-INCLUSION: {"Richard"=>"Excluded"} \| RAYYAN-EXCLUSION-REASONS: wrong outcome,wrong study design | 10.4269/ajtmh.13-0757 | Adolescent;Adult;Antigens, Protozoan/*blood;Burkina Faso;Child;Child, | PMC4385783 | https://pubmed.ncbi.nlm.nih.gov/25667051/ |
| rayyan-751571204 | Pfeil J and Heine JF and Mueller AK | Addition of histamine to subcutaneously injected Plasmodium berghei sporozoites increases the parasite liver load and could facilitate whole-parasite vaccination. | 2015 | Malaria journal | 14 | 36 | RAYYAN-INCLUSION: {"Richard"=>"Excluded"} \| RAYYAN-EXCLUSION-REASONS: wrong outcome,wrong study design | 10.1186/s12936-015-0552-3 | Animals;Histamine/*pharmacology;Liver/*drug effects/*parasitology;Malaria/i | PMC4318155 | https://pubmed.ncbi.nlm.nih.gov/25627880/ |
| rayyan-751571205 | Kateera F and Mens PF and Hakizimana E and Ingabire CM and Muragijemariya L and Karinda P and Grobusch MP and Mutesa L and van Vugt M | Malaria parasite carriage and risk determinants in a rural population: a malariometric survey in Rwanda. | 2015 | Malaria journal | 14 | 16 | RAYYAN-INCLUSION: {"Richard"=>"Excluded"} \| RAYYAN-EXCLUSION-REASONS: wrong outcome,wrong study design,wrong study duration | 10.1186/s12936-014-0534-x | Adolescent;Adult;Carrier State/*epidemiology/parasitology;Child;Child, | PMC4308829 | https://pubmed.ncbi.nlm.nih.gov/25604040/ |
| rayyan-751571206 | Hodgson SH and Juma E and Salim A and Magiri C and Kimani D and Njenga D and Muia A and Cole AO and Ogwang C and Awuondo K and Lowe B and Munene M and Billingsley PF and | Evaluating controlled human malaria infection in Kenyan adults with varying degrees of prior exposure to Plasmodium falciparum using sporozoites administered by intramuscular injection. | 2014 | Frontiers in microbiol | 5 | 686 | RAYYAN-INCLUSION: {"Richard"=>"Excluded"} \| RAYYAN-EXCLUSION-REASONS: wrong outcome,wrong study design | 10.3389/fmicb.2014.00686 | Humans;Humanities;Humanism;Injections, Intramuscular;Plasmodium | PMC4264479 | https://pubmed.ncbi.nlm.nih.gov/25566206/ |
| rayyan-751571207 | McCarthy KA and Wenger EA and Huynh GH and Eckhoff PA | Calibration of an intrahost malaria model and parameter ensemble evaluation of a pre-erythrocytic vaccine. | 2015 | Malaria journal | 14 | 6 | RAYYAN-INCLUSION: {"Richard"=>"Excluded"} \| RAYYAN-EXCLUSION-REASONS: wrong outcome,wrong study design | 10.1186/1475-2875-14-6 | Adolescent;Adult;Africa;Child; Child, Preschool;Disease Transmission, | PMC4326442 | https://pubmed.ncbi.nlm.nih.gov/25563798/ |
| rayyan-751571208 | Henszel Å and Janiec J and Izdebski R and Radziszewski F and PolaÅ„ski P | Emerging infectious diseases not covered by routine vaccination in Europe in 2010-2015--the review of WHO and ECDC notifications for the National IHR Focal Point in Poland. | 2015 | Przeglad epidemiologiczny | 69 | 679-86, 829-36 | RAYYAN-INCLUSION: {"Richard"=>"Excluded"} \| RAYYAN-EXCLUSION-REASONS: wrong outcome,wrong study design,wrong infection / disease | | Communicable Disease Control/*legislation & jurisprudence/methods;Comm | | https://pubmed.ncbi.nlm.nih.gov/27139344/ |
| rayyan-751571209 | Rasanathan K and MuÃ±iz M and Bakshi S and Kumar M and Solano A and Kariuki W and George A and Sylla M and Nefdt R and Young M and Diaz T | Community case management of childhood illness in sub-Saharan Africa - findings from a cross-sectional survey on policy and implementation. | 2014 | Journal of global health | 4 | 020401 | RAYYAN-INCLUSION: {"Richard"=>"Excluded"} \| RAYYAN-EXCLUSION-REASONS: wrong outcome,wrong study design,wrong study duration | 10.7189/jogh.04.020401 | Africa South of the Sahara;Cross-Sectional Studies;Cesarean Section;Case | PMC4267096 | https://pubmed.ncbi.nlm.nih.gov/25520791/ |
| rayyan-751571210 | Diggle E and Asgary R and Gore-Langton G and Nahashon E and Mungai J and Harrison R and Abagira A and Eves K and Grigoryan Z and Soti D and Juma E and Allan R | Perceptions of malaria and acceptance of rapid diagnostic tests and related treatment practises among community members and health care providers in Greater Garissa, North Eastern Province, Kenya. | 2014 | Malaria journal | 13 | 502 | RAYYAN-INCLUSION: {"Richard"=>"Excluded"} \| RAYYAN-EXCLUSION-REASONS: wrong outcome,wrong study design | 10.1186/1475-2875-13-502 | Adolescent;Adult;Animals;Diagnostic Tests, Routine/*methods;Female;*H | PMC4300559 | https://pubmed.ncbi.nlm.nih.gov/25519710/ |
| rayyan-751571211 | Mangham-Jefferies L and Wiseman V and Achonduh OA and Drake TL and Cundill B and Onwujekwe O and Mbacham W | Economic evaluation of a cluster randomized trial of interventions to improve health workers' practice in diagnosing and treating uncomplicated malaria in Cameroon. | 2014 | Value in health : the | 17 | 783-91 | RAYYAN-INCLUSION: {"Richard"=>"Excluded"} \| RAYYAN-EXCLUSION-REASONS: wrong outcome,wrong study design \| USER-NOTES: {"Richard"=>["Cost study"]} | 10.1016/j.jval.2014.07.010 | Adolescent;Adult;Antimalarials/*administration & dosage/economics/therapeuti | | https://pubmed.ncbi.nlm.nih.gov/25498773/ |
| rayyan-751571212 | Cairns ME and Leurent B and Milligan PJ | Composite endpoints for malaria case-management: not simplifying the picture? | 2014 | Malaria journal | 13 | 494 | RAYYAN-INCLUSION: {"Richard"=>"Excluded"} \| RAYYAN-EXCLUSION-REASONS: wrong outcome,wrong study design,wrong publication type | 10.1186/1475-2875-13-494 | Antimalarials/*therapeutic use;Case Management/statistics & | PMC4300677 | https://pubmed.ncbi.nlm.nih.gov/25496292/ |
| rayyan-751571213 | Ho MF and Baker J and Lee N and Luchavez J and Ariey F and Nhem S and Oyibo W and Bell D and GonzÃ¡lez I and Chiodini P and Gatton ML and Cheng Q and McCarthy JS | Circulating antibodies against Plasmodium falciparum histidine-rich proteins 2 interfere with antigen detection by rapid diagnostic tests. | 2014 | Malaria journal | 13 | 480 | RAYYAN-INCLUSION: {"Richard"=>"Excluded"} \| RAYYAN-EXCLUSION-REASONS: wrong outcome,wrong study design | 10.1186/1475-2875-13-480 | Adolescent;Adult;Aged;Antibodies, Protozoan/*blood;Antigens, | PMC4295572 | https://pubmed.ncbi.nlm.nih.gov/25481825/ |
| rayyan-751571214 | Edi CA and Koudou BG and Bellai L and Adja AM and Chouaibou M and Bonfoh B and Barry SJ and Johnson PC and MÃ¼ller P and Dongus S and N'Goran EK and Ranson H and | Long-term trends in Anopheles gambiae insecticide resistance in CÃ´te d'Ivoire. | 2014 | Parasites & vectors | 7 | 500 | RAYYAN-INCLUSION: {"Richard"=>"Excluded"} \| RAYYAN-EXCLUSION-REASONS: wrong outcome,wrong study design \| USER-NOTES: {"Richard"=>["Ento study"]} | 10.1186/s13071-014-0500-z | Animals;Anopheles/*drug effects/genetics/metabolism;Cote d'Ivoire;Insect | PMC4269959 | https://pubmed.ncbi.nlm.nih.gov/25429888/ |
| rayyan-751571215 | Ameyaw E and Nguah SB and Ansong D and Page I and Guillerm M and Bates I | The outcome of a test-treat package versus routine outpatient care for Ghanaian children with fever: a pragmatic randomized control trial. | 2014 | Malaria journal | 13 | 461 | RAYYAN-INCLUSION: {"Richard"=>"Excluded"} \| RAYYAN-EXCLUSION-REASONS: wrong outcome,wrong study design | 10.1186/1475-2875-13-461 | Ambulatory Care/methods;Anti-Bacterial Agents/*therapeutic | PMC4259007 | https://pubmed.ncbi.nlm.nih.gov/25428264/ |
| rayyan-751571216 | Maheu-Giroux M and Castro MC | Factors affecting providers' delivery of intermittent preventive treatment for malaria in pregnancy: a five-country analysis of national service provision assessment surveys. | 2014 | Malaria journal | 13 | 440 | RAYYAN-INCLUSION: {"Richard"=>"Excluded"} \| RAYYAN-EXCLUSION-REASONS: wrong outcome,wrong study design | 10.1186/1475-2875-13-440 | Africa South of the Sahara;Antimalarials/*administration & | PMC4247687 | https://pubmed.ncbi.nlm.nih.gov/25409680/ |
| rayyan-751571218 | Messenger LA and Larsen ML and Thomas JH and Rowland M | Installation of insecticide-treated durable wall lining: evaluation of attachment materials and product durability under field conditions. | 2014 | Parasites & vectors | 7 | 508 | RAYYAN-INCLUSION: {"Richard"=>"Excluded"} \| RAYYAN-EXCLUSION-REASONS: wrong outcome,wrong study design | 10.1186/s13071-014-0508-4 | Adhesives/chemistry;Animals; Construction Materials/*analysis;Culicidae/d | PMC4246572 | https://pubmed.ncbi.nlm.nih.gov/25403829/ |
| rayyan-751571219 | Revill PA and Walker S and Mabugu T and Nathoo KJ and Mugyenyi P and Kekitinwa A and Munderi P and Bwakura-Dangarembizi M and Musiime V and Bakeera-Kitaka S and | Opportunities for improving the efficiency of paediatric HIV treatment programmes. | 2015 | AIDS (London, England) | 29 | 201-10 | RAYYAN-INCLUSION: {"Richard"=>"Excluded"} \| RAYYAN-EXCLUSION-REASONS: wrong outcome,wrong study design,wrong infection / disease | 10.1097/QAD.0000000000000518 | Antiretroviral Therapy, Highly Active/economics;CD4 Lymphocyte Count;Child;Child, | PMC4318642 | https://pubmed.ncbi.nlm.nih.gov/25396263/ |
| rayyan-751571220 | Radeva-Petrova D and Kayentao K and ter Kuile FO and Sinclair D and Garner P | Drugs for preventing malaria in pregnant women in endemic areas: any drug regimen versus placebo or no treatment. | 2014 | The Cochrane database | 2014 | CD000169 | RAYYAN-INCLUSION: {"Richard"=>"Excluded"} \| RAYYAN-EXCLUSION-REASONS: wrong outcome,wrong study design,wrong publication type \| USER-NOTES: {"Richard"=>["Review"]} | 10.1002/14651858.CD000169.pub3 | Antimalarials/*therapeutic use;Female;Humans;Infant, Newborn;Malaria/drug | PMC4498495 | https://pubmed.ncbi.nlm.nih.gov/25300703/ |
| rayyan-751571221 | Hodgins S and D'Agostino A | The quality-coverage gap in antenatal care: toward better measurement of effective coverage. | 2014 | Global health, science | 2 | 173-81 | RAYYAN-INCLUSION: {"Richard"=>"Excluded"} \| RAYYAN-EXCLUSION-REASONS: wrong outcome,wrong study design,wrong publication type \| USER-NOTES: {"Richard"=>["Review"]} | 10.9745/GHSP-D-13-00176 | Blood Pressure;Data Collection;*Developing Countries;Female;Folic | PMC4168625 | https://pubmed.ncbi.nlm.nih.gov/25276575/ |
| rayyan-751571222 | Cox J and Sovannaroth S and Dy Soley L and Ngor P and Mellor S and Roca-Feltrer A | Novel approaches to risk stratification to support malaria elimination: an example from Cambodia. | 2014 | Malaria journal | 13 | 371 | RAYYAN-INCLUSION: {"Richard"=>"Excluded"} \| RAYYAN-EXCLUSION-REASONS: wrong outcome,wrong study design \| USER-NOTES: {"Richard"=>["Strat"]} | 10.1186/1475-2875-13-371 | Cambodia;Disease Eradication/*methods;*Health Information | PMC4177243 | https://pubmed.ncbi.nlm.nih.gov/25233886/ |
| rayyan-751571223 | Stuckey EM and Smith T and Chitnis N | Seasonally dependent relationships between indicators of malaria transmission and disease provided by mathematical model simulations. | 2014 | PLoS computational | 10 | e1003812 | RAYYAN-INCLUSION: {"Richard"=>"Excluded"} \| RAYYAN-EXCLUSION-REASONS: wrong outcome,wrong study design \| USER-NOTES: {"Richard"=>["Strat"]} | 10.1371/journal.pcbi.1003812 | Animals;Child;Child, Preschool;Computational Biology;*Computer | PMC4154642 | https://pubmed.ncbi.nlm.nih.gov/25187979/ |

| | | | | | | | | | | | |
|---|---|---|---|---|---|---|---|---|---|---|---|
| rayyan-751571224 | Boulay M and Lynch M and Koenker H | Comparing two approaches for estimating the causal effect of behaviour-change communication messages promoting insecticide-treated bed nets: an analysis of the 2010 Zambia malaria indicator survey. | 2014 | Malaria journal | 13 | 342 | RAYYAN-INCLUSION: {"Richard"=>"Excluded"} \| RAYYAN-EXCLUSION-REASONS: wrong outcome,wrong study design,wrong study duration | 10.1186/1475-2875-13-342 | Adolescent;Adult;Behavior Therapy/*methods;*Communication;Female;Health | PMC4161873 | https://pubmed.ncbi.nlm.nih.gov/25174278/ |
| rayyan-751571225 | Barbosa S and Gozze AB and Lima NF and Batista CL and Bastos Mda S and Nicolete VC and Fontoura PS and GonÃ§alves RM and Viana SA and Menezes MJ and Scopel KK | Epidemiology of disappearing Plasmodium vivax malaria: a case study in rural Amazonia. | 2014 | PLoS neglected tropical | 8 | e3109 | RAYYAN-INCLUSION: {"Richard"=>"Excluded"} \| RAYYAN-EXCLUSION-REASONS: wrong outcome,wrong study design | 10.1371/journal.pntd.0003109 | Adult;Brazil/epidemiology;Female;Humans;Malaria, Vivax/*epidemiology/parasitol | PMC4148206 | https://pubmed.ncbi.nlm.nih.gov/25166263/ |
| rayyan-751571226 | Kim JY and Goo YK and Ji SY and Shin HI and Han ET and Hong Y and Chung DI and Cho SH and Lee WJ | Development and efficacy of real-time PCR in the diagnosis of vivax malaria using field samples in the Republic of Korea. | 2014 | PloS one | 9 | e105871 | RAYYAN-INCLUSION: {"Richard"=>"Excluded"} \| RAYYAN-EXCLUSION-REASONS: wrong outcome,wrong study design | 10.1371/journal.pone.0105871 | Blood/parasitology;Humans;Limit of Detection;Malaria, Vivax/*diagnosis/genetics/par | PMC4141806 | https://pubmed.ncbi.nlm.nih.gov/25148038/ |
| rayyan-751571227 | Mishra N and Kaitholia K and Srivastava B and Shah NK and Narayan JP and Dev V and Phookan S and Anvikar AR and Rana R and Bharti RS and Sonal GS and Dhariwal AC and | Declining efficacy of artesunate plus sulphadoxine-pyrimethamine in northeastern India. | 2014 | Malaria journal | 13 | 284 | RAYYAN-INCLUSION: {"Richard"=>"Excluded"} \| RAYYAN-EXCLUSION-REASONS: wrong outcome,wrong study design | 10.1186/1475-2875-13-284 | Adolescent;Adult;*Antimalarials/administration & dosage/pharmacology/therape | PMC4127069 | https://pubmed.ncbi.nlm.nih.gov/25052385/ |
| rayyan-751571228 | Venkatesan M and Gadalla NB and Stepniewska K and Dahal P and Nsanzabana C and Moriera C and Price RN and MÃ¥rtensson A and Rosenthal PJ and Dorsey G and | Polymorphisms in Plasmodium falciparum chloroquine resistance transporter and multidrug resistance 1 genes: parasite risk factors that affect treatment outcomes for P. falciparum malaria after artemether-lumefantrine and artesunate-amodiaquine. | 2014 | The American journal of | 91 | 833-843 | RAYYAN-INCLUSION: {"Richard"=>"Excluded"} \| RAYYAN-EXCLUSION-REASONS: wrong outcome,wrong study design | 10.4269/ajtmh.14-0031 | Amino Acid Substitution;Amodiaquine/therapeutic | PMC4183414 | https://pubmed.ncbi.nlm.nih.gov/25048375/ |
| rayyan-751571229 | Faucher JF and Bellanger AP and Shaniya N and Hustache-Mathieu L and Hoen B and Chirouze C | Compliance to recommendations for the management of curative treatment of Plasmodium vivax/ovale infections. | 2014 | Medecine et maladies | 44 | 387-9 | RAYYAN-INCLUSION: {"Richard"=>"Excluded"} \| RAYYAN-EXCLUSION-REASONS: wrong outcome,wrong study design | 10.1016/j.medmal.2014.06.006 | Adolescent;Adult;Antimalarials/*therapeutic use;Child;Chloroquine/*therap | | https://pubmed.ncbi.nlm.nih.gov/25015307/ |
| rayyan-751571230 | Ruizendaal E and Dierickx S and Peeters Grietens K and Schallig HD and Pagnoni F and Mens PF | Success or failure of critical steps in community case management of malaria with rapid diagnostic tests: a systematic review. | 2014 | Malaria journal | 13 | 229 | RAYYAN-INCLUSION: {"Richard"=>"Excluded"} \| RAYYAN-EXCLUSION-REASONS: wrong outcome,wrong study design,wrong publication type \| USER-NOTES: {"Richard"=>["Review"]} | 10.1186/1475-2875-13-229 | Africa South of the Sahara;Case Management/organization & | PMC4084582 | https://pubmed.ncbi.nlm.nih.gov/24924295/ |
| rayyan-751571231 | Sikaala CH and Chinula D and Chanda J and Hamainza B and Mwenda M and Mukali I and Kamuliwo M and Lobo NF and Seyoum A and Killeen GF | A cost-effective, community-based, mosquito-trapping scheme that captures spatial and temporal heterogeneities of malaria transmission in rural Zambia. | 2014 | Malaria journal | 13 | 225 | RAYYAN-INCLUSION: {"Richard"=>"Excluded"} \| RAYYAN-EXCLUSION-REASONS: wrong outcome,wrong study design \| USER-NOTES: {"Richard"=>["Ento study"]} | 10.1186/1475-2875-13-225 | Adult;Animals;Anopheles/*growth & development/*parasitology;Ch | PMC4060139 | https://pubmed.ncbi.nlm.nih.gov/24906704/ |
| rayyan-751571232 | Manyando C and Njunju EM and Chileshe J and Siziya S and Shiff C | Rapid diagnostic tests for malaria and health workers' adherence to test results at health facilities in Zambia. | 2014 | Malaria journal | 13 | 166 | RAYYAN-INCLUSION: {"Richard"=>"Excluded"} \| RAYYAN-EXCLUSION-REASONS: wrong outcome,wrong study design | 10.1186/1475-2875-13-166 | Antimalarials/therapeutic use;Child, Preschool;Diagnostic Tests, | PMC4026818 | https://pubmed.ncbi.nlm.nih.gov/24885996/ |
| rayyan-751571233 | Kramer RA and Mboera LE and Senkoro K and Lesser A and Shayo EH and Paul CJ and Miranda ML | A randomized longitudinal factorial design to assess malaria vector control and disease management interventions in rural Tanzania. | 2014 | International journal of | 11 | 5317-32 | RAYYAN-INCLUSION: {"Richard"=>"Excluded"} \| RAYYAN-EXCLUSION-REASONS: wrong outcome,wrong study design,wrong publication type \| USER-NOTES: {"Richard"=>["Protocol"]} | 10.3390/ijerph110505317 | Animals;Community Health Workers;Cross-Sectional Studies;*Culicidae/growth & | PMC4053883 | https://pubmed.ncbi.nlm.nih.gov/24840349/ |
| rayyan-751571234 | Manyando C and Njunju EM and Mwakazanga D and Chongwe G and Mkandawire R and Champo D and Mulenga M and De Crop M and Claeys Y and Ravinetto RM and van Overmeir C | Safety of daily co-trimoxazole in pregnancy in an area of changing malaria epidemiology: a phase 3b randomized controlled clinical trial. | 2014 | PloS one | 9 | e96017 | RAYYAN-INCLUSION: {"Richard"=>"Excluded"} \| RAYYAN-EXCLUSION-REASONS: wrong outcome,wrong study design | 10.1371/journal.pone.0096017 | Adult;Antimalarials/*administration & dosage/adverse effects;Birth Weight;Drug | PMC4022666 | https://pubmed.ncbi.nlm.nih.gov/24830749/ |
| rayyan-751571235 | Johansson EW and Gething PW and Hildenwall H and Mappin B and Petzold M and Peterson SS and Selling KE | Diagnostic testing of pediatric fevers: meta-analysis of 13 national surveys assessing influences of malaria endemicity and source of care on test uptake for febrile children under five years. | 2014 | PloS one | 9 | e95483 | RAYYAN-INCLUSION: {"Richard"=>"Excluded"} \| RAYYAN-EXCLUSION-REASONS: wrong outcome,wrong study design \| USER-NOTES: {"Richard"=>["Review"]} | 10.1371/journal.pone.0095483 | Africa South of the Sahara/epidemiology;Child, Preschool;Cross-Sectional | PMC3991688 | https://pubmed.ncbi.nlm.nih.gov/24748201/ |
| rayyan-751571236 | VanBuskirk KM and Ofosu A and Kennedy A and Denno DM | Pediatric anemia in rural Ghana: a cross-sectional study of prevalence and risk factors. | 2014 | Journal of tropical pediatrics | 60 | 308-17 | RAYYAN-INCLUSION: {"Richard"=>"Excluded"} \| RAYYAN-EXCLUSION-REASONS: wrong outcome,wrong study design | 10.1093/tropej/fmu020 | Anemia/*epidemiology;Child, Preschool;Cross-Sectional Studies;Female;Ghana/epidem | | https://pubmed.ncbi.nlm.nih.gov/24728349/ |
| rayyan-751571237 | Matangila JR and Lufuluabo J and Ibalanky AL and InocÃªncio da Luz RA and Lutumba P and Van Geertruyden JP | Asymptomatic Plasmodium falciparum infection is associated with anaemia in pregnancy and can be more cost-effectively detected by rapid diagnostic test than by microscopy in Kinshasa, Democratic Republic of the Congo. | 2014 | Malaria journal | 13 | 132 | RAYYAN-INCLUSION: {"Richard"=>"Excluded"} \| RAYYAN-EXCLUSION-REASONS: wrong outcome,wrong study design | 10.1186/1475-2875-13-132 | Adult;Anemia/*diagnosis/epidemiology/parasitology;Asymptomatic | PMC3976674 | https://pubmed.ncbi.nlm.nih.gov/24690179/ |
| rayyan-751571238 | Rumisha SF and Smith TA and Masanja H and Abdulla S and Vounatsou P | Relationship between child survival and malaria transmission: an analysis of the malaria transmission intensity and mortality burden across Africa (MTIMBA) project data in Rufiji demographic surveillance system, Tanzania. | 2014 | Malaria journal | 13 | 124 | RAYYAN-INCLUSION: {"Richard"=>"Maybe"} \| RAYYAN-LABELS: Excluded (after full text),Excluded (outside study period) | 10.1186/1475-2875-13-124 | Animals;Anopheles/*parasitology/physiology;Bayes Theorem;Child, | PMC4021084 | https://pubmed.ncbi.nlm.nih.gov/24679119/ |
| rayyan-751571239 | Hamainza B and Moonga H and Sikaala CH and Kamuliwo M and Bennett A and Eisele TP and Miller J and Seyoum A and Killeen GF | Monitoring, characterization and control of chronic, symptomatic malaria infections in rural Zambia through monthly household visits by paid community health workers. | 2014 | Malaria journal | 13 | 128 | RAYYAN-INCLUSION: {"Richard"=>"Maybe"} \| RAYYAN-LABELS: Excluded (No intervention),Excluded (outside study period) | 10.1186/1475-2875-13-128 | *Community Health Workers/economics/statistics & numerical data;Delivery of | PMC4113135 | https://pubmed.ncbi.nlm.nih.gov/24678631/ |
| rayyan-751571240 | Gilliams EA and Jumare J and Claassen CW and Thesing PC and Nyirenda OM and Dzinjalamala FK and Taylor T and Plowe CV and Tracy LA and Laufer MK | Chloroquine-azithromycin combination antimalarial treatment decreases risk of respiratory- and gastrointestinal-tract infections in Malawian children. | 2014 | The Journal of infectious | 210 | 585-92 | RAYYAN-INCLUSION: {"Richard"=>"Excluded"} \| RAYYAN-EXCLUSION-REASONS: wrong outcome,wrong study design,wrong infection / disease | 10.1093/infdis/jiu171 | Antimalarials/*therapeutic use;Azithromycin/*therapeutic use;Child, | PMC4172042 | https://pubmed.ncbi.nlm.nih.gov/24652498/ |
| rayyan-751571241 | Senn N and Rarau P and Salib M and Manong D and Siba P and Rogerson S and Mueller I and Genton B | Use of antibiotics within the IMCI guidelines in outpatient settings in Papua New Guinean children: an observational and effectiveness study. | 2014 | PloS one | 9 | e90990 | RAYYAN-INCLUSION: {"Richard"=>"Excluded"} \| RAYYAN-EXCLUSION-REASONS: wrong infection / disease | 10.1371/journal.pone.0090990 | Anti-Bacterial Agents/*therapeutic use;Child, Preschool;Diarrhea/drug | PMC3953204 | https://pubmed.ncbi.nlm.nih.gov/24626194/ |
| rayyan-751571242 | Duke T and Fuller D | Randomised controlled trials in child health in developing countries: trends and lessons over 11 years. | 2014 | Archives of disease in | 99 | 615-20 | RAYYAN-INCLUSION: {"Richard"=>"Excluded"} \| RAYYAN-EXCLUSION-REASONS: wrong outcome,wrong study design,wrong publication type \| USER-NOTES: {"Richard"=>["Review"]} | 10.1136/archdischild-2013-305702 | Child;Child Welfare/*statistics & numerical data;*Developing Countries;Humans;Randomize | | https://pubmed.ncbi.nlm.nih.gov/24615625/ |
| rayyan-751571243 | Bei AK and Patel SD and Volkman SK and Ahouidi AD and Ndiaye D and Mboup S and Wirth DF | An adjustable gas-mixing device to increase feasibility of in vitro culture of Plasmodium falciparum parasites in the field. | 2014 | PloS one | 9 | e90928 | RAYYAN-INCLUSION: {"Richard"=>"Excluded"} \| RAYYAN-EXCLUSION-REASONS: wrong outcome,wrong study design | 10.1371/journal.pone.0090928 | Automation, Laboratory/*instrumentation;Carbon | PMC3946284 | https://pubmed.ncbi.nlm.nih.gov/24603696/ |
| rayyan-751571245 | Modrek S and Schatzkin E and De La Cruz A and Isiguzo C and Nwokolo E and Anyanti J and Ujuju C and Montagu D and Liu J | SMS messages increase adherence to rapid diagnostic test results among malaria patients: results from a pilot study in Nigeria. | 2014 | Malaria journal | 13 | 69 | RAYYAN-INCLUSION: {"Richard"=>"Excluded"} \| RAYYAN-EXCLUSION-REASONS: wrong outcome,wrong study design | 10.1186/1475-2875-13-69 | Adult;Diagnostic Tests, Routine/*statistics & numerical data;Early Growth | PMC3943452 | https://pubmed.ncbi.nlm.nih.gov/24564925/ |
| rayyan-751571246 | Steinhardt LC and Chinkhumba J and Wolkon A and Luka M and Luhanga M and Sande J and Oyugi J and Ali D and Mathanga D and Skarbinski J | Patient-, health worker-, and health facility-level determinants of correct malaria case management at publicly funded health facilities in Malawi: results from a nationally representative health facility survey. | 2014 | Malaria journal | 13 | 64 | RAYYAN-INCLUSION: {"Richard"=>"Excluded"} \| RAYYAN-EXCLUSION-REASONS: wrong outcome,wrong study design,wrong study duration | 10.1186/1475-2875-13-64 | Adolescent;Adult;Aged;Aged, 80 and over;*Case Management;Child;Child, | PMC3938135 | https://pubmed.ncbi.nlm.nih.gov/24555546/ |
| rayyan-751571247 | Hackett F and Berrang Ford L and FÃ¨vre E and Simarro P | Incorporating scale dependence in disease burden estimates: the case of human African trypanosomiasis in Uganda. | 2014 | PLoS neglected tropical | 8 | e2704 | RAYYAN-INCLUSION: {"Richard"=>"Excluded"} \| RAYYAN-EXCLUSION-REASONS: wrong infection / disease | 10.1371/journal.pntd.0002704 | Adult;*Cost of Illness;Disabled Persons;Female;Humans;Male;Quality-Adjusted Life | PMC3923749 | https://pubmed.ncbi.nlm.nih.gov/24551264/ |
| rayyan-751571248 | Foster D and Cox-Singh J and Mohamad DS and Krishna S and Chin PP and Singh B | Evaluation of three rapid diagnostic tests for the detection of human infections with Plasmodium knowlesi. | 2014 | Malaria journal | 13 | 60 | RAYYAN-INCLUSION: {"Richard"=>"Excluded"} \| RAYYAN-EXCLUSION-REASONS: wrong outcome,wrong study design | 10.1186/1475-2875-13-60 | Blood/*parasitology;Borneo;Diagnostic Errors;Diagnostic Tests, | PMC3931291 | https://pubmed.ncbi.nlm.nih.gov/24548805/ |
| rayyan-751571249 | Tao D and King JG and Tweedell RE and Jost PJ and Boddey JA and Dinglasan RR | The acute transcriptomic and proteomic response of HC-04 hepatoma cells to hepatocyte growth factor and its implications for Plasmodium falciparum sporozoite invasion. | 2014 | Molecular & cellular proteomi | 13 | 1153-64 | RAYYAN-INCLUSION: {"Richard"=>"Excluded"} \| RAYYAN-EXCLUSION-REASONS: wrong outcome,wrong study design | 10.1074/mcp.M113.035584 | Cell Line, Tumor/cytology/*parasitology;Epithelial-Mesenchymal | PMC4014276 | https://pubmed.ncbi.nlm.nih.gov/24532842/ |
| rayyan-751571250 | Tinto H and Diallo S and Zongo I and Guiraud I and Valea I and Kazienga A and Kpoda H and Sorgho H and OuÃ©draogo JB and GuiguemdÃ© TR and D'Alessandro U | Effectiveness of artesunate-amodiaquine vs. artemether-lumefantrine for the treatment of uncomplicated falciparum malaria in Nanoro, Burkina Faso: a non-inferiority randomised trial. | 2014 | Tropical medicine & | 19 | 469-75 | RAYYAN-INCLUSION: {"Richard"=>"Excluded"} \| RAYYAN-EXCLUSION-REASONS: wrong outcome,wrong study design | 10.1111/tmi.12274 | Amodiaquine/pharmacology/*therapeutic use;Antimalarials/pharmacolo | | https://pubmed.ncbi.nlm.nih.gov/24494602/ |
| rayyan-751571251 | Ayele DG and Zewotir TT and Mwambi HG | Semiparametric models for malaria rapid diagnosis test result. | 2014 | BMC public health | 14 | 31 | RAYYAN-INCLUSION: {"Richard"=>"Excluded"} \| RAYYAN-EXCLUSION-REASONS: wrong outcome,wrong study design | 10.1186/1471-2458-14-31 | Adolescent;Adult;Aged;Aged, 80 and over;Altitude;Child;Child, | PMC3898012 | https://pubmed.ncbi.nlm.nih.gov/24418514/ |
| rayyan-751571252 | Moiz B and Moatter T and Shaikh U and Adil S and Ali N and Mahar F and Shamsuddin N and Khurshid M | Estimating window period blood donations for human immunodeficiency virus Type 1, hepatitis C virus, and hepatitis B virus by nucleic acid amplification testing in Southern Pakistan. | 2014 | Transfusion | 54 | 1652-9 | RAYYAN-INCLUSION: {"Richard"=>"Excluded"} \| RAYYAN-EXCLUSION-REASONS: wrong outcome,wrong study design,wrong infection / disease | 10.1111/trf.12521 | Algorithms;HIV-1/*isolation & purification;Hepacivirus/*isolation & purification;Hepatitis B | | https://pubmed.ncbi.nlm.nih.gov/24383918/ |
| rayyan-751571253 | Alba S and Nathan R and Schulze A and Mshinda H and Lengeler C | Child mortality patterns in rural Tanzania: an observational study on the impact of malaria control interventions. | 2014 | International journal of | 43 | 204-15 | RAYYAN-INCLUSION: {"Richard"=>"Excluded"} \| RAYYAN-EXCLUSION-REASONS: wrong study duration | 10.1093/ije/dyt231 | Antimalarials/*administration & dosage;Child;*Child Mortality;Female;Food | PMC3937976 | https://pubmed.ncbi.nlm.nih.gov/24355745/ |
| rayyan-751571254 | Galadanci N and Wudil BJ and Balogun TM and Ogunrinde GO and Akinsulie A and Hasan-Hanga F and Mohammed AS and Kehinde MO and Olaniyi JA and Diaku-Akinwumi IN and | Current sickle cell disease management practices in Nigeria. | 2014 | International health | 6 | 23-8 | RAYYAN-INCLUSION: {"Richard"=>"Excluded"} \| RAYYAN-EXCLUSION-REASONS: wrong outcome,wrong study design,wrong infection / disease | 10.1093/inthealth/iht022 | Anemia, Sickle Cell/*therapy;Delivery of Health Care/*standards;Health | PMC6281355 | https://pubmed.ncbi.nlm.nih.gov/24114193/ |
